# Transmission of *Staphylococcus aureus* in the neonatal intensive care unit predicts invasive infection

**DOI:** 10.1101/2025.02.26.25322462

**Authors:** Qianxuan She, Lakshmi Srinivasan, Erin Theiller, Bianca E. Galis, T’Nia Napper, Andries Feder, Alex Arvanitis, Steven M. Jones, Ericka Hayes, Karen M. Puopolo, Robert N. Baldassano, Michael Z. David, Susan E. Coffin, Kathleen A. Gibbs, Kenneth Smith, Rebecca M. Harris, Joseph P. Zackular, Ahmed M. Moustafa, Paul J. Planet

**Author notes:** Corresponding authors. Joseph P. Zackular Ahmed M. Moustafa Paul J. Planet. These authors contributed equally to this work (Qianxuan She, Lakshmi Srinivasan).

## Abstract

**Background:** *Staphylococcus aureus* is a leading cause of healthcare-associated infection and is one of the most common pathogens causing serious invasive infection in the neonatal intensive care unit (NICU). Asymptomatic colonization by *S. aureus* presents a significant risk for subsequent infection of the colonized infant and for patient-to-patient transmission. Transmission of more virulent *S. aureus* strains in the NICU could increase infection risk in this vulnerable population.

**Methods:** We performed a three-year, unit-wide screening for *S. aureus* in the Children’s Hospital of Philadelphia (CHOP) NICU, with whole genome sequencing (WGS) of 1,670 isolates from patient colonization surveillance, blood cultures from both NICU and non-NICU patients, and environmental surfaces. We used comparative genomic epidemiological approaches to investigate the transmission dynamics of *S. aureus* in the NICU setting.

**Findings:** Our analyses revealed 490 unique strains with multiple, highly related, clones among NICU *S. aureus* genomes. We identified sixty-nine transmission clusters, of which 87% (60/69) were methicillin susceptible *S. aureus* (MSSA), and 28% (19/69) included isolates from both colonizing sites and bacteremia. The largest invasive cluster persisted over 2 years, involved 30 infants, and caused 4 cases of bacteremia. Spatial and temporal epidemiological links were found in 88% of clusters, and suggest that shared spaces served as the predominant means of transmission in the NICU setting. NICU environmental surveillance revealed *S. aureus* on surfaces; 3 of these isolates were also identified in colonizing and invasive clusters, suggesting potential environmental reservoirs. Remarkably, transmission clusters with both colonizing and invasive isolates were associated with higher rates of transmission, suggesting that specific transmission clusters pose a significantly higher risk for invasive infections in the NICU.

**Interpretations:** Genomic temporal-spatial epidemiologic analyses revealed high levels of *S. aureus* transmission in the NICU with multiple transmission clusters that persisted over long periods of time. Our findings demonstrate a strong association amongst colonization, transmission, and the development of invasive infections, underscoring the importance of targeted measures to prevent *S. aureus* infections in the NICU setting.

**Funding:** This work was supported by the Center for Microbial Medicine, Microbial Archive and Cryocollection and the Research Institute (CLABSI Innovation and OMICS Initiative) at CHOP.

## INTRODUCTION

Infants hospitalized in Neonatal Intensive Care Units (NICUs) are at increased risk for healthcare-associated infections (HAIs) due to factors including immature immune defenses, preterm birth, prolonged hospital stay, and use of invasive devices such as central venous catheters and endotracheal tubes^1–3^. HAIs are a major cause of morbidity, mortality, adverse long-term neurodevelopmental outcomes, and a significant contributor to the escalating cost of neonatal care ^1–3^. *Staphylococcus aureus* colonizes the skin and mucous membranes of infants early in life and is a major cause of systemic and invasive infection in this population^4–6^. A recent nationwide study involving over 100,000 very preterm infants born across the U.S. between 2018-2020 revealed that one-quarter of HAIs were attributed to *S. aureus* ^7^. Single and multicenter investigations of *S. aureus* infections in NICUs have identified significant rates of infection-specific morbidity and mortality, with one study reporting an approximately 10% case fatality rate ^7,8^. There is an urgent need for improved strategies and effective interventions to combat *S. aureus* colonization, transmission, and infection.

Currently, there are no standard approaches for NICU surveillance of *S. aureus* colonization, and there is little evidence for when to initiate or stop surveillance, how frequently to test, and which specific populations to target^9^. Current guidelines suggest that surveillance is likely to be most useful in the setting of outbreaks or ongoing healthcare-associated transmission, especially when results of surveillance will be used to direct infection prevention strategies such as cohorting, isolation, and decolonization^10^. Overall, the true impact of surveillance guided prevention strategies in decreasing rates of *S. aureus* transmission and infection remains unclear^11^.

Traditional molecular epidemiology methods, such as multi-locus sequence typing (MLST), *spa*-typing, or SCC*mec*-typing, lack the granularity required to address these questions, as genetically distinct *S. aureus* clones may differ by as few as ∼100 single nucleotide polymorphisms (SNPs)^12–16^. Whole-genome sequencing (WGS) offers a distinct advantage by enabling accurate assessment of the clonality of bacterial isolates. WGS is becoming increasingly cost-effective for tracking transmission clusters in clinical settings. While prior WGS studies have primarily focused on outbreak investigations of methicillin-resistant *S. aureus* (MRSA), few have monitored the colonization dynamics of both MRSA and methicillin-susceptible *S. aureus* (MSSA) over longer periods ^17–20^.

Here we report the results of a three-year, unit-wide surveillance screening for *S. aureus* across the Children’s Hospital of Philadelphia (CHOP) NICU, encompassing WGS of 1670 isolates from surveillance of NICU patients, blood cultures from both NICU and non-NICU patients at CHOP, and NICU environmental samples. Our findings provide clear evidence of *S. aureus* transmission and persistence within the NICU and reveal that transmission clusters within the NICU directly associate with risk for subsequent bloodstream infection, highlighting new opportunities to control colonization to prevent serious invasive infection. We also show that physical and temporal patient proximity likely drive transmission. Remarkably, clusters linked to invasive disease also exhibited enhanced transmissibility. Together, these findings highlight the need for whole genome sequencing to identify transmission in the NICU and provide evidence that targeted intervention around transmitted strains could significantly reduce invasive disease.

## METHODS

### Study Setting and Surveillance

This study was approved by the CHOP Institutional Review Board (IRB 022889 & 17-014648). CHOP NICU system comprises two facilities: a 102-bed quaternary care unit and an 18-bed Level III NICU. As a major referral center serving the mid-Atlantic region, these units collectively manage approximately 2,900 patient-days monthly and 12,000 central line-days annually.

In collaboration with the CHOP Infectious Disease Diagnostic Laboratory (IDDL), a comprehensive *S. aureus* surveillance program was implemented across both facilities. At quaternary care unit, we conducted both MRSA screening and the MRSA/MSSA monthly surveillance over 33 months, collecting 5,466 surveillance swabs from 2,092 patients across seven distinct NICU sections. In the Level III NICU, monthly MRSA/MSSA surveillance yielded 133 surveillance swabs from 95 patients.

### Study Definitions

**Bloodstream infections (BSI)** were defined as per Centers for Disease Control National Healthcare Safety Network (CDC NHSN) definitions for lab confirmed bloodstream infections (LCBI)^21^. BSI was defined as primary when no secondary source was identified, or secondary when identified to be seeded from a site-specific infection. Central line-associated bloodstream infections **(CLABSI)** are defined as a BSI in the setting of a central line in place for more than two consecutive calendar days. For *S. aureus* a single positive blood culture is sufficient to diagnose BSI.

### Whole Genome Sequencing and Bioinformatic analysis

Detailed methods are provided in Supplementary Methods (appendix p2-4). We performed whole genome sequencing on single colony isolates, with 1,446 of 1,670 assembled genomes passing quality control. A Core-genome phylogeny was constructed using panaroo and IQ-TREE. Through hierarchical clustering, we partitioned genomes into 28 groups, then performed SNP-based analysis on groups with ≥2 genomes to identify 69 transmission clusters. Genomic temporal-spatial analyses were performed using customized R scripts, and molecular clock analysis using BEAST v2.7.6 for time to the most recent common ancestor.

### Statistical analysis

The persistence duration and the number of patients involved were compared between invasive and colonizing clusters. To assess the statistical significance of differences in patient numbers and persistence between invasive and colonizing clusters, the nonparametric Mann–Whitney–Wilcoxon test with continuity correction was employed.

## RESULTS

### High prevalence of *S. aureus* colonization and infections in the NICU

We conducted active surveillance for *S. aureus* colonization in the CHOP NICU over a period of 33 months. Throughout the surveillance period, *S. aureus* was consistently detected with recurrent positive swabs from patients in the same bed locations (Figure 1A). The burden of invasive disease was substantial, with *S. aureus* isolated from 43 blood cultures taken from 41 infants across 38 distinct bed locations, spanning all seven NICU sections (Figure 1A). Stable MSSA and MRSA colonization rates were observed over time across all NICU locations. MSSA colonization rates (mean 9·04 colonized patients/1,000 patient-days; SD: 5·06; median: 10·82; IQR: 7·92–11·89) significantly exceeded those of MRSA (mean 2·18 colonized patients/1,000 patient-days; SD: 1·15; median: 1·98; IQR: 1·60–2·89) (Figure 1D). This pattern was mirrored in bloodstream infection (BSI) rates, where MSSA BSIs (mean: 0·37 BSIs/1,000 patient-days; SD: 0·28; median: 0·33; IQR: 0·32-0·36) occurred more frequently than MRSA BSIs (mean: 0·06 BSIs/1,000 patient-days; SD: 0·13; median: 0; IQR: 0–0) (Figure 1D). Together, results from this surveillance effort demonstrate high prevalence of *S. aureus* colonization and infections in the NICU over the period of our study.

**Figure 1.**
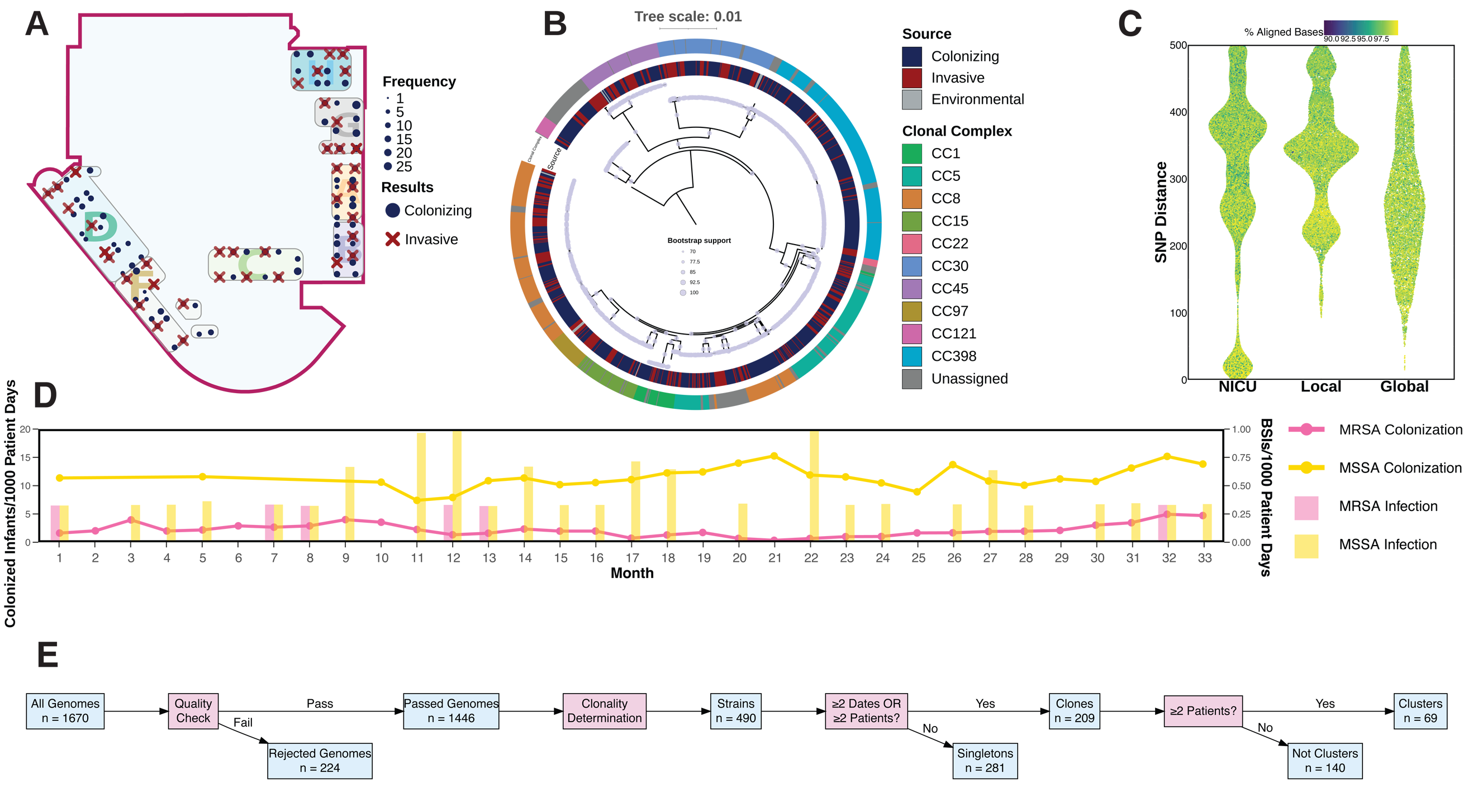
Spatial distribution and genomic diversity of *S. aureus* isolates in the NICU setting. (A) NICU floorplan visualization of *S. aureus* MRSA/MSSA surveillance results: positive *S. aureus* colonizing samples (blue dots, size proportional to frequency) and *S. aureus* bacteremia samples (red crosses). (B) Core-genome phylogenetic analysis with source distribution (inner ring) and clonal complex assignment (outer ring). Genomes assigned to Clonal Complexes 1, 5, and 8 were paraphyletic in the phylogenetic analysis. Bootstrap values ≥70 support 66·4% of tree branches. (C) Pair-wise SNP analysis reveals clonal relationships among *S. aureus* genomes by comparing genomes across three datasets: (1) within NICU genomes (NICU), (2) against a local Philadelphia pediatric database (Local), and (3) against a curated global publicly available database (Global). (D) Monthly MSSA/MRSA colonization and infection rates. (E) Workflow schematic for identifying strains, clones, and transmission clusters. Red boxes represent decision points, while blue boxes indicate process outputs, serving as inputs for subsequent analytical steps.

### Genomic Diversity and Clonality among NICU *S. aureus* isolates

We performed WGS on 1,670 isolates, comprising 1,170 *S. aureus-*positive surveillance isolates, 482 NICU and non-NICU *S. aureus-*positive blood isolates, and 18 isolates from *S. aureus-*positive environmental sampling isolates.

Following *de novo* assembly and stringent quality control, 1,446 high-quality genome assemblies were retained for subsequent analysis (982 surveillance, 449 blood cultures, and 15 environmental isolates). This comprehensive genomic dataset allowed high-resolution investigation of clonality and transmission dynamics. The 1,446 genomes belonged to ten clonal complexes (CCs) and 66 sequence types (ST). CC398, CC8, and CC5 were the predominant clonal complexes. A core-genome maximum-likelihood tree (Figure 1B) indicated substantial genomic diversity of *S. aureus* isolates in the NICU setting. Notably, genomes from surveillance swabs (colonizing), positive blood cultures (invasive), and environmental samples were interspersed across all major phylogenetic clades, showing that the ability to cause invasive infections was not restricted to specific lineages, remaining a potential outcome for various genetic backgrounds.

We identified 328 patients with two or more genomes, yielding a total of 1,142 genomes. While most of those genomes tended to form monophyletic groups (median 3 genomes/patient, IQR 2-4), 158 genomes from 36 patients (median 4 genomes/patient, IQR 2-5) were from distinct phylogenetic clades. Twenty-four distinct phylogenetic clades of highly related genomes were identified, encompassing 1442 genomes (median: 26 genomes/clade, IQR 4-73) from 657 patients (median: 11 patients/clade, IQR 2-36).

Clonal relationships were further determined through comprehensive single nucleotide polymorphism (SNP) analysis using three datasets. We constructed pairwise SNP distance matrices comparing NICU genomes against: (1) themselves (NICU matrix), (2) a local database of 349 pediatric *S. aureus* genomes collected in Philadelphia (Local matrix)^22^, and (3) a curated global database of 68,298 high-quality publicly available *S. aureus* genomes^23^. The NICU matrix uniquely demonstrated a significant cluster of closely related genomes (SNP distance <100) (Figure 1C).

Based on the phylogenetic pattern, clonality, and high rates of *S. aureus* colonization across the NICU, we hypothesized that at least some infants may have acquired *S. aureus* through transmission within the NICU. To rigorously test for transmission, we used a SNP threshold approach corrected for phylogenetic structure, which allowed for flexible SNP thresholds across different groups (appendix p8). This approach revealed that conventional single, arbitrary SNP thresholds occasionally did not capture the clonal relationships within the phylogenetic trees, as putatively linked genomes were not always monophyletic (appendix p11). By incorporating phylogeny-corrected group-specific SNP thresholds, we refined the approach to better reflect the phylogenetic inference. This approach also improved upon the conventional method by avoiding the limitations of using a single threshold across all genomes from different genetic backgrounds. Instead, it yielded a set of group-specific thresholds tailored to various genetic backgrounds, which remain in line with standards set across numerous other studies^12,13,15,16^.

### Analysis of widespread transmission revealed strong link to infections and suggested source of transmission

We defined a “strain” as either a group of genomes within a SNP threshold or singletons with no SNP distance matches. A “clone” was designated as a strain detected in a sample obtained on more than one date or patient, while a “transmission cluster” refers to a clone found in more than one patient. Using the above thresholds, we identified 490 strains (360 NICU, 130 non-NICU), 209 clones (150 NICU, 59 non-NICU), and 69 transmission clusters (69 NICU) (Figure 1E, appendix p9,10). We observed 609 NICU colonization events, and 180 (29·56%) were identified as transmission-acquired after excluding 69 index patient cases (the first patient in which the cluster was observed) (appendix p12). CC398 MSSA was most prevalent, with 62/490 strains and 14/69 clusters. This observation aligns with many studies reporting the recent global spread of the CC398 MSSA lineage^24^, as well as findings from other NICUs in the US^17^. Of 69 transmission clusters, 19 “invasive clusters” were identified based on the detection of the strain from any BSI (Figure 2A). Among these invasive clusters, CC398 MSSA accounted for 5/19 (26%) invasive clusters, highlighting the association between CC398 MSSA and HAIs and bloodstream infections in the NICU setting. Of the 26 BSI from the 19 invasive clusters, 17 infections (26.98%) were attributed to transmission after excluding 9 index patients (appendix p12). Cluster 1 was the largest transmission cluster, involving 30 patients, and four patients were diagnosed with bacteremia infections associated with this clone.

**Figure 2.**
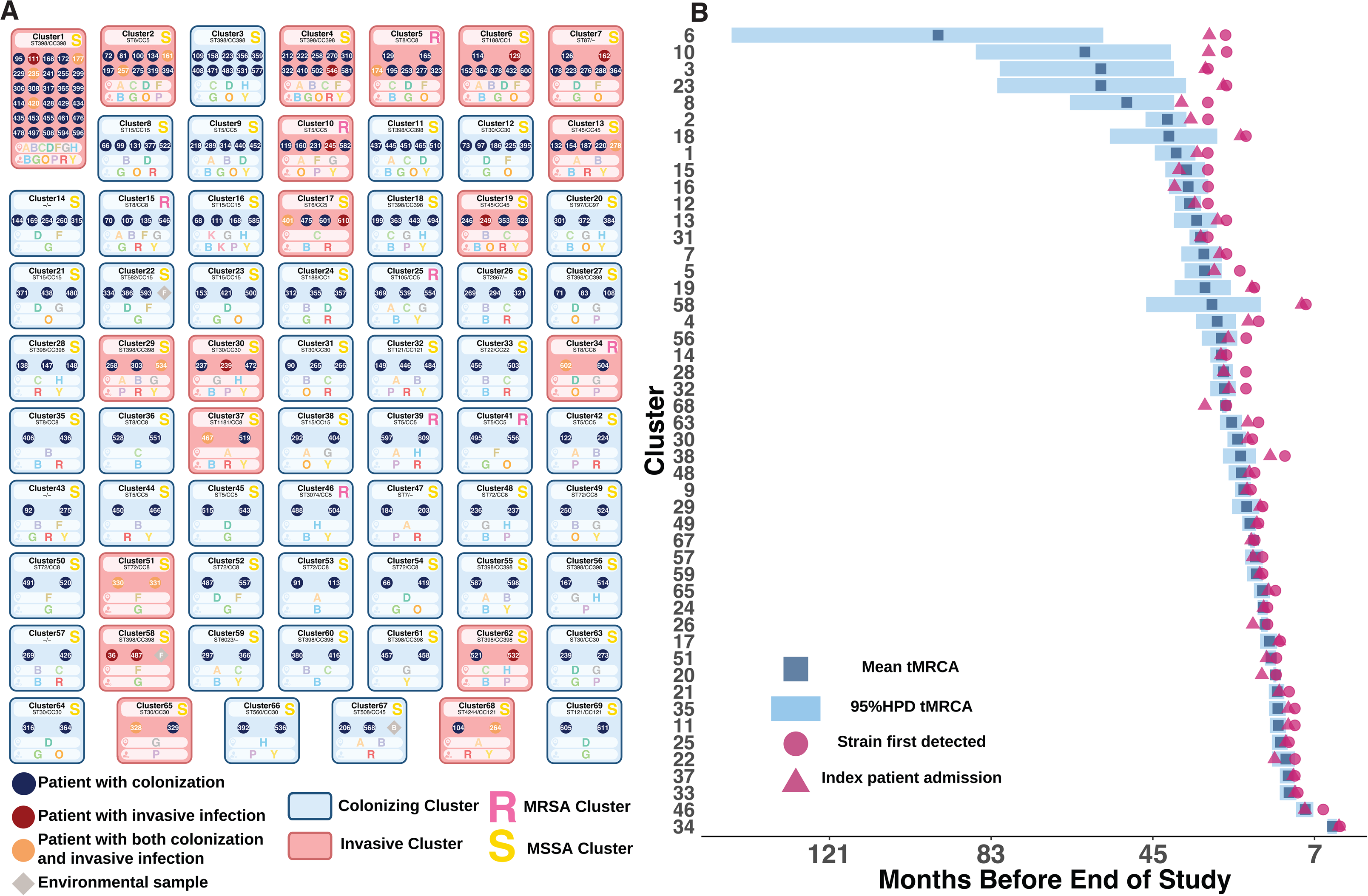
Cluster characterization and origin analysis. (A) Transmission clusters (n=69) represented as boxes, colored according to invasive status (red: invasive clusters; blue: colonizing clusters) and annotated with methicillin resistance status (R: MRSA; S: MSSA). Within each cluster, patient status is indicated by colored dots (blue: colonization only; red: infection only; yellow: both colonization and infection), with individual patient IDs shown as numbers. Environmental isolates are marked by grey diamonds. Two bars below each cluster box indicate: (1) the specific NICU section where the cluster was detected, and (2) the assigned treatment team at the time of detection. Among all transmission clusters identified, 19 were classified as invasive. (B) Comparative analysis of cluster tMRCA (time to most recent common ancestor) versus index patient admission dates, revealing 50% of clusters originating from index patient admission. X-axis (Months Before End of Study) represents the number of months prior to completion of study period.

We hypothesized that some transmission clusters might have been introduced via the admission of index patients pre-colonized by the respective strains. This was tested by comparing the admission date of the putative index patient to the time of the most recent common ancestor (tMRCA) using a molecular clock approach performed on 48 evaluable transmission clusters, as this approach requires a minimum of four genomes per transmission cluster (Figure 2B). In 50% (24/48) of the clusters, admission dates fell within the 95% highest posterior density (HPD) interval of the tMRCA, indicating a high likelihood that the transmission clusters were introduced through the admission of index patients colonized by each strains. For the remaining 38% of clusters (16/42), the tMRCA predated or postdated the index patient’s admission date, suggesting the possibility that those transmission clusters were introduced into the NICU environment through alternative routes, or by unsampled index patients. Together, these findings demonstrate that transmission accounted for a substantial proportion of both colonization and infection events in our NICU setting, and that transmission clusters predominantly originated from the index patients upon admission, highlighting the potential for reducing invasive disease through early identification and targeted interventions focused on transmission.

### Geospatial and temporal proximity drives transmission

Only eight of the 69 transmission clusters were MRSA (appendix p3), with three invasive clusters and five colonizing clusters, suggesting that antibiotic resistance alone does not fully explain transmission and persistence (Figure 2A). We hypothesized that geospatial and temporal proximity would be the largest driver of transmission. As exemplified in Cluster 1 (Figure 3AC), temporal and geospatial proximity was observed among patients in Section H, generating a strong epidemiological link in the cluster. The geospatial proximity was also found in Section F and D. Serving as a potential proxy for unsampled healthcare workers, the shared treatment team assignments revealed potential healthcare worker-mediated transmission routes, complementing the epidemiological link in the cluster. For instance, despite physical separation in distinct NICU sections, patients 229 and 235 were linked through concurrent care by the same NICU treatment team. This pattern was also found in patients 414 and 399, who shared the same treatment team during their stay in different sections (Figure 3B). Besides Cluster 1, similar links were seen in 88% of transmission clusters (appendix p13-80). Notably, Cluster 58 represented the sole instance in which *S. aureus* spread between NICU and non-NICU sections. For the 12% of transmission clusters lacking strong epidemiological links, transmission may have occurred through unsampled intermediaries, shared equipment or environmental reservoirs not observed in this study. These epidemiological links demonstrate that shared NICU spaces and care teams serve as the primary drivers of transmission, underscoring the critical role of geospatial and temporal proximity in transmission dynamics.

**Figure 3.**
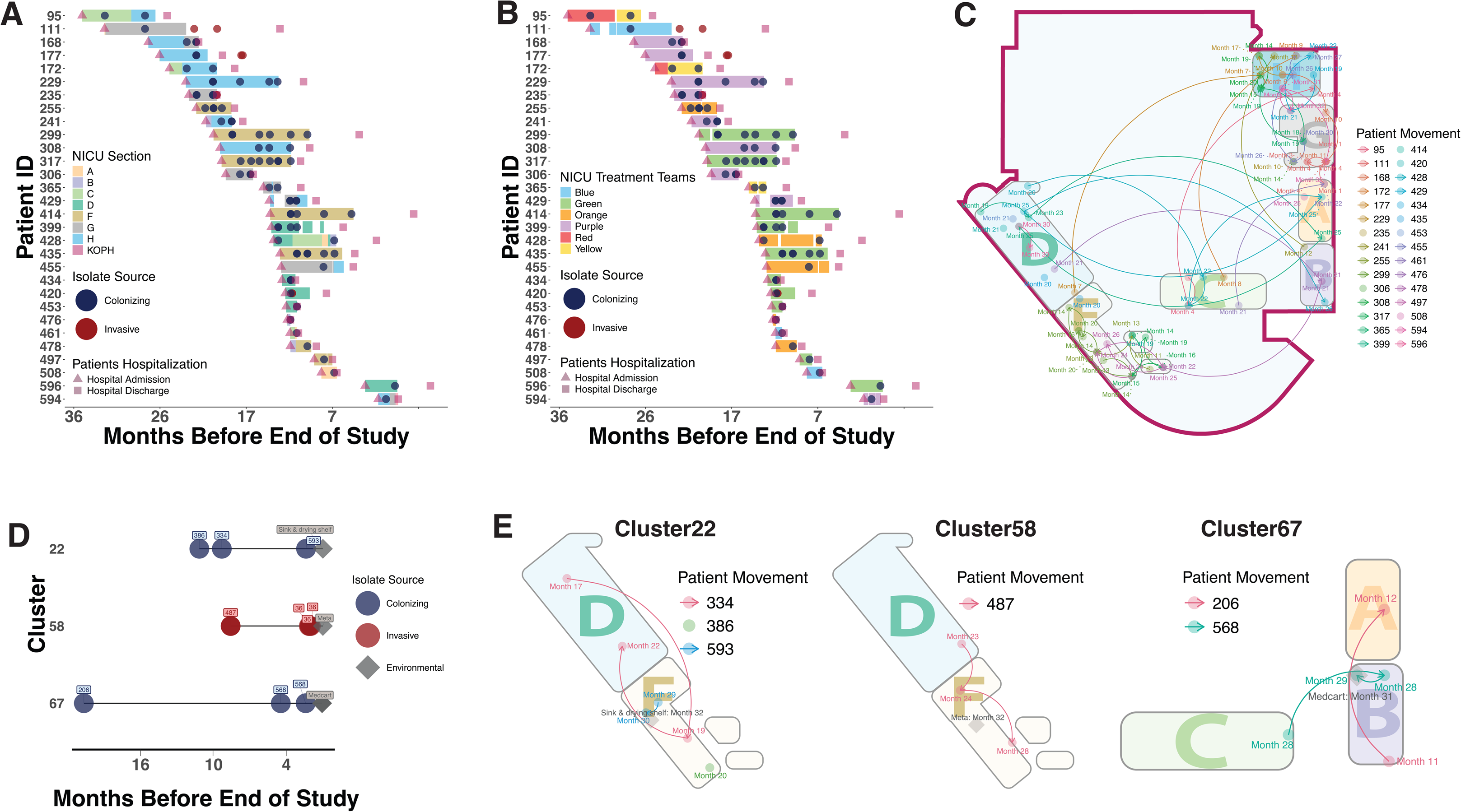
Geospatial and temporal analysis of NICU transmission dynamics and persistence of environmental reservoirs Fig 3. (A-B) Temporal analysis of Cluster 1 by location and treatment team assignments. (A) Patient timeline showing NICU sections (colored rectangles), with admission (triangles) and discharge dates (squares). (B) Corresponding treatment team assignments over time (colored rectangles). In both A and B panels, sampling events are indicated by dots (red: invasive isolates; blue: colonizing isolates). (C) NICU floorplan visualization of patient movements. Geospatial and temporal proximity serves as the primary driver of transmission, as visualized in the floorplan. This visualization reveals that certain NICU sections had a high concentration of patient movements, posing increased risk for transmission and infection. (D) Temporal distribution of isolate sampling (red dot: invasive, blue dot: colonizing, grey diamond: environmental), demonstrating the prolonged persistence of environmental reservoirs. (E) Spatial mapping of environmental reservoirs (grey squares) relative to patient movement within the clusters. For Panel A, B, and D, X-axis (Months Before End of Study) represents the number of months remaining until the study period concludes.

To further detect potential reservoirs of transmission, we surveyed 210 surfaces and sites in all seven NICU sections using swabs and broad-based culture methods. We collected 192 bacterial isolates (multiple species), and 15 of 18 *S. aureus* isolates produced high-quality *S. aureus* genomes (sampling locations in appendix p217). Remarkably, we recovered environmental isolates from three of the previously defined transmission clusters (Clusters 22, 58, 67) (Figure 3 DE). There was high spatial proximity between environmental isolate collection sites and patient beds in the cluster for all three, strongly implicating environmental reservoirs (Figure 3E). Persistence in the NICU environment was observed in these clusters, as evidenced by the time intervals between the most recent cluster detections and the corresponding environmental isolate collection dates (30-42 days). Persistence occurred despite routine NICU sterilization procedures, indicating the resilience of potential environmental reservoirs.

### Persistence is associated with increased invasive infection

We examined the length of cluster persistence in the NICU by measuring the time between first isolation date of a transmission cluster and the most recent date of detection. Invasive clusters, defined by at least one BSI isolate in the cluster, demonstrated significantly higher persistence (median 421 days, IQR 212-570) than that of the 50 colonizing clusters (median 128 days, IQR 35.25-394) (Figure 4A). To further assess the persistence of transmission clusters in the NICU, we employed a molecular clock approach to calculate the tMRCA for 48 transmission clusters with more than four genomes (18 invasive, 30 colonizing). The average and median molecular clock persistence times of 18 invasive clusters (median 576 days, IQR 304.5-907.25) were higher than those for the 30 colonizing clusters (median 383 days, IQR 195.5-655.25) (appendix p9,10). These analyses suggest that persistence in the NICU is highly associated with subsequent invasive infection. Additionally, our analyses revealed transitions from colonizing to invasive states in several invasive clusters. For instance, Clusters 1, 2, and 4, which were the top three most persistent invasive clusters, initially manifested as colonizing clusters for the first few months before causing invasive disease (Figure 4D). These findings suggest that strains with enhanced persistence and transmission capabilities pose a significantly higher risk for invasive infections in the NICU setting. This increased risk may be mediated by bacterial virulence factors that promote both persistence and invasive disease, as well as by host characteristics that render the infant more vulnerable to persistent colonization and infection.

**Figure 4.**
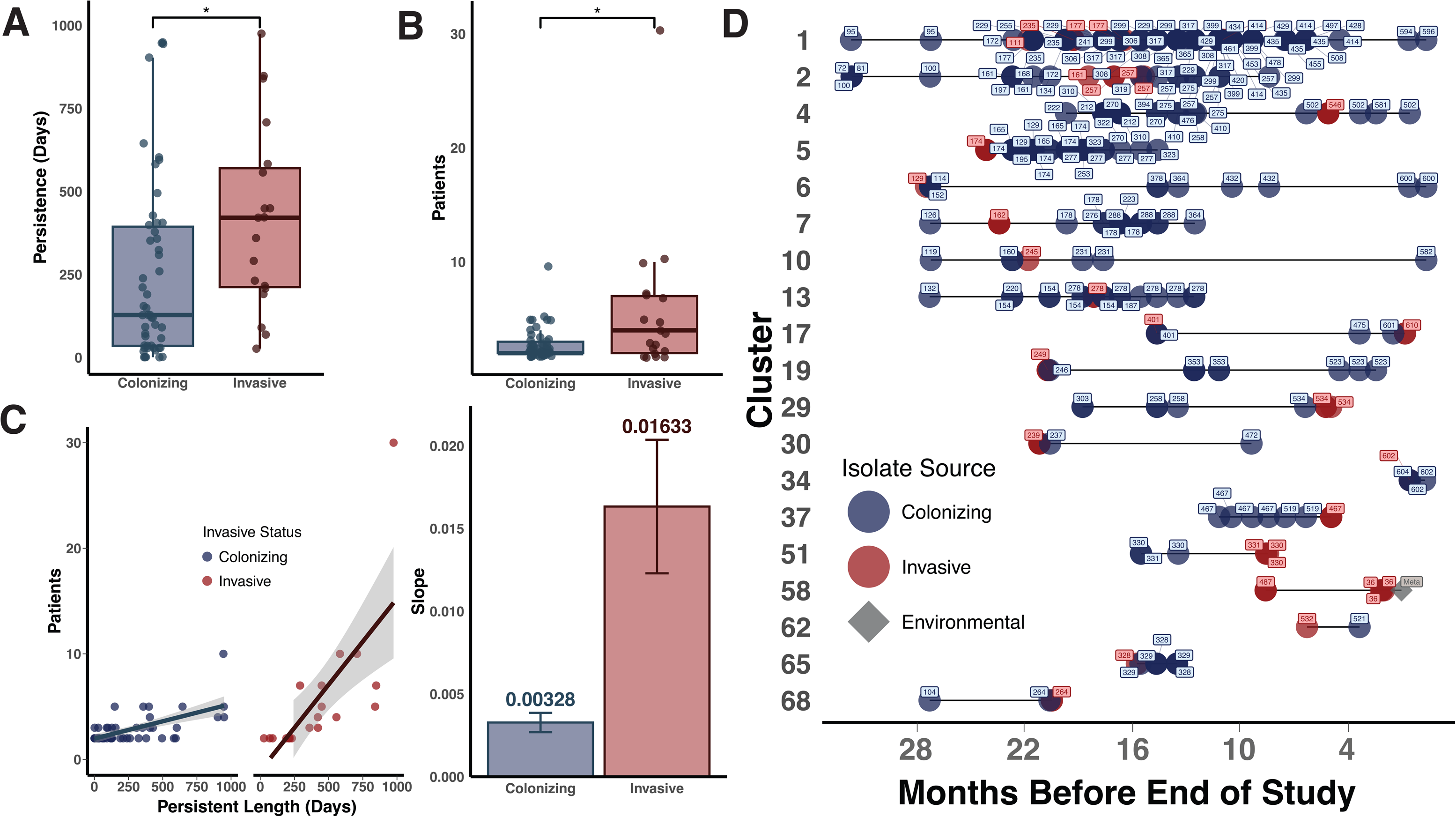
Distinct epidemiological characteristics of invasive clusters predict infection. (A) Comparison of persistence times of invasive versus colonizing clusters (Wilcoxon test, *p < 0·05). (B) Comparison of number of patients involved between invasive and colonizing clusters (Wilcoxon test, *p < 0·05) (C) Transmissibility comparison quantified using linear regression slopes shows five-fold higher transmissibility in invasive clusters, serving as a critical predictor for subsequent infection of a high transmissible strain. (D) Temporal distribution of isolate sampling, with invasive (red) and colonizing (blue) sources, demonstrating transitions from colonizing to invasive status after prolonged persistence in the NICU setting. X-axis (Months Before End of Study) represents the number of months prior to completion of study period.

### Enhanced transmissibility predicts risk of invasive infections

Invasive clusters persist longer than the colonizing clusters (median 4 patients, IQR 2-7) and involve more patients (median 2 patients, IQR 2-3) (Figure 4B). To test if invasive clusters involve more patients across equivalent time periods, we quantified transmissibility by using the slopes of linear regression models fitted to the correlation between the number of patients and the duration of persistence. Invasive clusters exhibited markedly enhanced transmissibility with a transmissibility slope that was approximately five-fold higher than colonizing clusters. (Figure 4C). This striking finding suggests that certain clones pose significantly greater risk by being more transmissible and also more invasive and provides a clear rationale for prioritizing surveillance and prevention of these particular strains. Together, this comprehensive study of *S. aureus* in the NICU setting creates a new paradigm for our understanding of colonization, transmission, and invasive infection of *S. aureus* in high-risk neonates and provides a roadmap for future surveillance and intervention strategies.

## DISCUSSION

Infections due to *S. aureus* remain a major cause of morbidity and mortality in the NICU setting^7^. Colonization is associated with increased risk of infection, and exposure to the NICU environment and equipment, healthcare workers, and parents are likely the major factors that impact transmission and colonization^25–27^. Despite decades of infection prevention efforts and collaborative implementation of bundles of care, rates of invasive infections such as central line-associated bloodstream infections have plateaued, especially in complex and high-risk populations in children’s hospitals^27^. New strategies are needed to mitigate *S. aureus* transmission and infection in the NICU. In this study, we employed WGS of samples from surveillance screening and prospective collection of all invasive infections in the hospital to measure and model the dynamics of introduction, transmission, and persistence of *S. aureus* in our NICU.

For determining transmission or persistence of clones most studies rely on SNP thresholds that are usually set at a single value across groups of genomes, which can lead to false negative and positive assignment of genomes to clonal groups^28^. In addition, the length of time a strain has been persistent, and variations in mutation rate could make single cutoffs problematic^29^. We show that even conservative cutoffs can produce transmission clusters that are incongruent with clonal groups on a phylogenetic tree. Thus, we used a strategy that allows for different cutoffs across groups and corrects for phylogenetic discrepancy. Our technique also presents the inferred number of transmission clusters at every threshold value, which allows for assessment of more conservative as well as broader thresholds (appendix p11). We also compared our genomes to non-study bacterial genomes, showing that our chosen thresholds are robust to many other genome comparisons (Figure 1C). This new method captures clear clonal relationships within the NICU genomes, serving as evidence of the robustness of the inference of “sameness” and providing a more comprehensive understanding of transmission dynamics and clonal structures.

WGS has been used previously for tracking *S. aureus* transmissions in NICU settings ^17–19^. However, previous studies have lacked analyses combining both MRSA and MSSA, and only a small number of studies have identified links between transmission and invasive infection^19,30–34^. Our study leveraged a comprehensive collection of isolates from multiple sources in the CHOP NICU: routine surveillance samples, infant blood culture and environmental samples, generating genomes from both MRSA and MSSA samples. The prolonged length of surveillance and the large number of samples sequenced ensure the robustness of the inference of the dynamics of transmission, persistence, and introduction. For instance, the extended surveillance period allowed for the granular detection of the largest transmission cluster that was persistent for 975 days and involved 30 patients. A total of 69 transmission clusters were identified, with 26 BSIs linked to 19 clusters. We also mapped transmission within NICU sections to assess geospatial impacts, a novel approach in this context.

Currently, most infection prevention interventions are uniformly applied across the NICU and include standard measures such as improving hand hygiene compliance and applying bundles of care to prevent device-associated infections^35^. Patient-level interventions to reduce transmission of specific organisms, such as patient care cohorting and contact precautions, have primarily been used to limit transmission of MRSA^36,37^. Decolonization regimens such as use of short courses of intranasal mupirocin and topical chlorhexidine have also been used for MRSA decolonization, though some centers have employed them in patients colonized with MSSA, especially in the setting of outbreaks or increased rates of infection^38–41^. In this study, we demonstrate several findings that could be key to defining new approaches to infection prevention: that is, the use of surveillance for specific bacterial strains associated with higher risk of transmission and invasive infection. Our findings suggest that there are inherent properties of some bacterial strains that confer both increased transmissibility and risk of invasive infection. Thus, identification of highly transmitted strains, and devising targeted strategies to mitigate transmission, could reduce the risk of invasive infection by *S. aureus.* Identification of the biological and genomic properties of more invasive and transmissible strains will be a major focus of future work. We also identified links between environmental sources of *S. aureus* and invasive clusters, highlighting that environmental deep cleaning efforts may need to be part of intensive efforts to eliminate high risk strains of *S. aureus* from their environmental reservoirs.

This study has several potential limitations. As a single-center investigation, our findings may not be fully generalizable, particularly to lower-acuity settings. Additionally, our persistence hypothesis might underestimate instances of strain reintroduction to the NICU. Previous studies have reported that healthcare workers can be sources of transmission to patients in clinical settings^12^. The epidemiological characteristics of some of our transmission clusters suggest this possibility. However, we did not sample healthcare workers and this limits our ability to test this hypothesis.

Our study shows extensive transmission, persistence, and introduction in the NICU of both MRSA and MSSA, and that MSSA represents a larger problem for both colonization and invasive disease than MRSA. To our knowledge this is the first report that links high rates of transmission to invasive *S. aureus* clusters in the NICU setting. This observation suggests that detecting colonization and transmission is critical to infection prevention and control and also provides support for interventions that target highly transmissible strains in real time. In NICU settings where standard infection prevention measures are not accomplishing further reduction in infection rates, WGS directed interventions may provide additional infection prevention strategies in the following ways – (1) WGS can broadly inform NICUs of the burden of transmission, persistence and environmental reservoirs, to help direct efforts and resources towards the most important interventions; (2) WGS can aid in targeting strains at highest risk of causing invasive infections, and (3) WGS can assess the effectiveness of interventions. Targeted interventions could include heightened surveillance of patients and environmental sites, enhanced approaches to reducing transmission such as patient care cohorting and contact precautions for strains identified as high risk, and enhanced decolonization and environmental cleaning efforts. A WGS based approach could provide a new framework for defining patients at highest risk of infection, by evaluating underlying patient risk factors as well as the presence of high-risk bacterial strains.

## Data sharing

Genome assemblies and associated limited metadata have been deposited in the National Center for Biotechnology Information (NCBI) under BioProjects PRJNA1218714, PRJNA1218736, and PRJNA1219099. The analysis scripts are publicly accessible through our GitHub repository (https://github.com/qianxuans/CHOP-NICU-Scripts). The transmission cluster identification pipeline is accessible as a command-line interface through GitHub (https://github.com/microbialARC/THRESHER).

## Acknowledgments

The authors would like to acknowledge the Microbiome Sequencing Center, the Microbial Archive and Cryocollection (MicrobialARC), and the Center for Microbial Medicine, and the Research Institute at the Children’s Hospital of Philadelphia. We would also like to acknowledge the support and efforts provided by the Infectious Disease Diagnostics Laboratory (IDDL) and Neonatal Intensive Care Unit (NICU) at the Children’s Hospital of Philadelphia.

