## Supplementary Appendix for "Transmission of *Staphylococcus aureus* in the neonatal intensive care unit predicts invasive infection"

#### Supplementary Methods

##### Whole Genome Sequencing

Single colony isolates from the CHOP IDDL were stored at −80°C at the CHOP microbial ARchive and Cryo-collection (microbialARC) strain bank and were sequenced at the CHOP Microbiome Center. DNA from all isolates were extracted using the Illumina Direct from Colony protocol. Extracted DNA was quantified with the Quant-iT PicoGreen® dsDNA assay kit (ThermoFisher) before library generation. Libraries were generated from 7.5 ng DNA using the Illumina DNA Prep kit and IDT for Illumina unique dual indexes at 1:4 scale reaction volumes. Library success was assessed by Quant-iT PicoGreen® dsDNA assay kit and samples with library yields < 1 ng/ul were re-prepped as needed. After all samples for a given pool were prepped, an equal volume of library was pooled from every sample and then the pool was sequenced using a 300 cycle Nano kit on the Illumina MiSeq. Libraries were then repooled based on the demultiplexing statistics of the MiSeq Nano run. Final libraries were sequenced on an Illumina NovaSeq 6000 v1.5 flow cells, producing 2x150 bp paired end reads. Extraction blanks and nucleic acid-free water were processed along with experimental samples to empirically assess environmental and reagent contamination. A laboratory-generated mock community consisting of DNA from *Vibrio campbellii* and Lambda phage were included as a positive sequencing control.

##### Genome Assembly and Quality Control

Raw reads underwent quality control using Sunbeam 4.3.7, including adapter trimming and host decontamination [1]. Isolate genomes were assembled using SPAdes v3.15.5 [2], while 102 metagenomic samples were processed through the Anvi'o v8 pipeline using MEGAHIT v1.2.9 [3,4]. Assembly quality was assessed using CheckM with the following criteria: ≥95% completeness, ≤5% contamination, and "Staphylococcus (UID301)" lineage classification. Species-level contamination was evaluated using Mash[5]. Assemblies were filtered to retain genomes between 2.55 Mb and 3.15 Mb. Of 1,670 assembled genomes, 1,446 passed quality criteria and were used for downstream analyses (https://github.com/qianxuans/CHOP-NICU-Scripts/blob/main/Genome_assembly_and_quality_control/passed_genomes.xlsx). Genomes rejected from quality control appeared to be a random subset of the data; we found no temporal, patient-based, location, sequence type, or anatomical collection site associations (https://github.com/qianxuans/CHOP-NICU-Scripts/blob/main/Genome_assembly_and_quality_control/failed_genomes.xlsx). All passed assemblies are available on NCBI (Accession numbers available in https://github.com/qianxuans/CHOP-NICU-Scripts/blob/main/Genome_assembly_and_quality_control/passed_genomes.xlsx).

The genome assemblies demonstrated robust sequencing depth with mean coverage of 90·22X (median: 76·79X; IQR: 42·78-115·92X) and assembly contiguity as reflected by contig length averaging 18,541.07 base pairs (median: 18·361 bp; IQR: 13,357·5-22,566·5 bp). This comprehensive genomic dataset allowed high-resolution investigation of clonality and transmission dynamics. The genome assemblies deposited in the NCBI GenBank database may exhibit minor variations from those employed in our analyses, due to GenBank's standard processing pipeline which includes sequence trimming and the exclusion of contigs below 200 base pairs.

##### Core-genome Phylogeny Inference

The 1,446 high-quality genome assemblies were annotated using Bakta v1.9.2 with Database v5.1 [6]. The resulting gff3 files were utilized for core-genome alignment, which was carried out using panaroo 1.5.0 [7]. This alignment was then used to infer the maximum likelihood phylogeny in IQ-TREE version 2.3.0 [8], employing the general time-reversible (GTR) substitution model [9] and accounting for among-site rate heterogeneity using the Gamma distribution and four rate categories [10].

The phylogeny inference was performed with 100 initial parsimony trees. Unbiased branch support values were provided by UFBoot using 10,000 bootstrap replicates[11]. SH-like approximate likelihood ratio test was implemented with 10,000 replicates to assess branch support[12]. The resulting phylogeny was midpoint-rooted, and the Interactive Tree of Life (iTOL) was used for visualization and annotation [13].

##### Clonality Determination

Two analyses were performed to produce the input for the analysis to determine strain compositions.

On one hand, pairwise SNP distances were calculated using the wrapper dnadiff in MUMmer 4.0.0rc [14], and the gSNP in the dnadiff output was used as the number for SNP distance. We constructed pairwise SNP distance matrices comparing NICU genomes against: (1) themselves (NICU matrix), (2) a local database of 349 pediatric *S. aureus* genomes collected in Philadelphia (Local matrix)[15], and (3) a curated global database of 68,298 high-quality publicly available *S. aureus* genomes[16]. A close clonal relationship can be observed in the SNP distance matrices, where only NICU matrix presents a unique cluster below 100 SNP distances.

On the other hand, we used a hierarchical clustering approach to partition all genomes into 28 groups using the core-genome phylogenetic tree described previously in **Core-genome phylogeny inference**. During the hierarchical clustering process, we started by computing the pairwise distances between any pair of genomes using branch lengths in the phylogenetic tree using the cophenetic.phylo function in R package ape[17]. The generating distance matrix was used to test every possible grouping pattern using function hclust from stats, a built-in R package. To evaluate the quality of the grouping pattern, we calculated the average silhouette scores using function silhouette from R package cluster[18], and grouping pattern with the highest average silhouette scores was used to

partition the genomes. Every group with no less than two genomes was supported with 100 ultrafast bootstrap support [11]and 100 SH-like approximate likelihood ratio test support (SH-aLRT)[19], expect for Group2 with 97 SH-aLRT support (appendix p6).
In total 28 groups were determined with hierarchical clustering. A quality check step was conducted before the downstream analysis. If there is only one genome in a group, that genome is considered a singleton. For the groups with no less than 2 genomes, an arbitrary threshold at 1000 SNP, informed by the natural gaps in the patterns of SNP distance matrices, was used to detect the potential singleton members in the group. In a group, if any member has no less than 1000 SNP distances to any of the rest members, the member is considered a singleton. If every member has no less than 1000 SNP distance to any of the rest members, every member is considered singleton. Removing those singletons minimize the possibility of the presence of extra-long branches in the phylogenetic trees due to the outliner members. Those long branches could compromise the accuracy of the analysis.
Twenty-four out of 28 groups consisted of no less than two genomes, averaging 60 genomes (median 26, IQR 4-73). After excluding three singletons from three groups, subsequent fine-grained phylogenetic analysis was implemented. For each of the 24 groups, we used the curated database of 68,298 high-quality *S. aureus* genomes mentioned previously to identify five most closely related publicly available genomes for every study genome in the group. Those publicly available genomes restricted the thresholds to focus on potential transmission clusters within the NICU. Excessively high thresholds including public genomes in the strains were considered unsuitable, as they would likely extend beyond the scope of intra-NICU transmission. The multi-sequence alignments containing study genomes and closest publicly available genomes were subsequently produced using Snippy 4.6.0[20], and the study genome with highest N50 in each group was used as the reference genome. The resulting alignments were used as the input for inferring the reference-based maximum-likelihood phylogenetic trees with IQ-TREE version 2.3.0 [8], employing the general time-reversible (GTR) substitution model [9] and accounting for among-site rate heterogeneity using the Gamma distribution and four rate categories [10], and supported with standard bootstrap supports[21]. We implemented a single linkage clustering (SLC) algorithm to determine closely related strains at every possible potential SNP threshold from the minimal distance in the group to 500. At each SNP threshold tested, the strain compositions determined by pairwise SNP distances were mapped to the corresponding reference-based phylogenetic tree of the group. Interestingly, even at conservative SNP thresholds the strains determined by SNP thresholds were sometimes not monophyletic on the phylogenetic tree (Multi-SNP-Threshold Plot). To correct for this problem, we expanded our strains to include the smallest clade that included all SLC members as well as any non-SLC members in that clade when the bootstrap support of this clade is no less than 70. We then selected the SNP threshold at which the number and composition of strains plateaued. The plateaus of 24 groups averaged 21.58 (median 18.5, IQR 10.75-27.25), and 486 strains were determined (223 singletons and 263 clones).
The analytical codes are publicly available in the project's GitHub repository (https://github.com/qianxuans/CHOP-NICU-Scripts). The ready-to-use pipeline performing the analysis is publicly available for download at https://github.com/microbialARC/THRESHER

**Cluster analysis and visualizations**

If any bacteremia isolate was present in a transmission cluster, it was defined as an invasive cluster; otherwise, it was referred to as a colonizing cluster.
Methicillin resistance in transmission clusters was confirmed through clinical laboratory testing and detection of the mecA gene using BLASTx 2.14.0+ against a customized protein database containing 13 PBP2a family β-lactam-resistant peptidoglycan transpeptidase amino acid sequences from the NCBI protein database [22,23]. The accession numbers of the sequences were WP_057521704.1, WP_063852670.1, WP_063852677.1, WP_063852683.1, WP_000721309.1, WP_000721306.1, WP_063852626.1, WP_063852617.1, WP_000721310.1, WP_063851348.1, WP_012655867.1, WP_063852710.1, and WP_000725529.1.
We used customized R scripts to create visualizations. These included plots depicting patient metadata, invasive status, methicillin resistance, and cluster persistence. Additionally, we generated swimmer plots and annotated NICU floor plans using separate customized R scripts. All scripts are available from https://github.com/qianxuans/CHOP-NICU-Scripts.

**Molecular Clock Analysis**

For genomes identified as belonging to the same strain, we generated whole-genome alignments and masked putative recombination regions using ClonalFrameML [24]. Time to most recent common ancestor (tMRCA) was estimated using BEAST v2.7.6 [25], employing an HKY substitution model and strict molecular clock. The analysis incorporated a coalescent constant population model with clock rates following log-normal distributions (1.0E-6 to 1.0E-7) as Bayesian priors. Markov Chain Monte Carlo (MCMC) chains were run up to 500 million steps to ensure effective sample sizes exceeded 200 for all parameters. The resulting phylogenetic trees were generated in NEXUS format and can be accessed at https://github.com/qianxuans/CHOP-NICU-Scripts/tree/main/Molecular_Clock_Analysis/BEAST2_Tree_Visualization/nexus. All analysis scripts and visualization tools are publicly available in the project's GitHub repository (https://github.com/qianxuans/CHOP-NICU-Scripts).

#### Extended Discussion

##### Pathogen Trends in BSIs

While bloodstream infections due to coagulase-negative staphylococci have declined in our NICU, *S. aureus* (mostly MSSA) remains the predominant pathogen. Point prevalence testing in the CHOP NICU indicates high *S. aureus* colonization rates among NICU infants, primarily MSSA with a few MRSA cases.

##### Infection Prevention Measures

Current infection prevention measures are broadly applied, but our findings suggest strain-specific approaches may be more effective. Certain *S. aureus* strains exhibit both high transmissibility and invasive potential, highlighting the need for targeted mitigation strategies such as enhanced environmental cleaning and selective decolonization. Identifying these high-risk strains could refine prevention efforts beyond standard measures. Alternatively, other factors may facilitate transmission, allowing specific strains to colonize a larger pool of vulnerable hosts. When conditions permit (e.g., a breakdown of physical or immunological barriers), these strains can cause invasive infections.

##### Limitations

Our strain determination method, which integrates SNP thresholds with phylogenetic structure, tends to expand rather than contract transmission clusters. The phylogenetic correction only adds members missed by the clustering algorithm rather than removing them, potentially overestimating cluster sizes. However, our within-cluster SNP distances align with other studies.

Since most screening swabs were collected from nasal sites, the available data to assess the correlation between colonized body sites and infection risk is limited.

#### Supplementary figures and tables

| Group | Number of Genomes | Bootstrap Support | SH-aLRT Support | CC | Threshold |
|---|---|---|---|---|---|
| 1 | 1 | N/A | N/A | Unassigned | N/A |
| 2 | 163 | 100 | 97 | CC5 | 34 |
| 3 | 311 | 100 | 100 | CC398 | 72 |
| 4 | 3 | 100 | 100 | Unassigned | 15 |
| 5 | 154 | 100 | 100 | CC30 | 40 |
| 6 | 104 | 100 | 100 | CC45 | 19 |
| 7 | 4 | 100 | 100 | CC45 | 8 |
| 8 | 67 | 100 | 100 | Unassigned | 19 |
| 9 | 22 | 100 | 100 | CC121 | 31 |
| 10 | 9 | 100 | 100 | CC22 | 19 |
| 11 | 6 | 100 | 100 | Unassigned | 10 |
| 12 | 3 | 100 | 100 | Unassigned | 11 |
| 13 | 245 | 100 | 100 | CC8 | 32 |
| 14 | 45 | 100 | 100 | CC97 | 15 |
| 15 | 83 | 100 | 100 | CC15 | 56 |
| 16 | 3 | 100 | 100 | CC1 | 14 |
| 17 | 14 | 100 | 100 | CC1 | 18 |
| 18 | 35 | 100 | 100 | CC1 | 21 |
| 19 | 1 | N/A | N/A | CC1 | N/A |
| 20 | 30 | 100 | 100 | Unassigned | 9 |
| 21 | 4 | 100 | 100 | Unassigned | 6 |
| 22 | 46 | 100 | 100 | CC5 | 22 |
| 23 | 11 | 100 | 100 | CC8 | 12 |
| 24 | 73 | 100 | 100 | CC8 | 26 |
| 25 | 2 | 100 | 100 | Unassigned | 6 |
| 26 | 1 | N/A | N/A | Unassigned | N/A |
| 27 | 1 | N/A | N/A | Unassigned | N/A |
| 28 | 5 | 100 | 100 | CC1 | 3 |

Table S1: Phylogenetically corrected SNP Thresholds for Hierarchical Groups

This table summarizes the hierarchical clustering groups. The table provides the number of genomes, bootstrap and SH-aLRT supports for the grouping, clonal complex assignment, and the SNP thresholds. Groups with only one genome lack bootstrap and SH-aLRT supports (N/A), as these require at least 2 genomes to form a clade in the phylogeny. Nine groups were designated as "Unassigned" as they were not assigned any sequence types.

| Cluster | Invasive Status | Observed Persistence (Days) | Molecular Clock Persistence (Days) |
|---|---|---|---|
| 1 | Invasive | 975 | 1201 |
| 2 | Invasive | 708 | 998 |
| 3 | Colonizing | 944 | 1710 |
| 4 | Invasive | 583 | 879 |
| 5 | Invasive | 291 | 542 |
| 6 | Invasive | 848 | 2905 |
| 7 | Invasive | 449 | 610 |
| 8 | Colonizing | 947 | 1526 |
| 9 | Colonizing | 399 | 449 |
| 10 | Invasive | 840 | 1854 |
| 11 | Colonizing | 149 | 267 |
| 12 | Colonizing | 645 | 776 |
| 13 | Invasive | 449 | 661 |
| 14 | Colonizing | 358 | 393 |
| 15 | Colonizing | 949 | 1099 |
| 16 | Colonizing | 903 | 1045 |
| 17 | Invasive | 421 | 497 |
| 18 | Colonizing | 406 | 957 |
| 19 | Invasive | 557 | 909 |
| 20 | Colonizing | 155 | 159 |
| 21 | Colonizing | 99 | 183 |
| 22 | Colonizing | 309 | 373 |
| 23 | Colonizing | 603 | 1501 |
| 24 | Colonizing | 128 | 157 |
| 25 | Colonizing | 406 | 466 |
| 26 | Colonizing | 29 | 46 |
| 28 | Colonizing | 91 | 250 |
| 29 | Invasive | 422 | 535 |
| 30 | Invasive | 360 | 465 |
| 31 | Colonizing | 428 | 479 |
| 32 | Colonizing | 352 | 507 |
| 33 | Colonizing | 128 | 189 |
| 34 | Invasive | 27 | 80 |

| | | | |
|---|---|---|---|
| 35 | Colonizing | 22 | 143 |
| 37 | Invasive | 191 | 238 |
| 38 | Colonizing | 29 | 345 |
| 46 | Colonizing | 35 | 163 |
| 48 | Colonizing | 120 | 215 |
| 49 | Colonizing | 128 | 189 |
| 51 | Invasive | 216 | 251 |
| 56 | Colonizing | 495 | 673 |
| 57 | Colonizing | 379 | 430 |
| 58 | Invasive | 231 | 902 |
| 59 | Colonizing | 190 | 234 |
| 63 | Colonizing | 29 | 220 |
| 65 | Invasive | 69 | 161 |
| 67 | Colonizing | 595 | 602 |
| 68 | Invasive | 208 | 214 |

Table S2: Characteristics of 69 Identified Transmission Clusters
This table presents invasive status and temporal metrics for identified transmission clusters. For each cluster, the table reports invasive status, earliest and most recent detection dates, time to most recent common ancestor (tMRCA), observed persistence period in days, and molecular clock-estimated persistence in days.

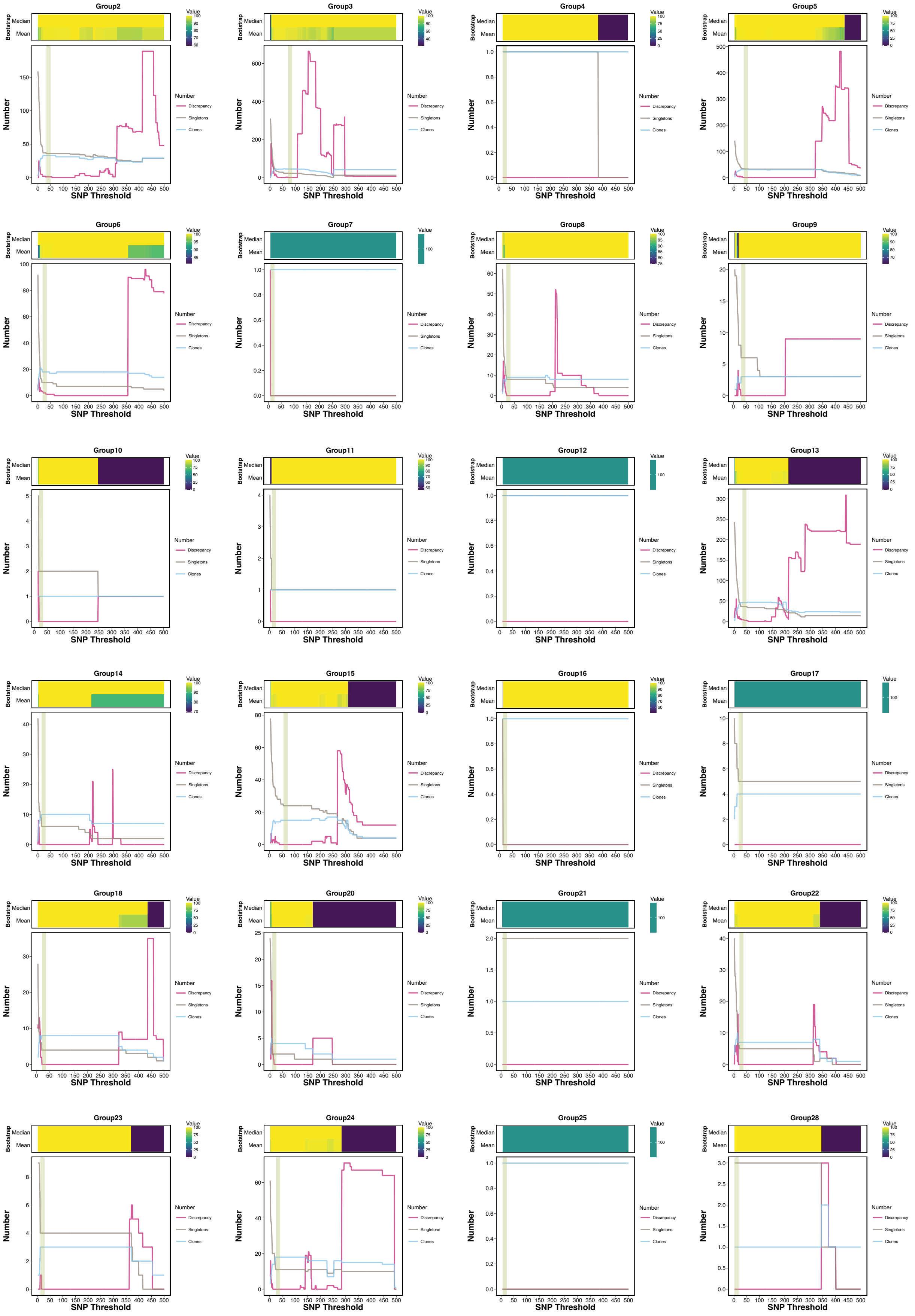

Figure S1: Multi-SNP Threshold Plots for Hierarchical Groups containing at Least Two Genomes

For each plot, the x-axis represents tested SNP threshold values. The upper panel displays average and median bootstrap support values for clones identified at each threshold. The lower panel quantifies discrepancy genomes (purple), clones (blue), and singletons (gray) across the tested threshold range. The green bar indicates the plateau where the strain compositions were finalized.

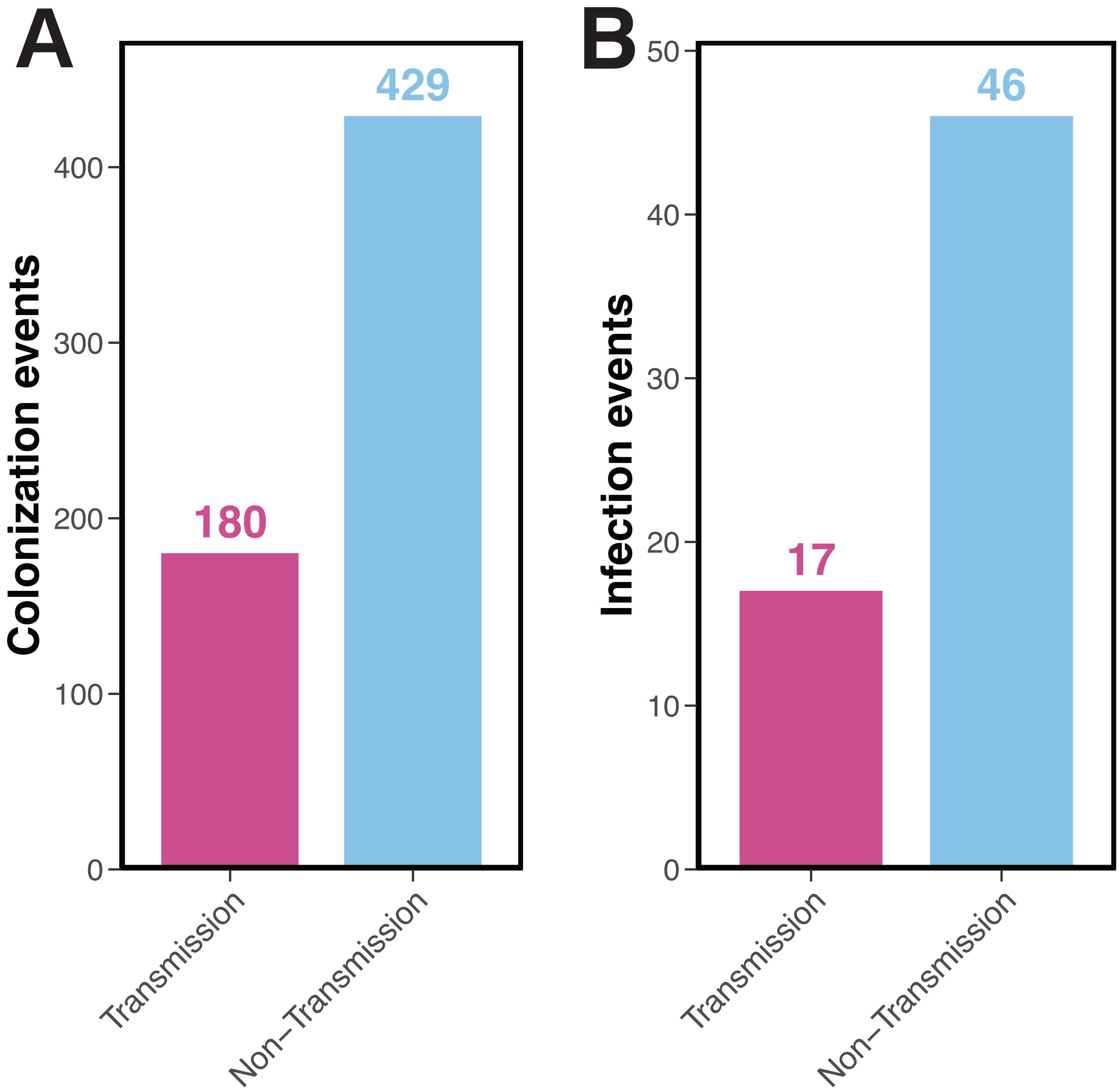

Figure S2: Distribution of Transmission-acquired Colonization and Infection Events

(A) Distribution of colonization events in the NICU (n=609), showing 180 cases (29·56%) identified as transmission-acquired after excluding index patients. (B) Distribution of infection events across CHOP departments (n=63), with 17 cases (26·98%) identified as transmission-acquired.

Figure S3: NICU Floorplans Demonstrating Patient Movements in the Identified Clusters

Visualization of patient movements across the NICU for each identified transmission cluster. Patient locations are mapped chronologically, with connecting lines indicating the patient movements.

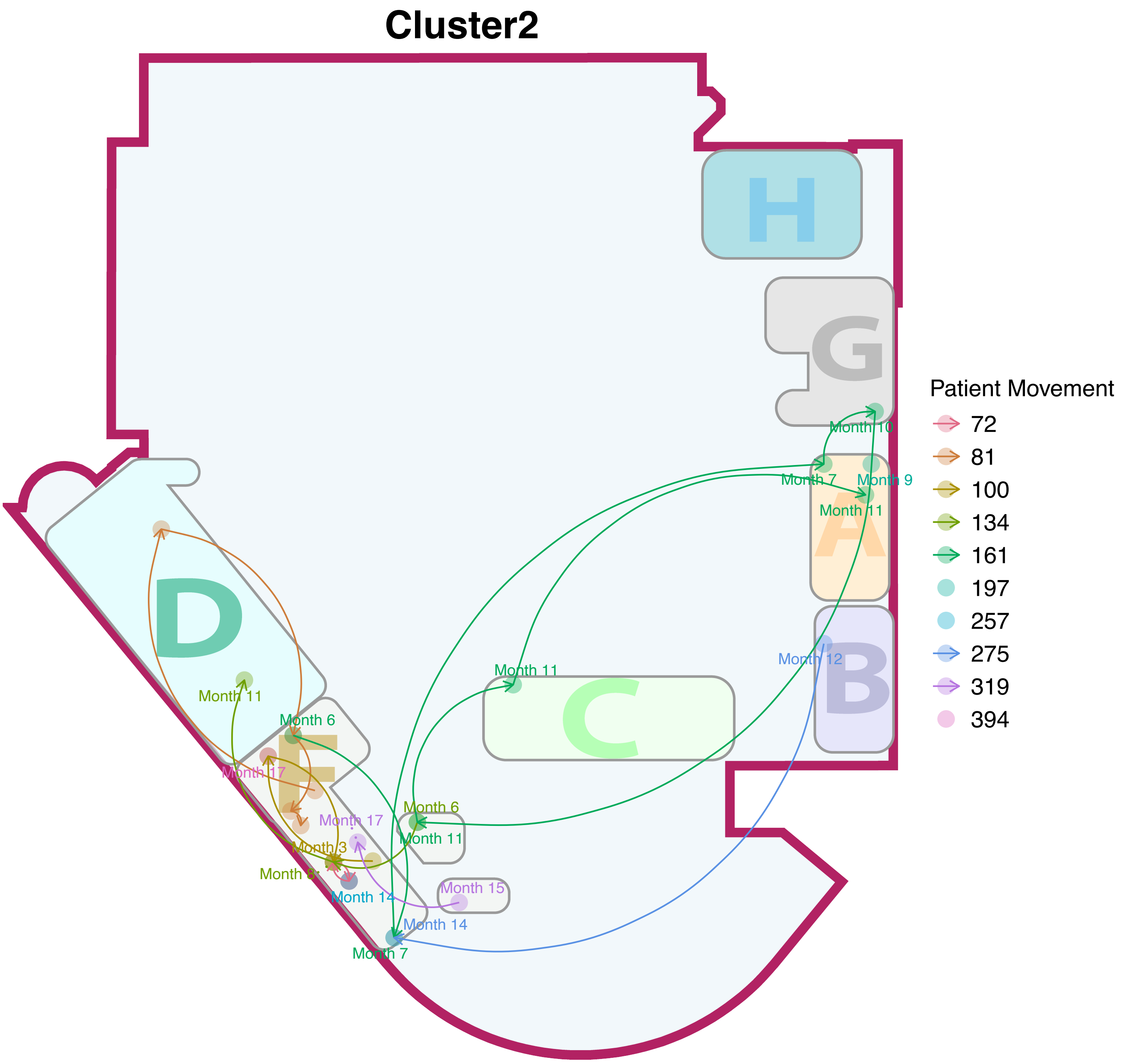

#### Cluster3

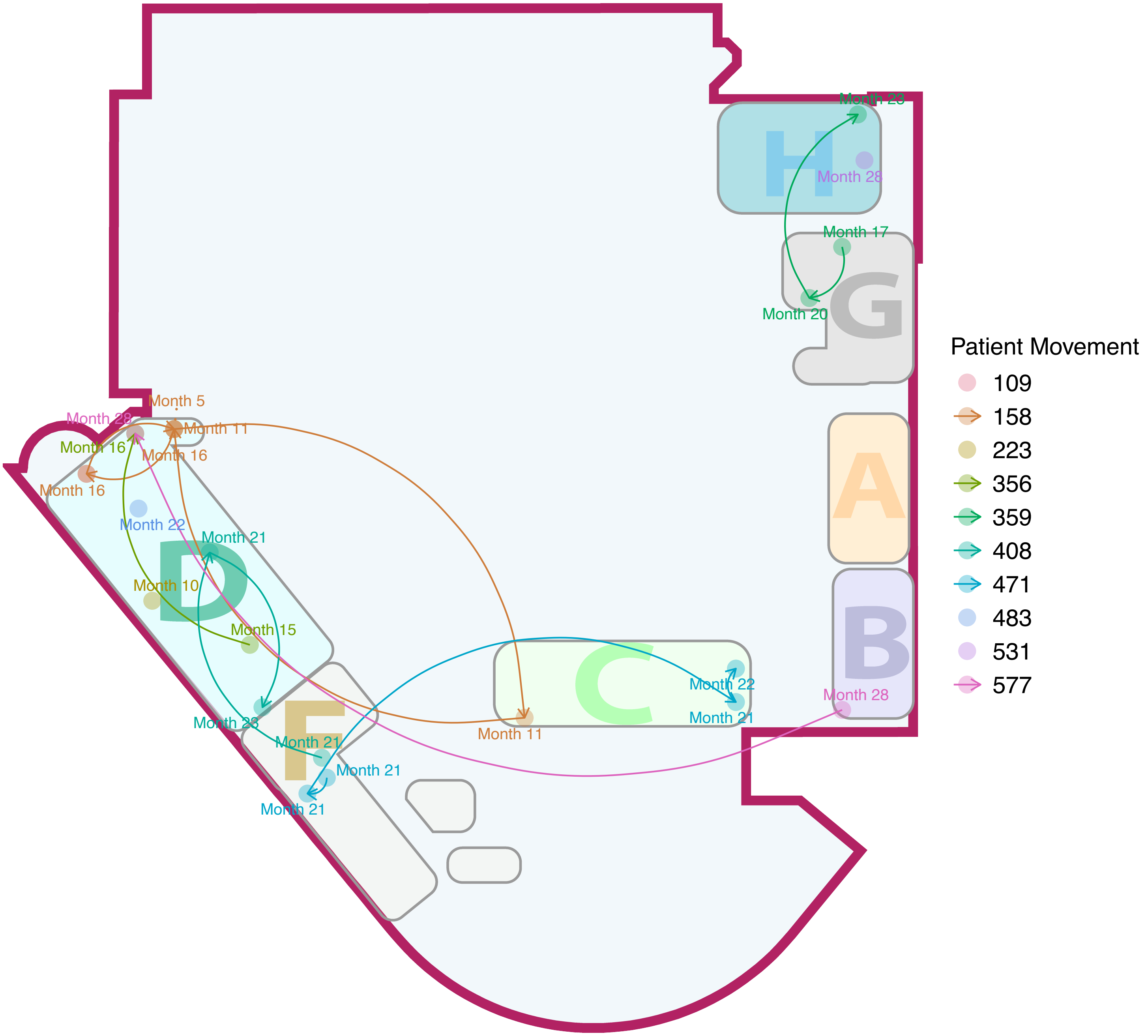
Month 23
Month 28
Month 17
Month 20
Month 5
Month 28
Month 11
Month 16
Month 16
Month 16
Month 22
Month 21
Month 10
Month 15
Month 22
Month 21
Month 23
Month 11
Month 28
Month 21
Month 21
Month 21
H
G
A
B
C
D
F
Patient Movement
109
158
223
356
359
408
471
483
531
577

### Cluster4

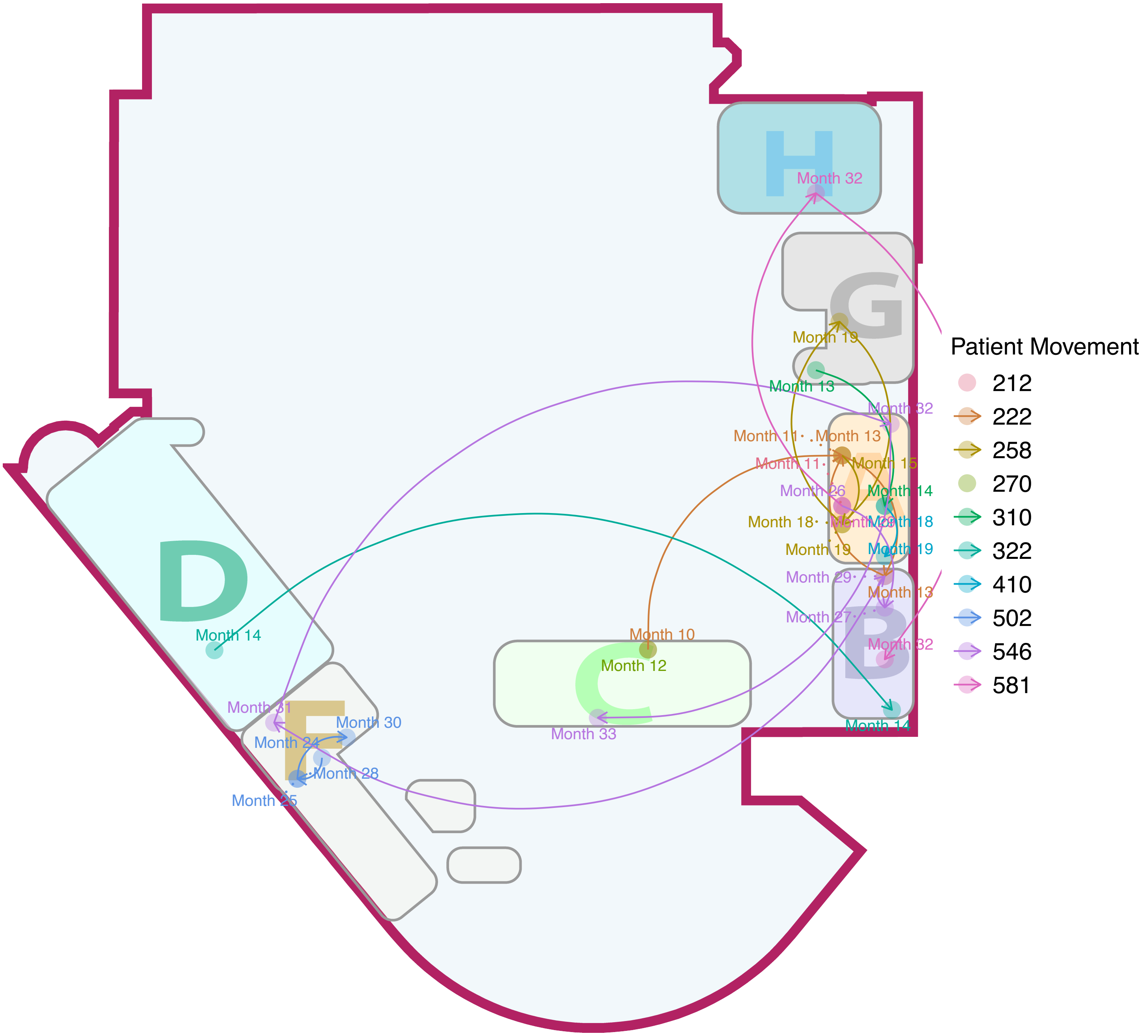
H
Month 32
G
Month 19
Month 13
Month 32
Month 11
Month 13
Month 15
Month 26
Month 14
Month 18
Month 19
Month 29
Month 27
B
Month 32
Month 14
D
Month 14
Month 10
C
Month 12
Month 33
Month 31
Month 30
Month 24
Month 28
Month 25
F
Patient Movement
212
222
258
270
310
322
410
502
546
581

### Cluster5

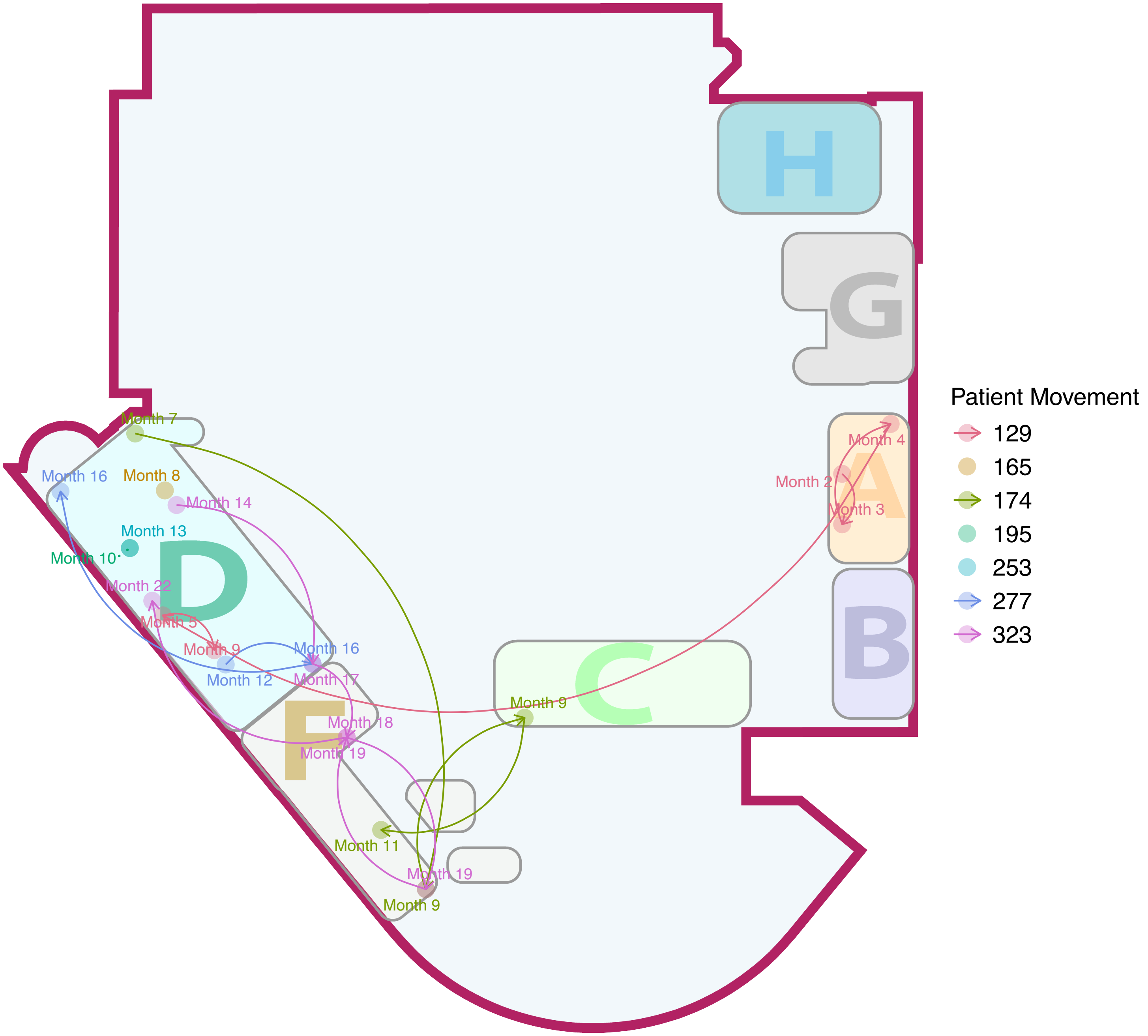

H
G
A
B
C
D
F
Month 4
Month 2
Month 3
Month 7
Month 16
Month 8
Month 14
Month 13
Month 10
Month 22
Month 5
Month 9
Month 16
Month 12
Month 17
Month 18
Month 19
Month 9
Month 11
Month 19
Month 9
Patient Movement
129
165
174
195
253
277
323

### Cluster6

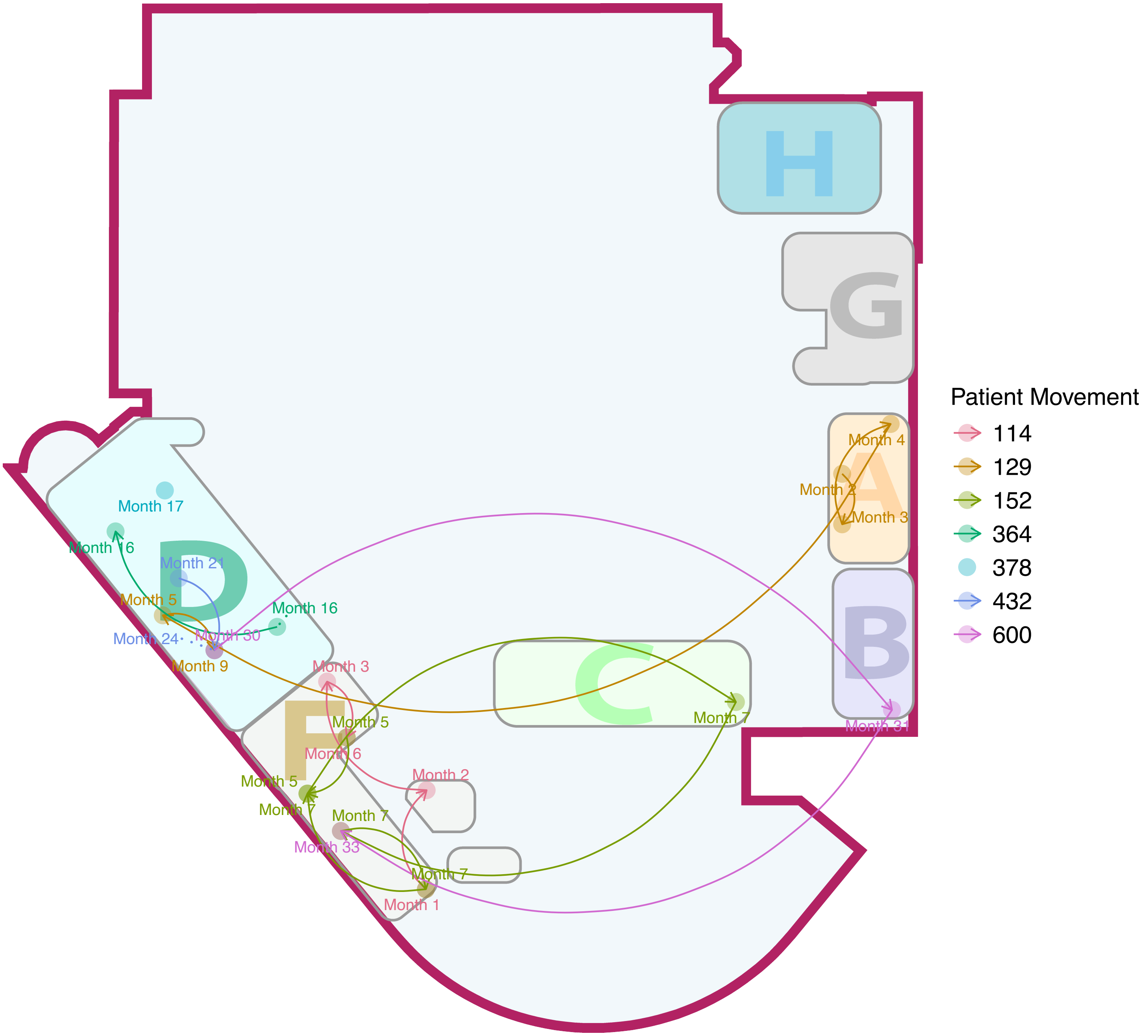

H
G
A
B
C
D
F
Month 17
Month 16
Month 21
Month 5
Month 16
Month 24
Month 30
Month 9
Month 3
Month 5
Month 6
Month 5
Month 7
Month 7
Month 33
Month 2
Month 7
Month 1
Month 7
Month 4
Month 2
Month 3
Month 31
Patient Movement
114
129
152
364
378
432
600

### Cluster7

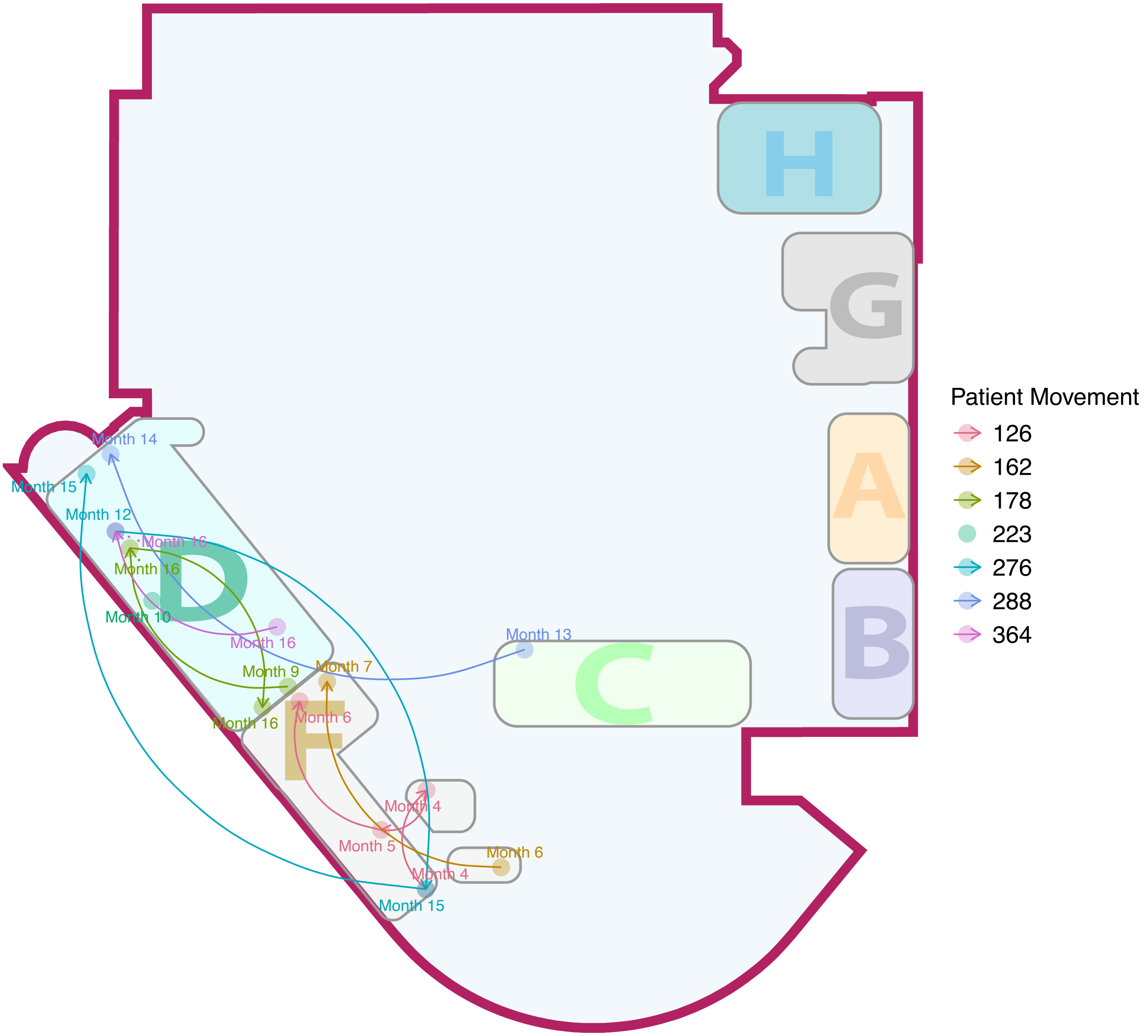

H
G
A
B
C
D
F
Month 14
Month 15
Month 12
Month 16
Month 16
Month 10
Month 16
Month 13
Month 9
Month 7
Month 16
Month 6
Month 4
Month 5
Month 6
Month 4
Month 15
Patient Movement
126
162
178
223
276
288
364

### Cluster8

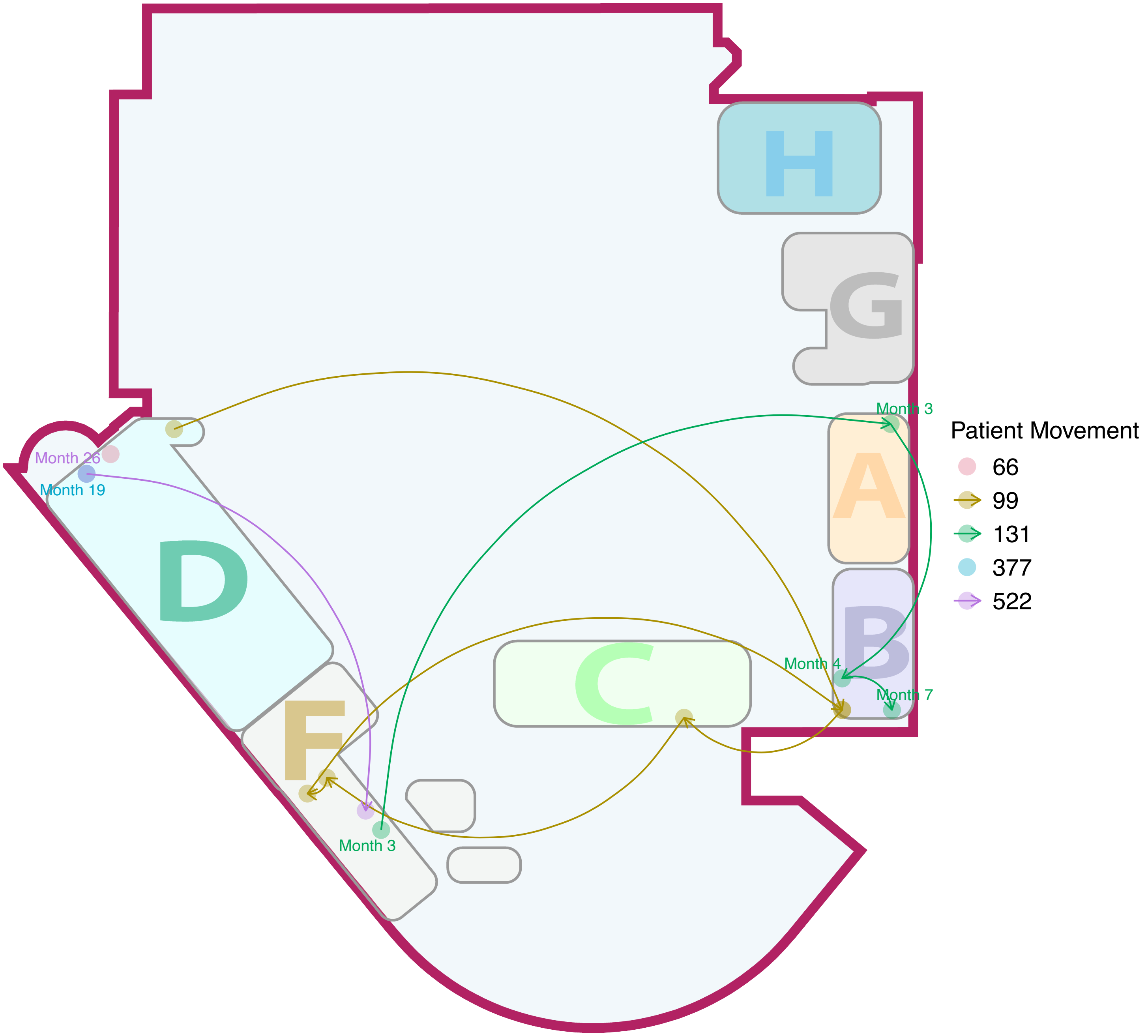

H
G
Month 3
A
Patient Movement
66
99
131
377
522
Month 26
Month 19
D
B
Month 4
Month 7
C
F
Month 3

### Cluster9

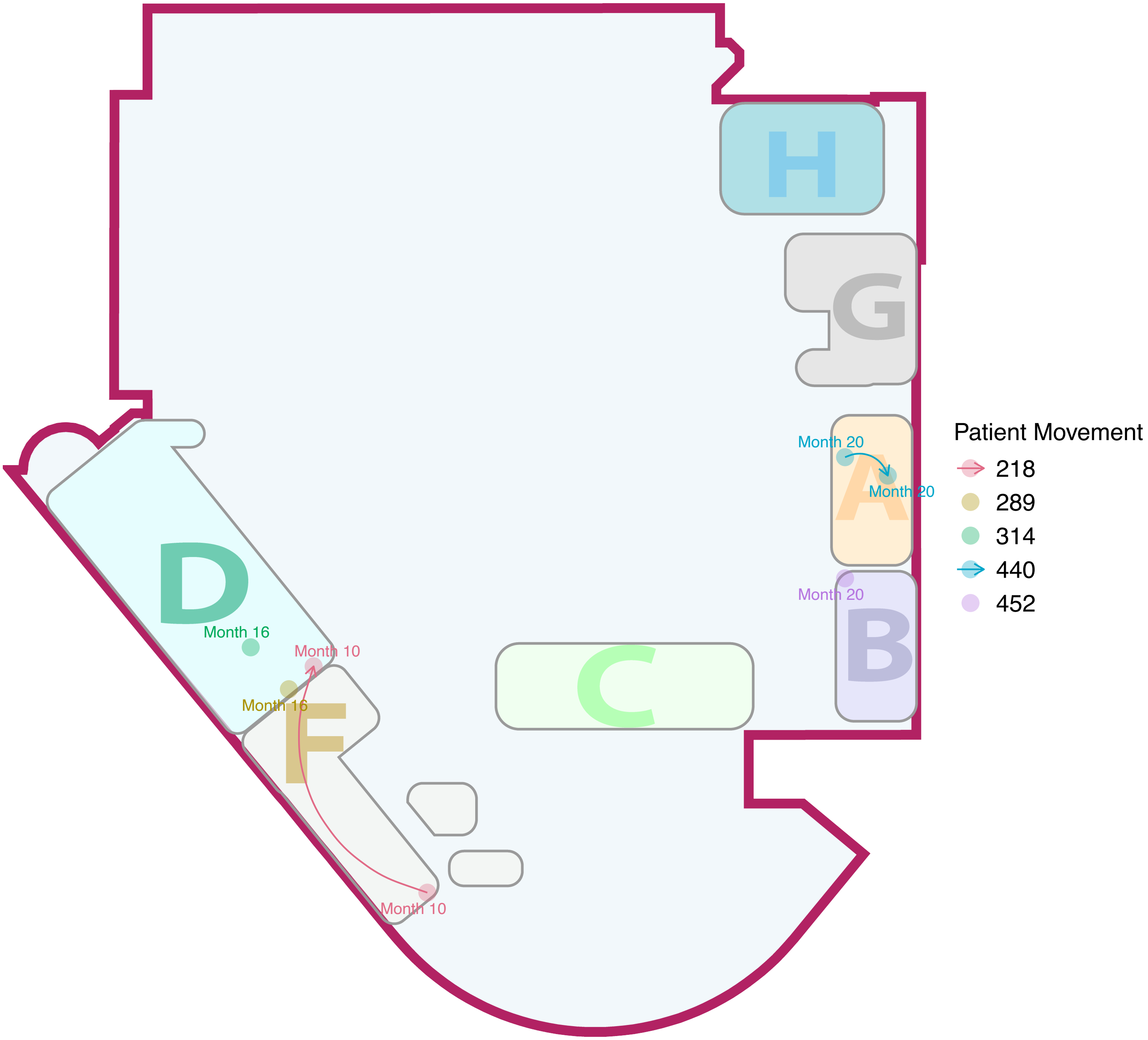

H
G
Month 20
A
Month 20
Month 20
B
D
Month 16
Month 10
Month 16
F
C
Month 10
Patient Movement
218
289
314
440
452

### Cluster10

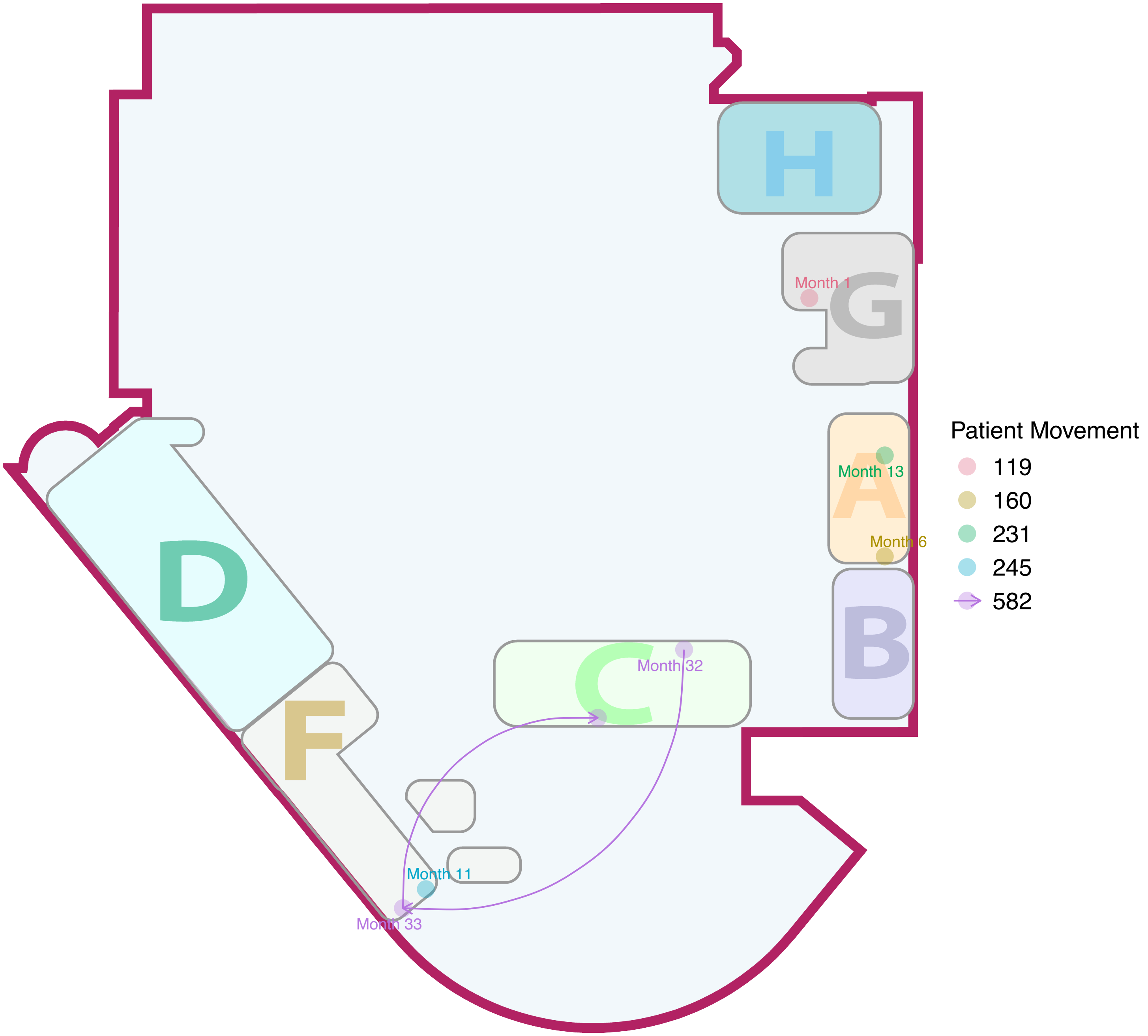

H
G
Month 1
A
Month 13
Month 6
B
D
C
Month 32
F
Month 11
Month 33
Patient Movement
119
160
231
245
582

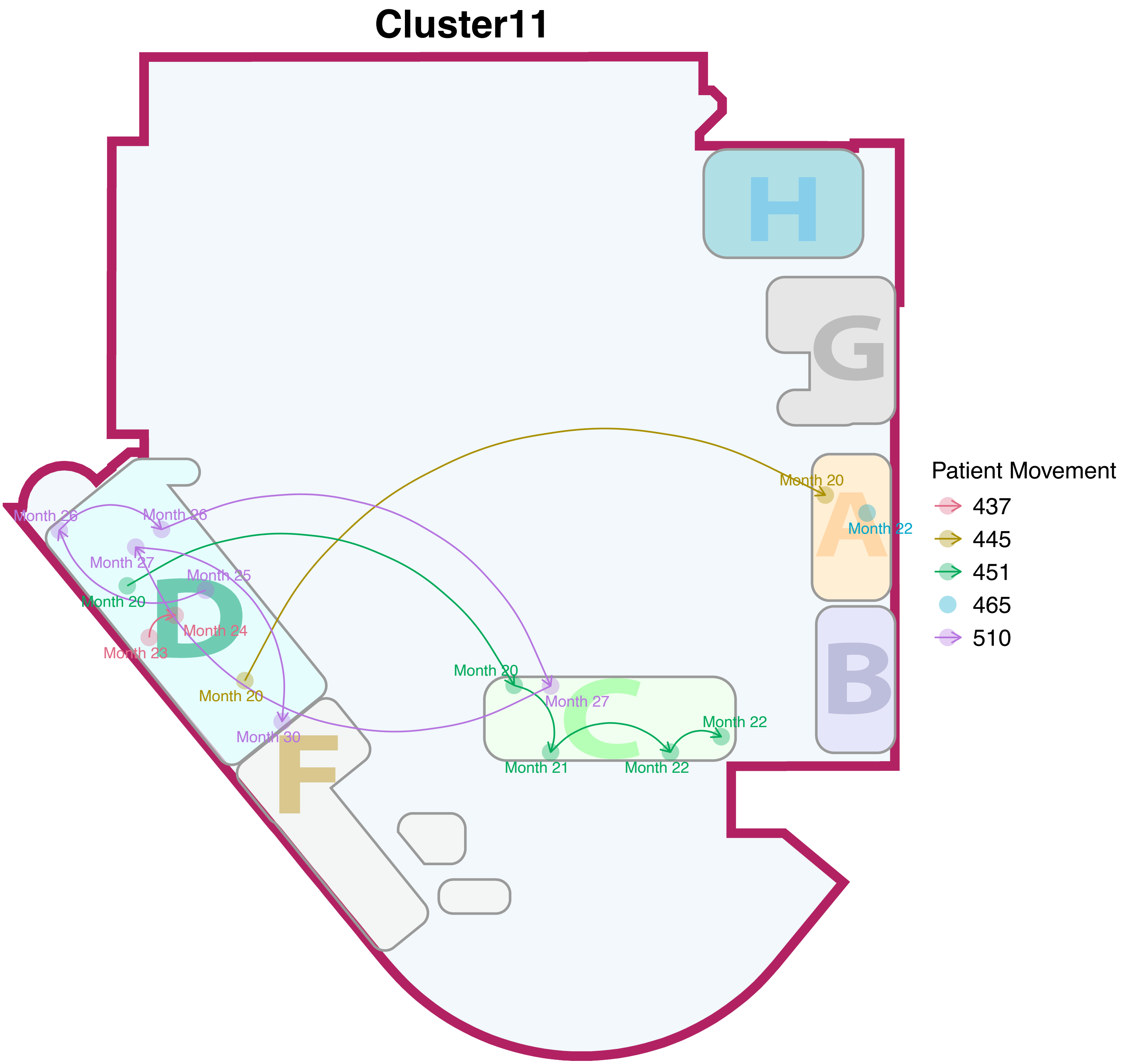
Cluster11
H
G
A
B
C
D
F
Patient Movement
437
445
451
465
510
Month 20
Month 22
Month 26
Month 27
Month 25
Month 24
Month 23
Month 30
Month 21

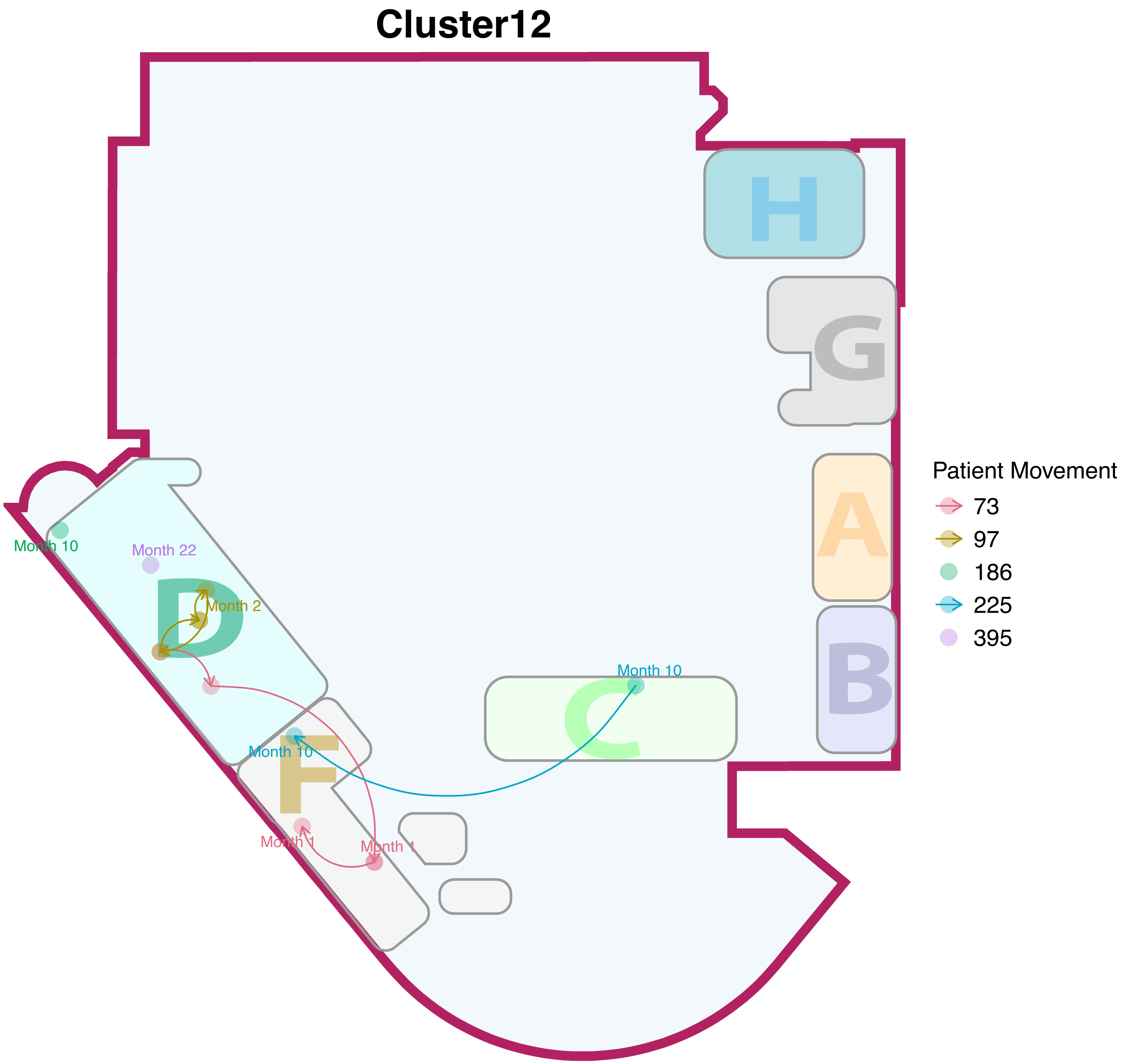

Cluster12
H
G
A
B
C
D
F
Month 10
Month 22
Month 2
Month 10
Month 10
Month 1
Month 1
Patient Movement
73
97
186
225
395

### Cluster13

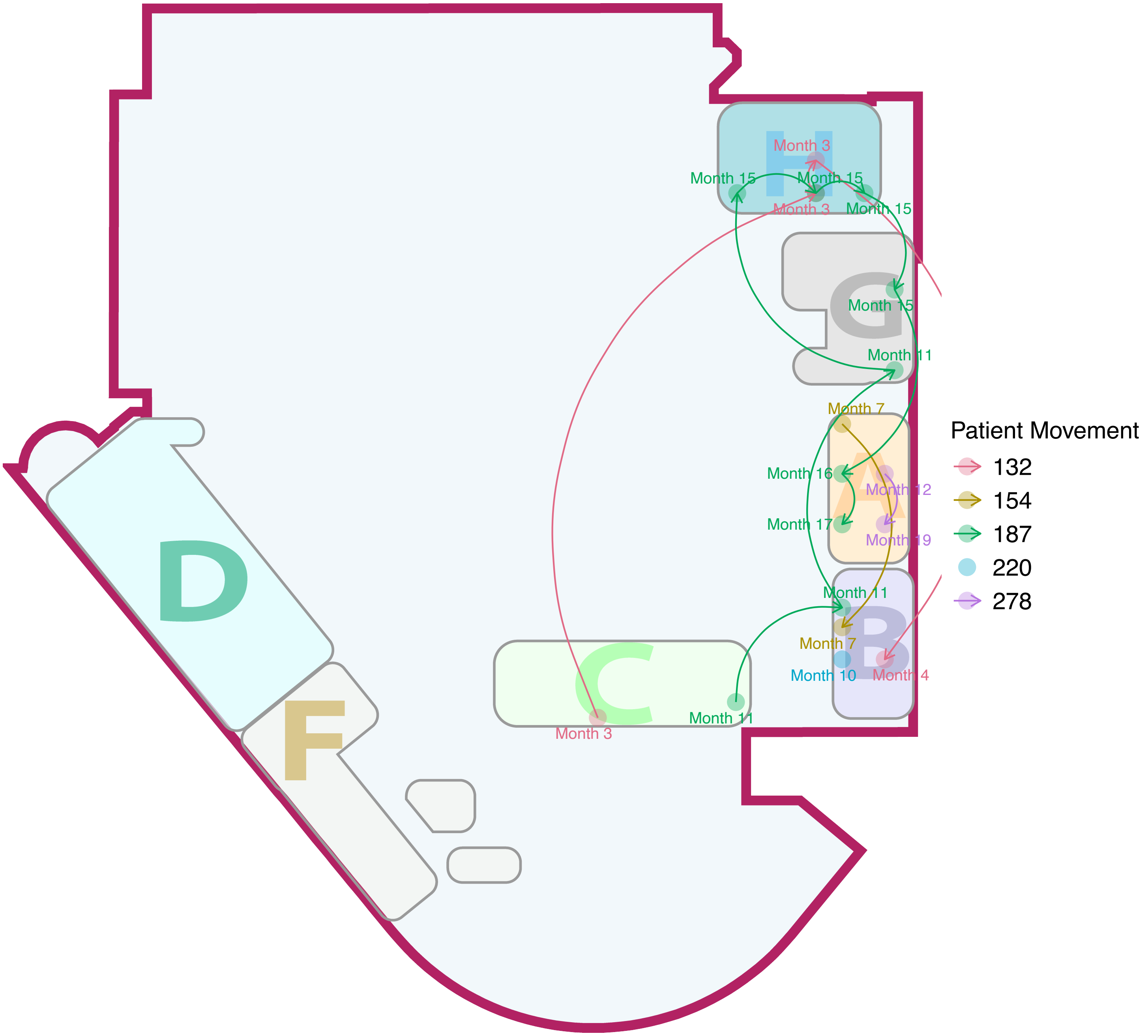

H
G
A
B
C
D
F
Month 3
Month 15
Month 15
Month 3
Month 15
Month 15
Month 11
Month 7
Month 16
Month 12
Month 17
Month 19
Month 11
Month 7
Month 10
Month 4
Month 11
Month 3
Patient Movement
132
154
187
220
278

### Cluster14

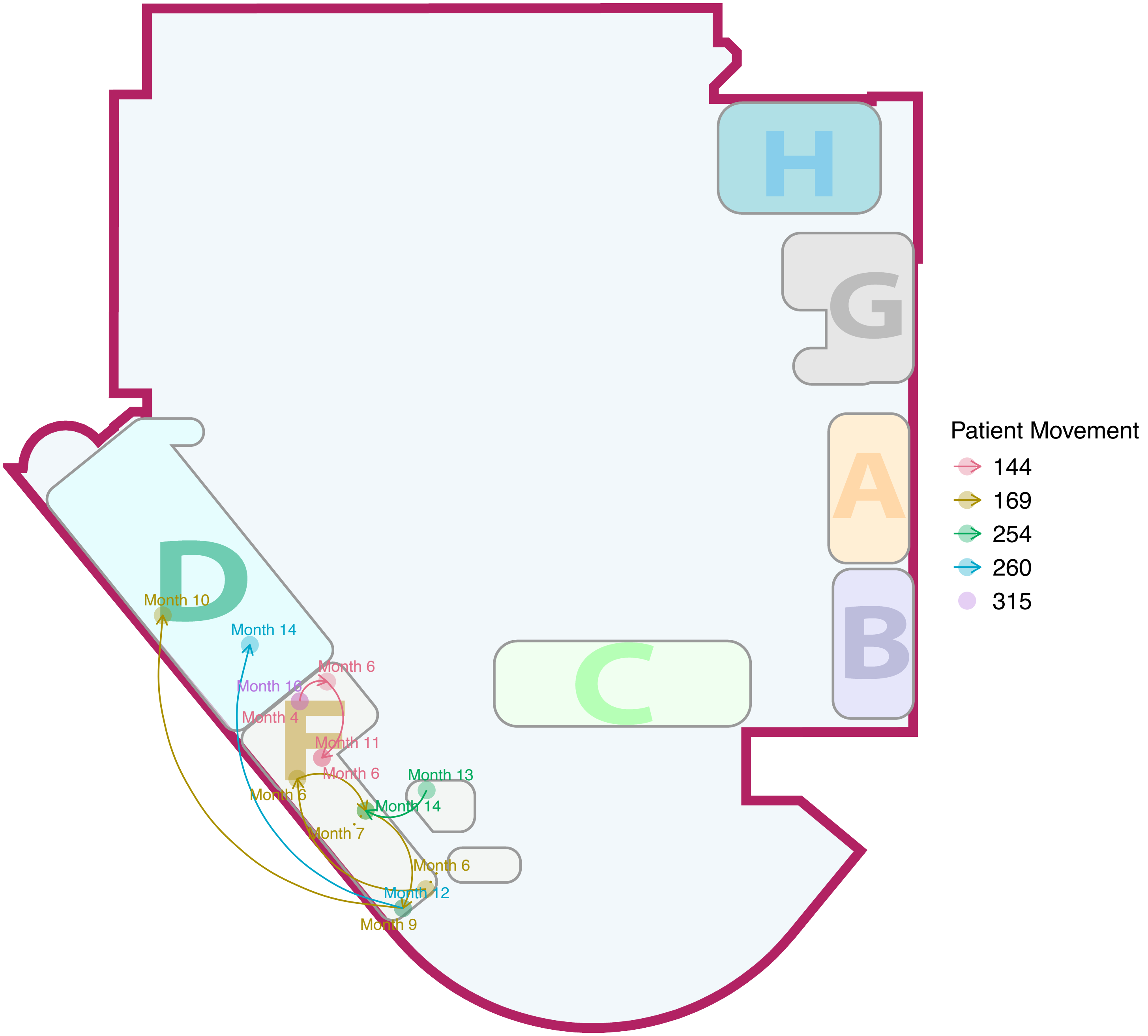

H
G
A
B
C
D
F
Month 10
Month 14
Month 6
Month 16
Month 4
Month 11
Month 6
Month 13
Month 6
Month 14
Month 7
Month 6
Month 12
Month 9
Patient Movement
144
169
254
260
315

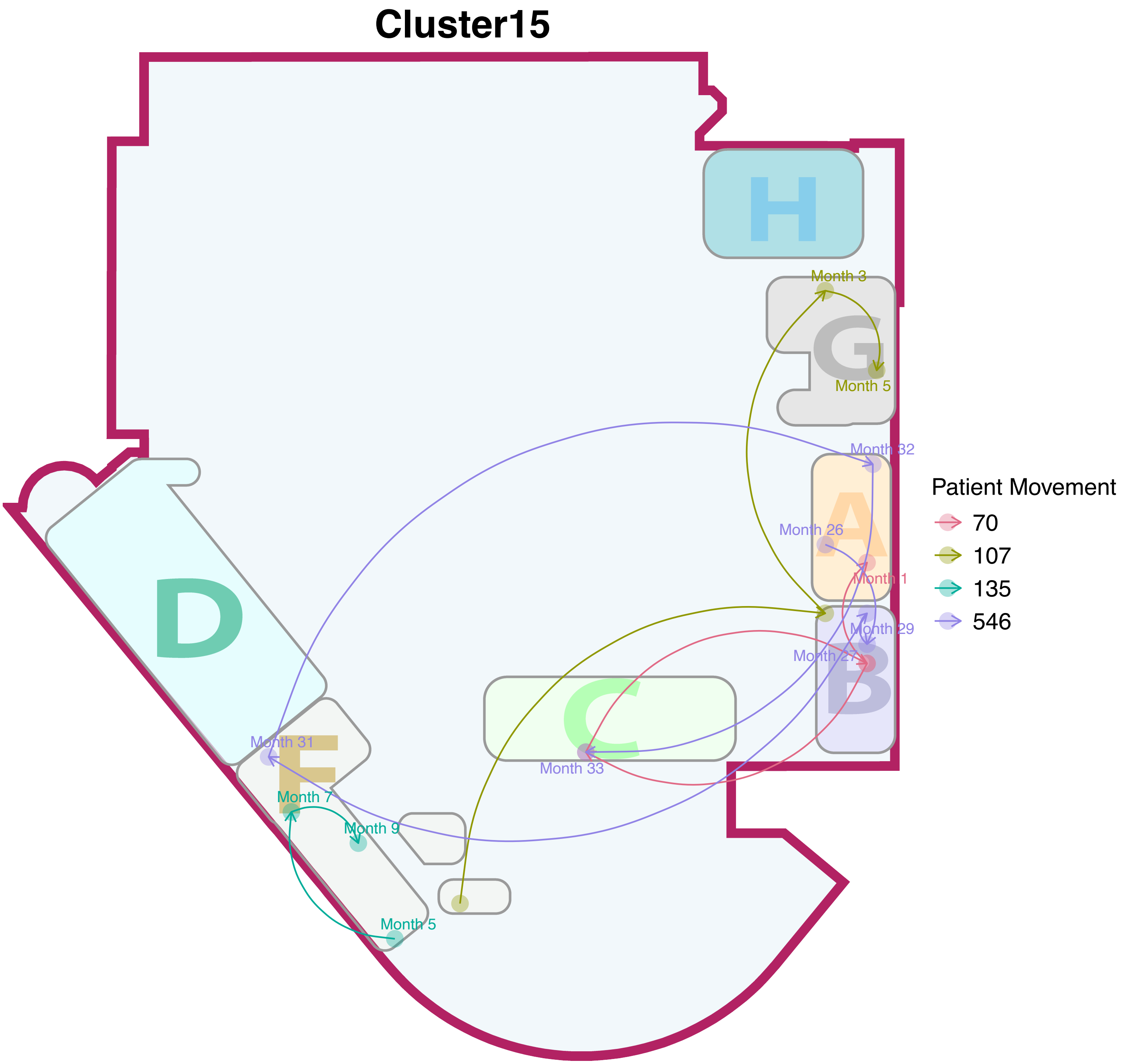

Cluster15
H
G
Month 3
Month 5
Month 32
A
Month 26
Month 1
Month 29
Month 27
B
D
C
Month 31
Month 33
F
Month 7
Month 9
Month 5
Patient Movement
70
107
135
546

### Cluster16

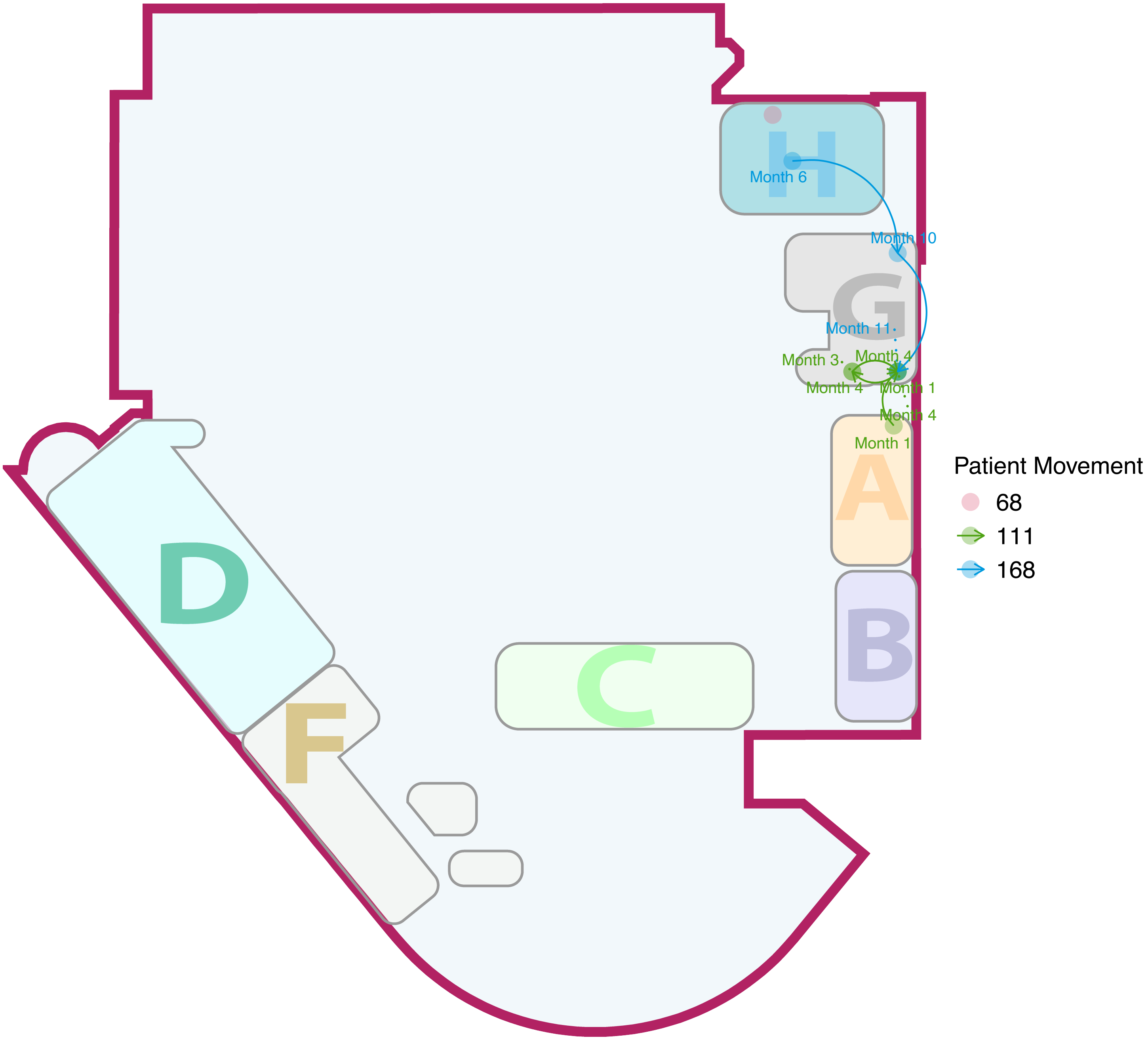
H
Month 6
Month 10
G
Month 11
Month 3
Month 4
Month 4
Month 1
Month 4
Month 1
A
B
C
D
F
Patient Movement
68
111
168

#### Cluster17

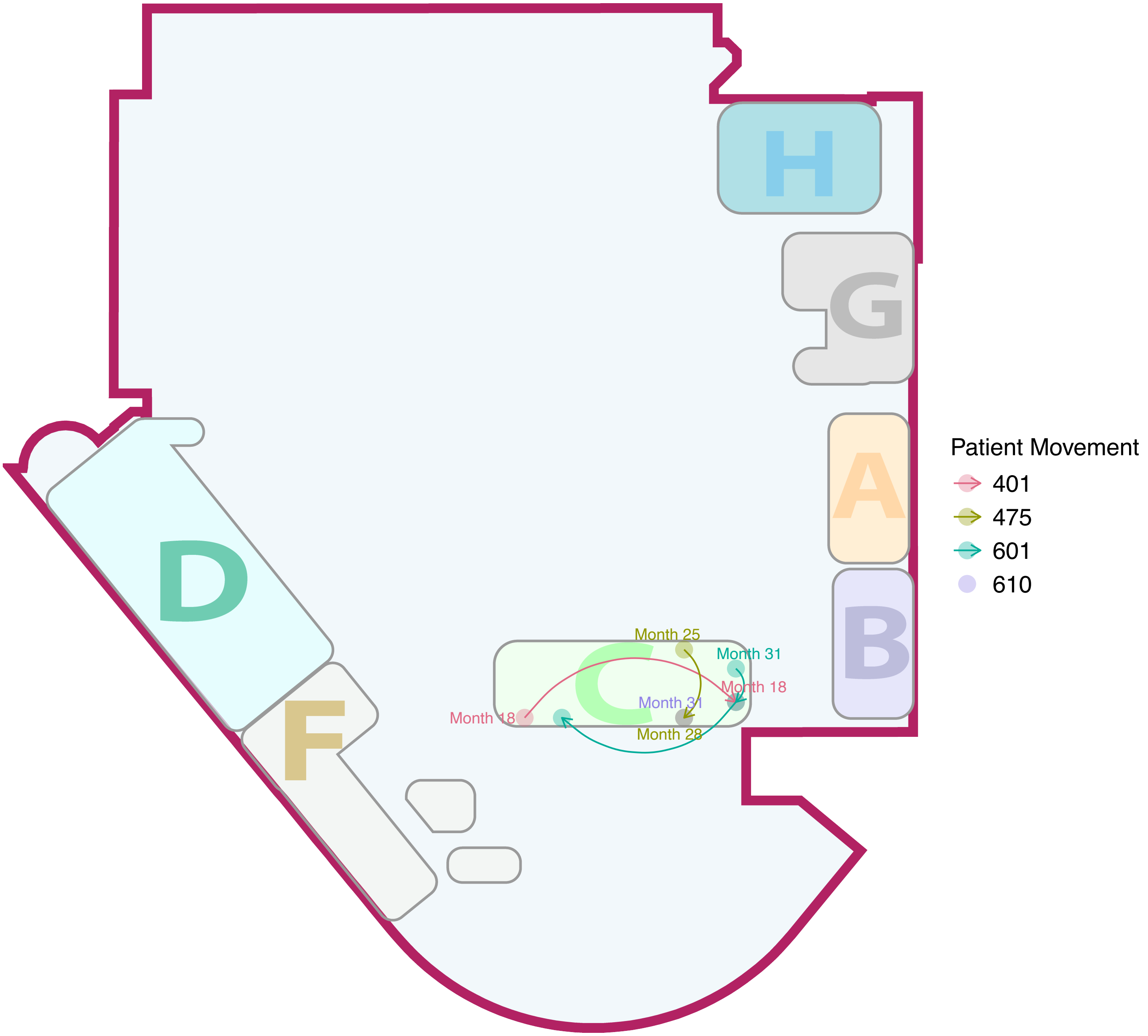

H
G
A
B
D
C
F
Month 25
Month 31
Month 18
Month 31
Month 18
Month 28
Patient Movement
401
475
601
610

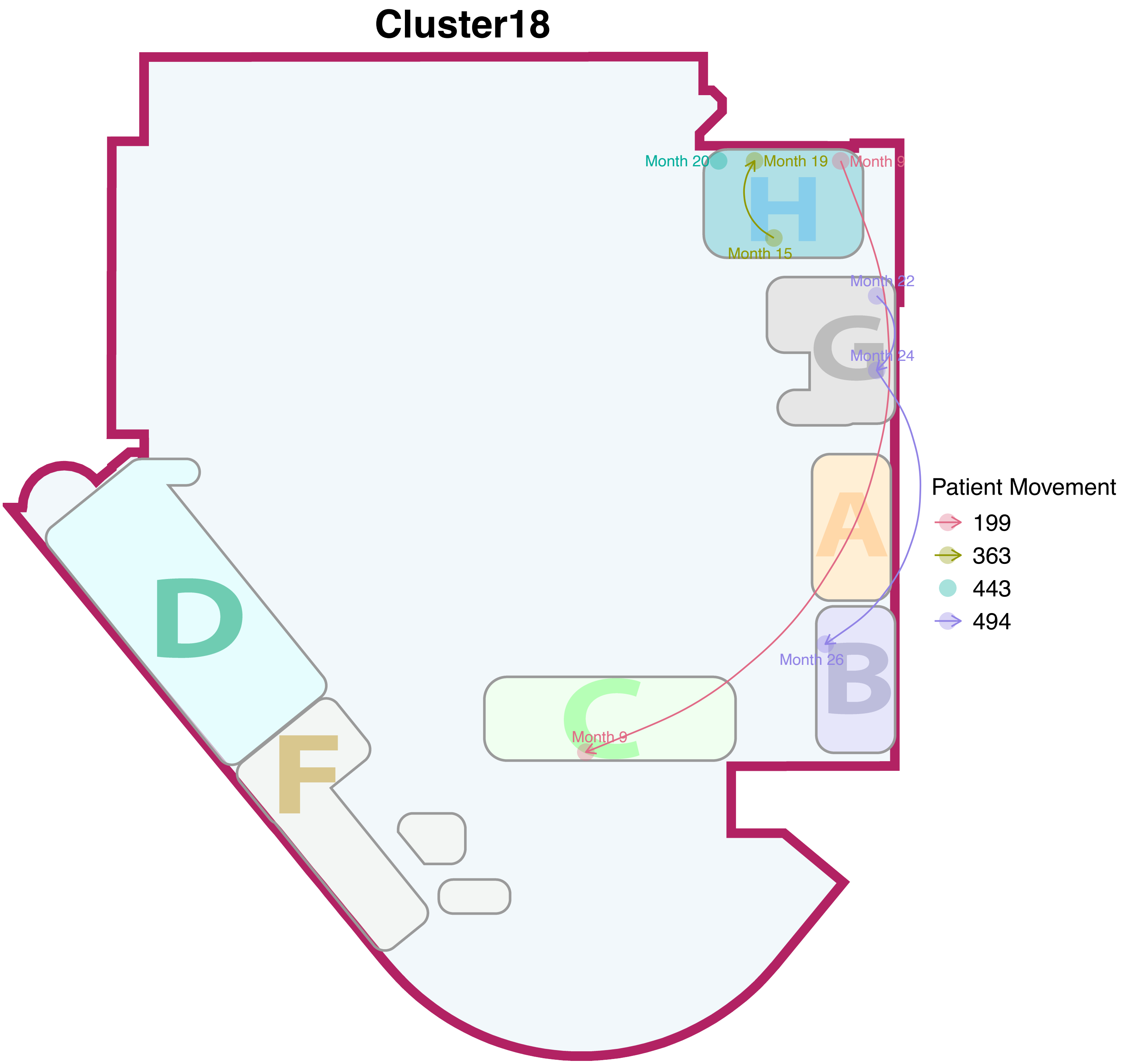

Cluster18
Month 20
Month 19
Month 9
Month 15
Month 22
Month 24
Month 26
Month 9
H
G
A
B
C
D
F
Patient Movement
199
363
443
494

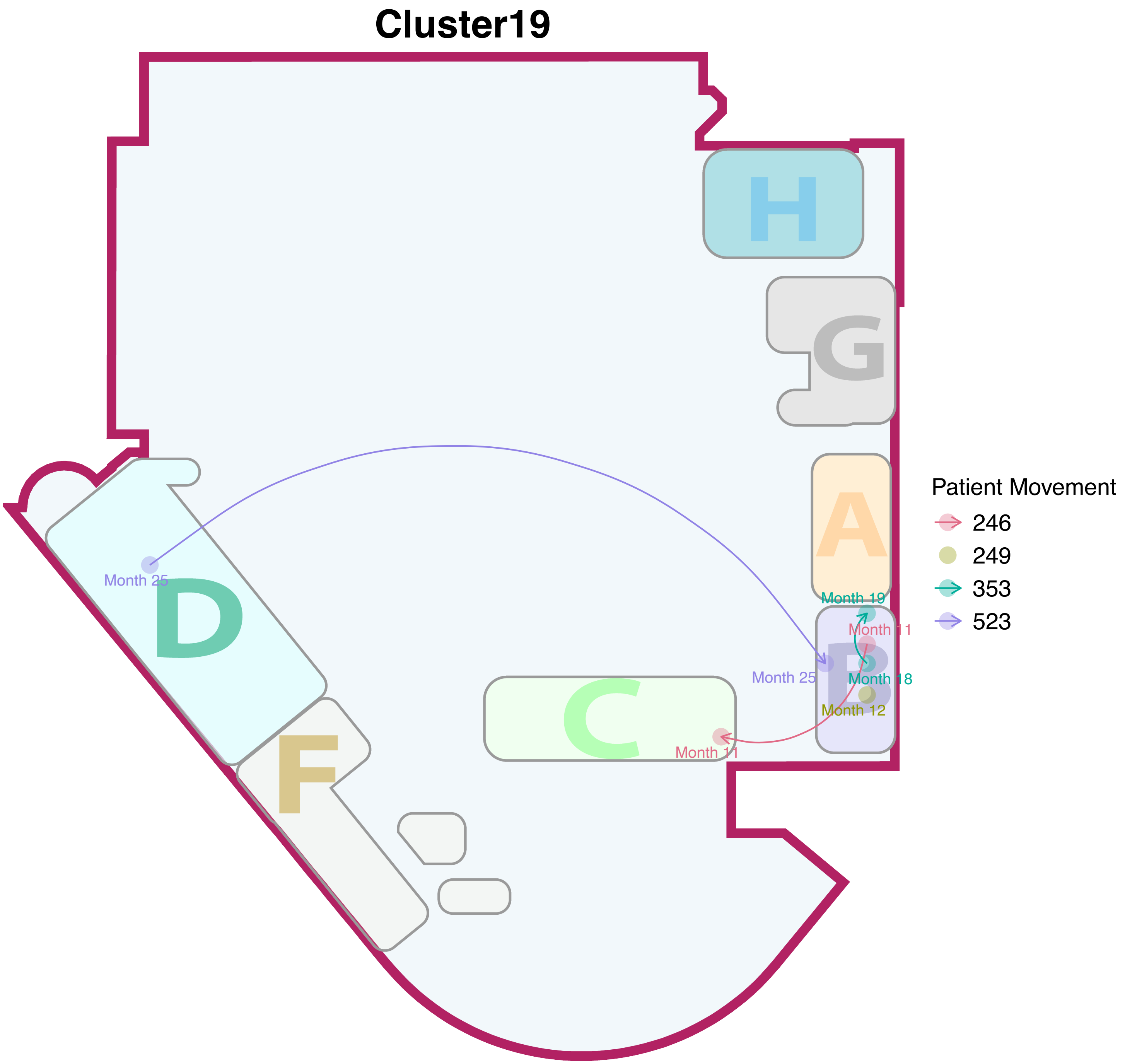
Cluster19
H
G
A
D
C
F
B
Month 25
Month 19
Month 11
Month 25
Month 18
Month 12
Month 11
Patient Movement
246
249
353
523

### Cluster20

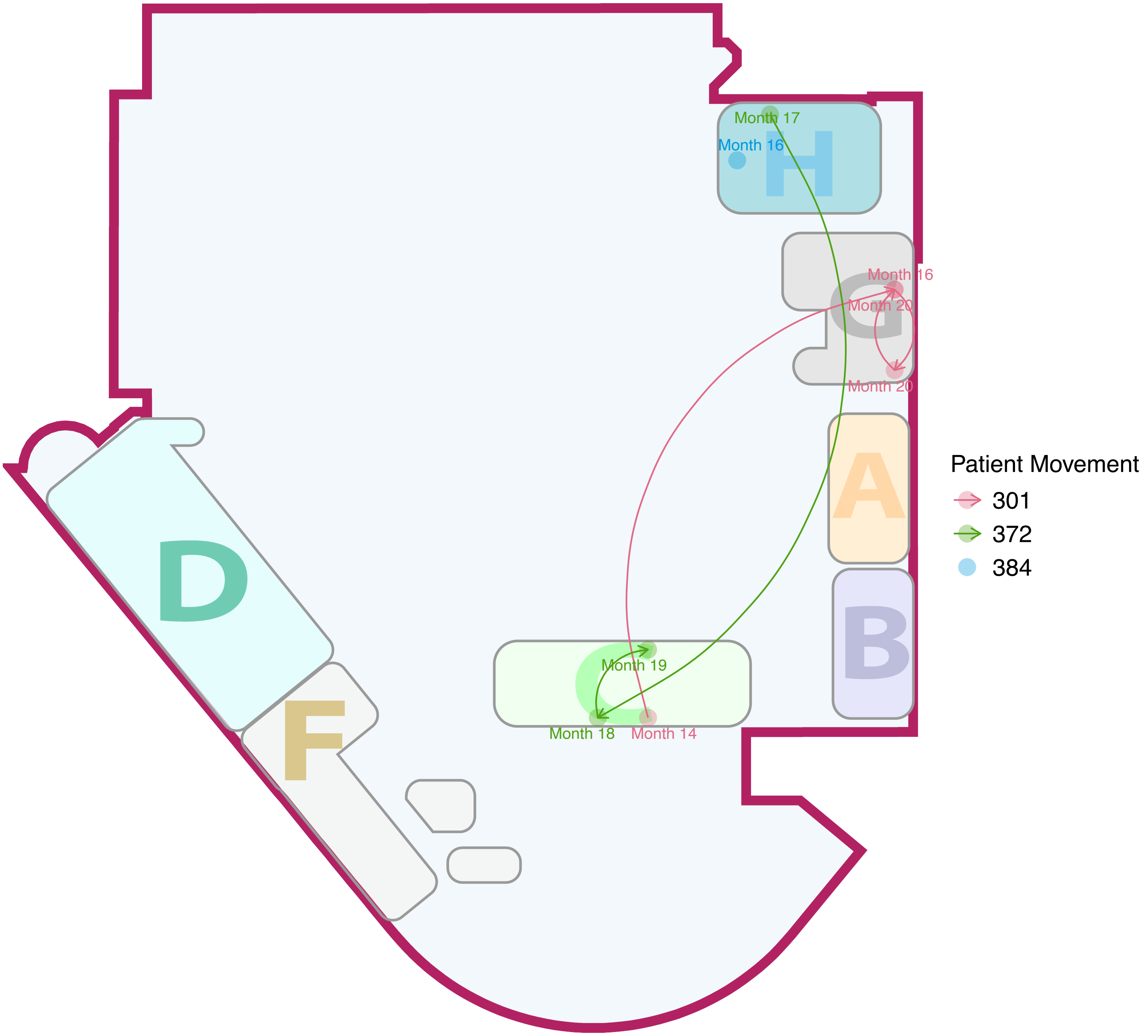

Month 17
Month 16
H
Month 16
Month 20
G
Month 20
A
B
D
F
Month 19
Month 18
Month 14
Patient Movement
301
372
384

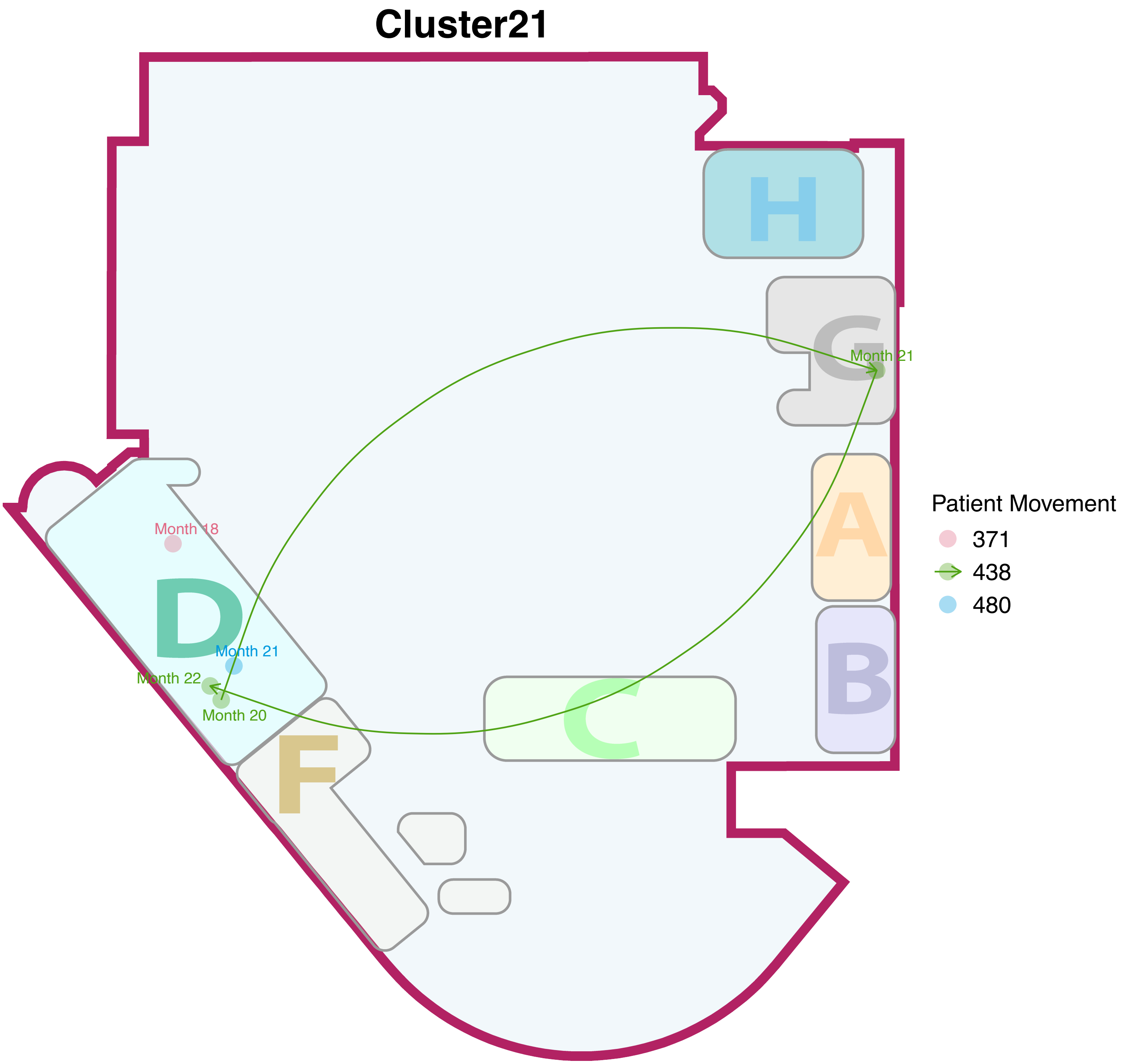
Cluster21
H
G
Month 21
A
B
C
D
F
Month 18
Month 21
Month 22
Month 20
Patient Movement
371
438
480

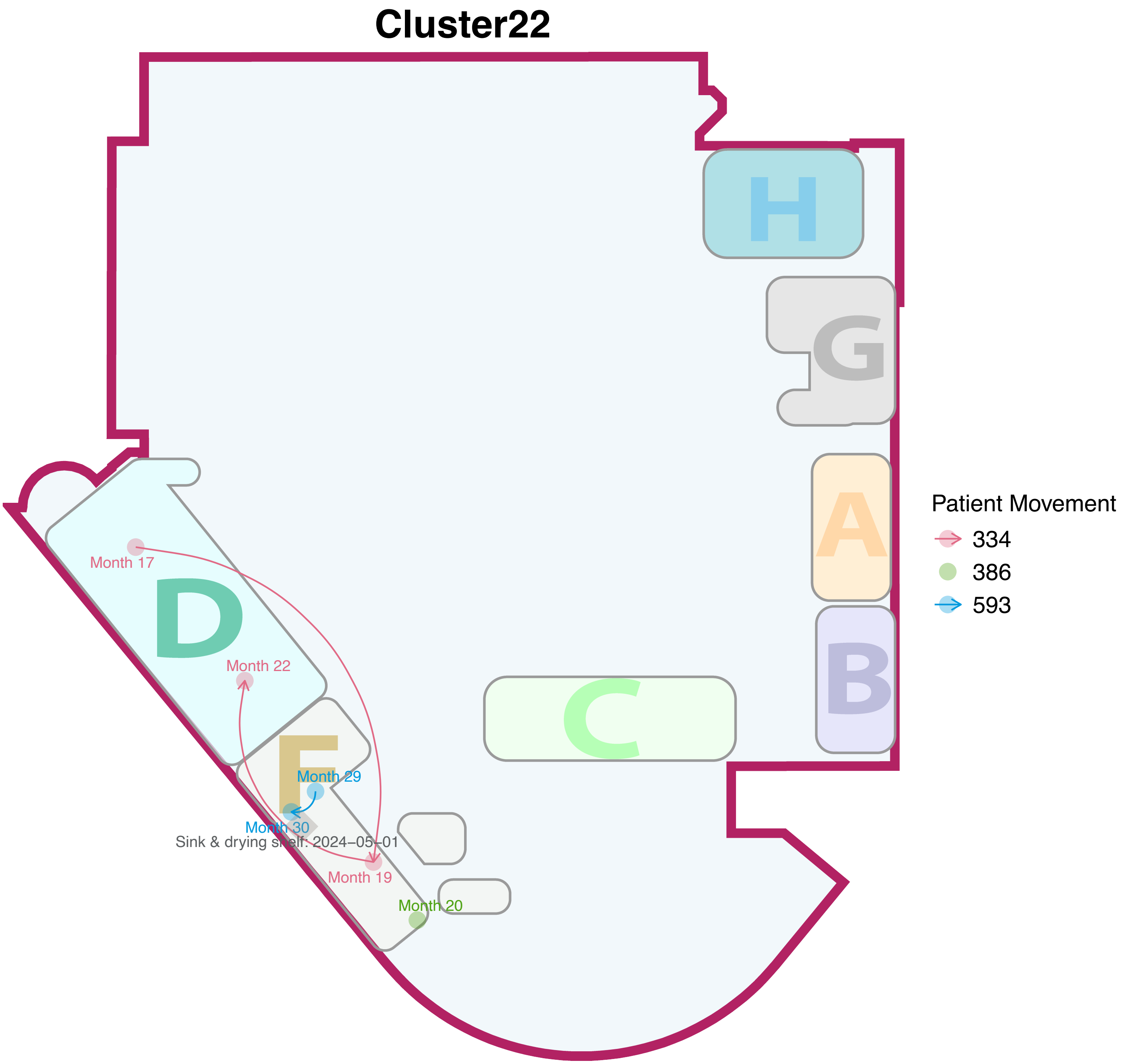

Cluster22
H
G
A
B
C
D
F
Month 17
Month 22
Month 29
Month 30
Sink & drying shelf: 2024–05–01
Month 19
Month 20
Patient Movement
334
386
593

### Cluster23

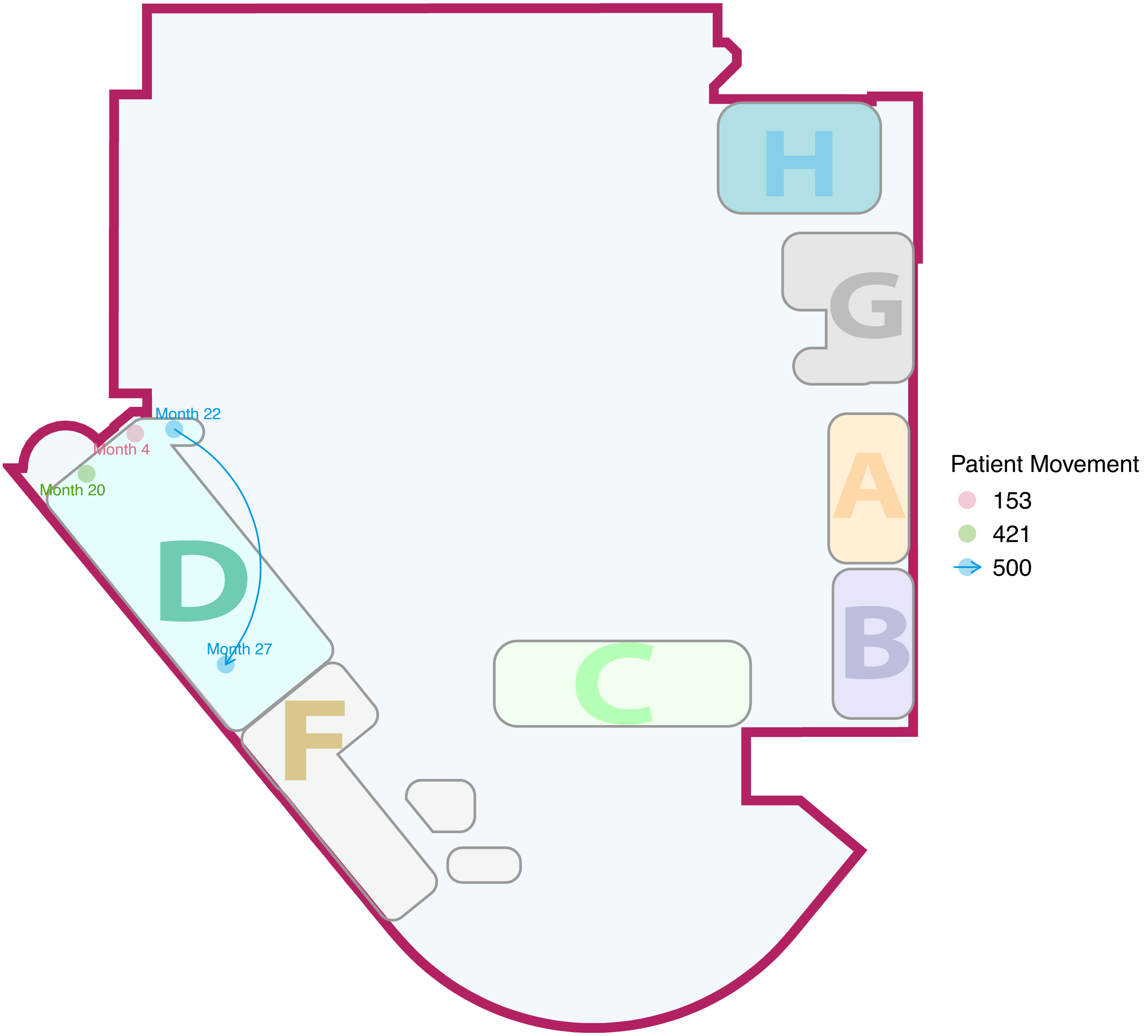

H
G
A
B
C
D
F
Month 22
Month 4
Month 20
Month 27
Patient Movement
153
421
500

### Cluster24

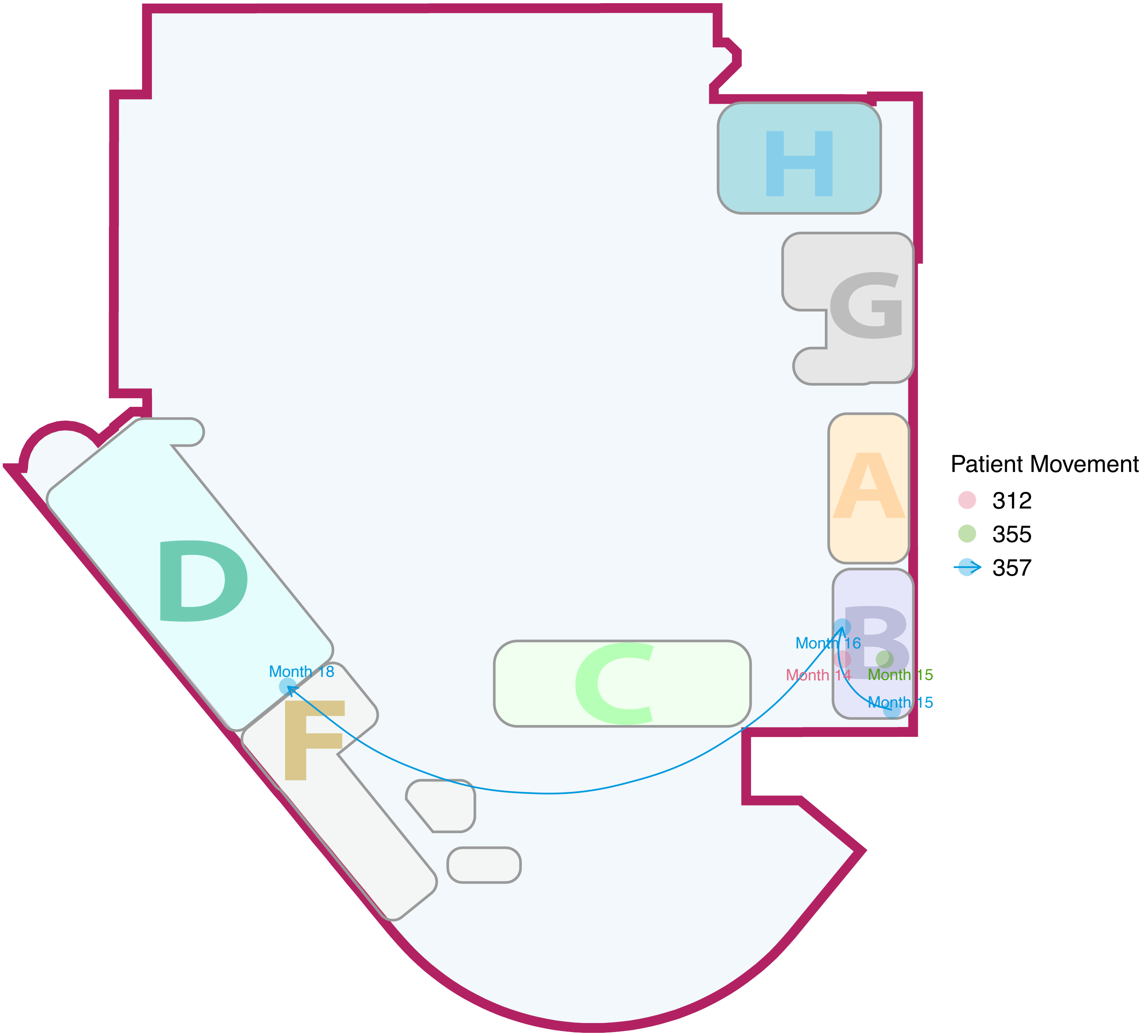

H
G
A
B
C
D
F
Month 18
Month 16
Month 14
Month 15
Month 15
Patient Movement
312
355
357

### Cluster25

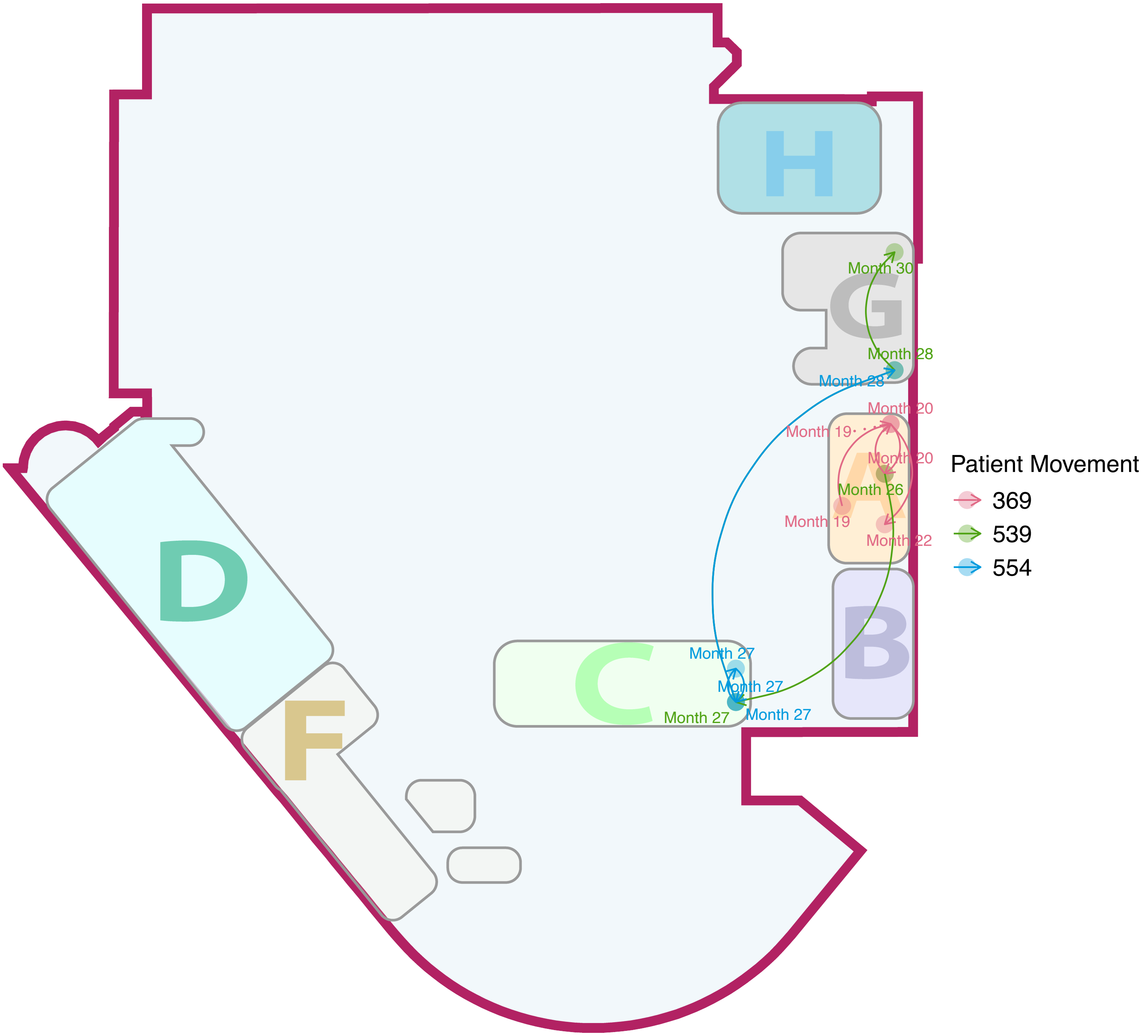

H
G
Month 30
Month 28
Month 28
Month 20
Month 19
Month 20
Month 26
Month 19
Month 22
A
B
C
Month 27
Month 27
Month 27
Month 27
D
F
Patient Movement
369
539
554

### Cluster26

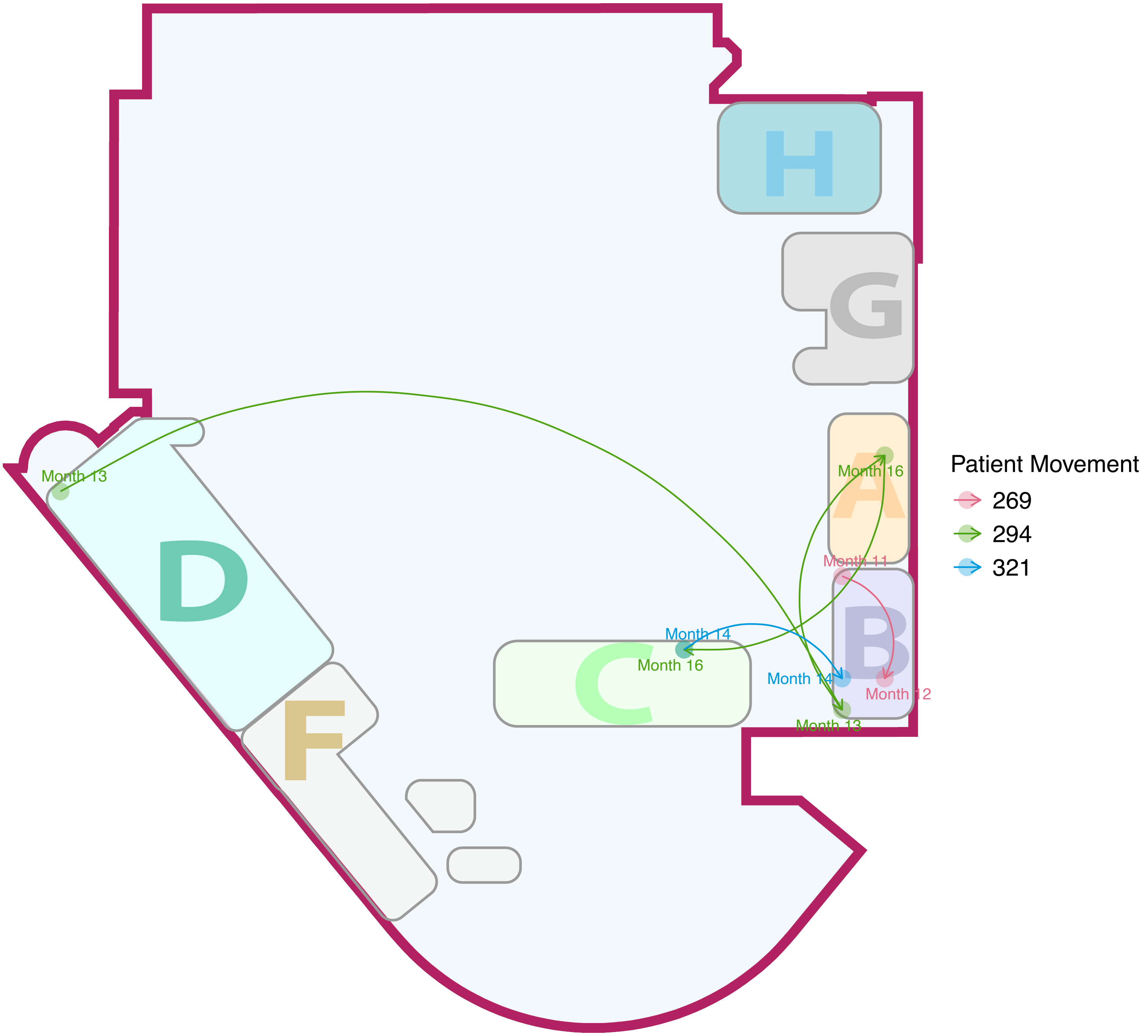

H
G
A
B
C
D
F
Month 13
Month 16
Month 11
Month 14
Month 16
Month 14
Month 12
Month 13
Patient Movement
269
294
321

#### Cluster27

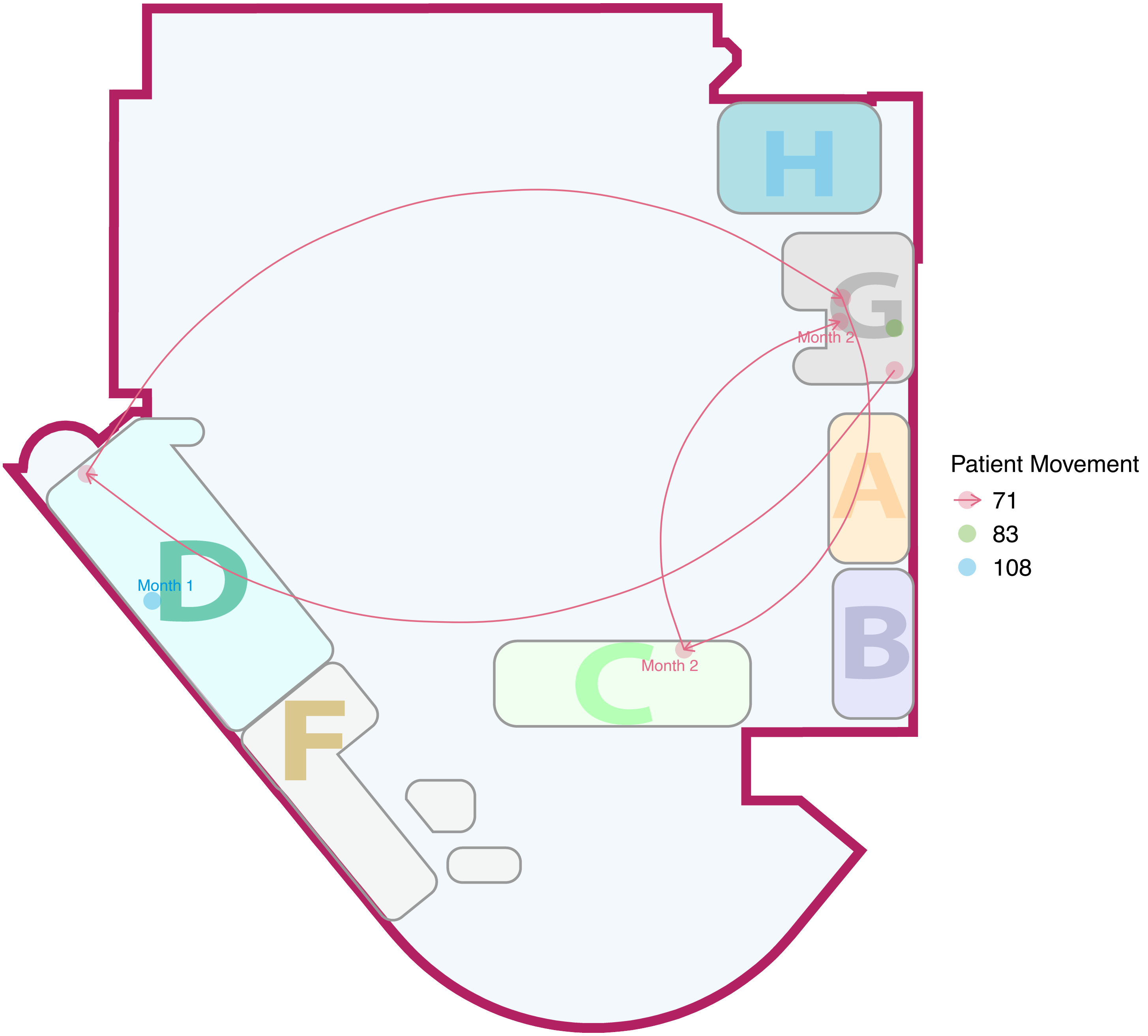

#### Cluster28

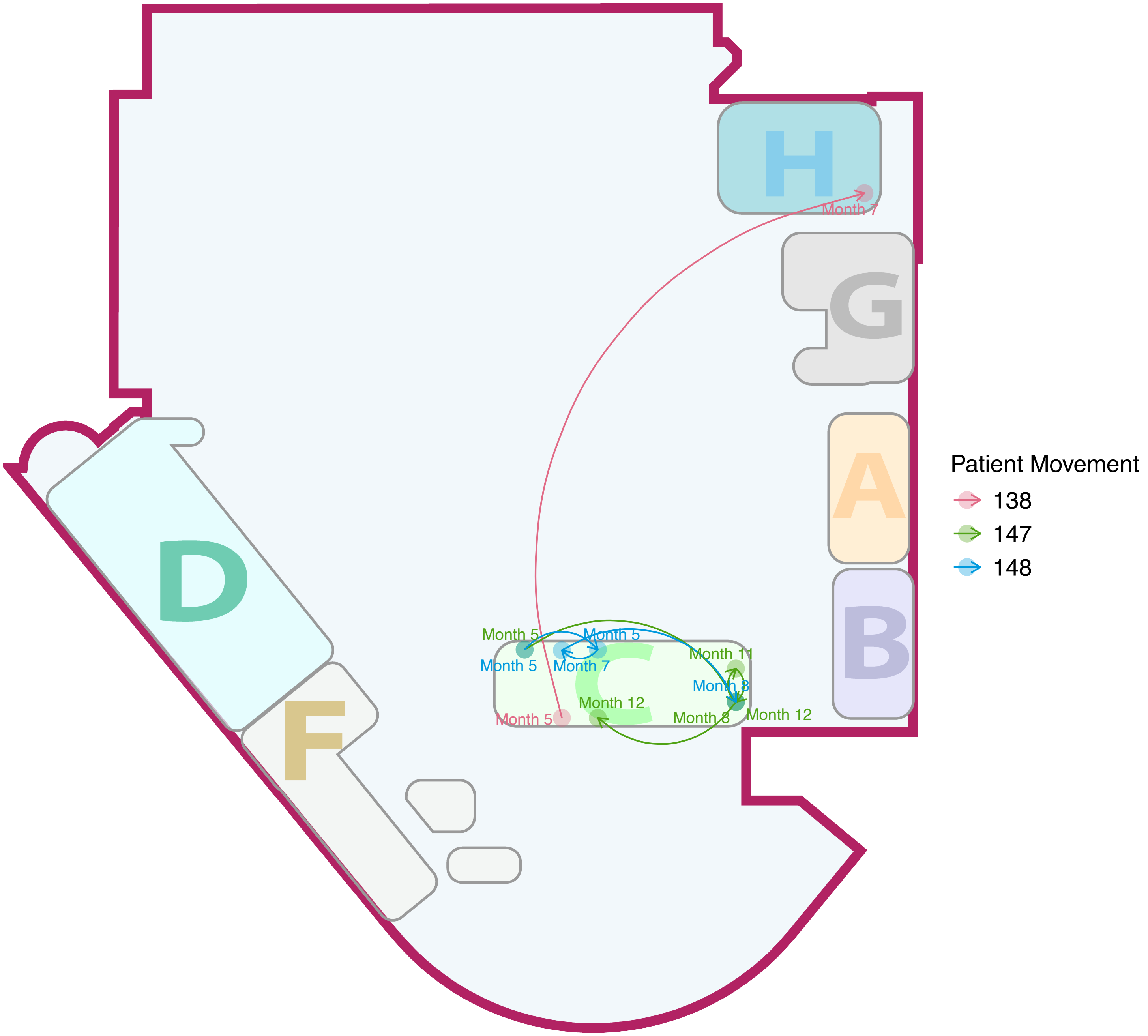

H
G
A
B
D
F
C
Month 7
Month 5
Month 5
Month 5
Month 7
Month 11
Month 8
Month 12
Month 5
Month 8
Month 12
Patient Movement
138
147
148

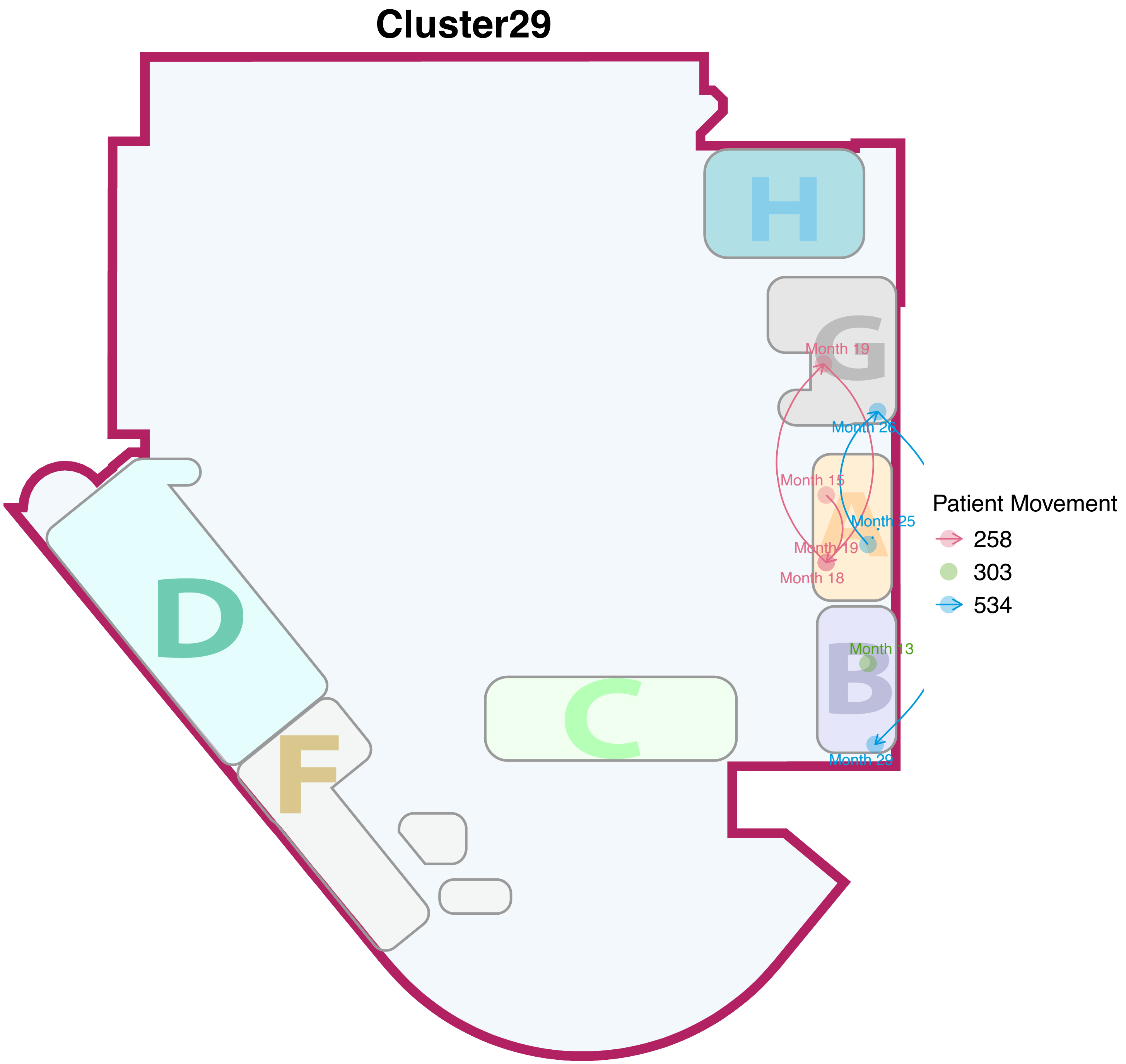
Cluster29
H
G
Month 19
Month 26
Month 15
Month 25
Month 19
Month 18
Month 13
B
Month 29
C
D
F
Patient Movement
258
303
534

#### Cluster30

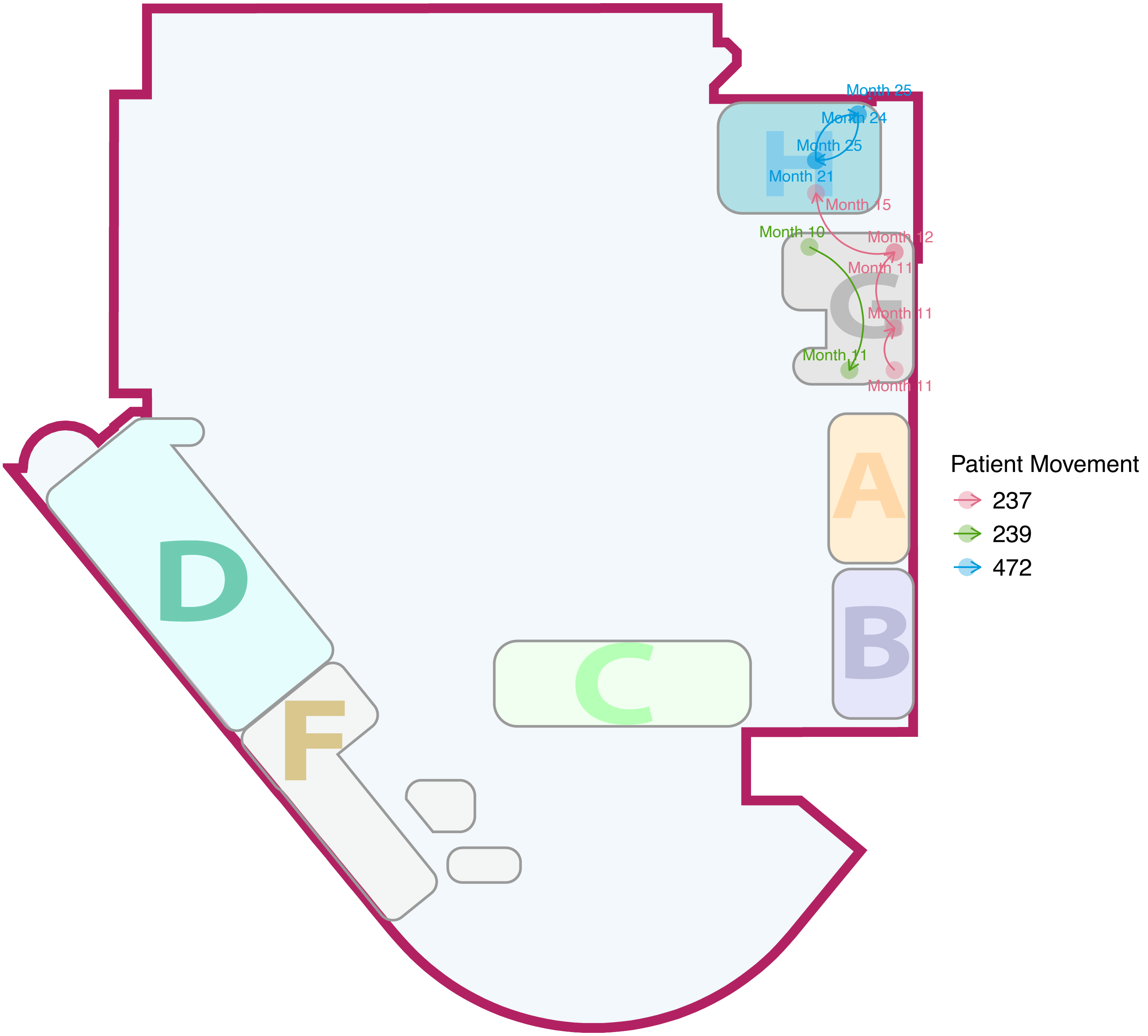

Month 25
Month 24
Month 25
Month 21
Month 15
Month 10
Month 12
Month 11
Month 11
Month 11
Month 11
H
G
A
B
C
D
F
Patient Movement
237
239
472

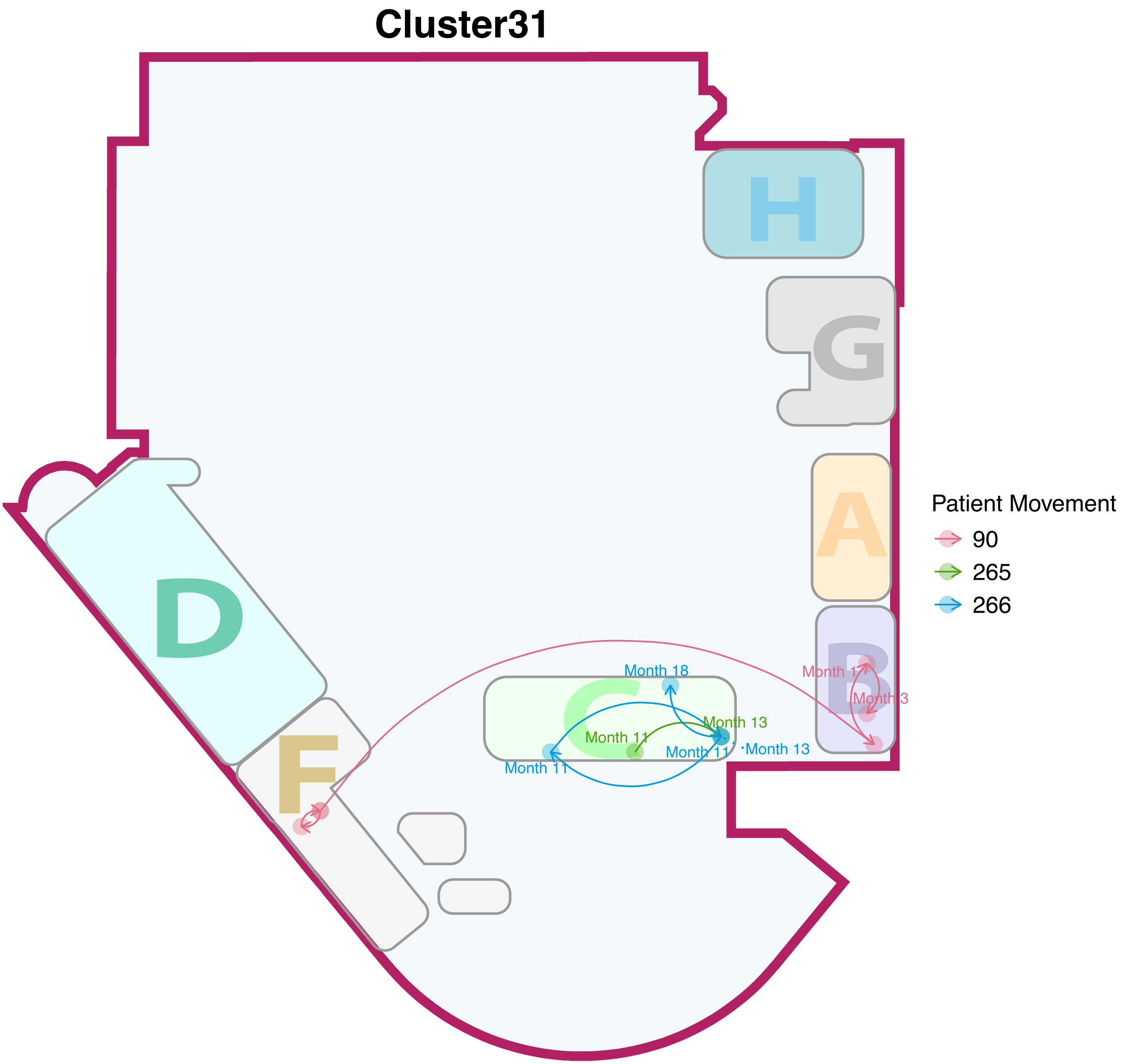

Cluster31
H
G
A
D
B
C
F
Month 18
Month 17
Month 3
Month 13
Month 11
Month 11
Month 13
Month 11
Patient Movement
90
265
266

### Cluster32

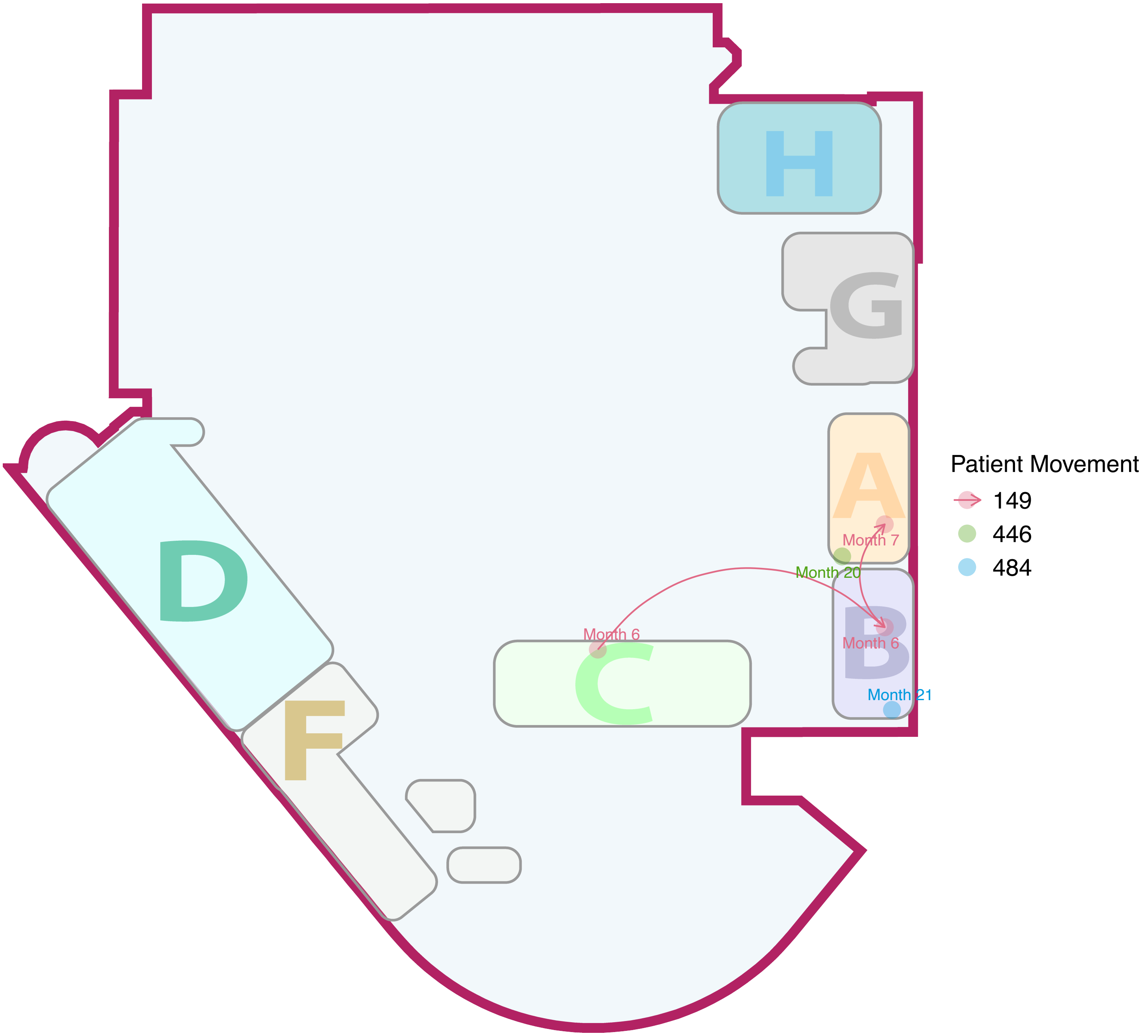

H
G
A
B
C
D
F
Month 7
Month 20
Month 6
Month 6
Month 21
Patient Movement
149
446
484

### Cluster33

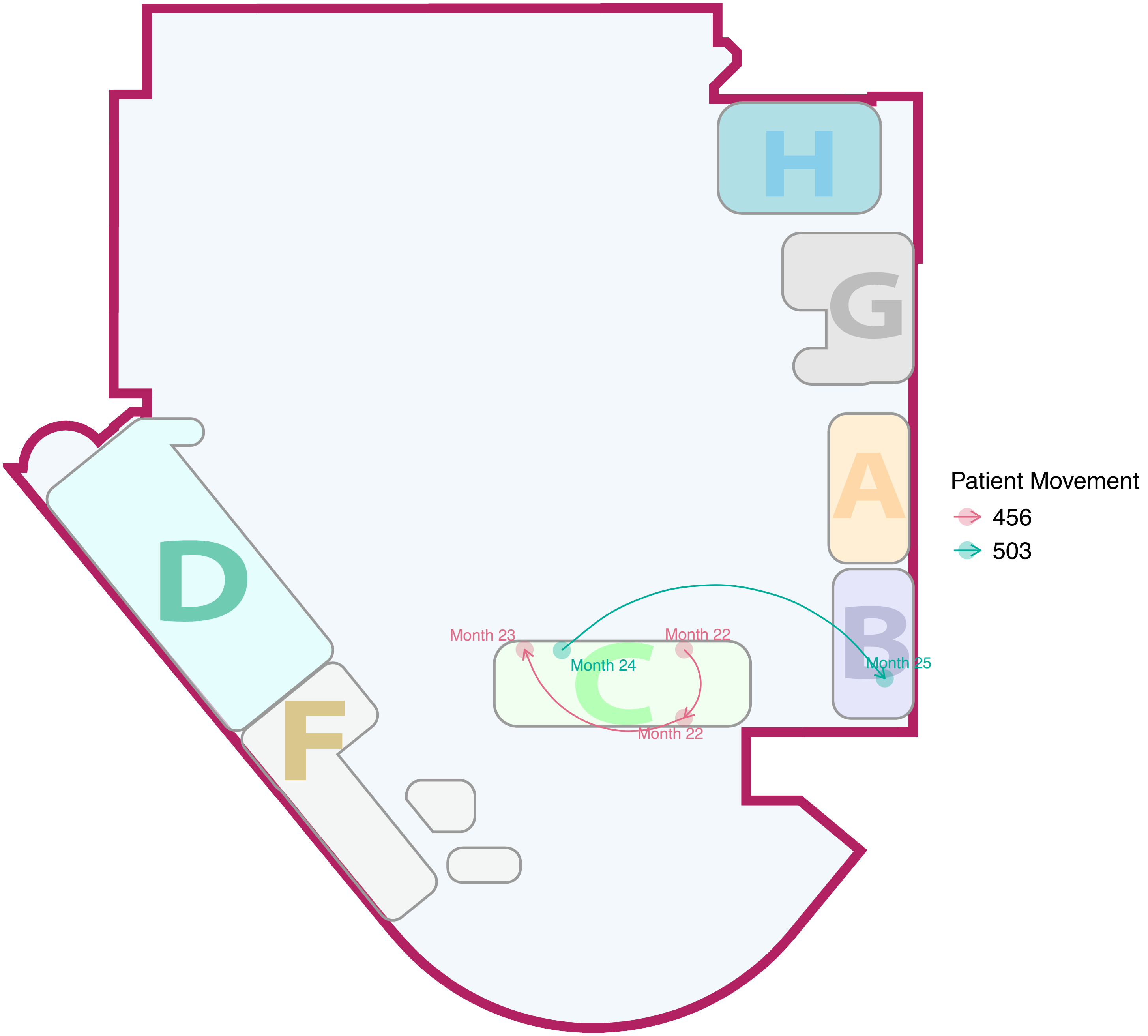
H
G
A
B
C
D
F
Month 23
Month 22
Month 24
Month 25
Month 22
Patient Movement
456
503

### Cluster34

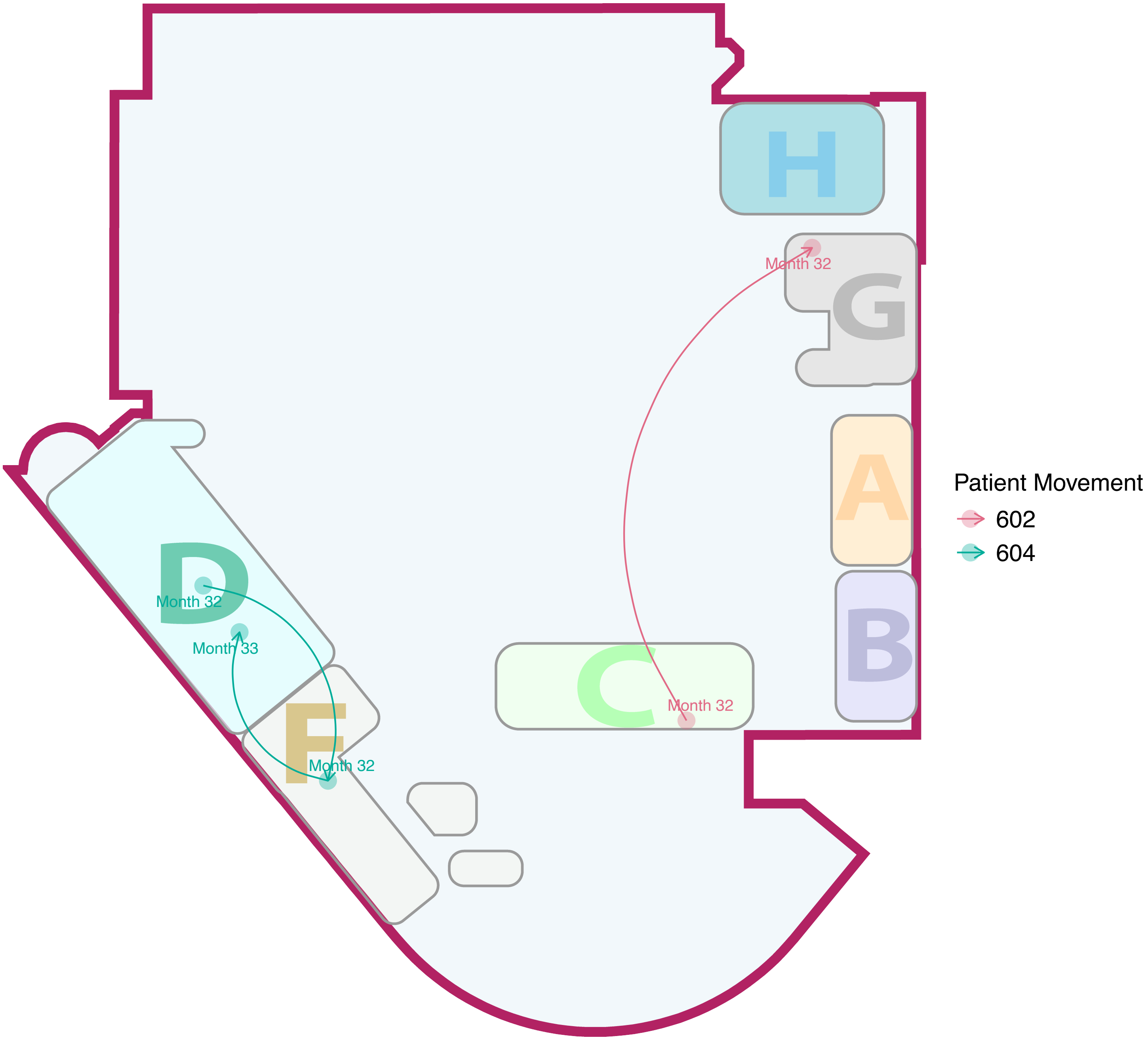

H
G
Month 32
A
B
C
Month 32
D
Month 32
Month 33
F
Month 32
Patient Movement
602
604

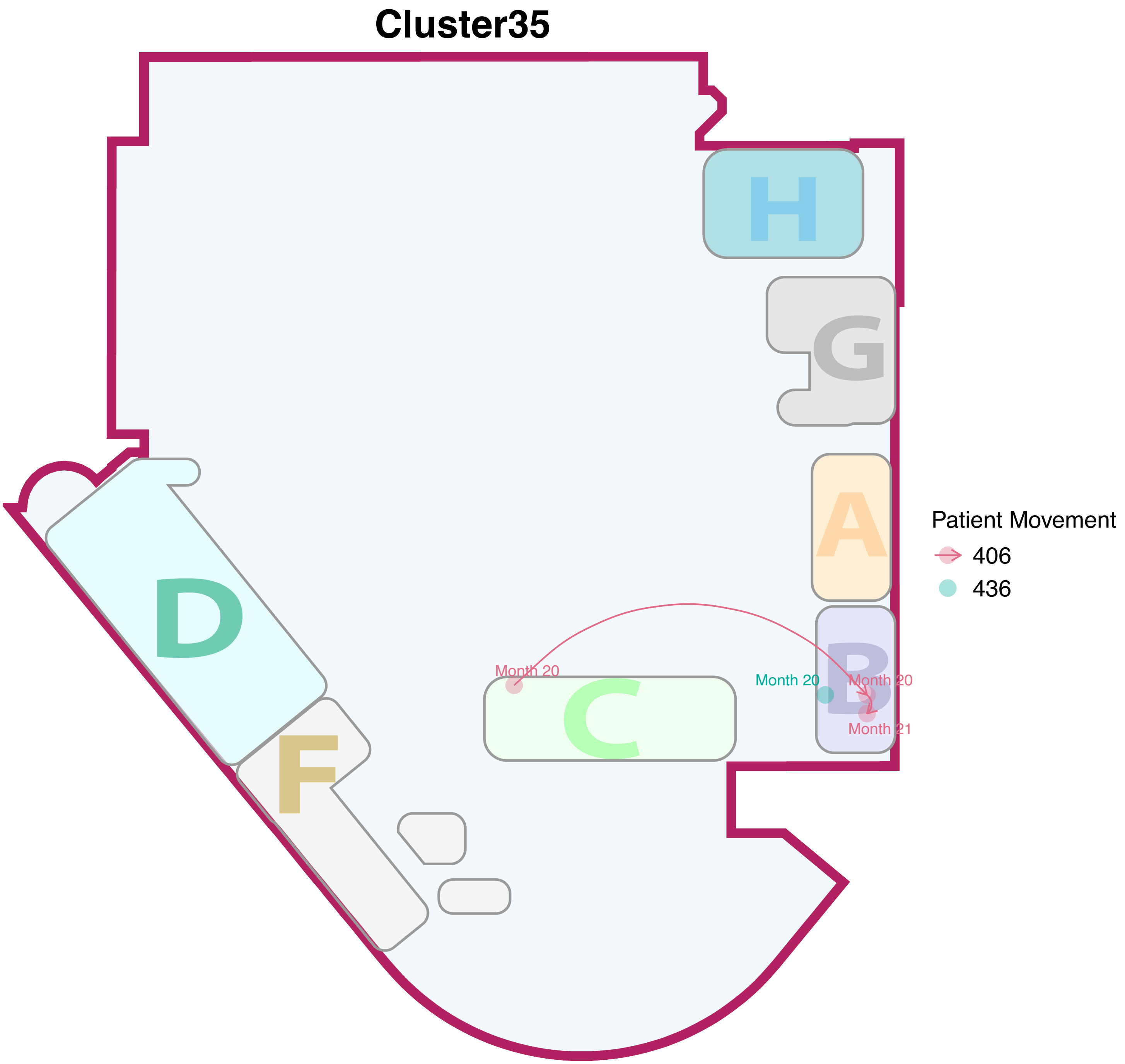
Cluster35
H
G
A
D
B
C
F
Month 20
Month 20
Month 20
Month 21
Patient Movement
406
436

### Cluster36

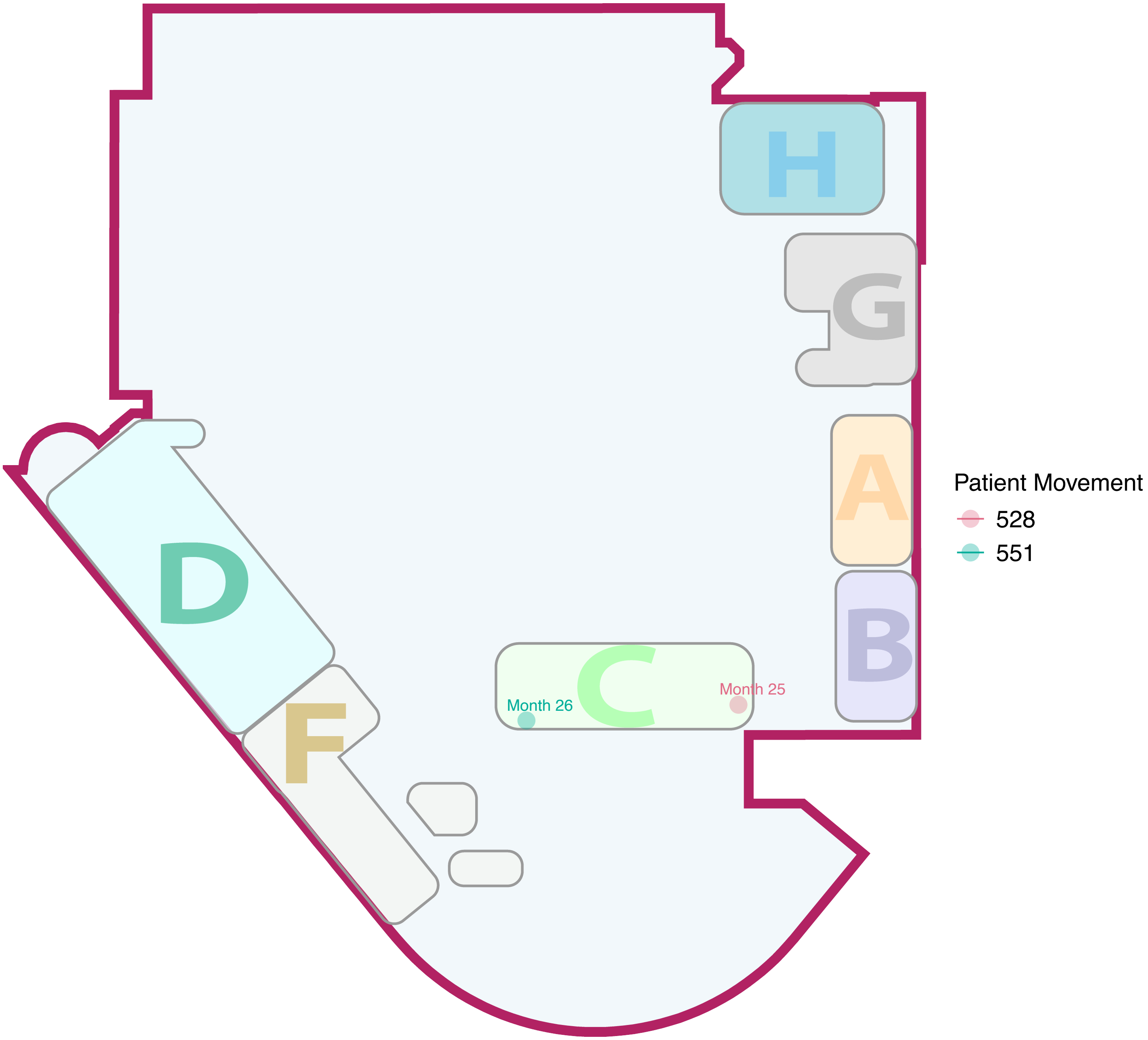

### Cluster37

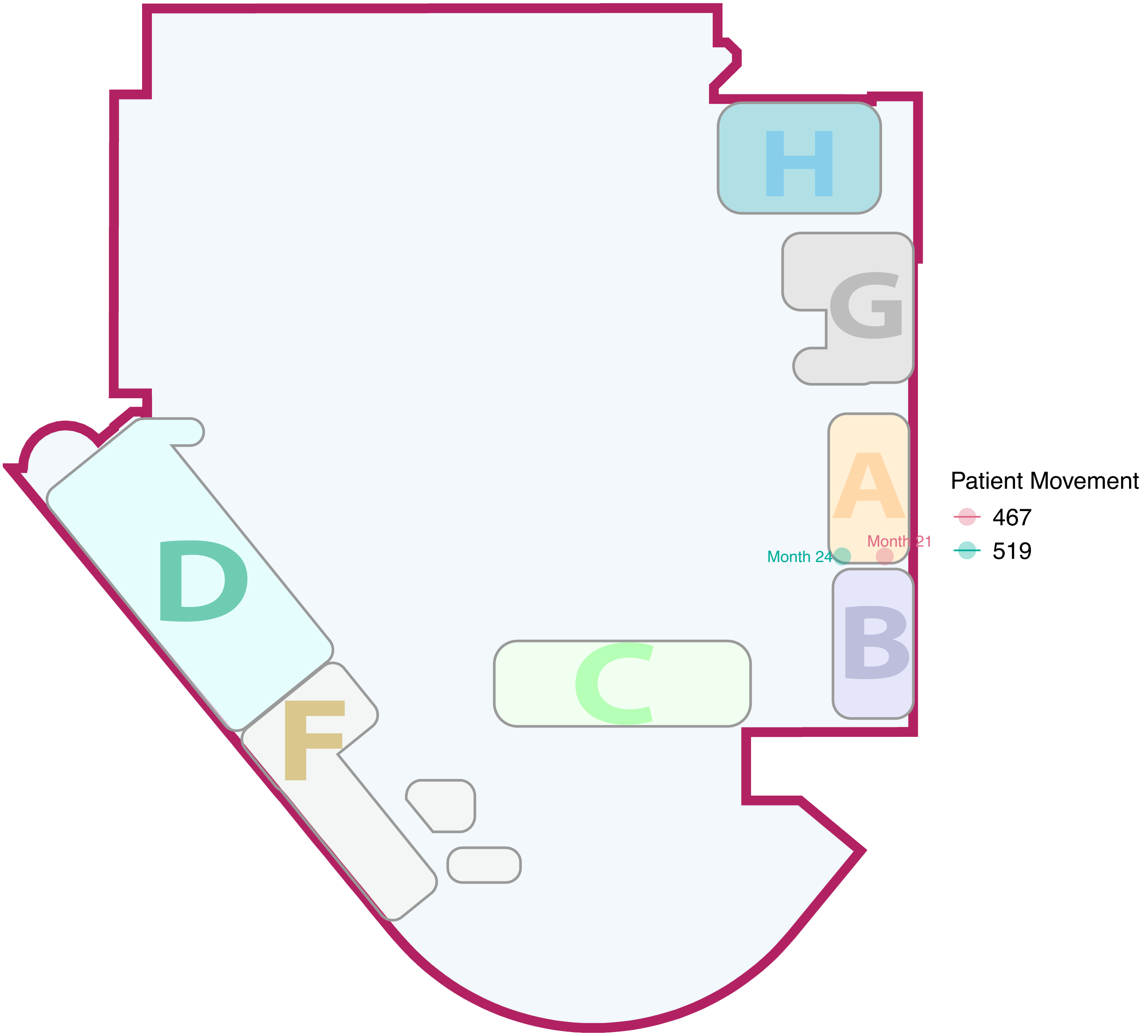

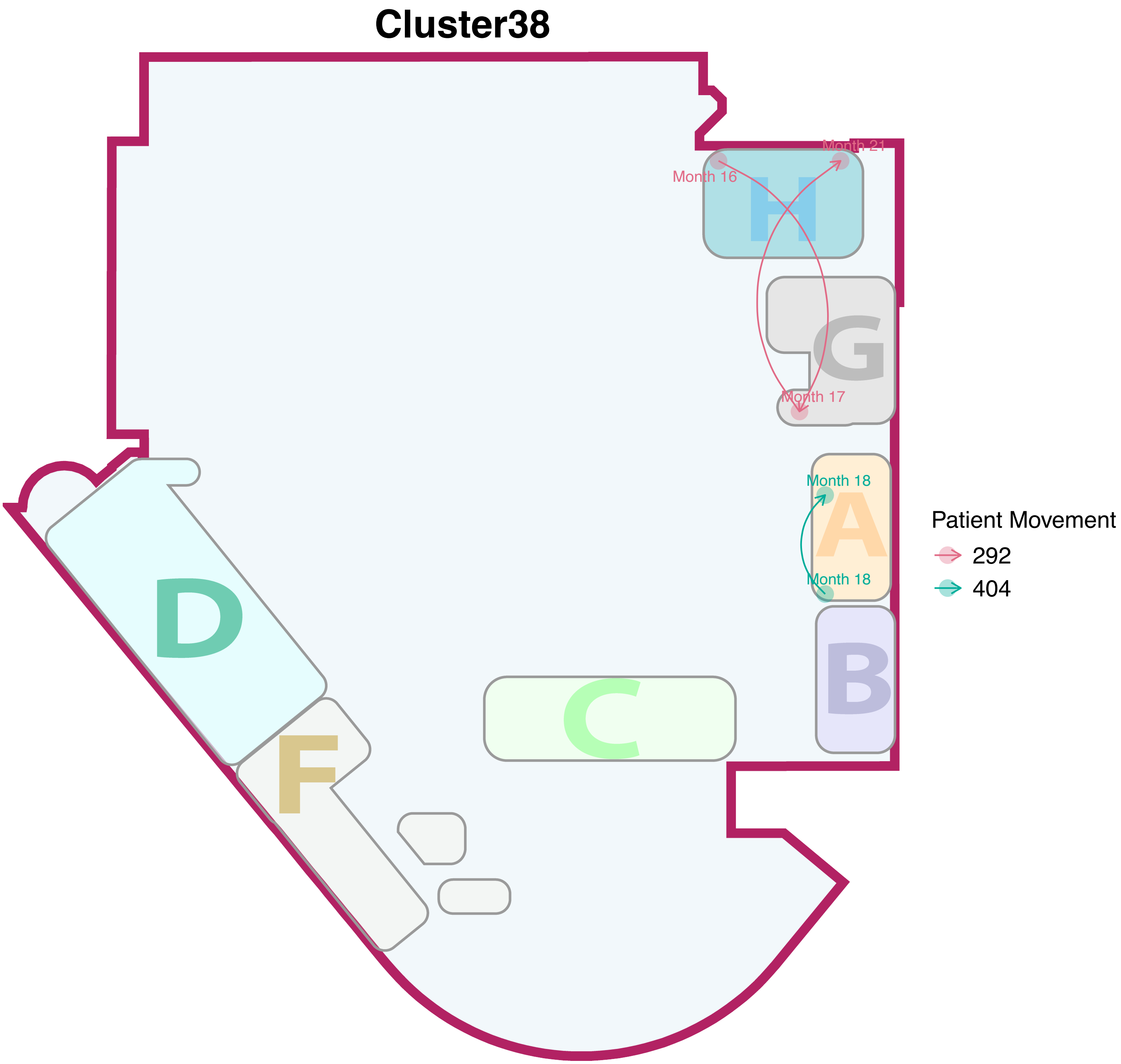

Cluster38
Month 16
Month 21
Month 17
Month 18
Month 18
H
G
A
B
C
D
F
Patient Movement
292
404

### Cluster39

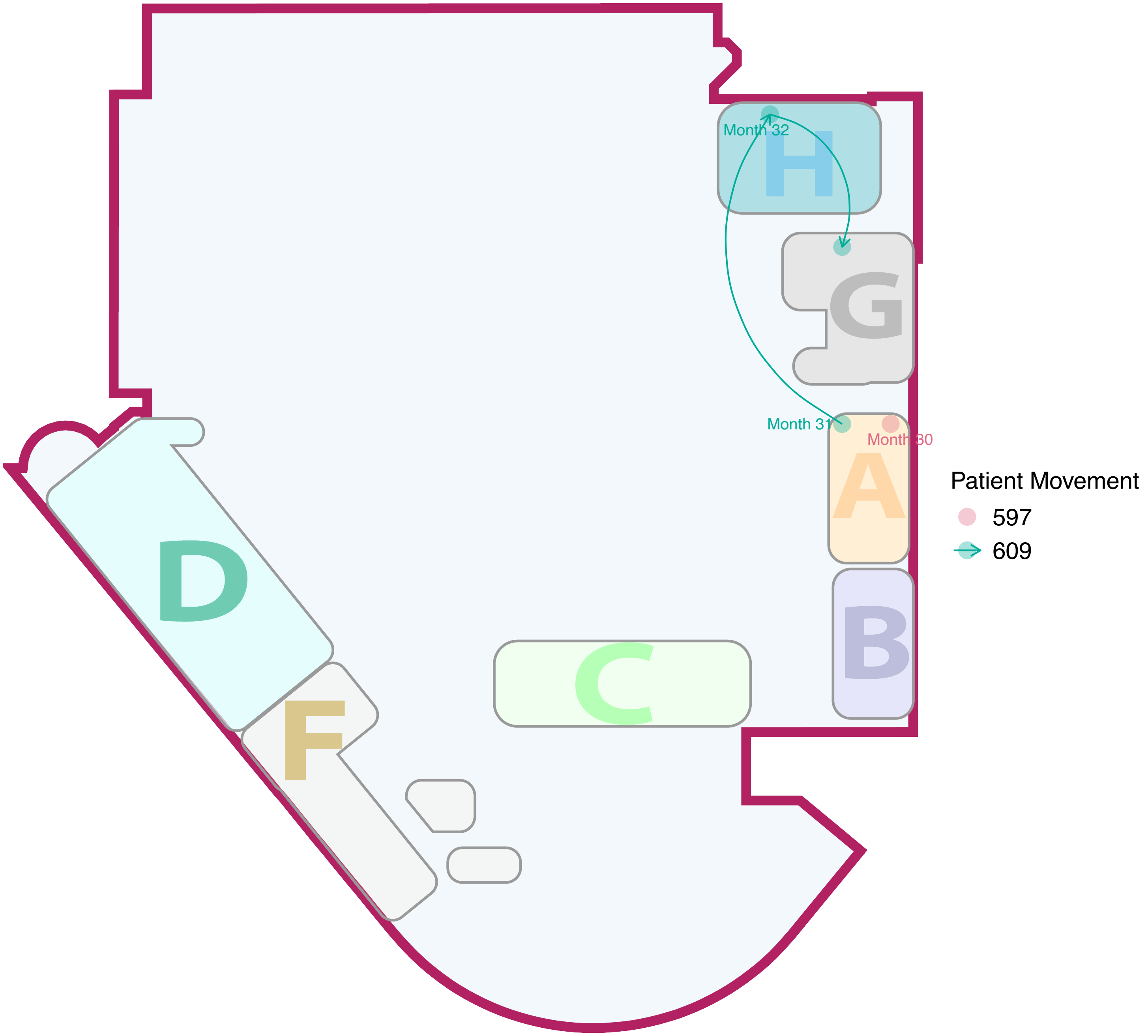

### Cluster40

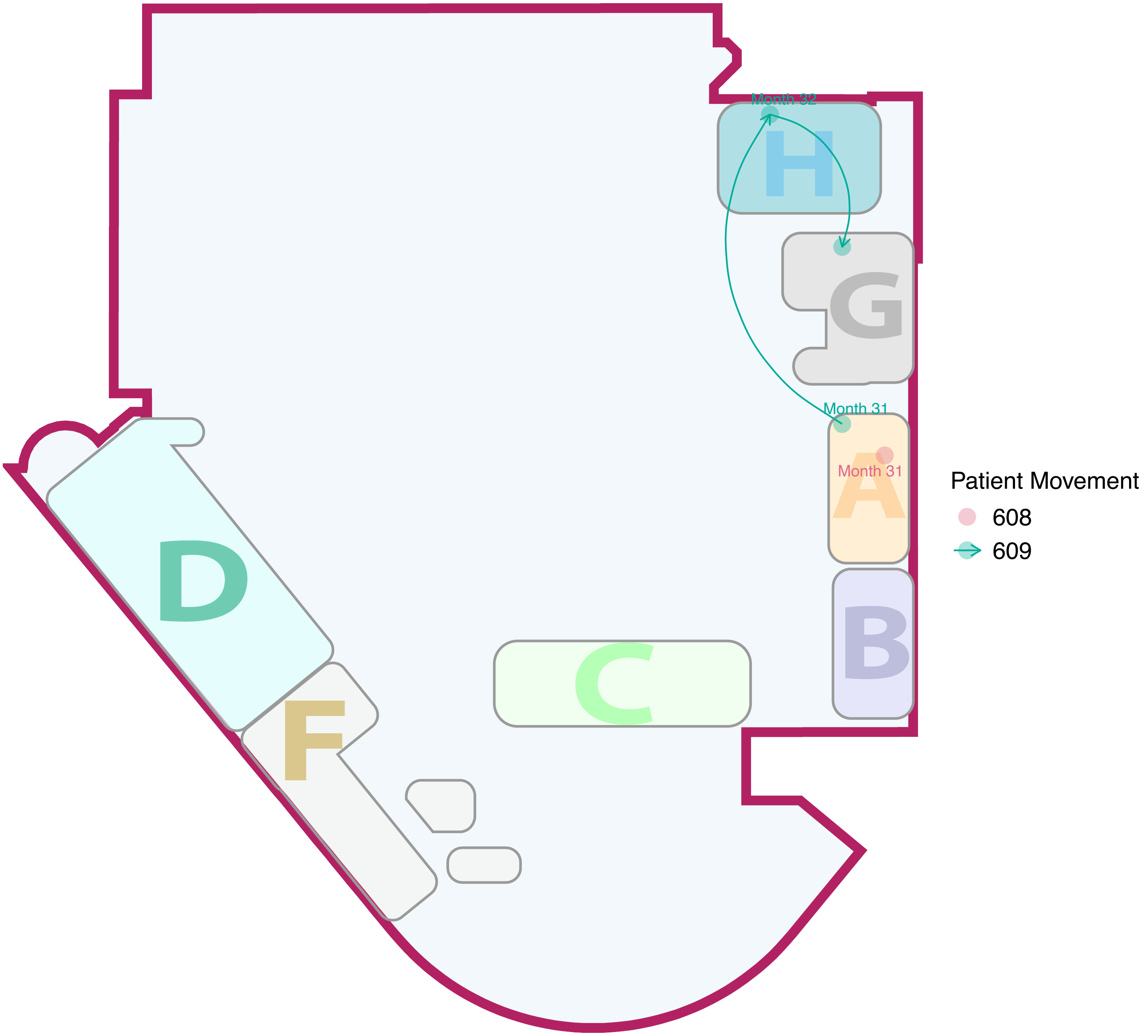

### Cluster41

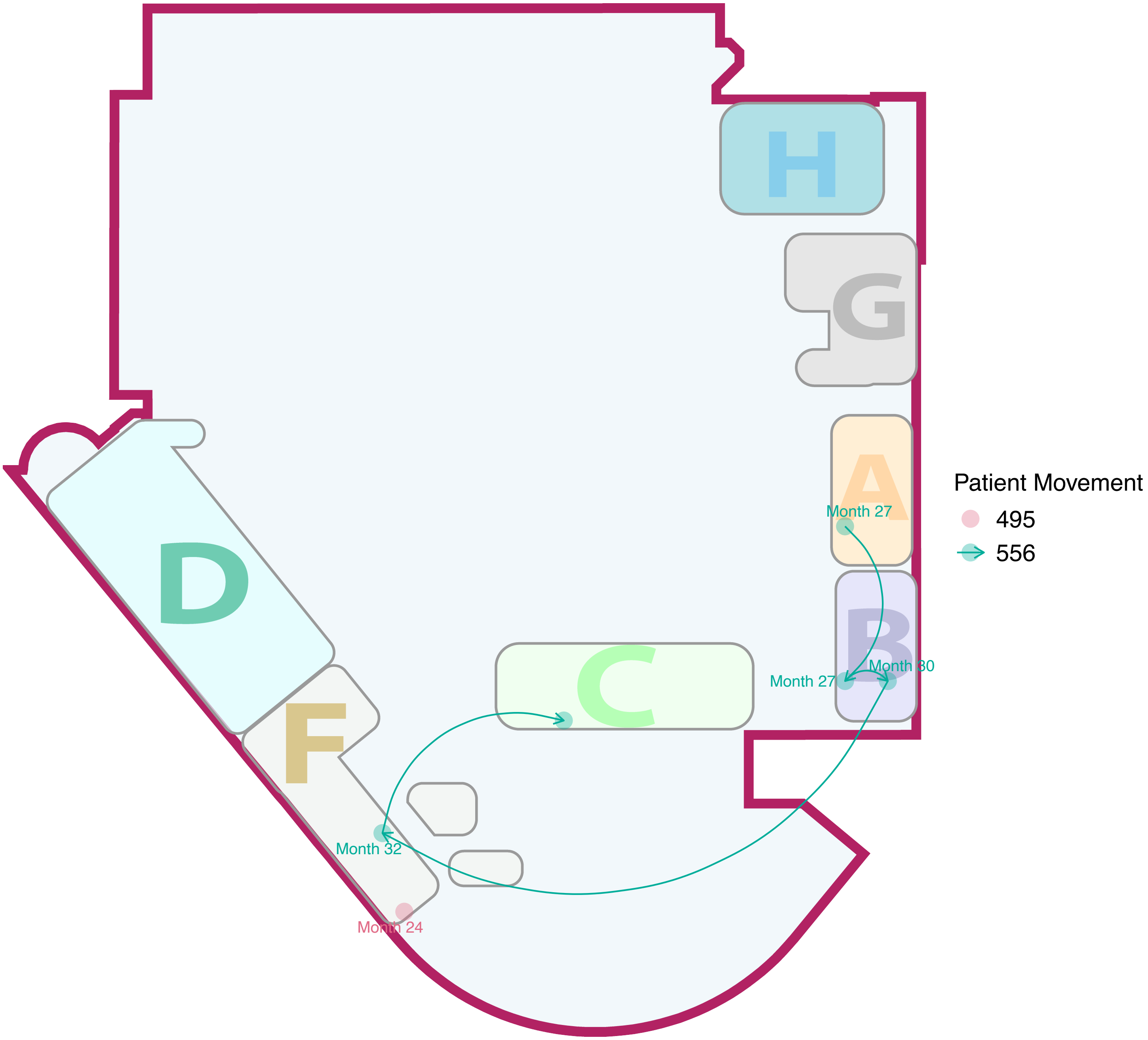

H
G
A
Month 27
B
Month 27
Month 30
C
D
F
Month 32
Month 24
Patient Movement
495
556

#### Cluster42

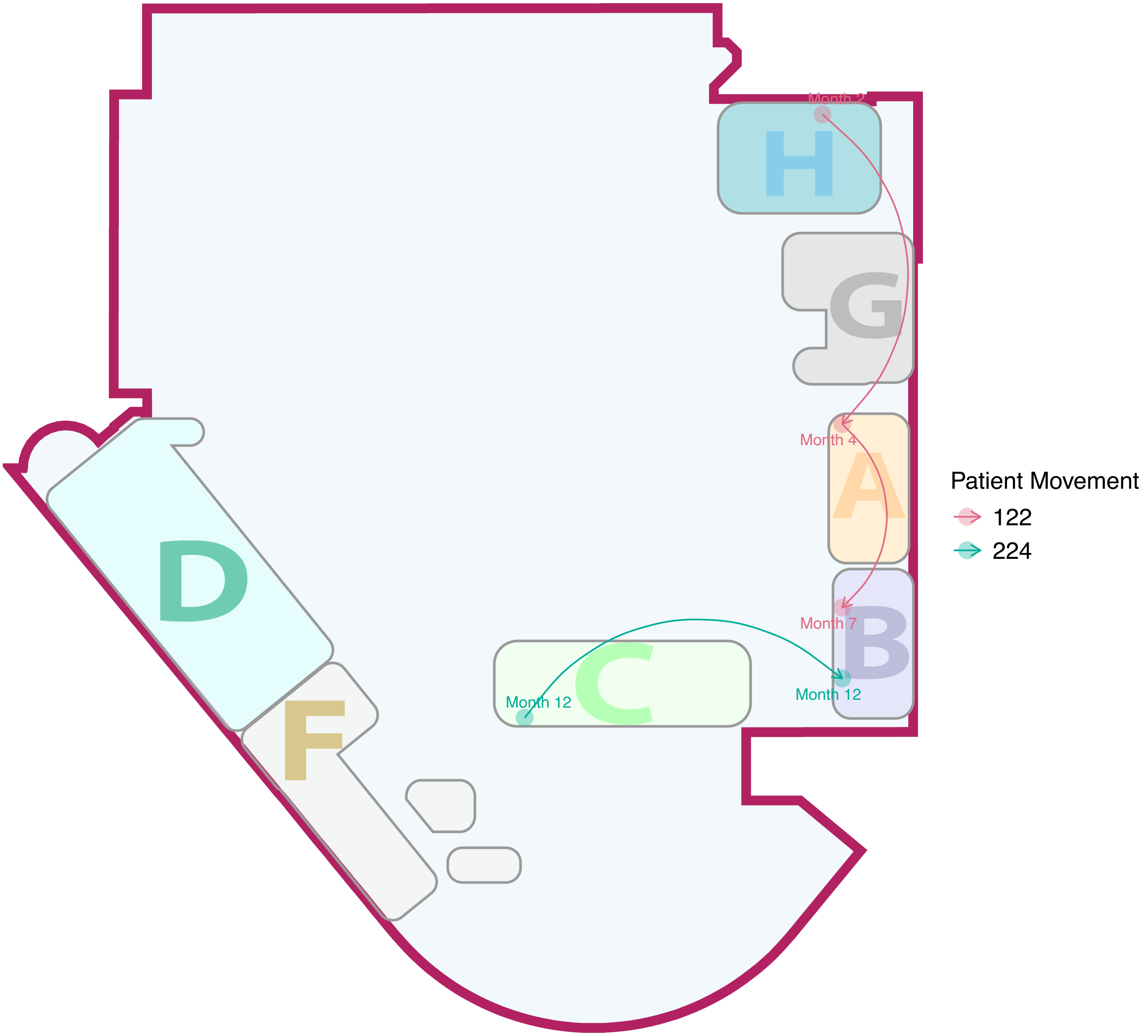

Month 2
H
G
Month 4
A
Month 7
B
Month 12
C
Month 12
D
F
Patient Movement
122
224

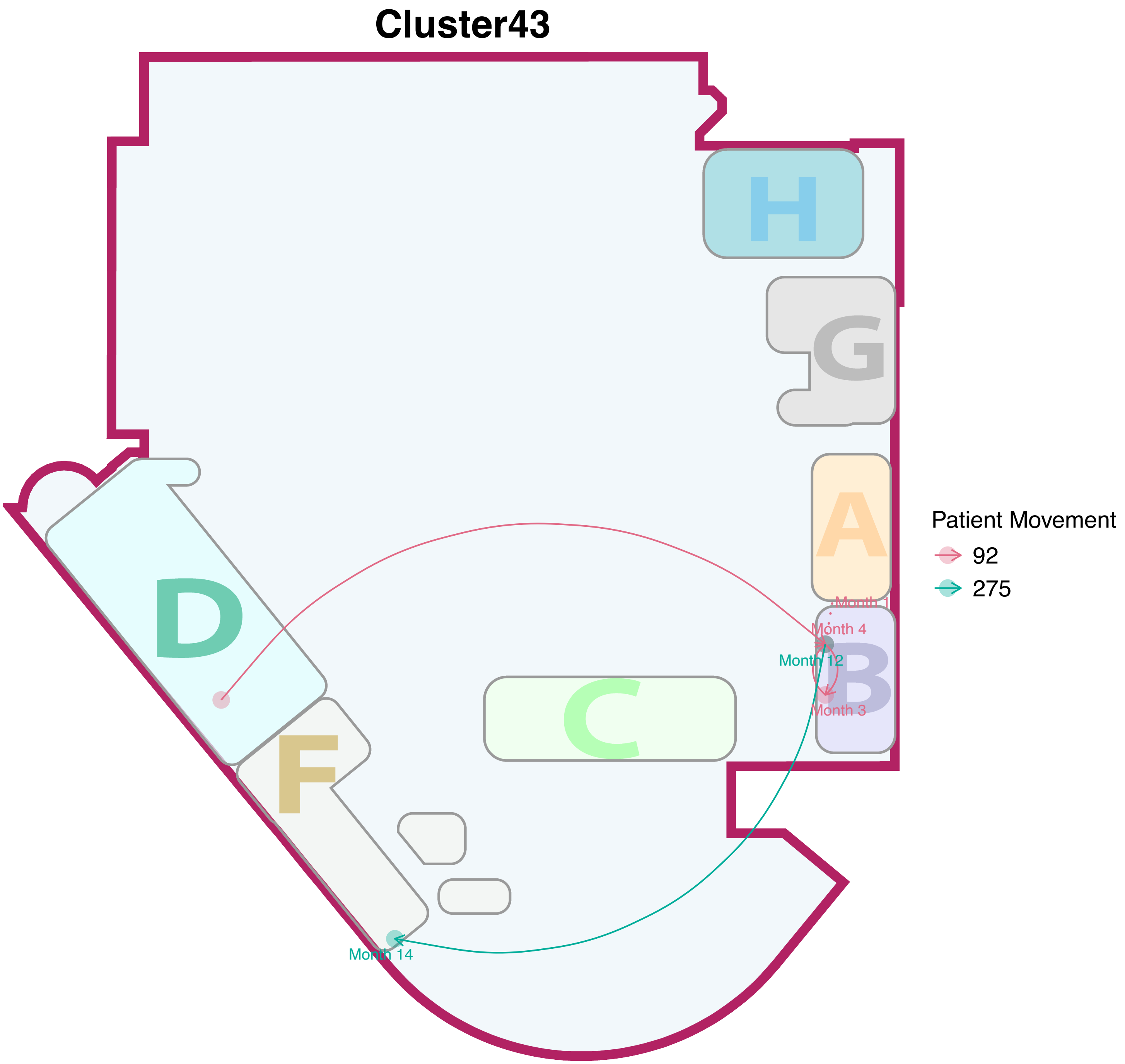

Cluster43
H
G
A
B
C
D
F
Month 4
Month 12
Month 3
Month 14
Patient Movement
92
275

### Cluster44

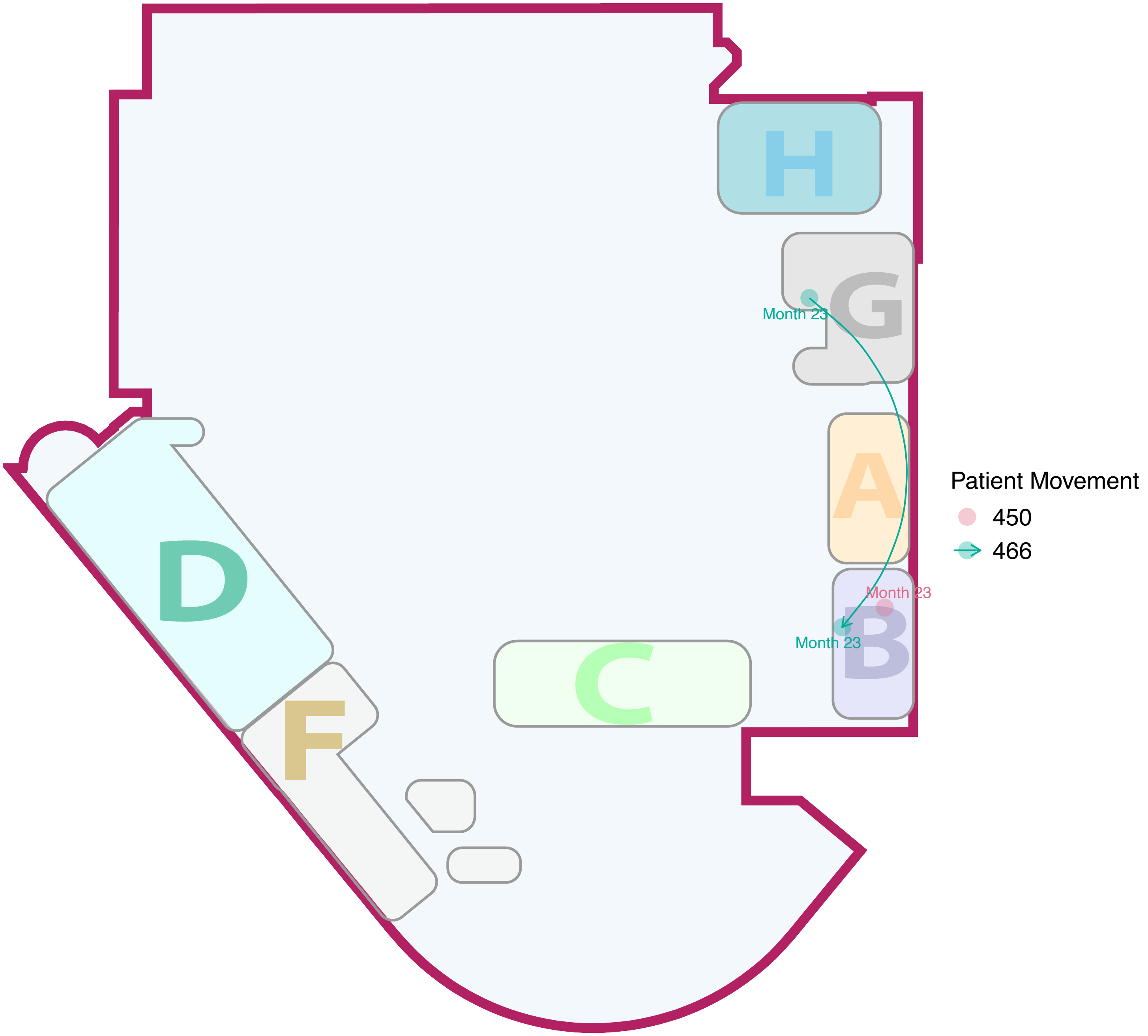

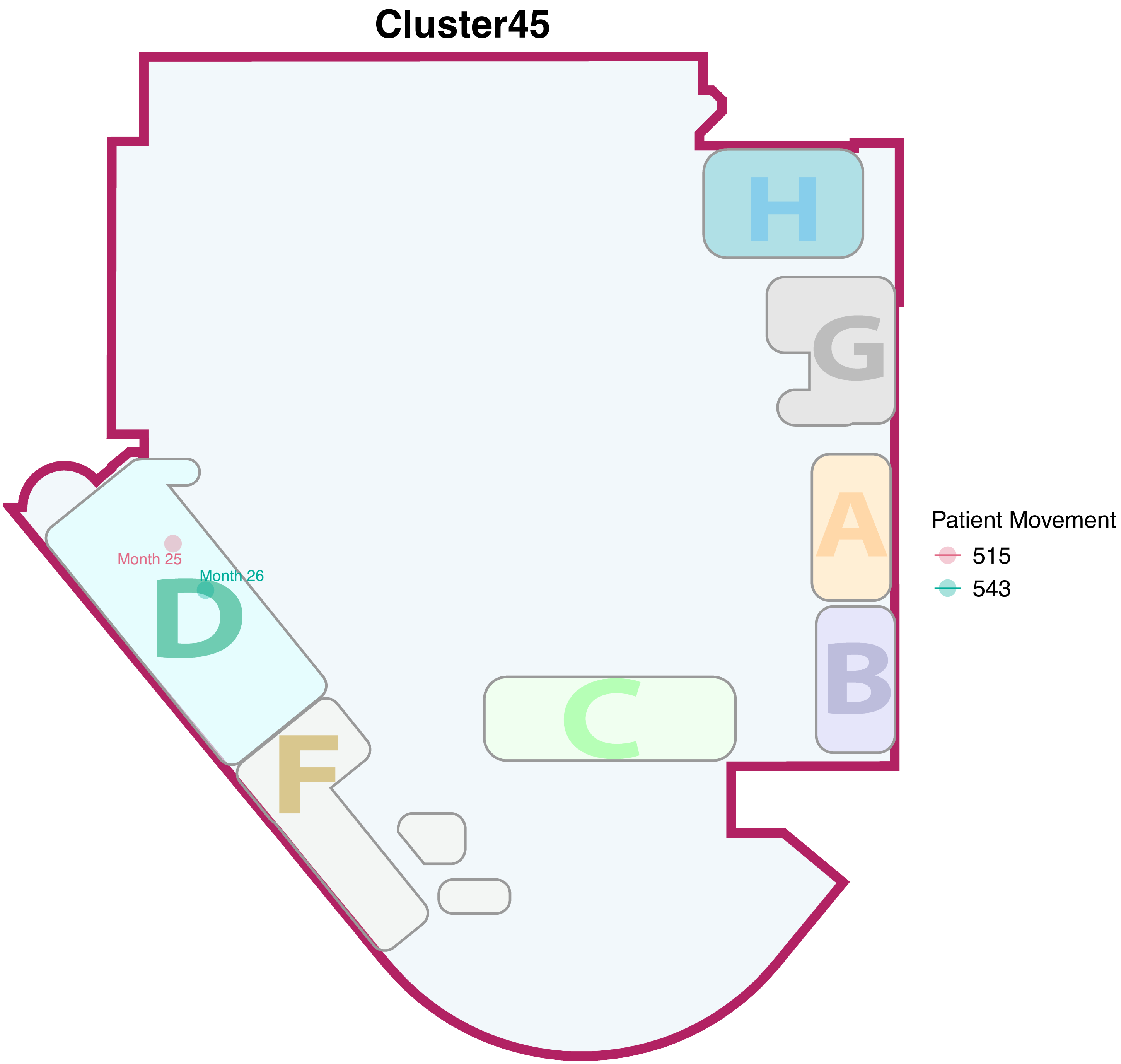
Cluster45
H
G
A
B
C
D
F
Month 25
Month 26
Patient Movement
515
543

### Cluster46

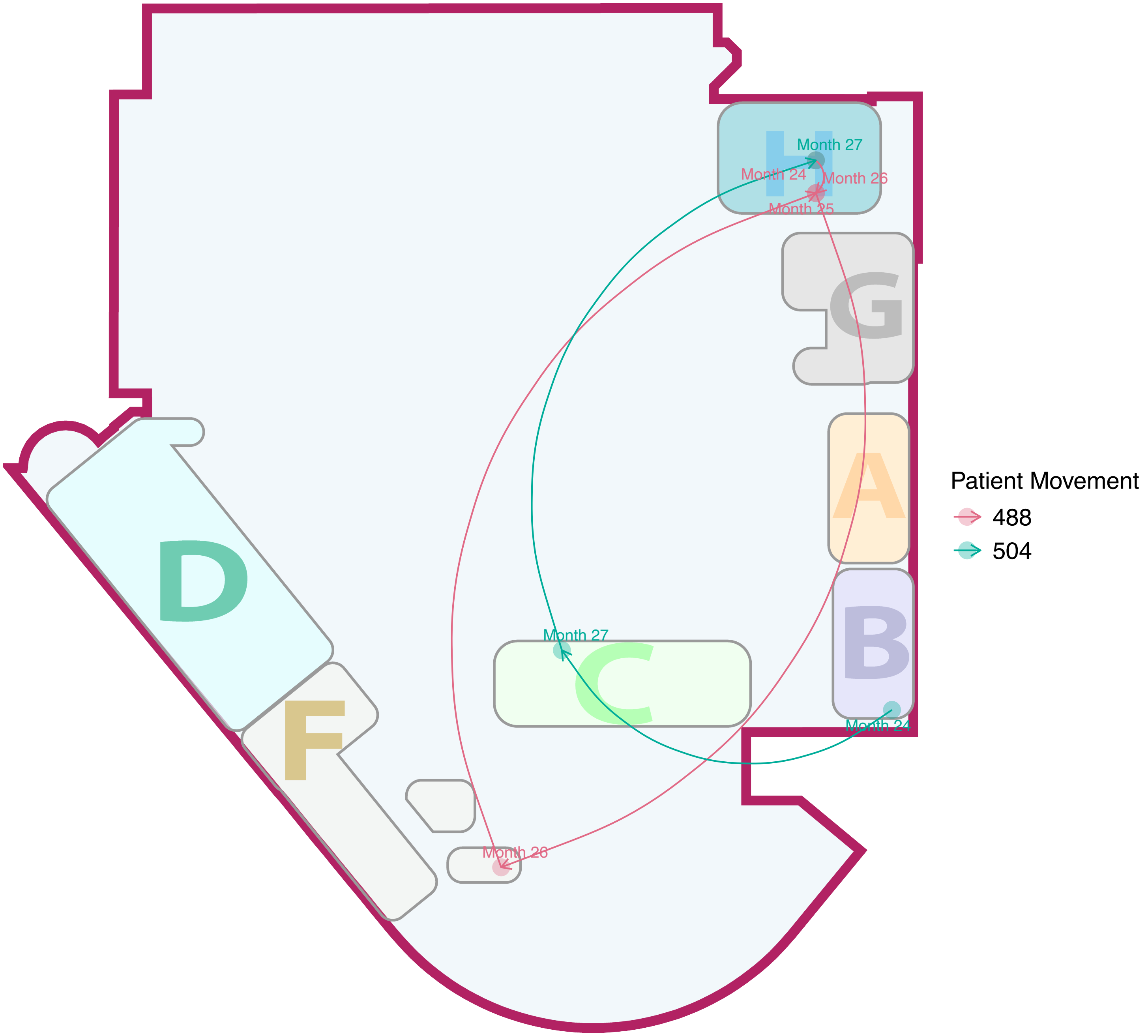

H
G
A
B
C
D
F
Month 27
Month 24
Month 26
Month 25
Month 27
Month 24
Month 26
Patient Movement
488
504

### Cluster47

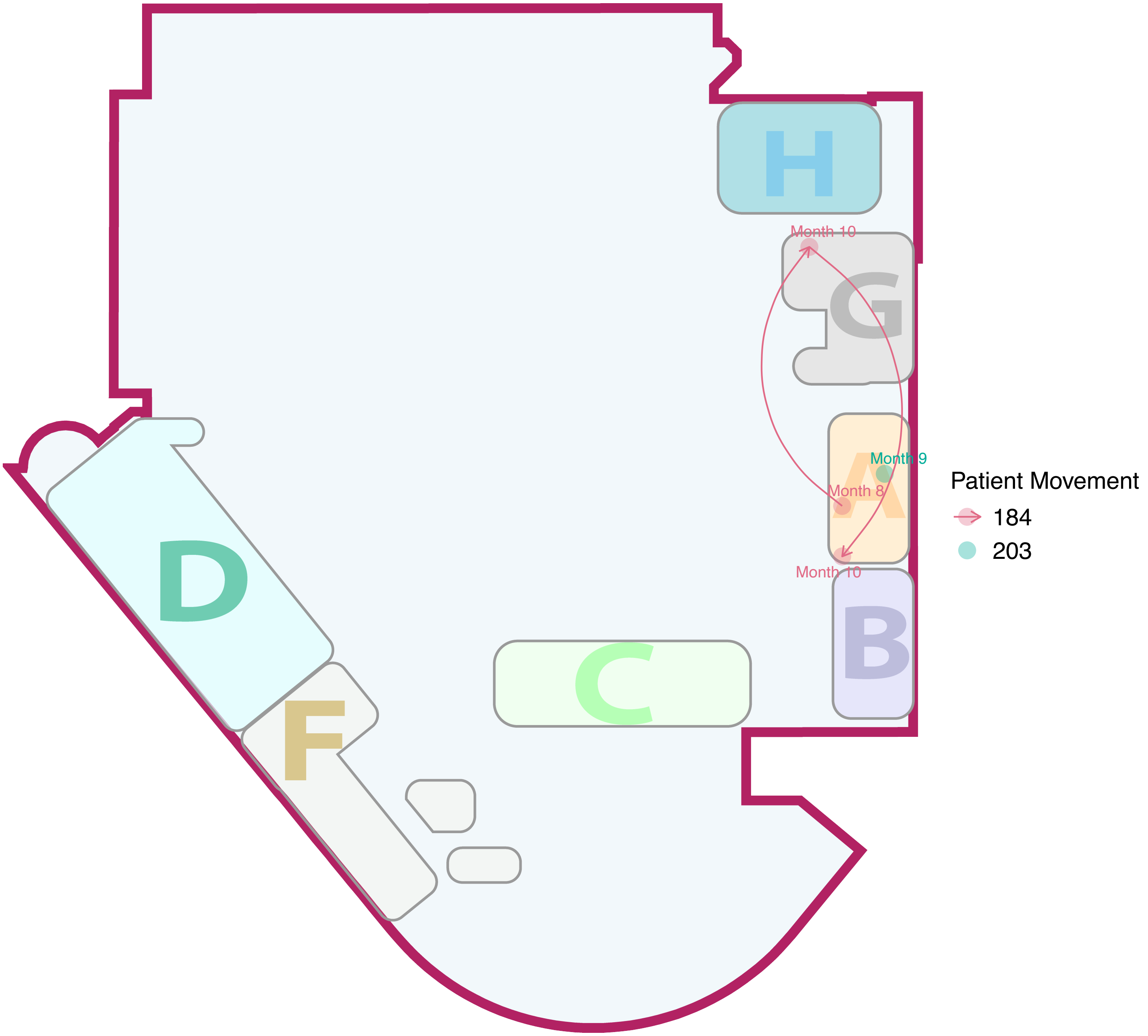
H
Month 10
G
Month 9
Month 8
A
Month 10
B
D
C
F
Patient Movement
184
203

### Cluster48

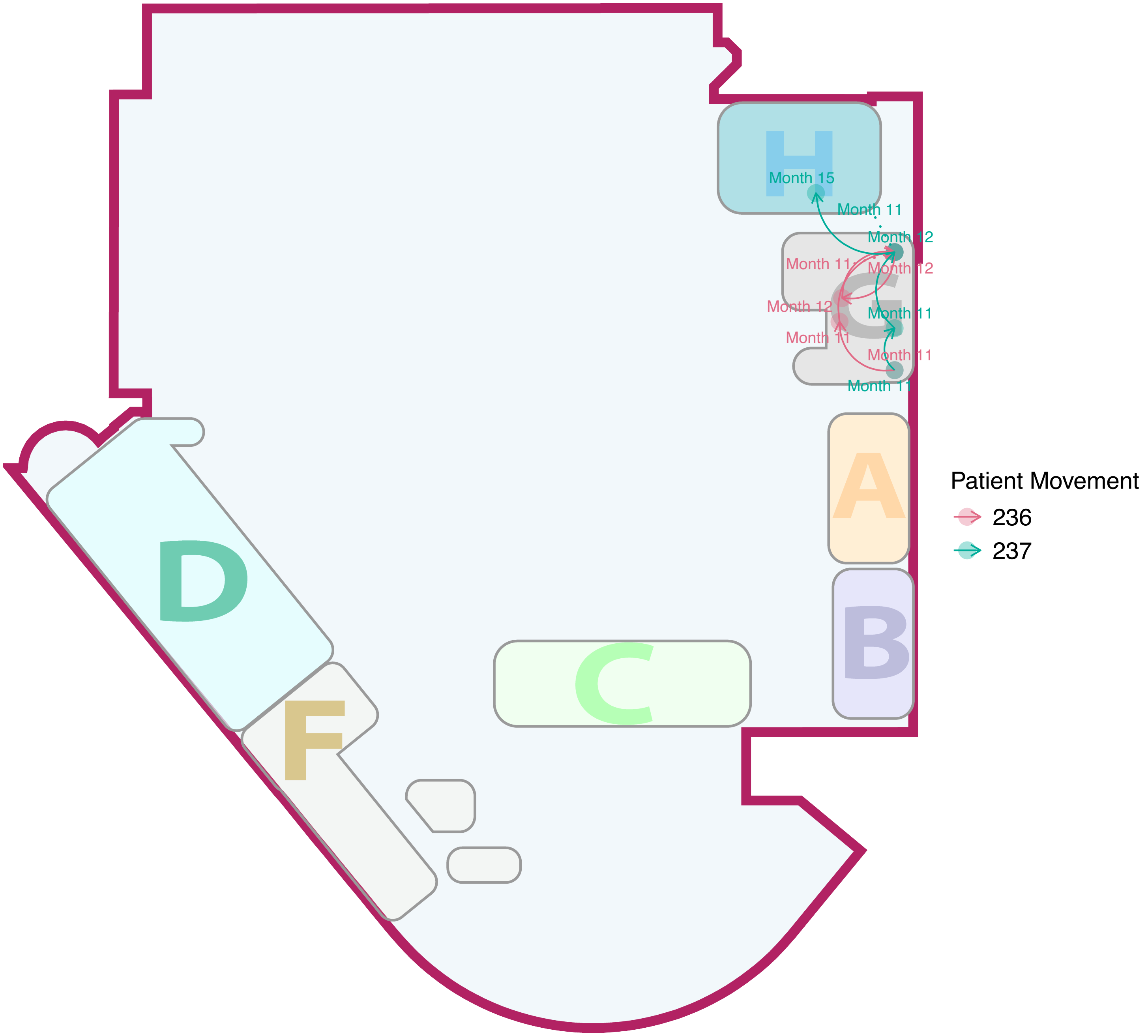

H
Month 15
Month 11
Month 12
Month 11
Month 12
Month 12
G
Month 11
Month 11
Month 11
Month 11
A
B
C
D
F
Patient Movement
236
237

### Cluster49

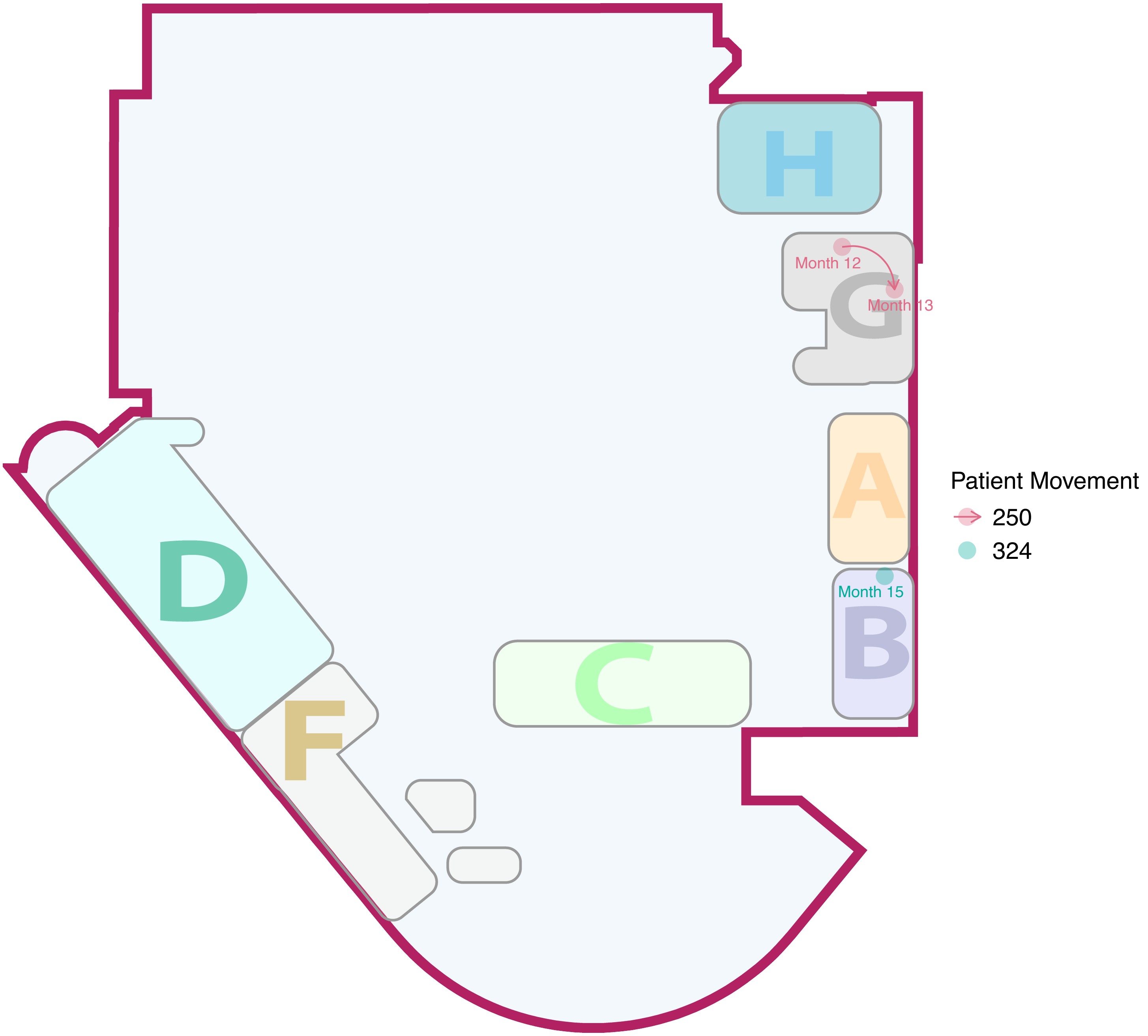

### Cluster50

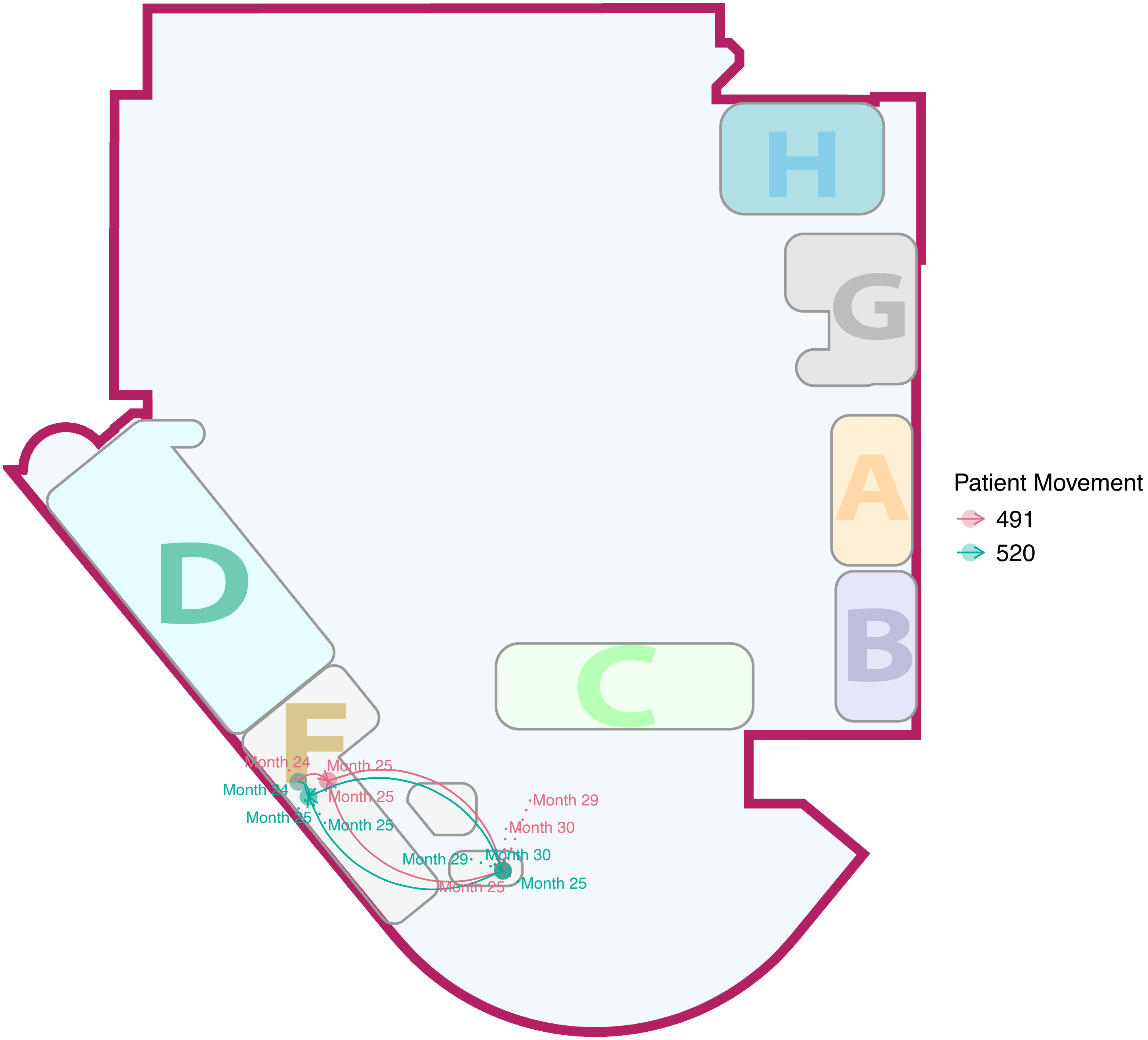

H
G
A
B
C
D
F
Month 24
Month 25
Month 25
Month 24
Month 25
Month 25
Month 29
Month 30
Month 29
Month 30
Month 25
Month 25
Patient Movement
491
520

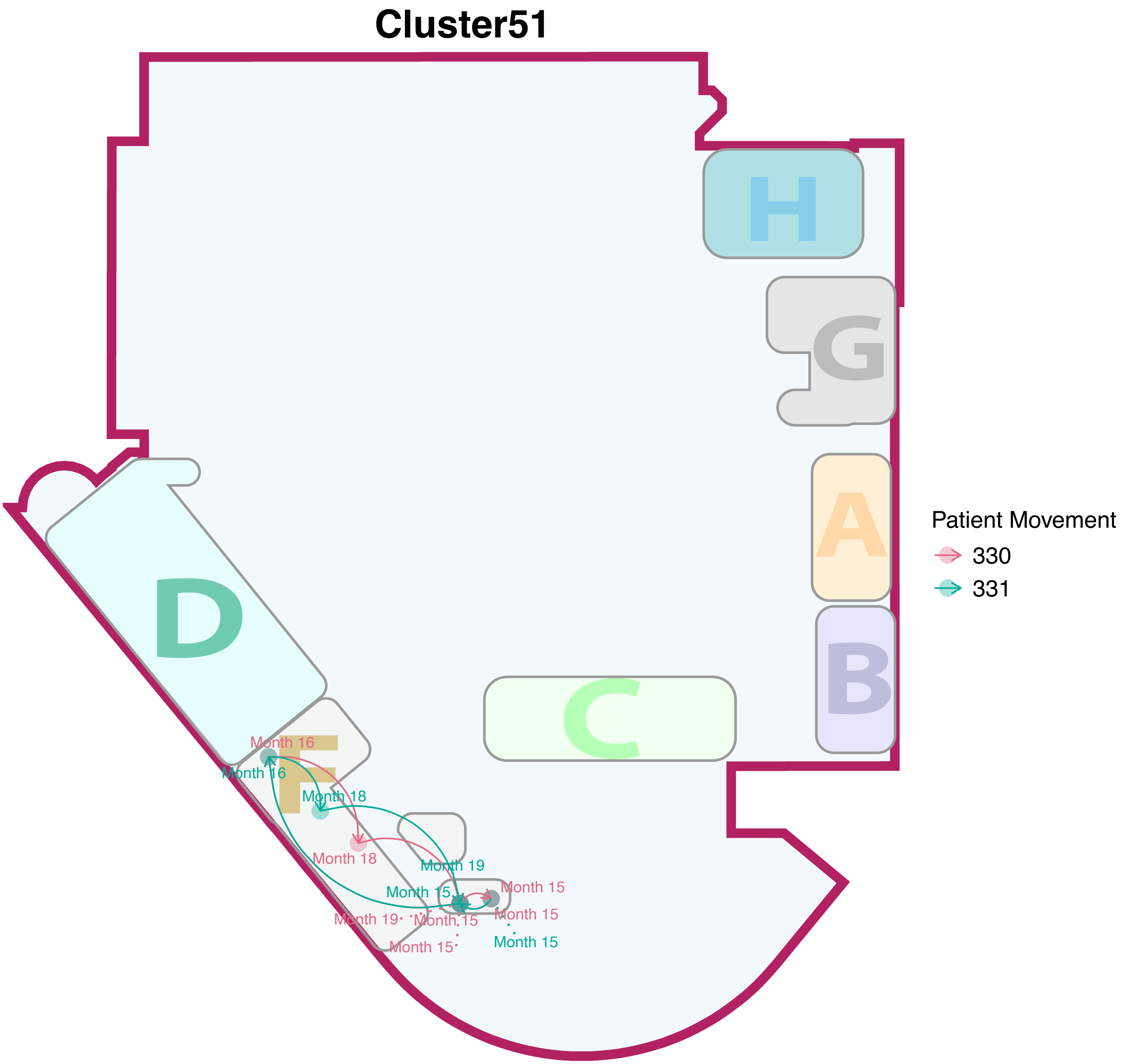

Cluster51
H
G
A
B
C
D
F
Patient Movement
330
331
Month 16
Month 16
Month 18
Month 18
Month 19
Month 15
Month 15
Month 19
Month 15
Month 15
Month 15
Month 15

### Cluster52

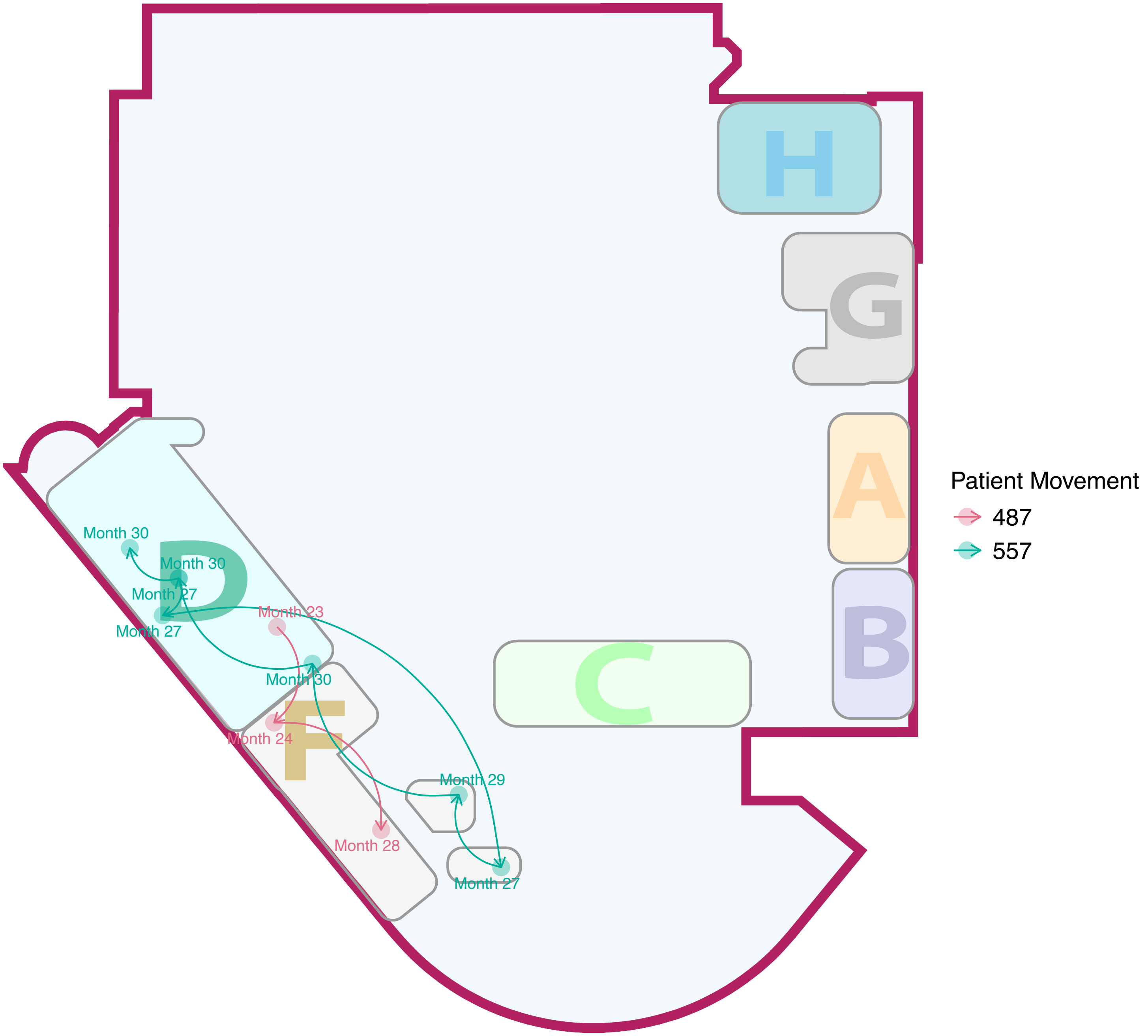

H
G
A
B
C
D
F
Month 30
Month 30
Month 27
Month 27
Month 23
Month 30
Month 24
Month 29
Month 28
Month 27
Patient Movement
487
557

#### Cluster53

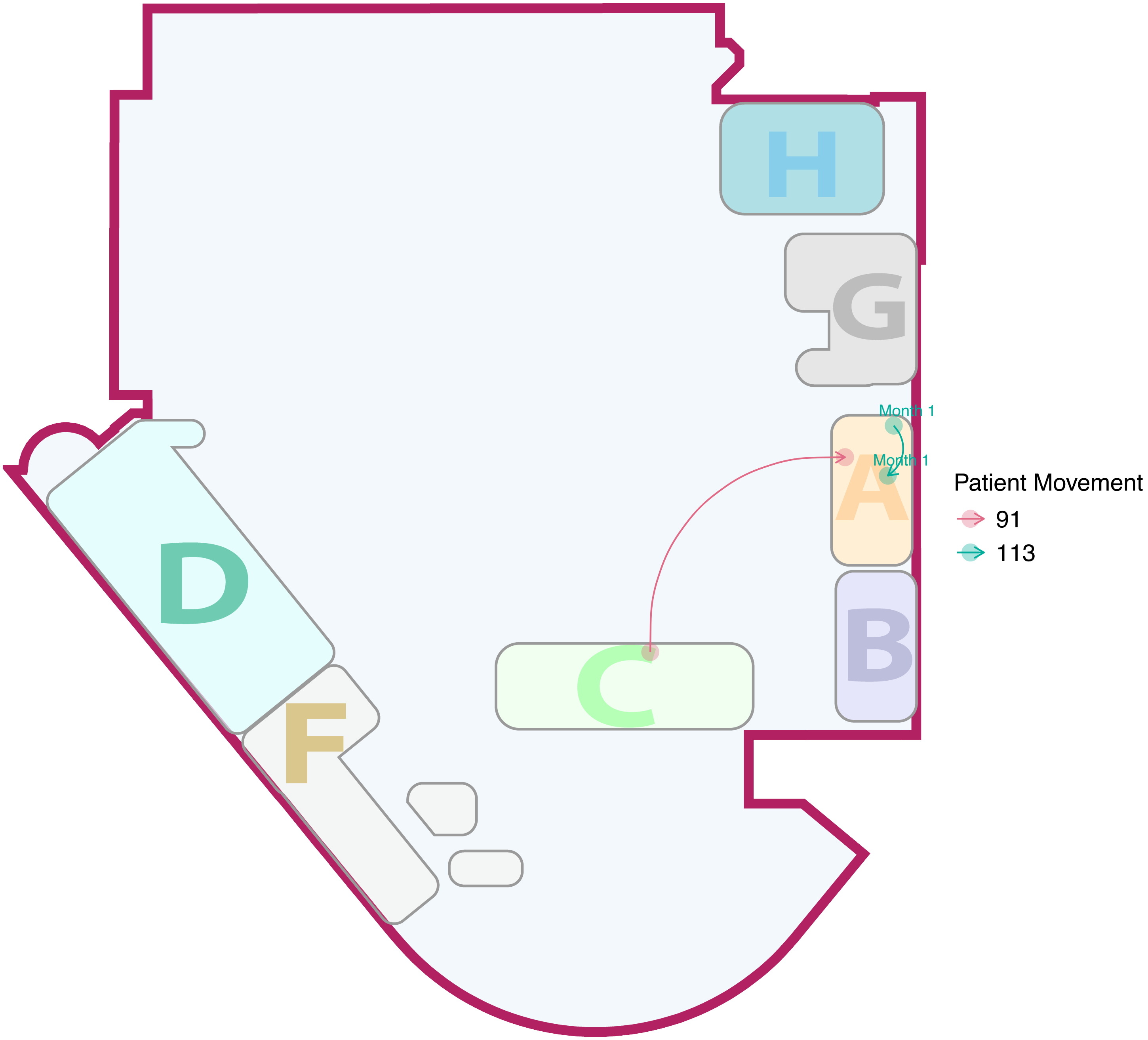

### Cluster54

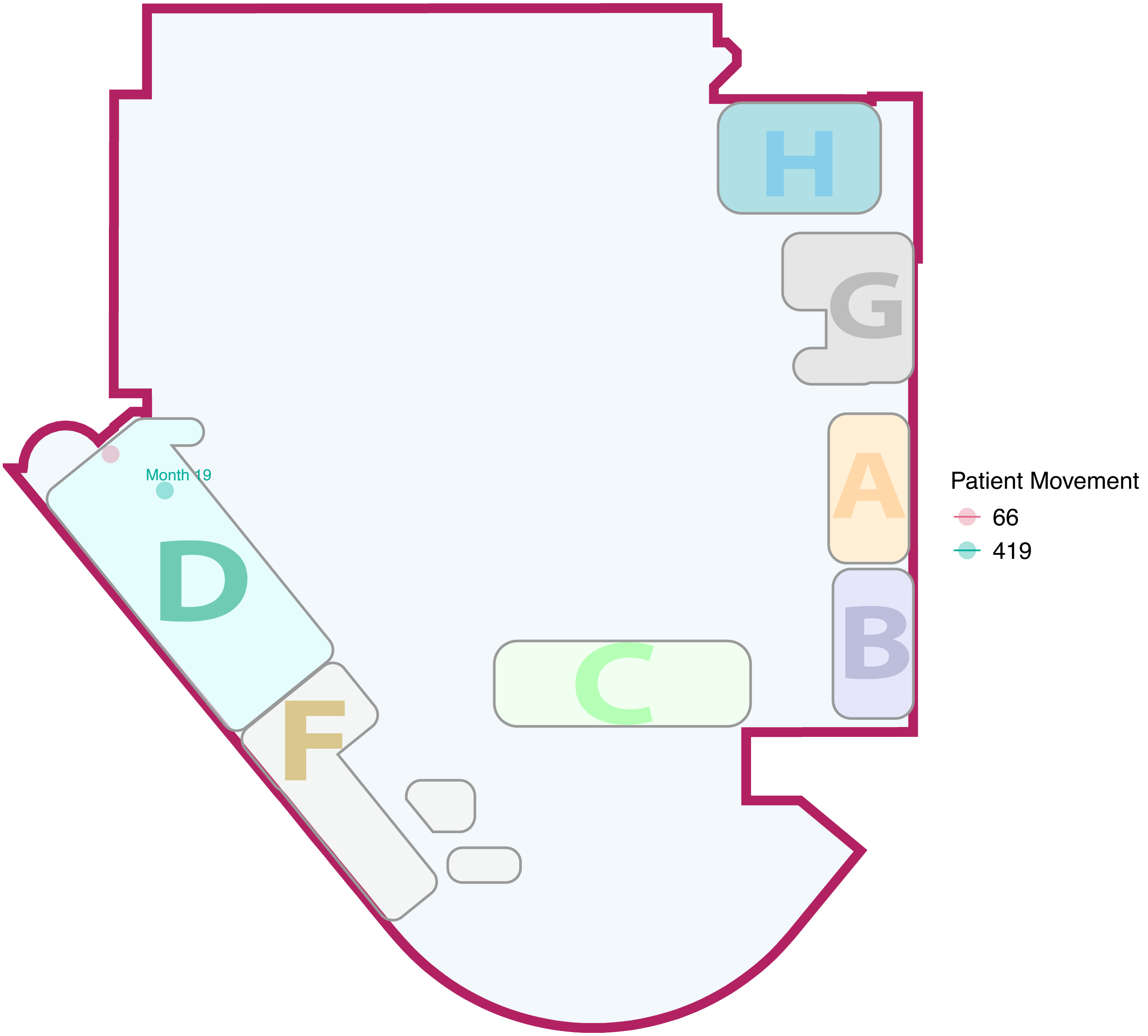

### Cluster55

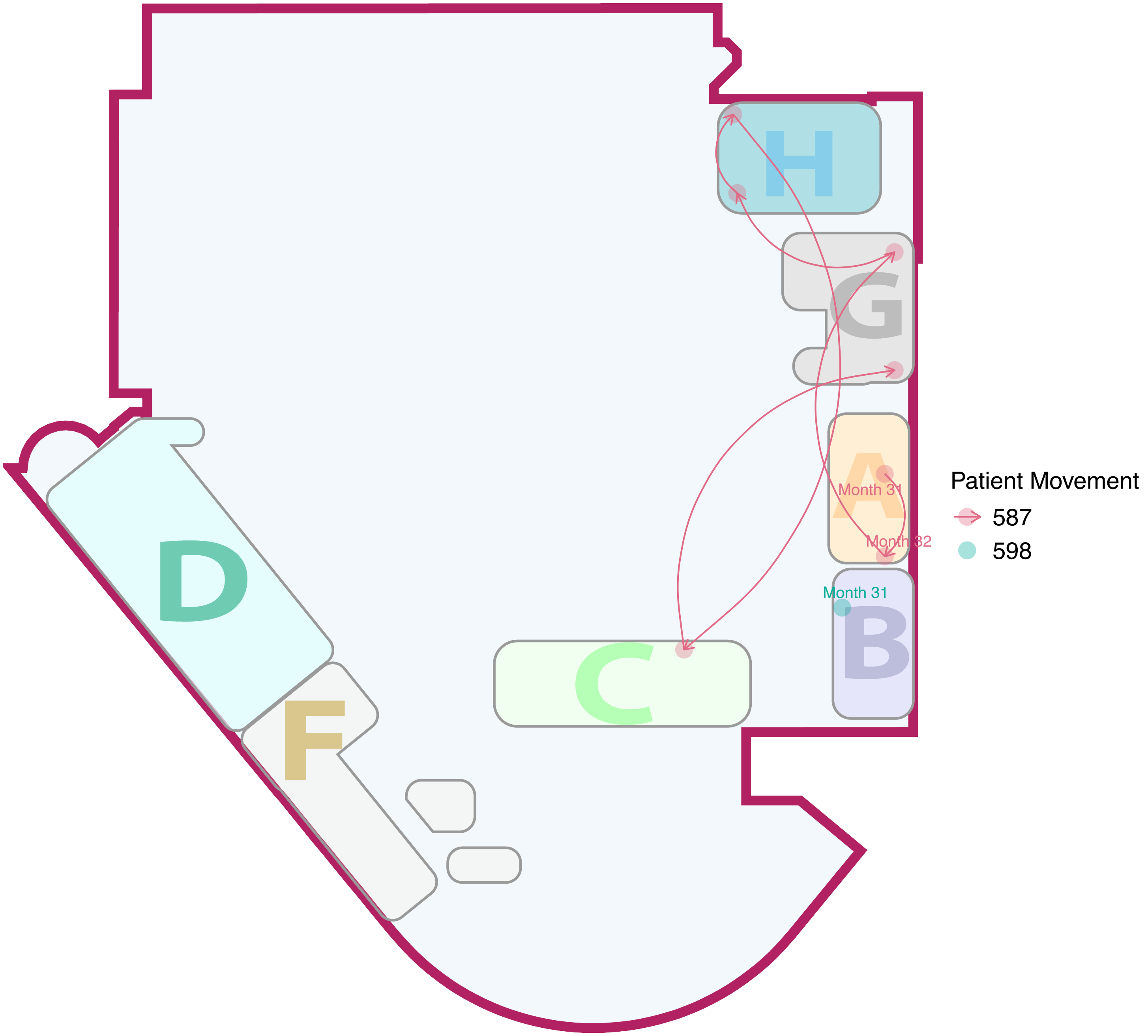

### Cluster56

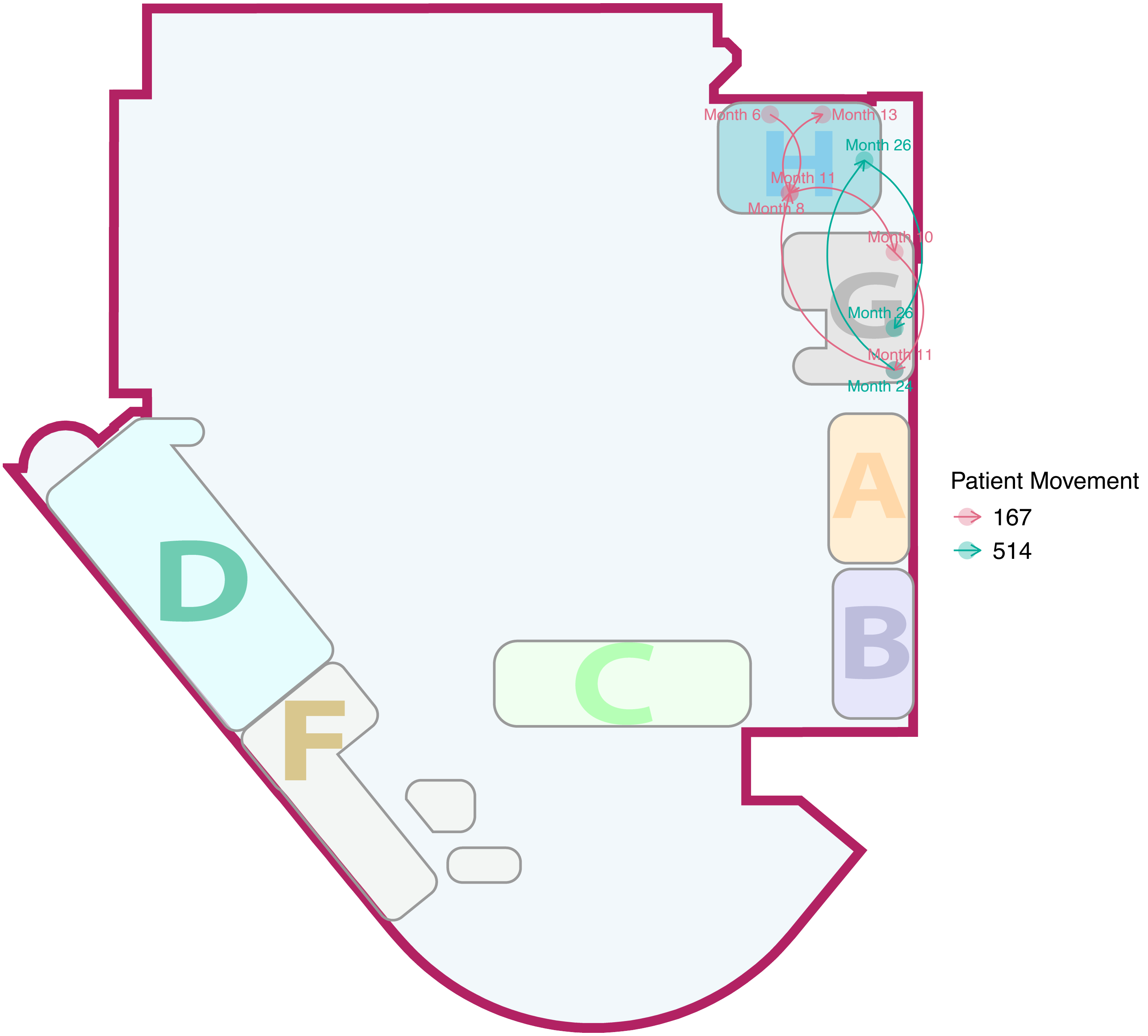

Month 6
Month 13
Month 26
Month 11
Month 8
Month 10
Month 26
Month 11
Month 24
H
G
A
B
C
D
F
Patient Movement
167
514

#### Cluster57

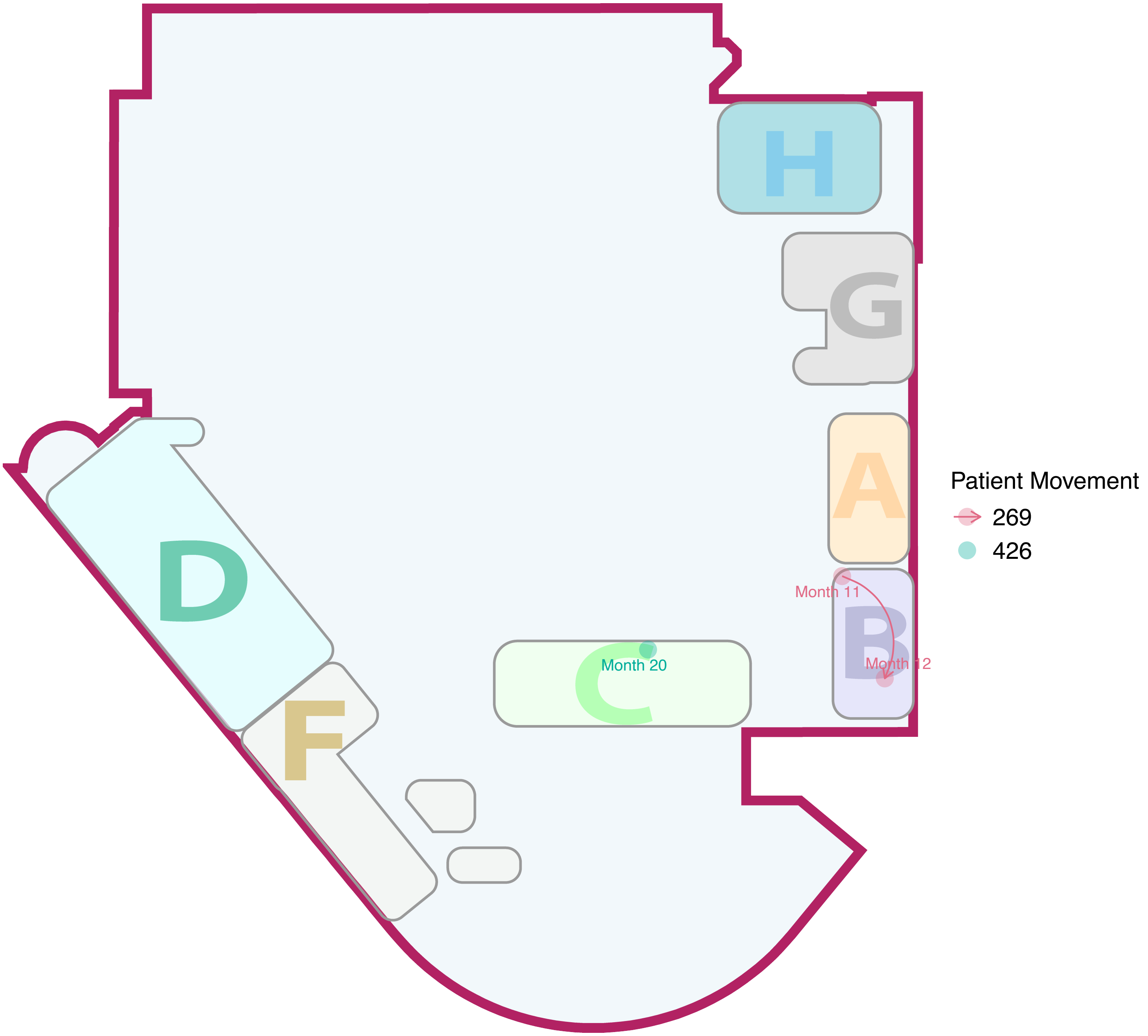

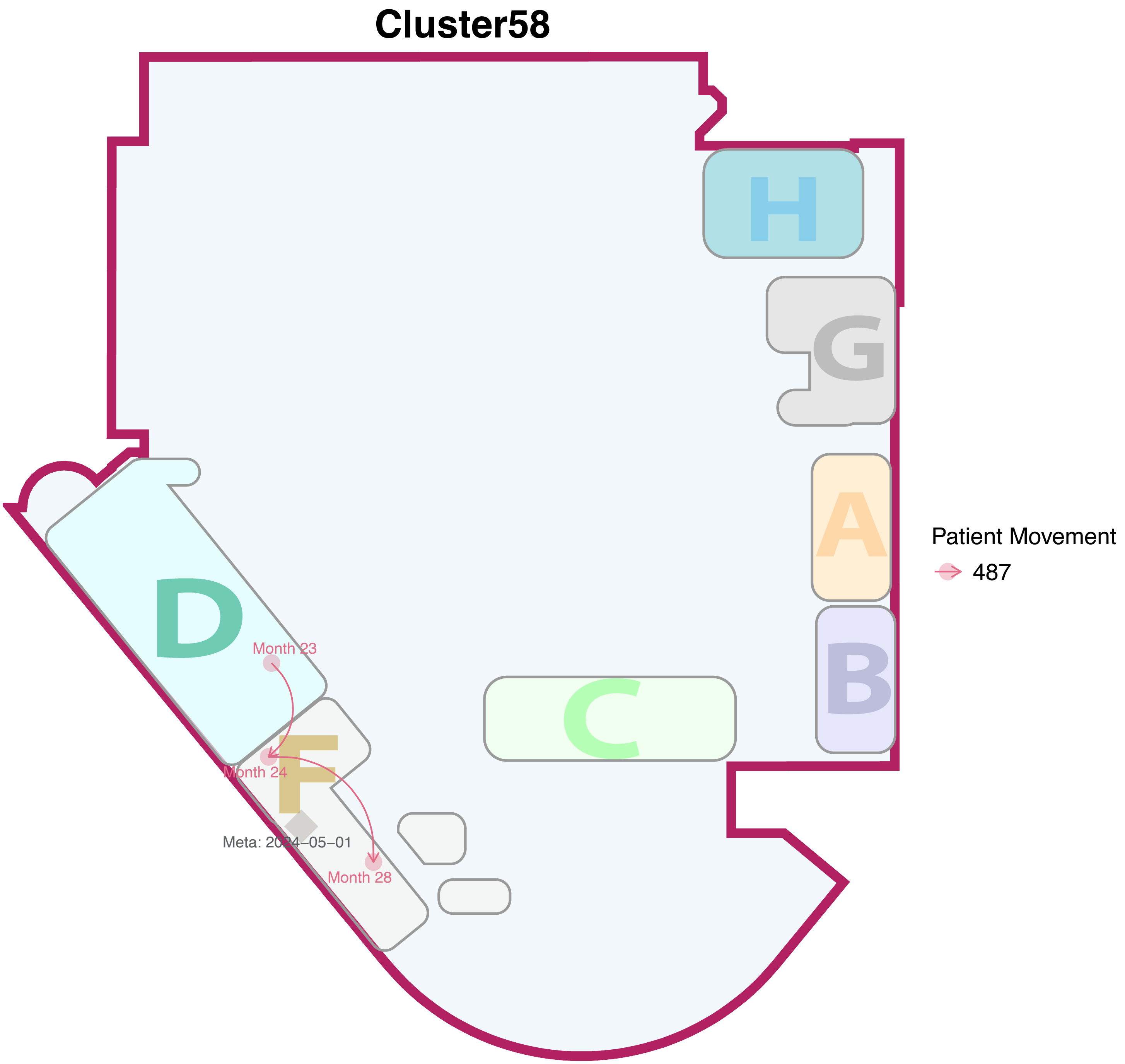

Cluster58
H
G
A
B
C
D
F
Month 23
Month 24
Month 28
Meta: 2024−05−01
Patient Movement
487

### Cluster59

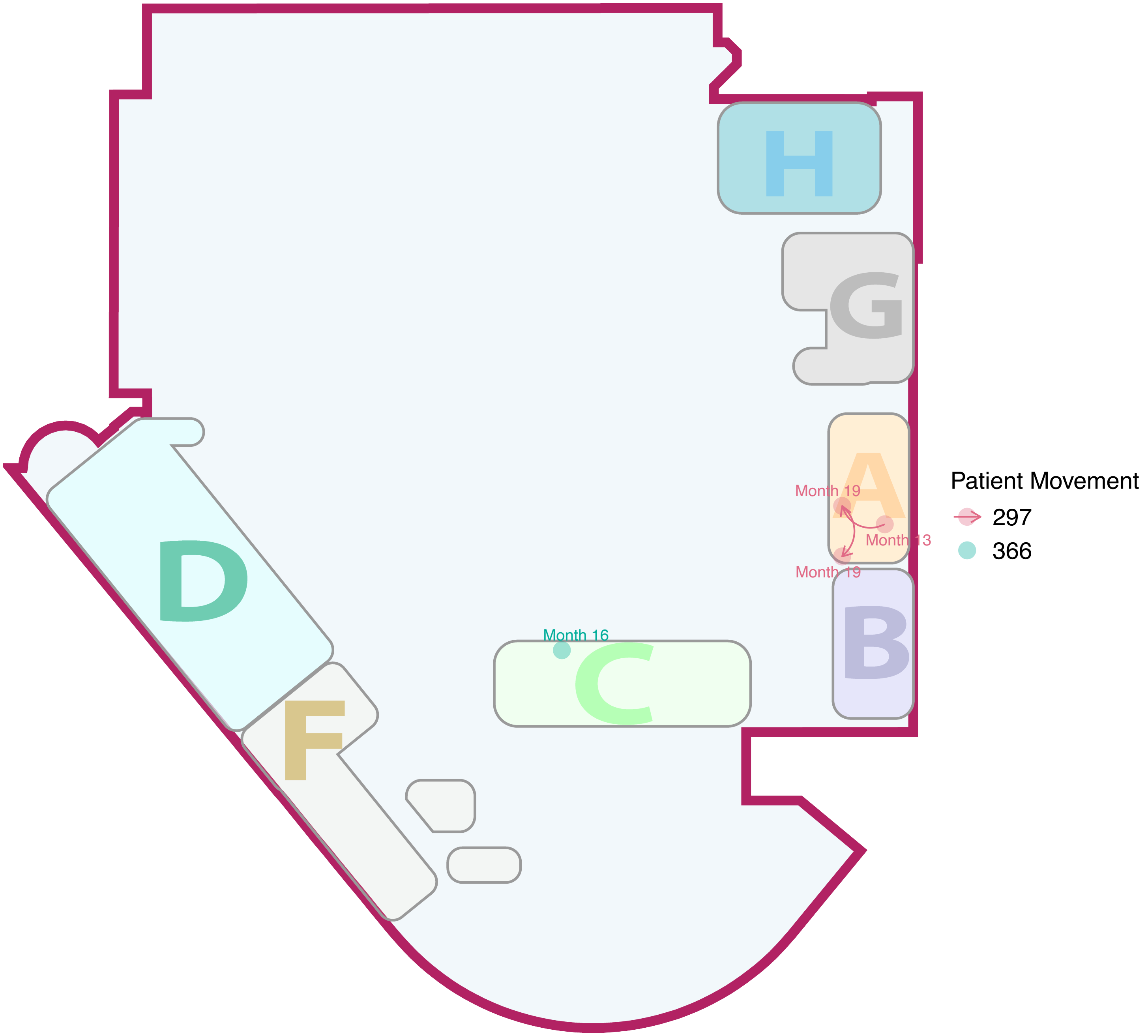

H
G
A
B
C
D
F
Month 19
Month 13
Month 19
Month 16
Patient Movement
297
366

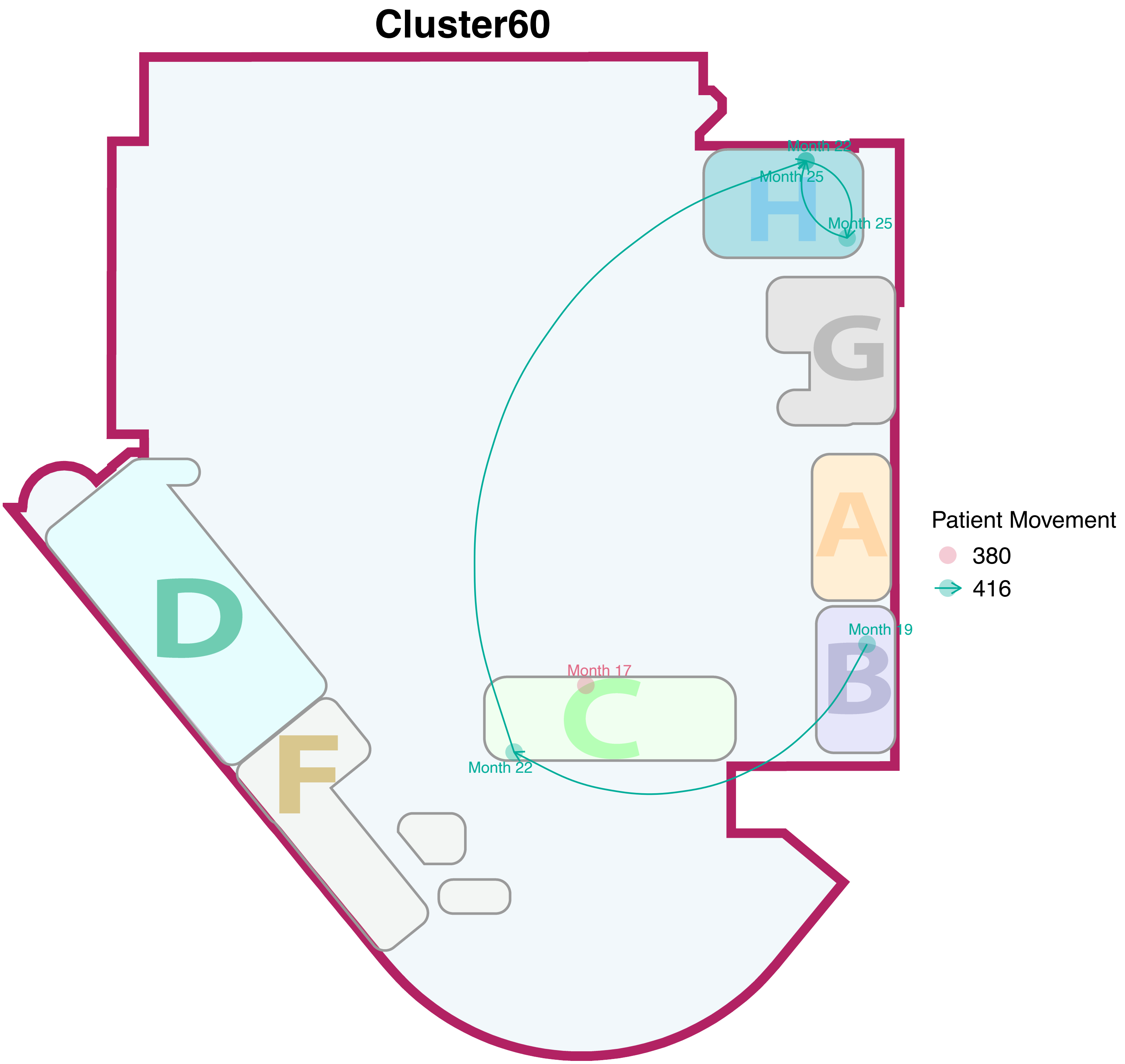
Cluster60
Month 22
Month 25
Month 25
H
G
A
Patient Movement
380
416
Month 19
B
Month 17
C
Month 22
D
F

### Cluster61

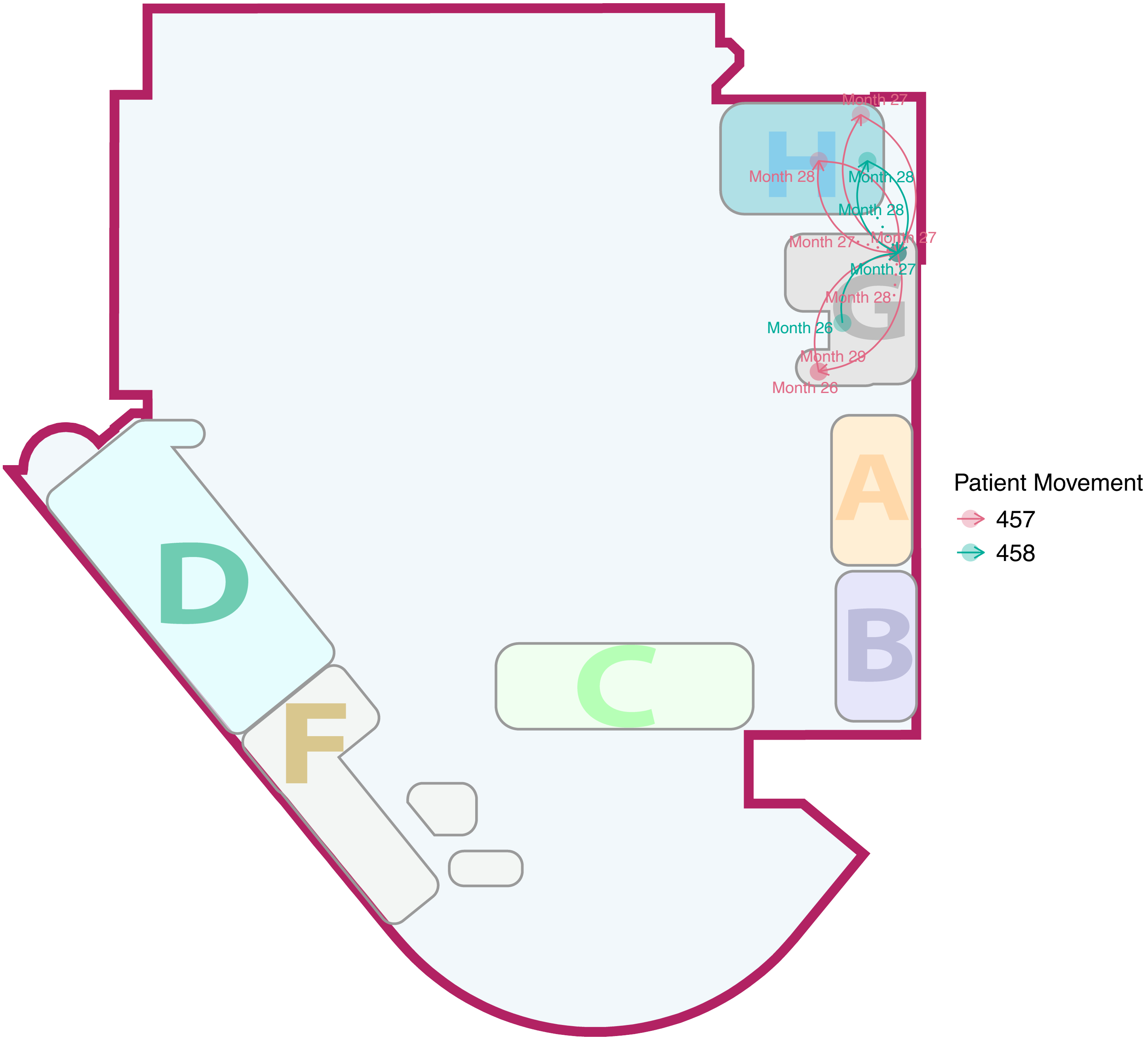

Month 27
Month 28
Month 28
Month 28
Month 27
Month 27
Month 27
Month 28
Month 26
Month 29
Month 26
H
G
A
B
C
D
F
Patient Movement
457
458

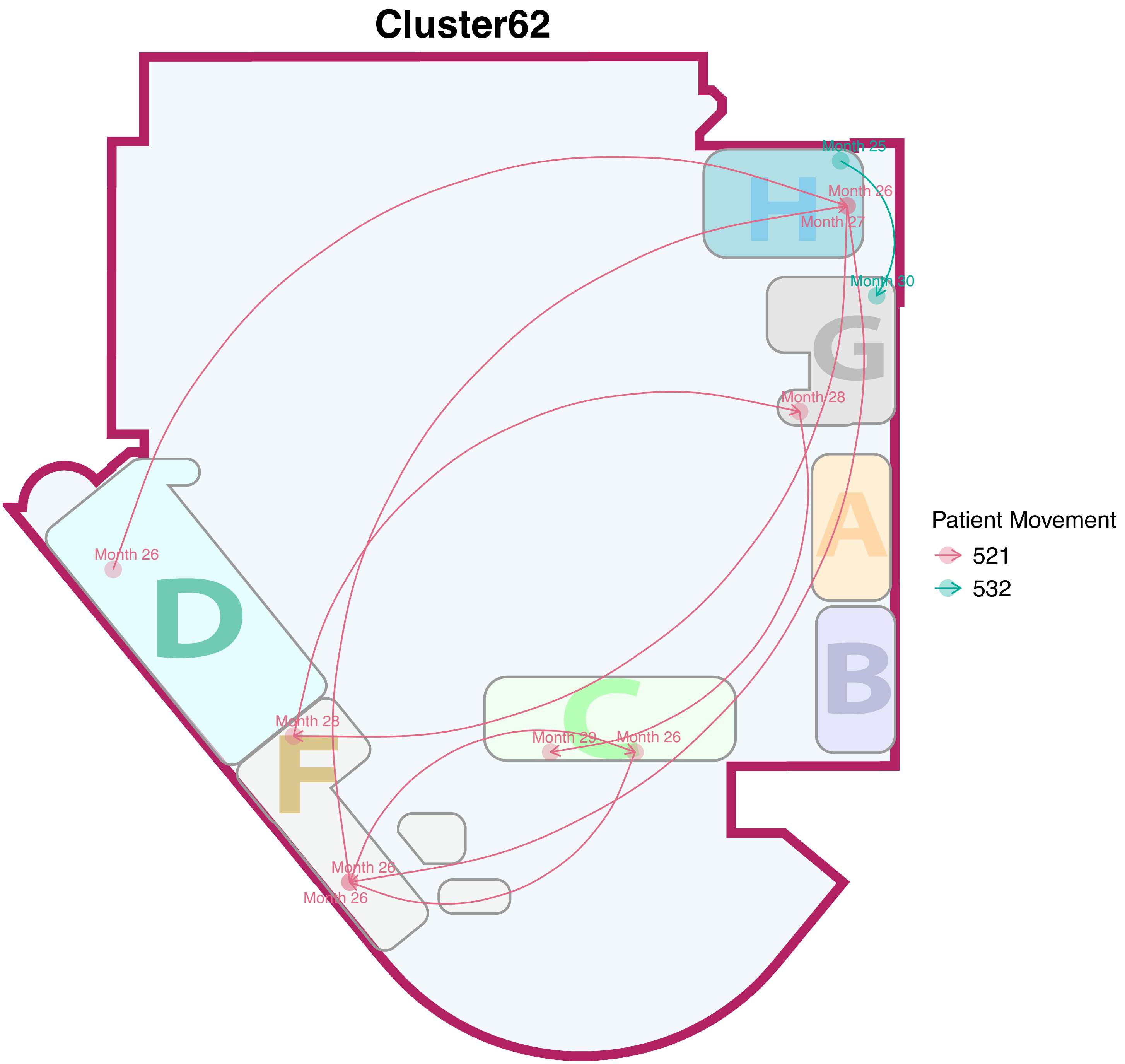

Cluster62
H
G
A
B
C
D
F
Month 25
Month 26
Month 27
Month 30
Month 28
Month 26
Month 28
Month 29
Month 26
Month 26
Month 26
Patient Movement
521
532

### Cluster63

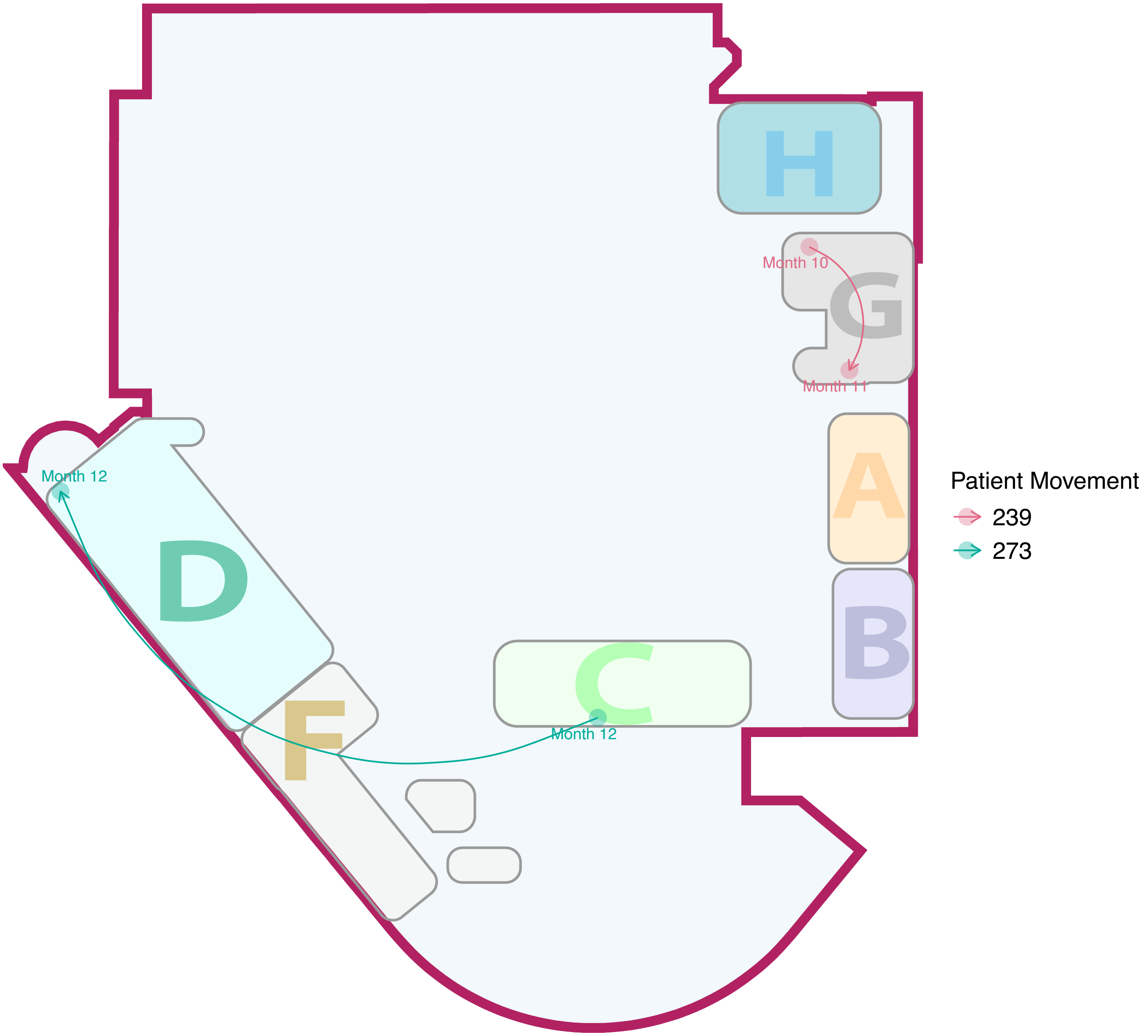

H
G
Month 10
Month 11
A
B
C
D
F
Month 12
Month 12
Patient Movement
239
273

#### Cluster64

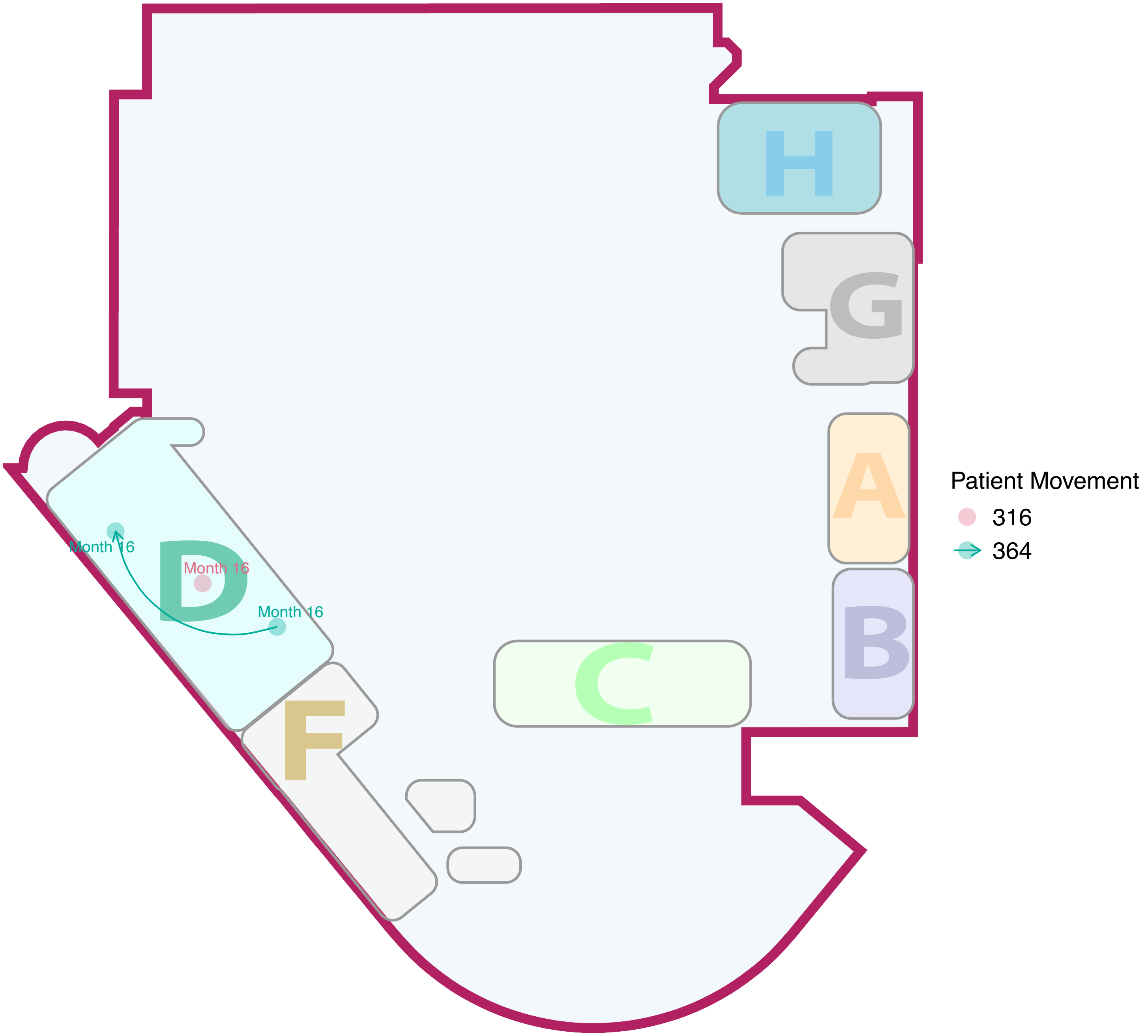

#### Cluster65

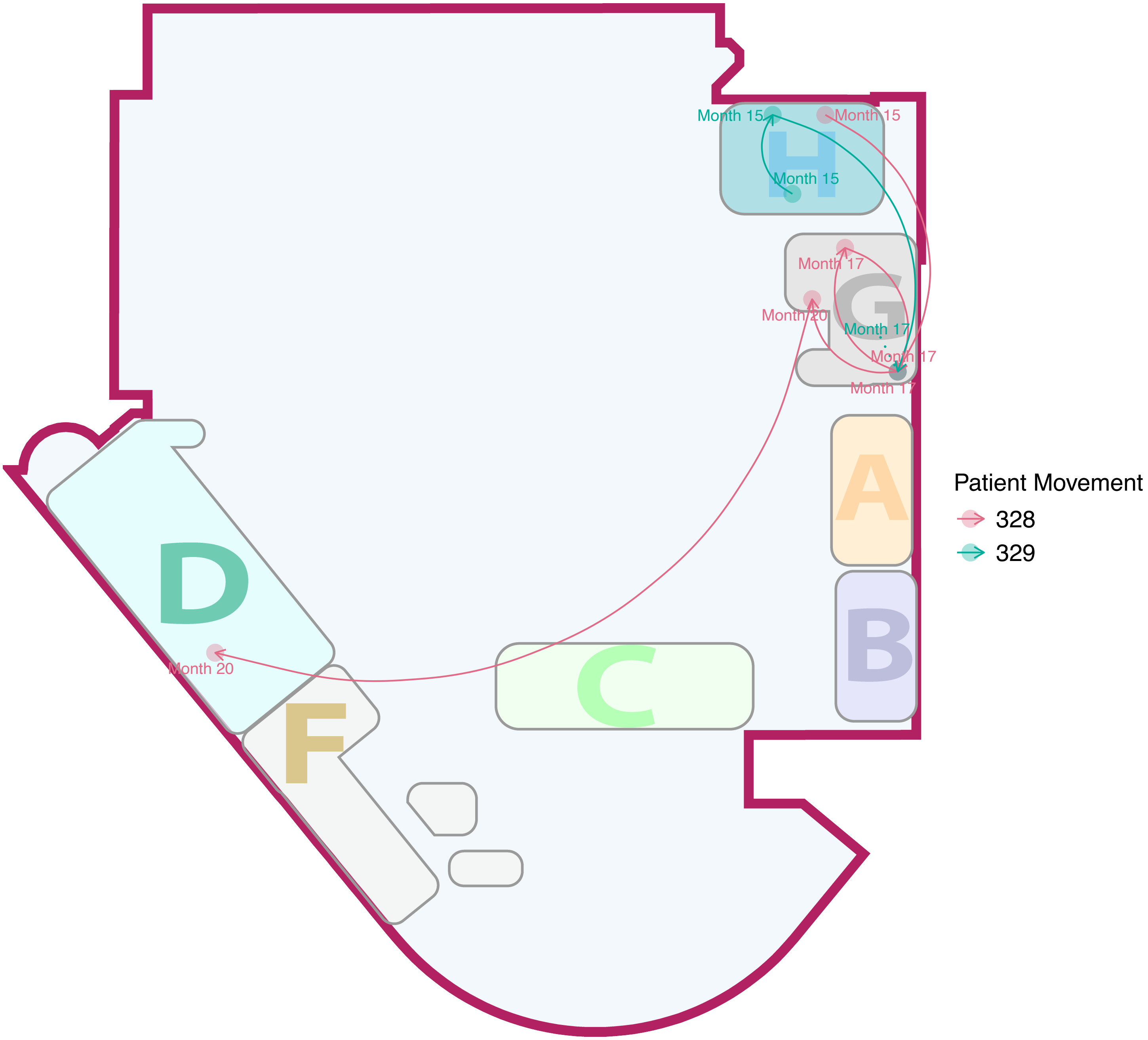

H
G
A
B
C
D
F
Month 15
Month 15
Month 15
Month 17
Month 20
Month 17
Month 17
Month 17
Month 20
Patient Movement
328
329

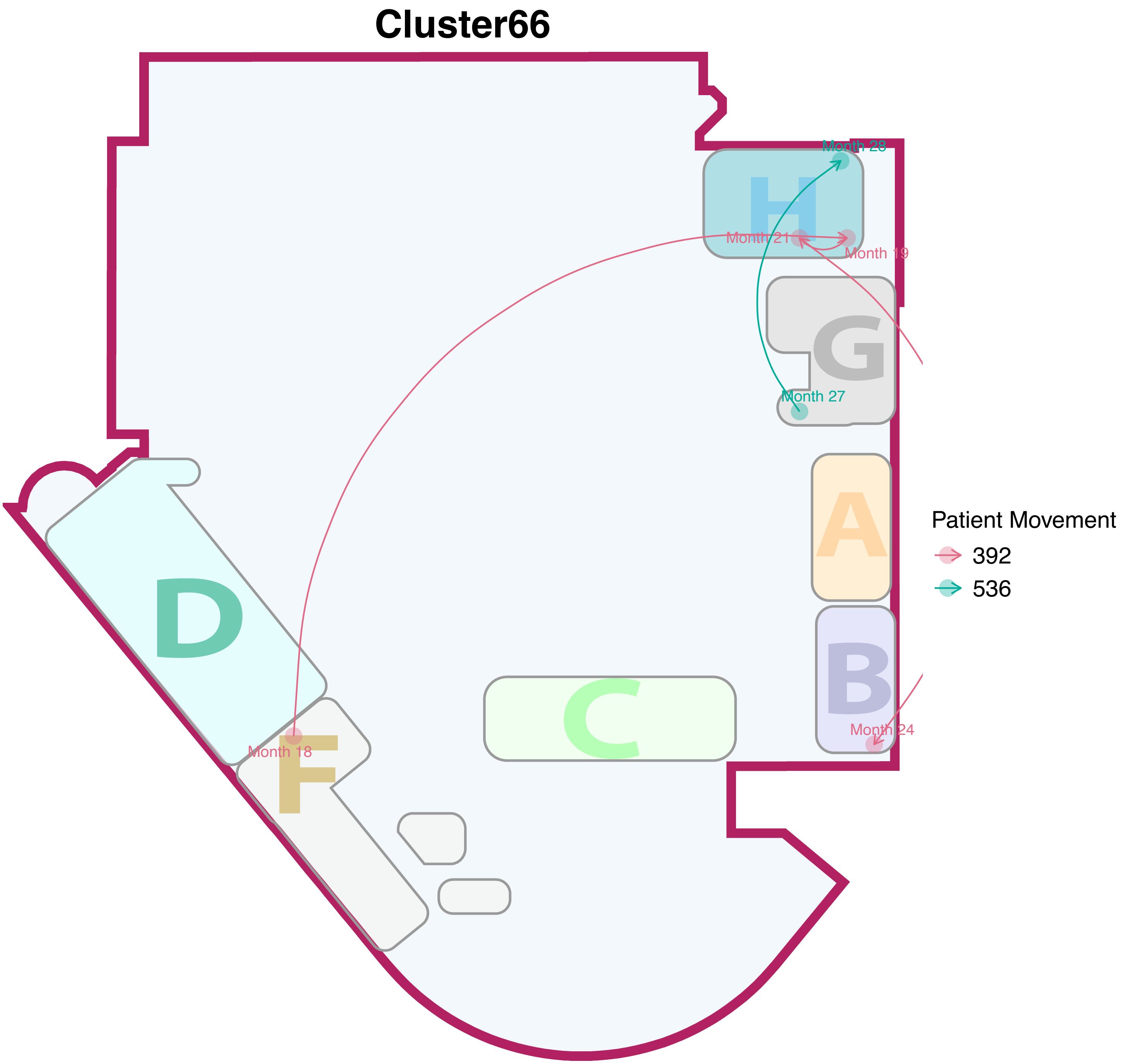
Cluster66
Month 28
H
Month 21
Month 19
G
Month 27
A
Patient Movement
392
536
D
B
Month 24
C
Month 18
F

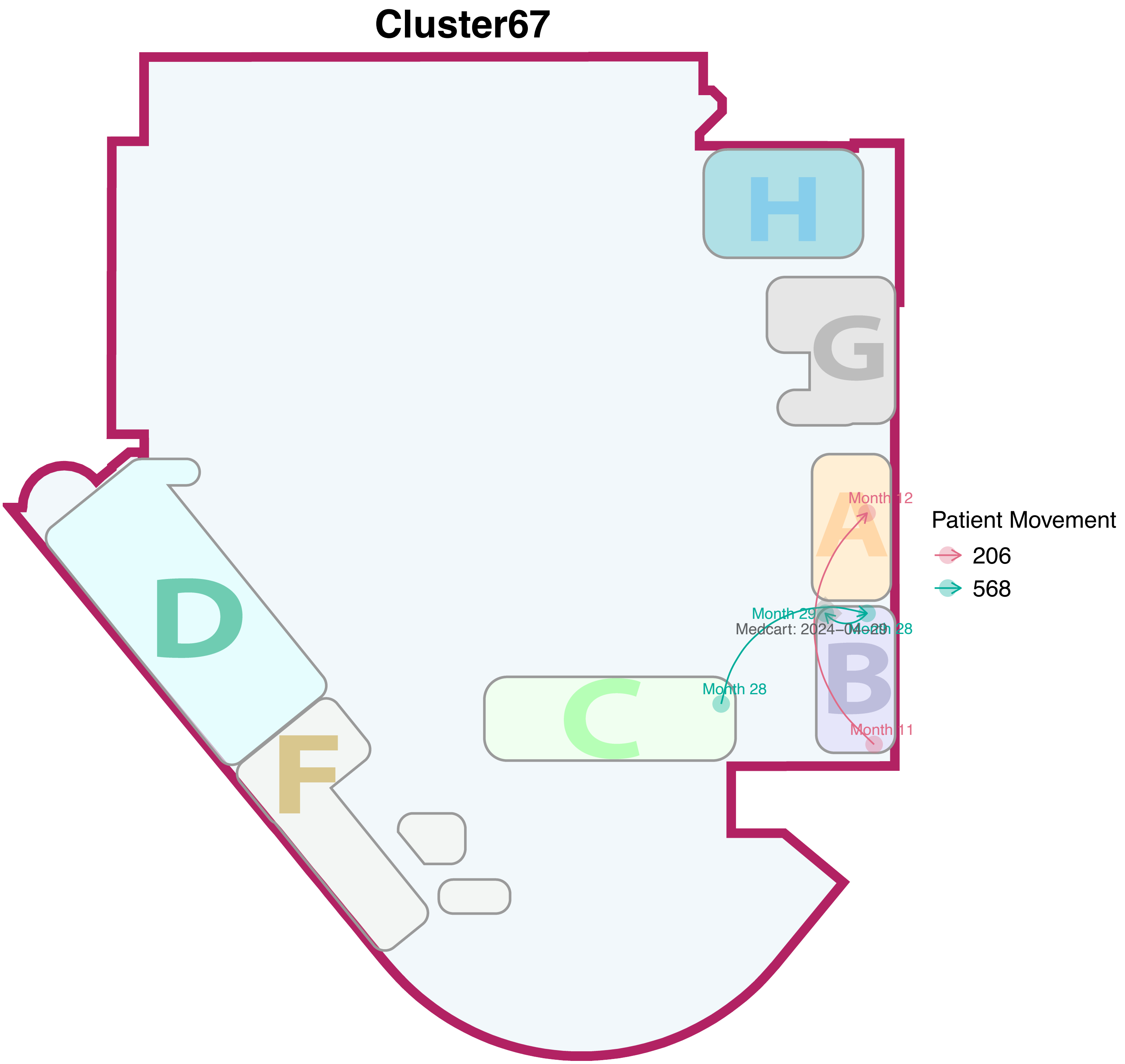
Cluster67
H
G
A
B
C
D
F
Month 12
Month 29
Medcart: 2024–04–29
Month 28
Month 28
Month 11
Patient Movement
206
568

### Cluster68

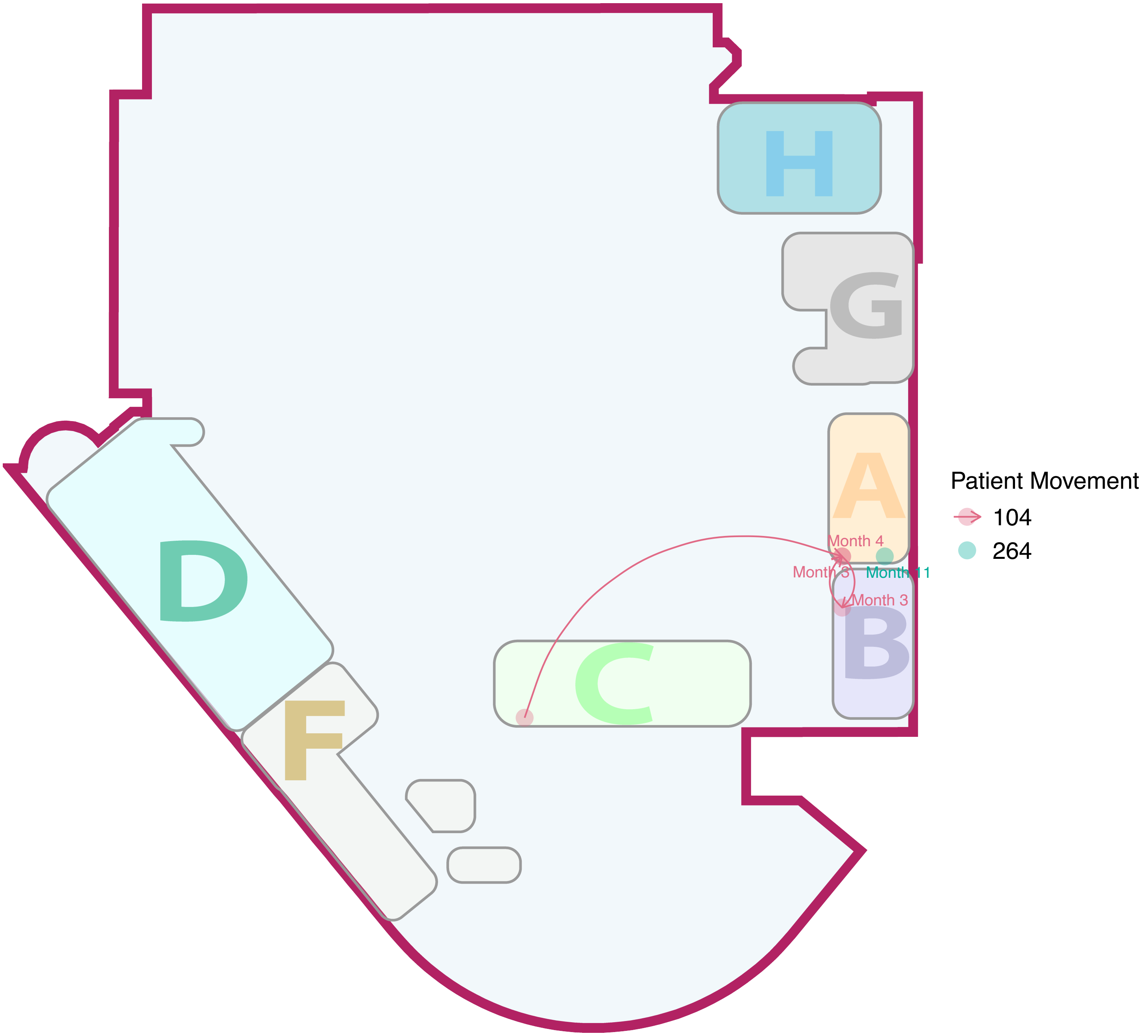

H
G
A
D
C
B
F
Month 4
Month 3
Month 11
Month 3
Patient Movement
104
264

### Cluster69

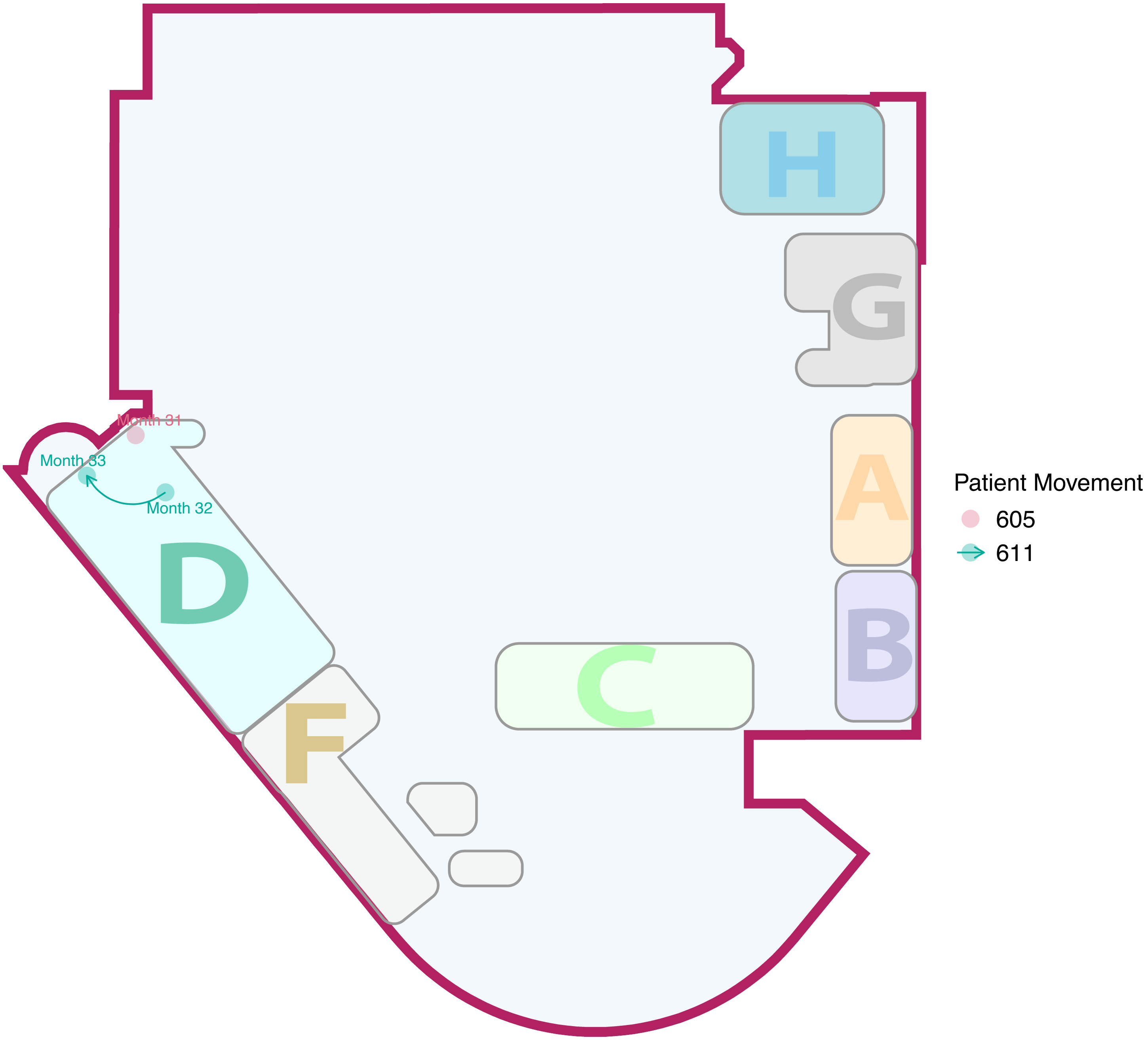

Figure S4: Temporal analysis for each identified transmission cluster by location

Each bar represents a single patient's NICU stay, with colored rectangles indicating different NICU sections. Admission dates are marked with triangles and discharge dates with squares. X-axis (Months Before End of Study) represents the number of months prior to completion of study period.

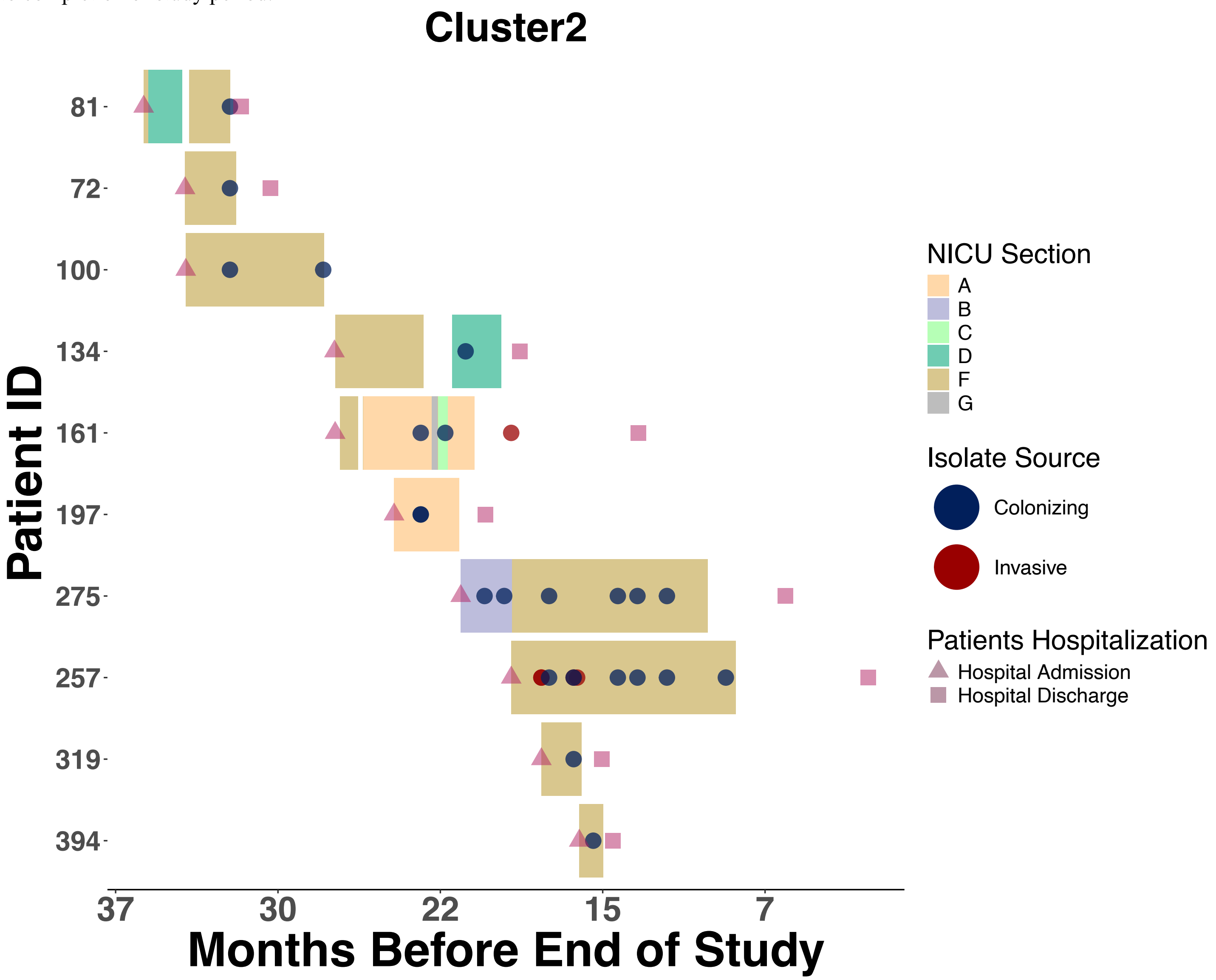

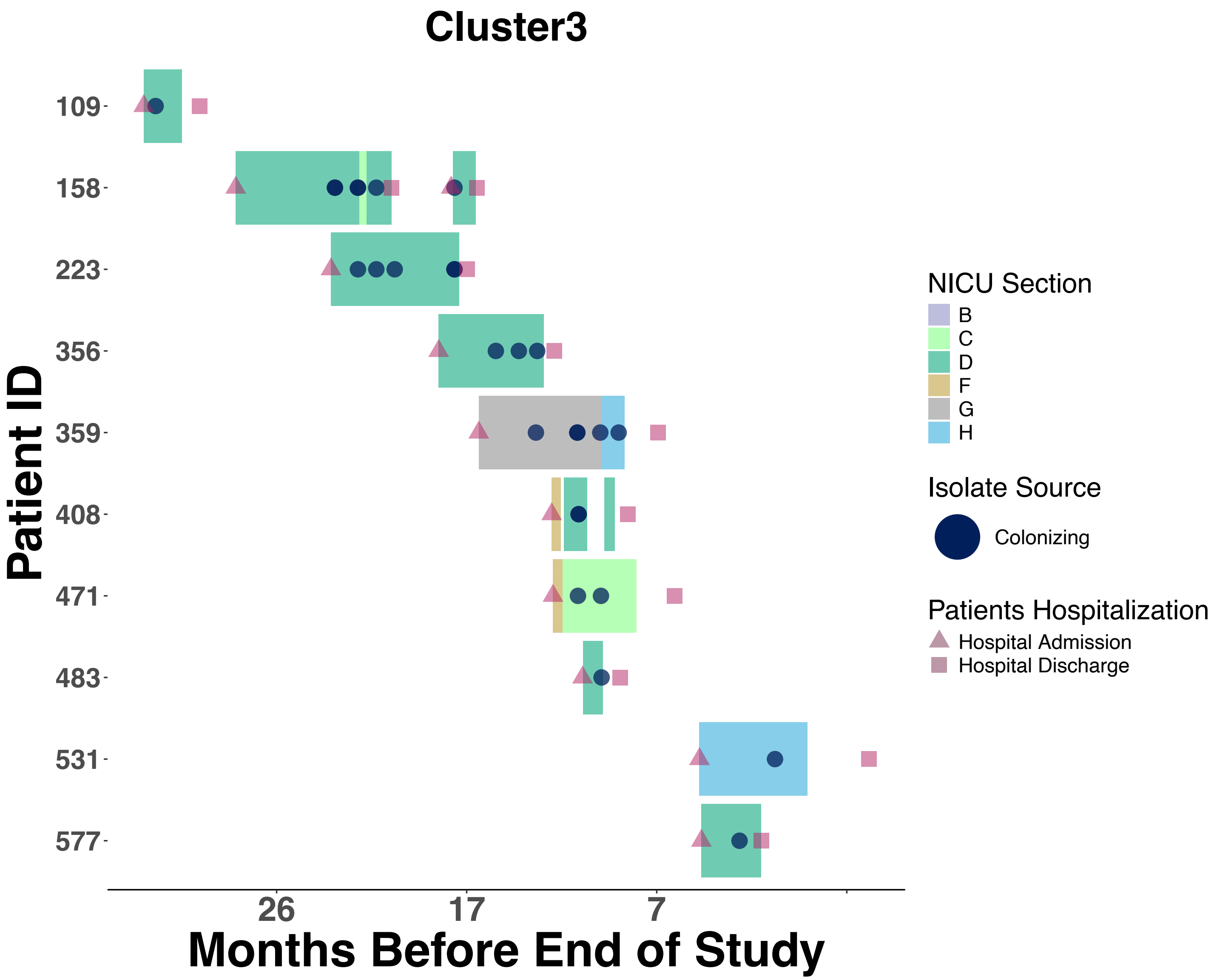
Cluster3
Patient ID
109
158
223
356
359
408
471
483
531
577
26
17
7
Months Before End of Study
NICU Section
B
C
D
F
G
H
Isolate Source
Colonizing
Patients Hospitalization
Hospital Admission
Hospital Discharge

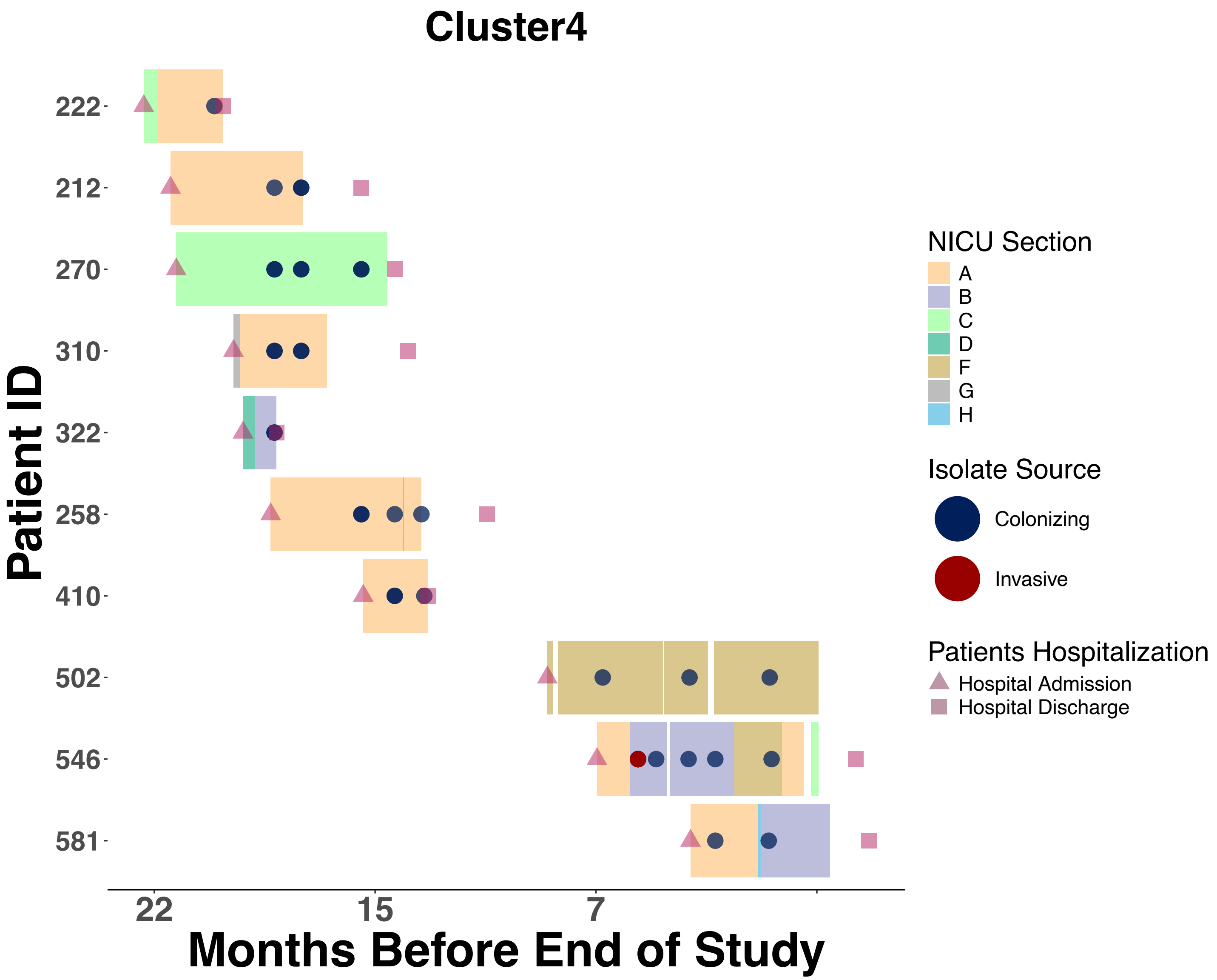

Cluster4
Patient ID
222
212
270
310
322
258
410
502
546
581
22
15
7
Months Before End of Study
NICU Section
A
B
C
D
F
G
H
Isolate Source
Colonizing
Invasive
Patients Hospitalization
Hospital Admission
Hospital Discharge

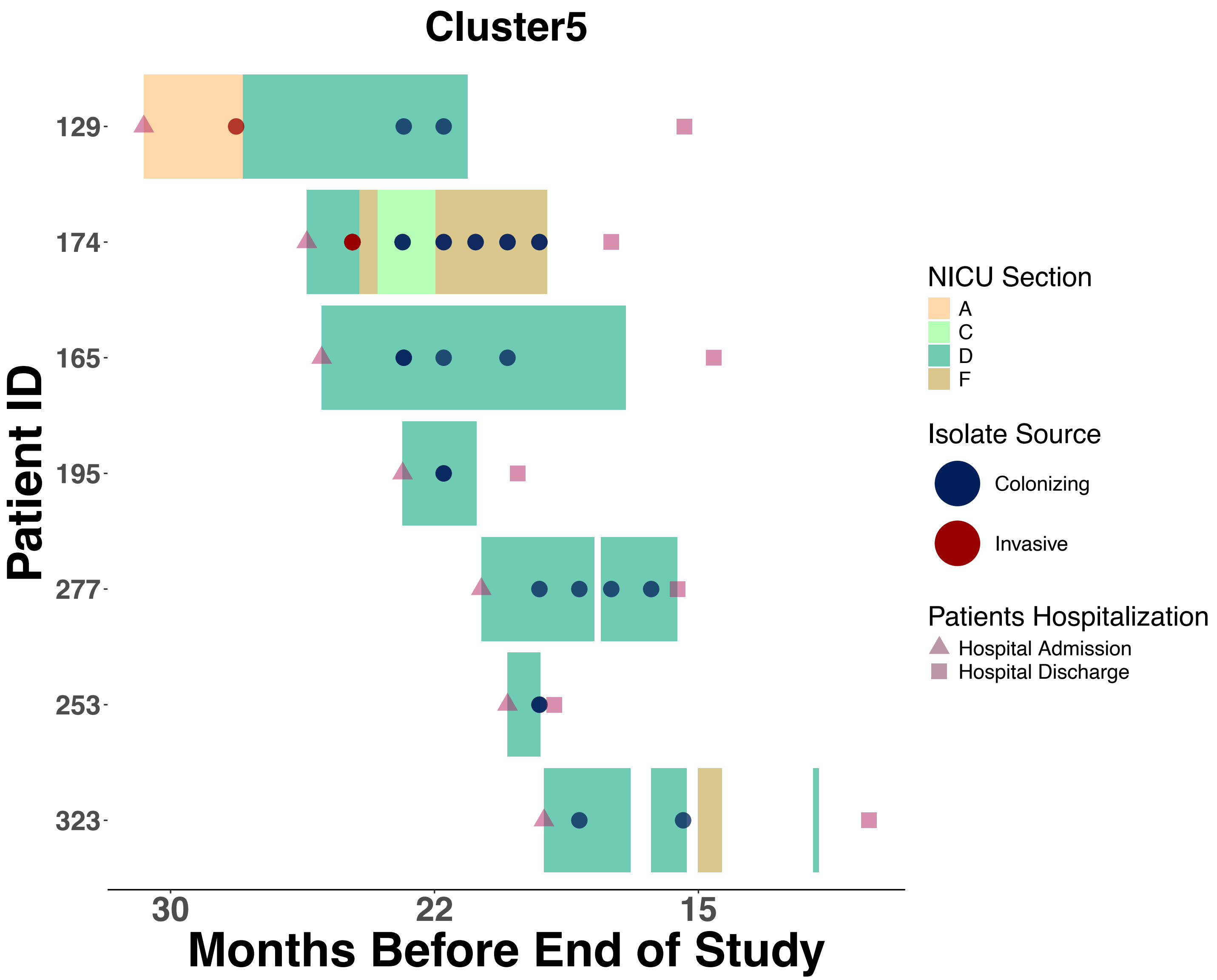
Cluster5
Patient ID
129
174
165
195
277
253
323
30
22
15
Months Before End of Study
NICU Section
A
C
D
F
Isolate Source
Colonizing
Invasive
Patients Hospitalization
Hospital Admission
Hospital Discharge

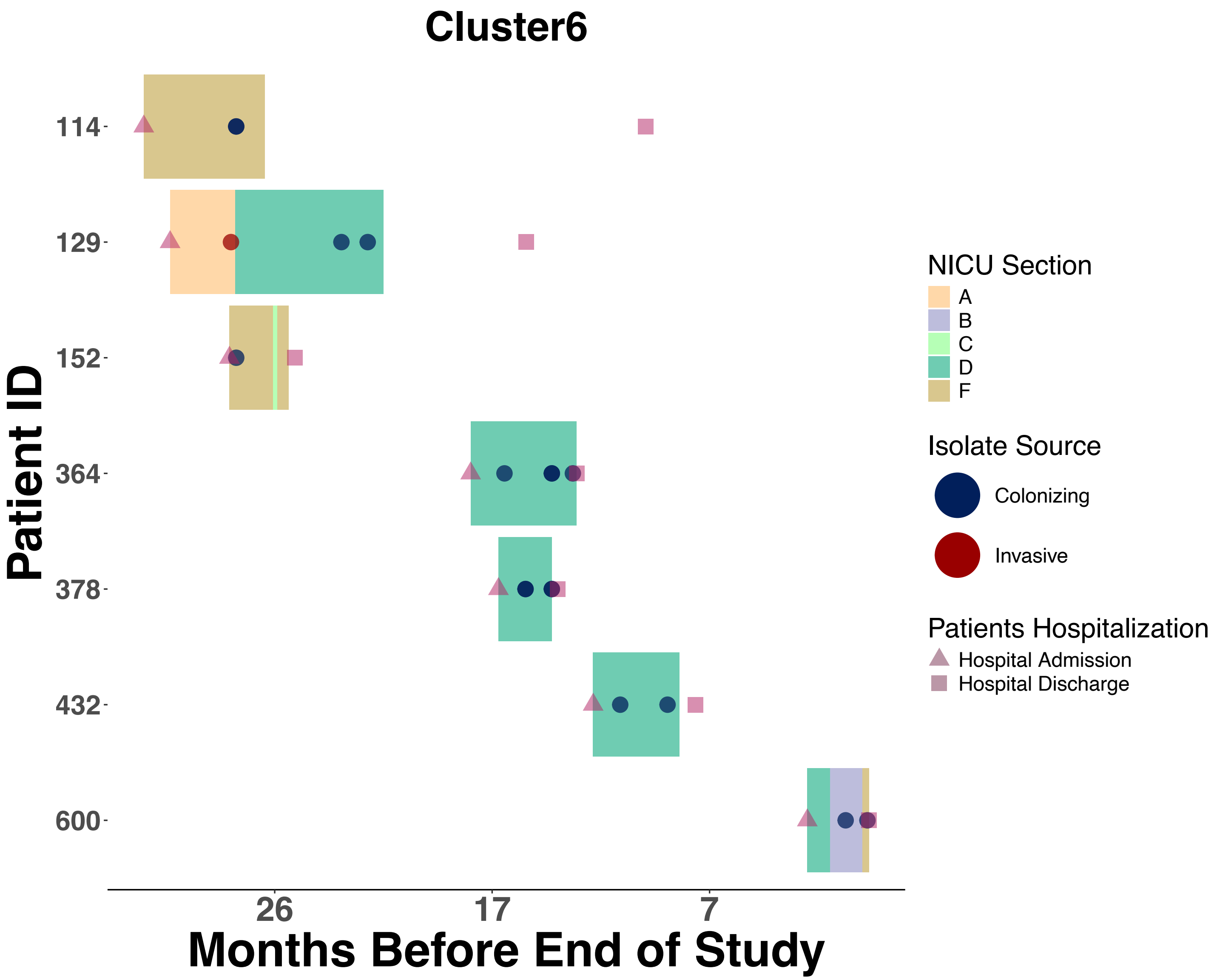

Cluster6
Patient ID
114
129
152
364
378
432
600
26
17
7
Months Before End of Study
NICU Section
A
B
C
D
F
Isolate Source
Colonizing
Invasive
Patients Hospitalization
Hospital Admission
Hospital Discharge

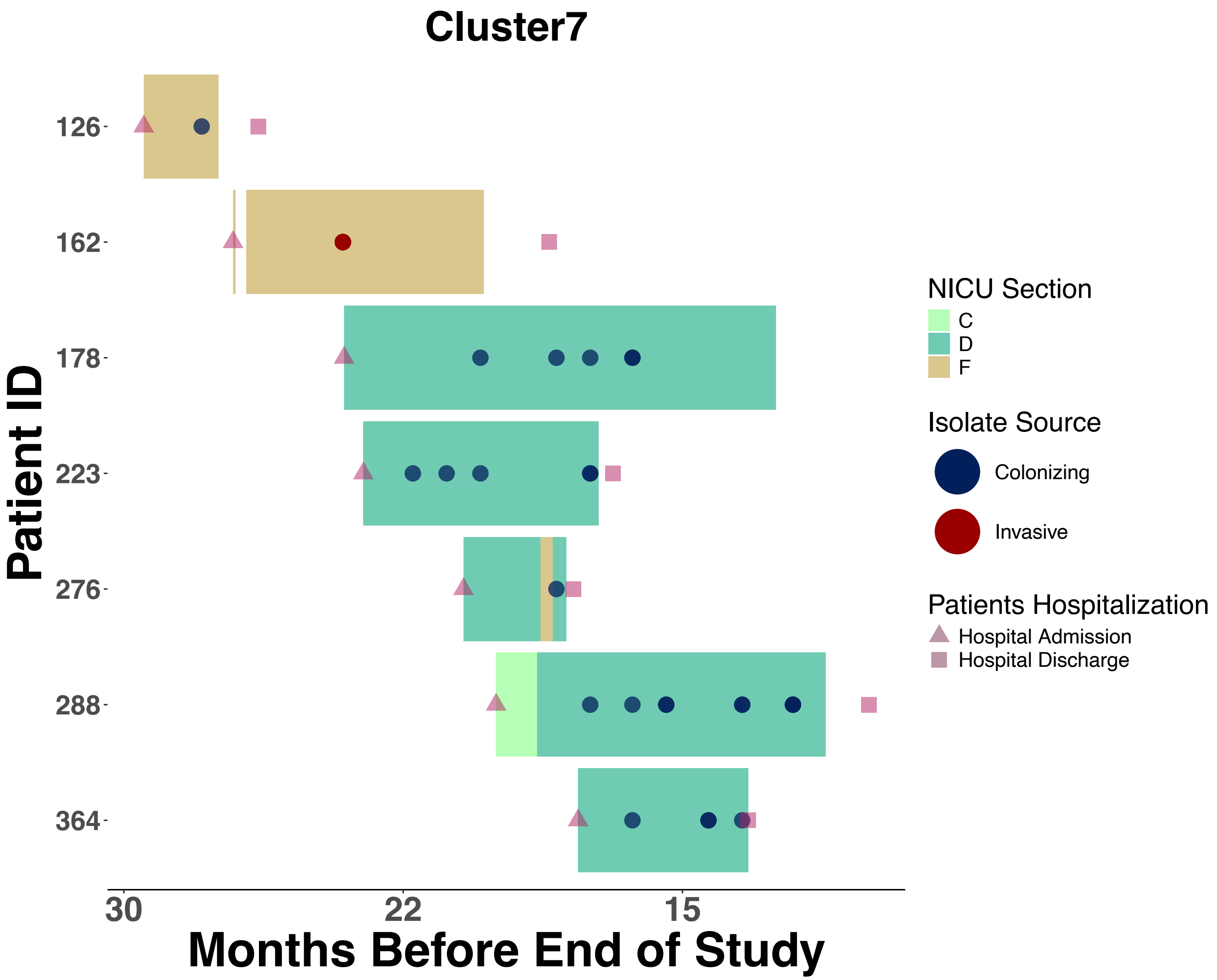
Cluster7
Patient ID
126
162
178
223
276
288
364
30
22
15
Months Before End of Study
NICU Section
C
D
F
Isolate Source
Colonizing
Invasive
Patients Hospitalization
Hospital Admission
Hospital Discharge

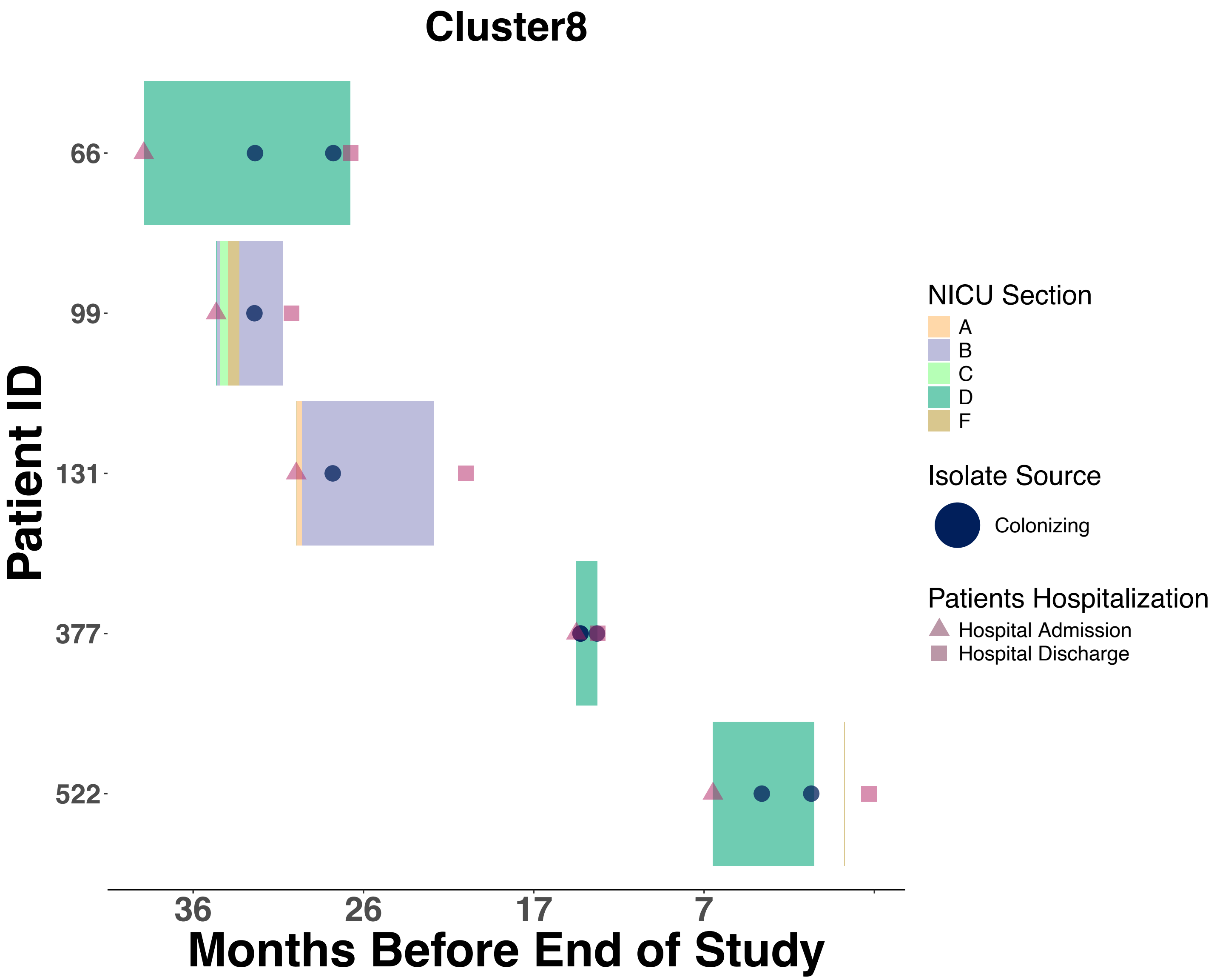

Cluster8
Patient ID
66
99
131
377
522
36
26
17
7
Months Before End of Study
NICU Section
A
B
C
D
F
Isolate Source
Colonizing
Patients Hospitalization
Hospital Admission
Hospital Discharge

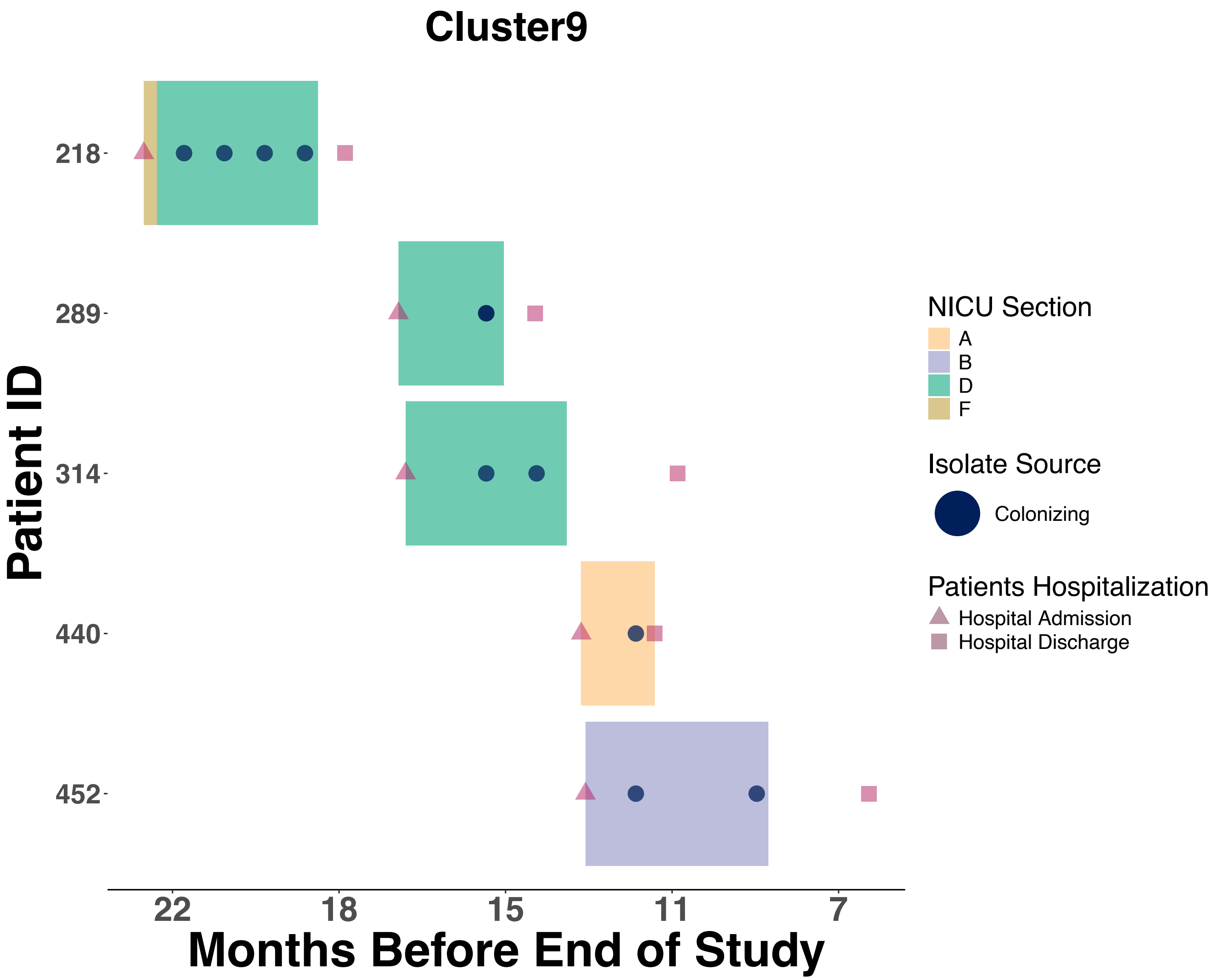
Cluster9
Patient ID
218
289
314
440
452
22
18
15
11
7
Months Before End of Study
NICU Section
A
B
D
F
Isolate Source
Colonizing
Patients Hospitalization
Hospital Admission
Hospital Discharge

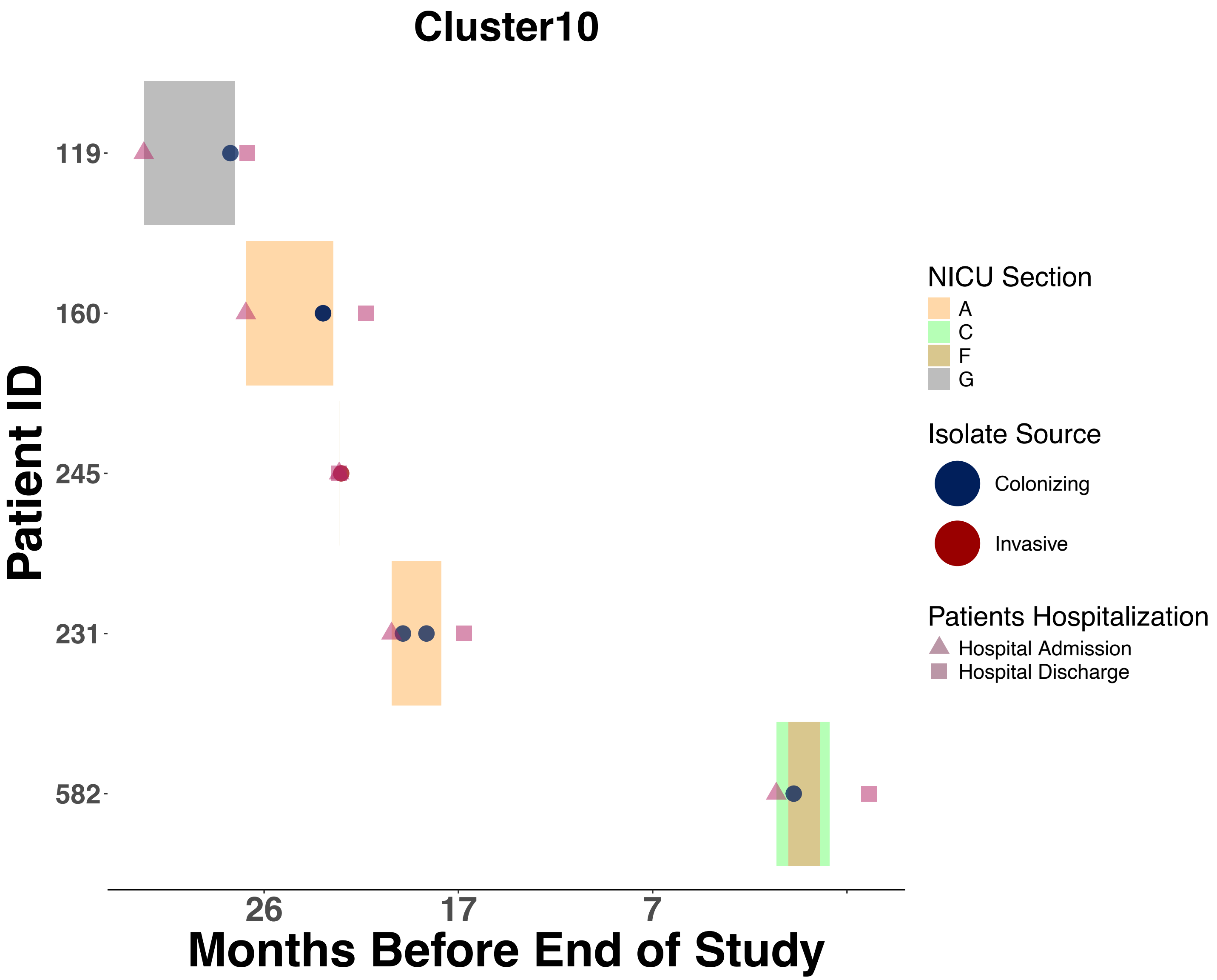
Cluster10
Patient ID
119
160
245
231
582
26
17
7
Months Before End of Study
NICU Section
A
C
F
G
Isolate Source
Colonizing
Invasive
Patients Hospitalization
Hospital Admission
Hospital Discharge

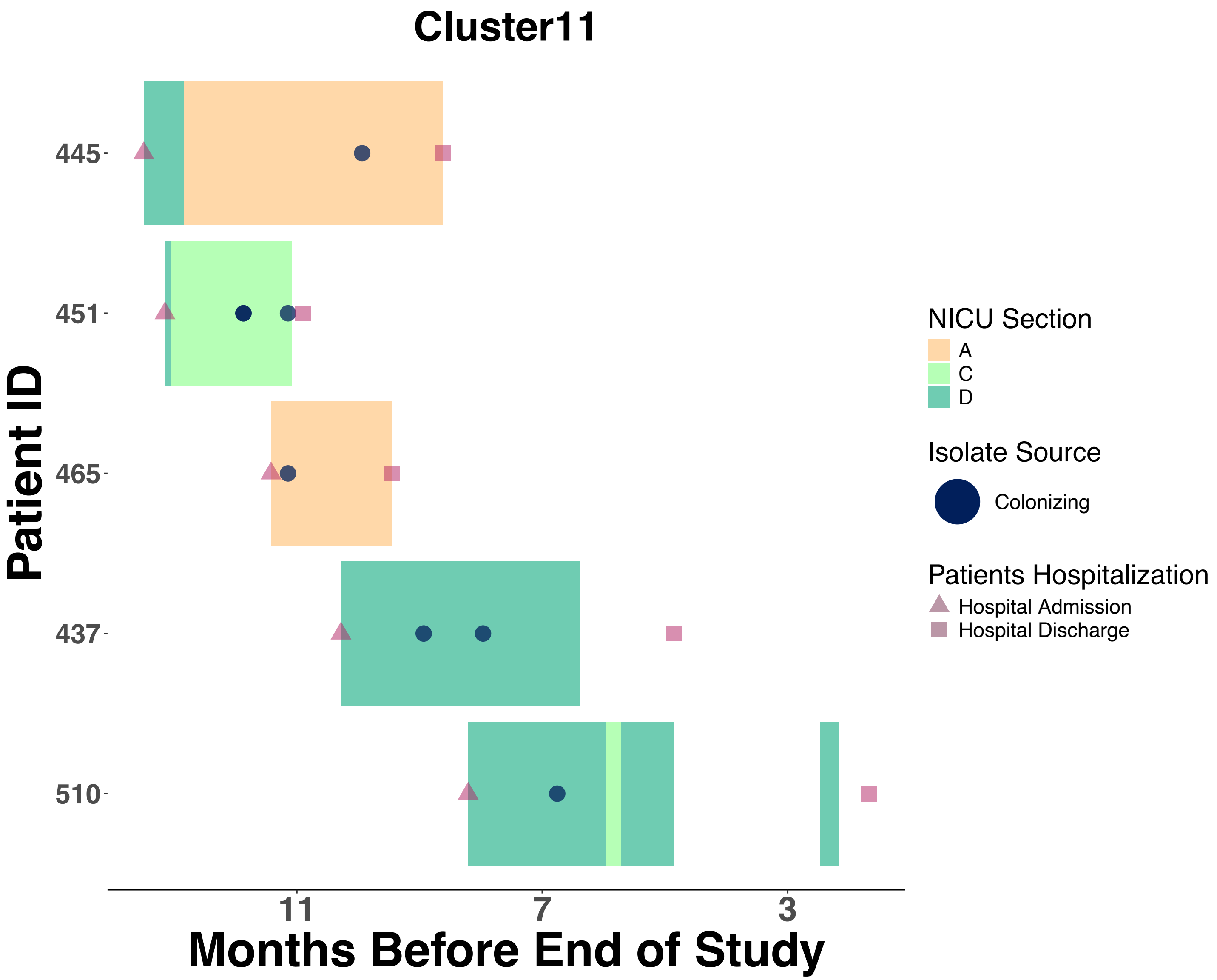
Cluster11
Patient ID
445
451
465
437
510
11
7
3
Months Before End of Study
NICU Section
A
C
D
Isolate Source
Colonizing
Patients Hospitalization
Hospital Admission
Hospital Discharge

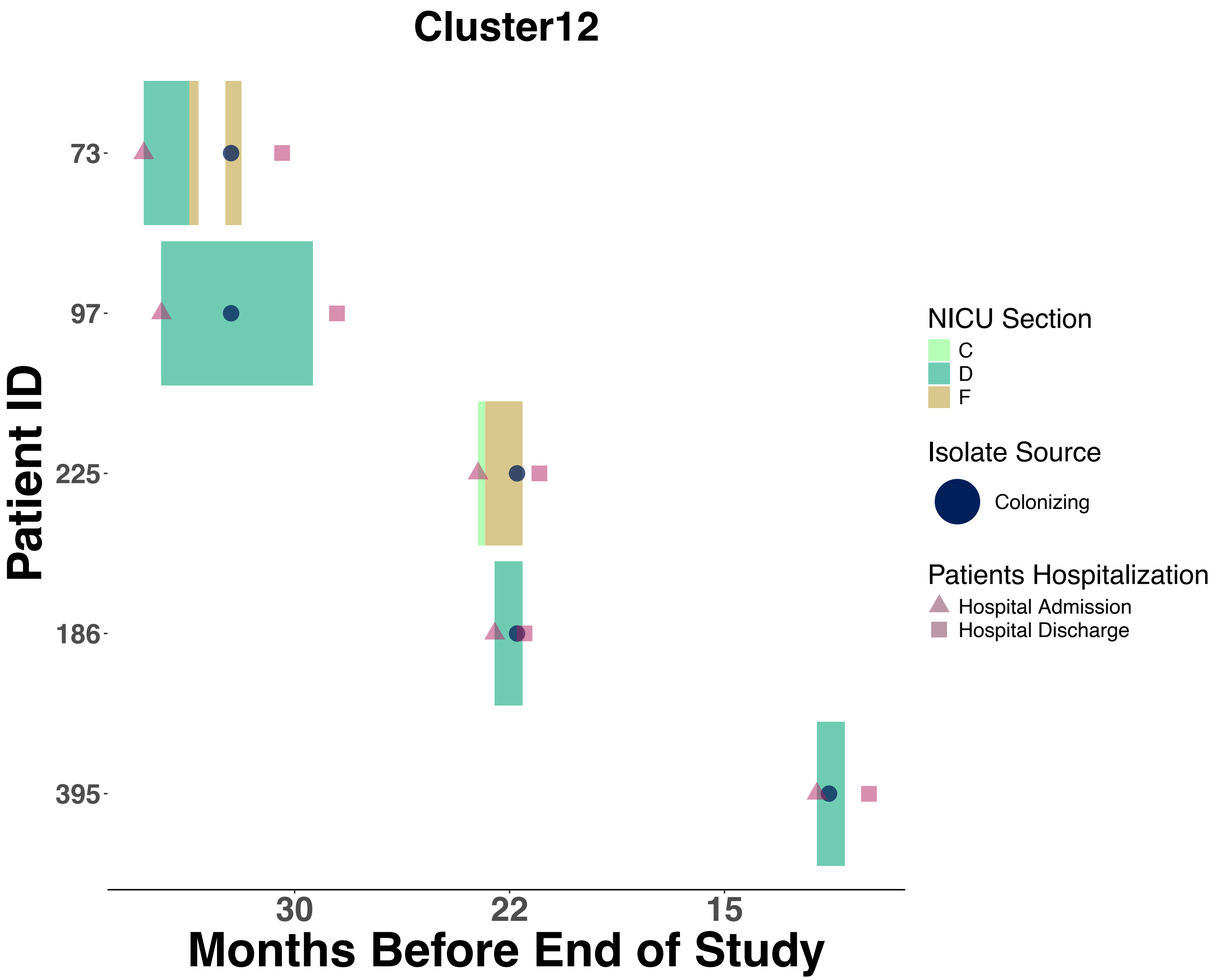
Cluster12
Patient ID
73
97
225
186
395
30
22
15
Months Before End of Study
NICU Section
C
D
F
Isolate Source
Colonizing
Patients Hospitalization
Hospital Admission
Hospital Discharge

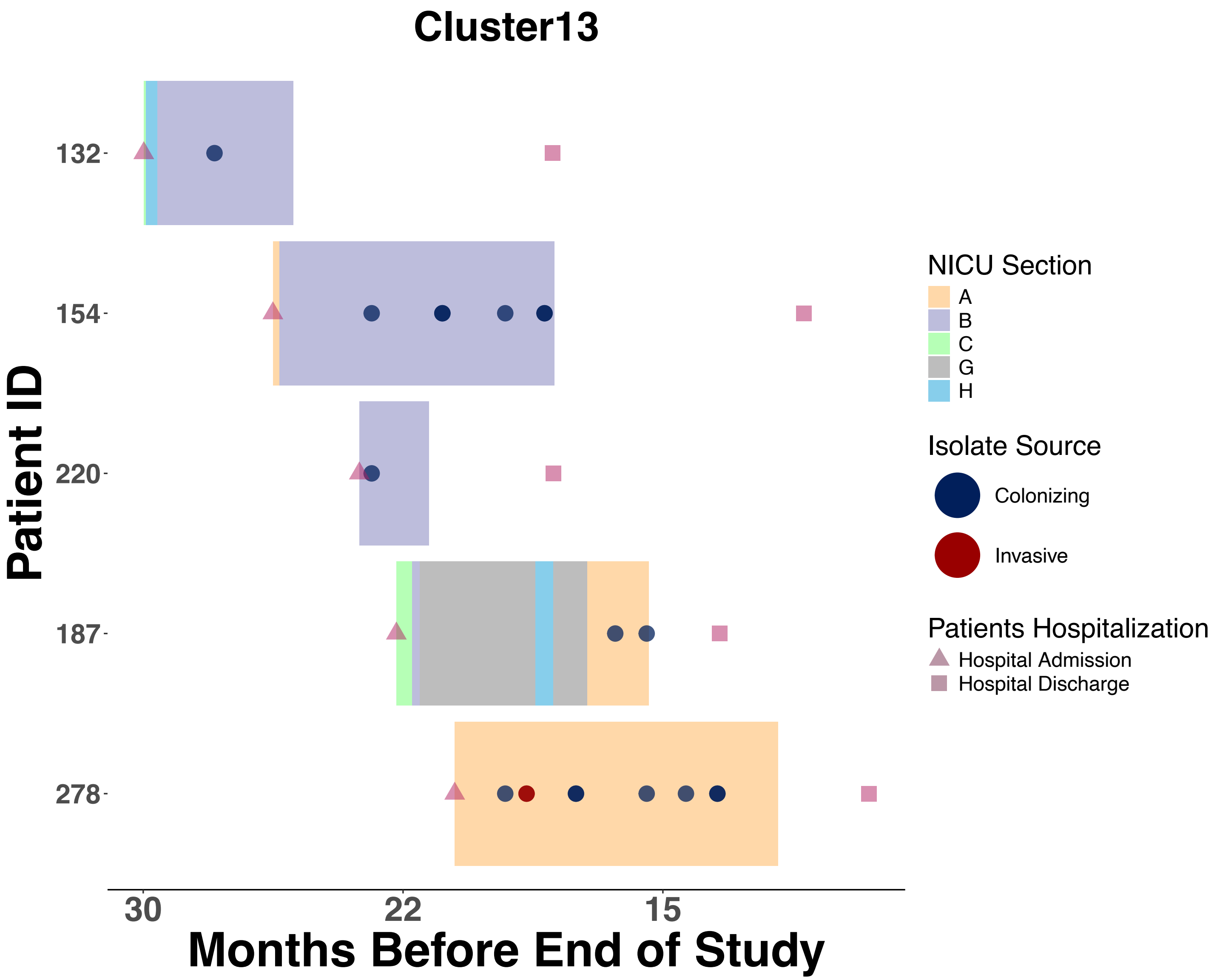

Cluster13
Patient ID
132
154
220
187
278
30
22
15
Months Before End of Study
NICU Section
A
B
C
G
H
Isolate Source
Colonizing
Invasive
Patients Hospitalization
Hospital Admission
Hospital Discharge

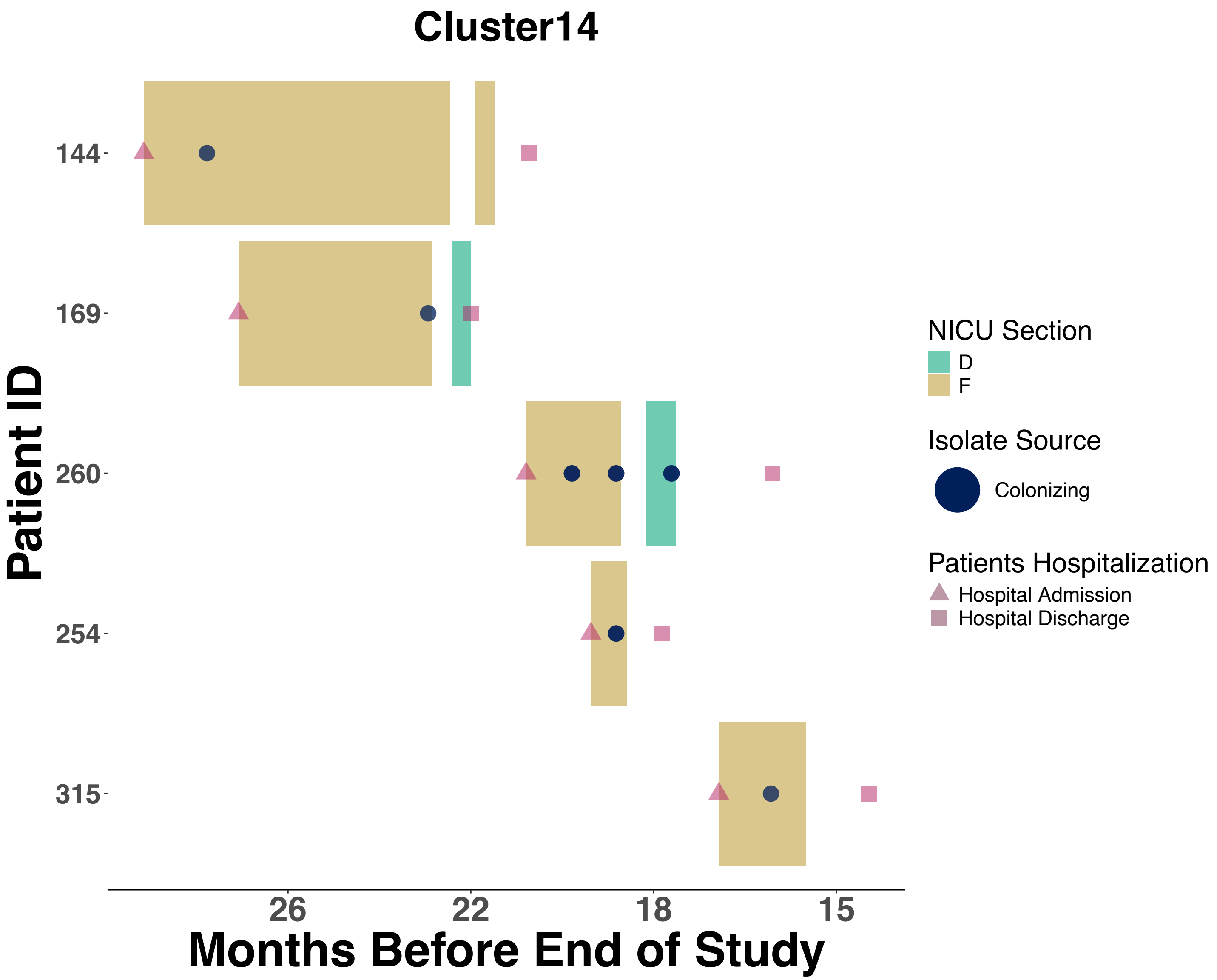

Cluster14
Patient ID
144
169
260
254
315
26
22
18
15
Months Before End of Study
NICU Section
D
F
Isolate Source
Colonizing
Patients Hospitalization
Hospital Admission
Hospital Discharge

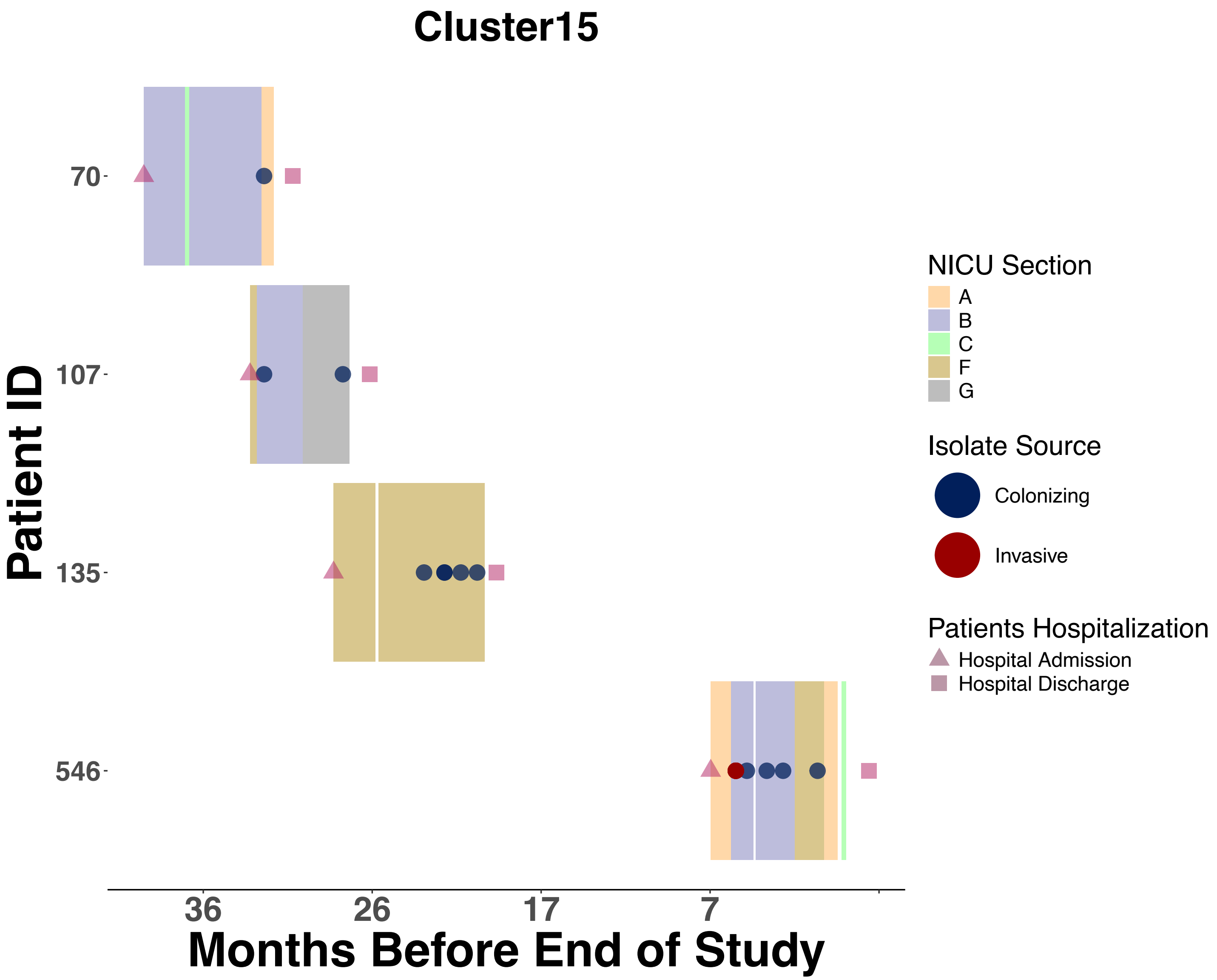

Cluster15
Patient ID
70
107
135
546
36
26
17
7
Months Before End of Study
NICU Section
A
B
C
F
G
Isolate Source
Colonizing
Invasive
Patients Hospitalization
Hospital Admission
Hospital Discharge

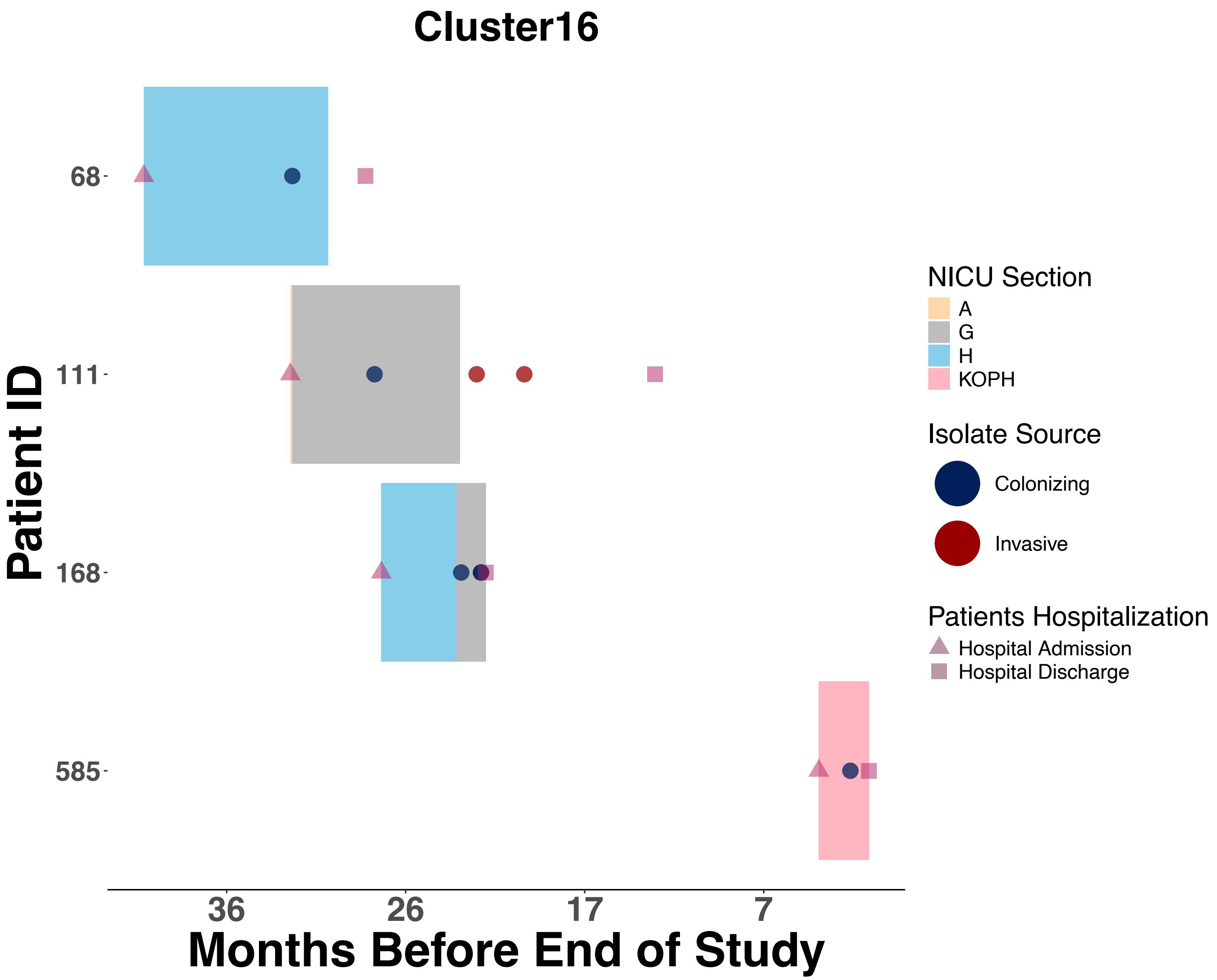

Cluster16
68
111
168
585
Patient ID
36
26
17
7
Months Before End of Study
NICU Section
A
G
H
KOPH
Isolate Source
Colonizing
Invasive
Patients Hospitalization
Hospital Admission
Hospital Discharge

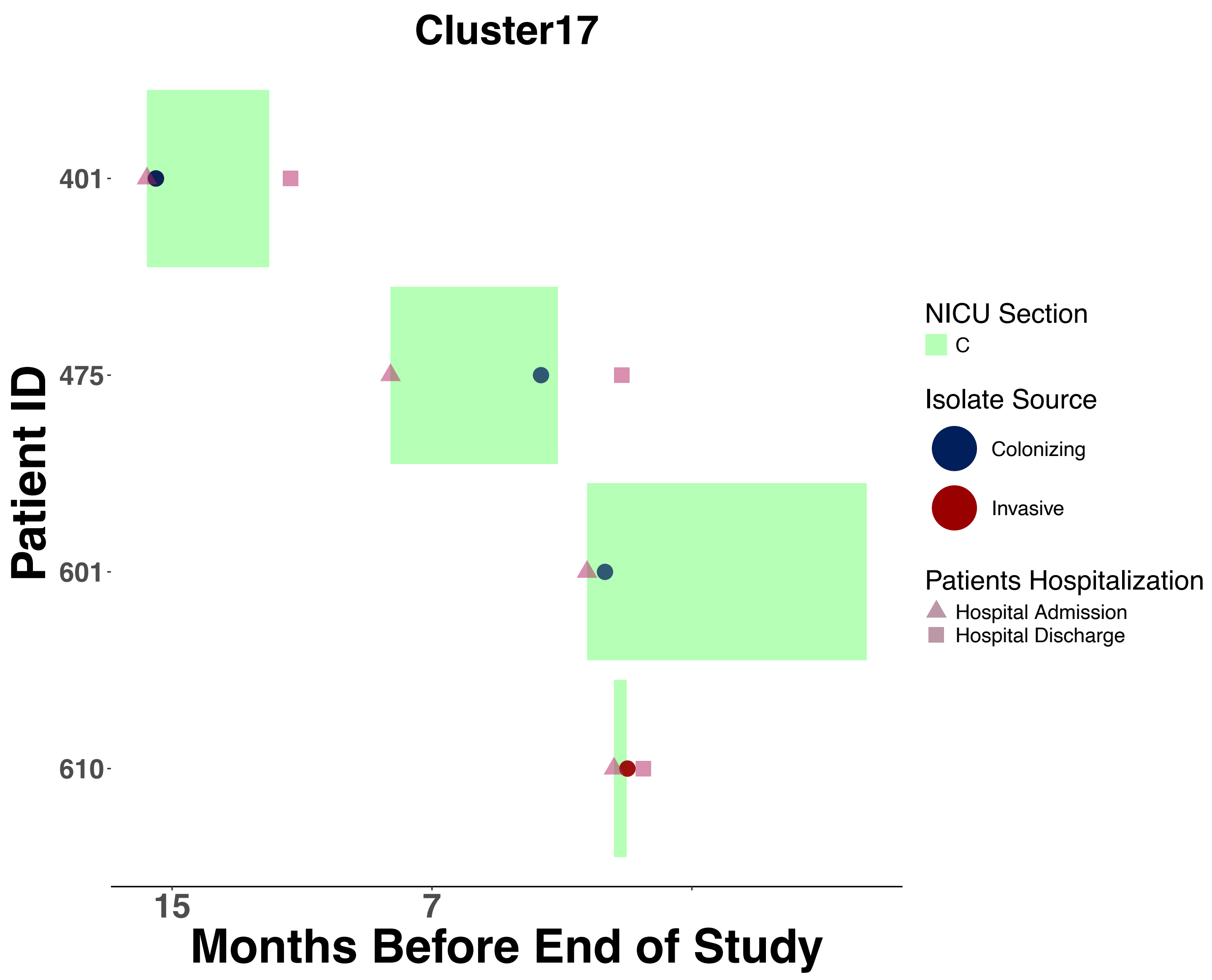

Cluster17
Patient ID
401
475
601
610
15
7
Months Before End of Study
NICU Section
C
Isolate Source
Colonizing
Invasive
Patients Hospitalization
Hospital Admission
Hospital Discharge

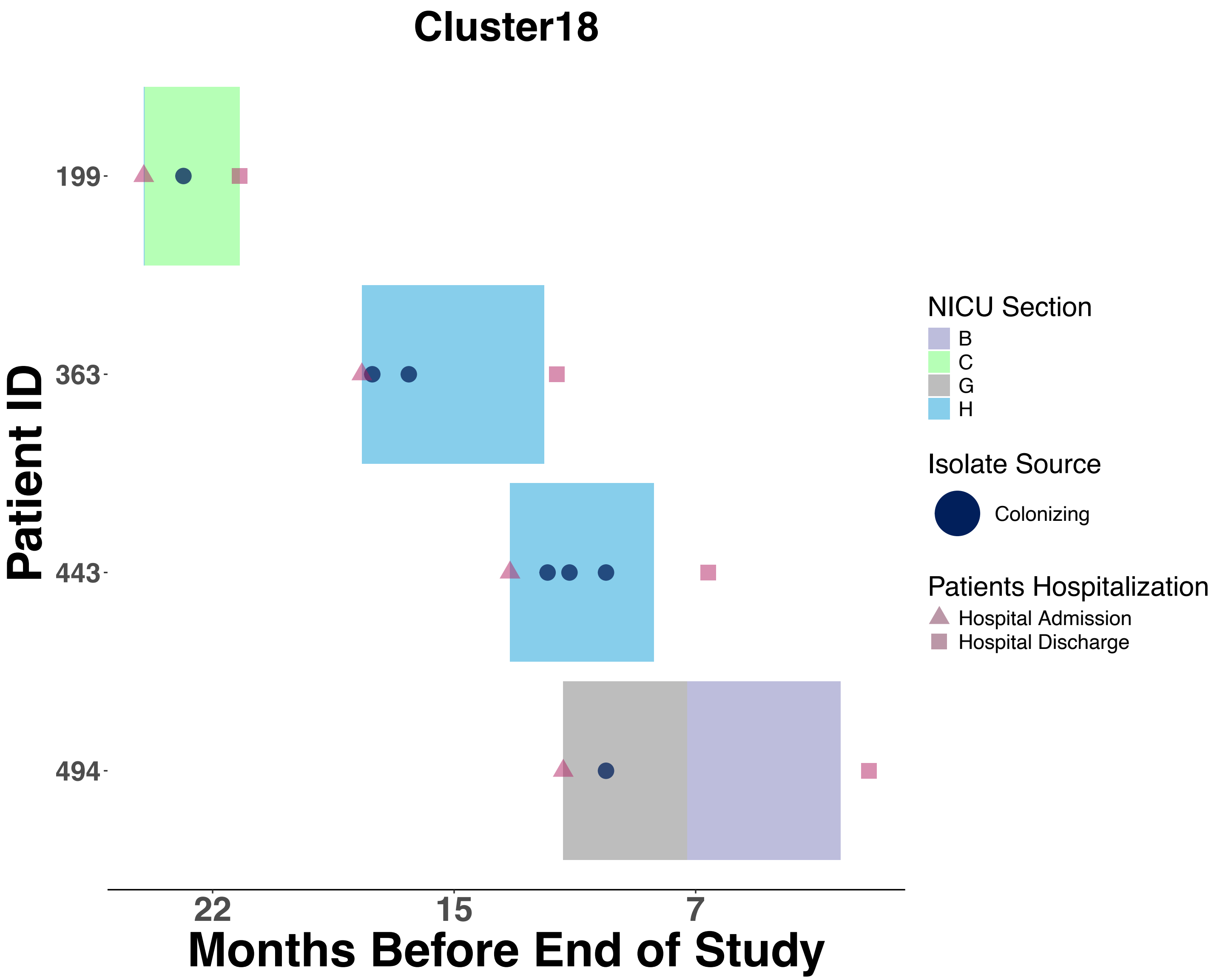

Cluster18
Patient ID
199
363
443
494
22
15
7
Months Before End of Study
NICU Section
B
C
G
H
Isolate Source
Colonizing
Patients Hospitalization
Hospital Admission
Hospital Discharge

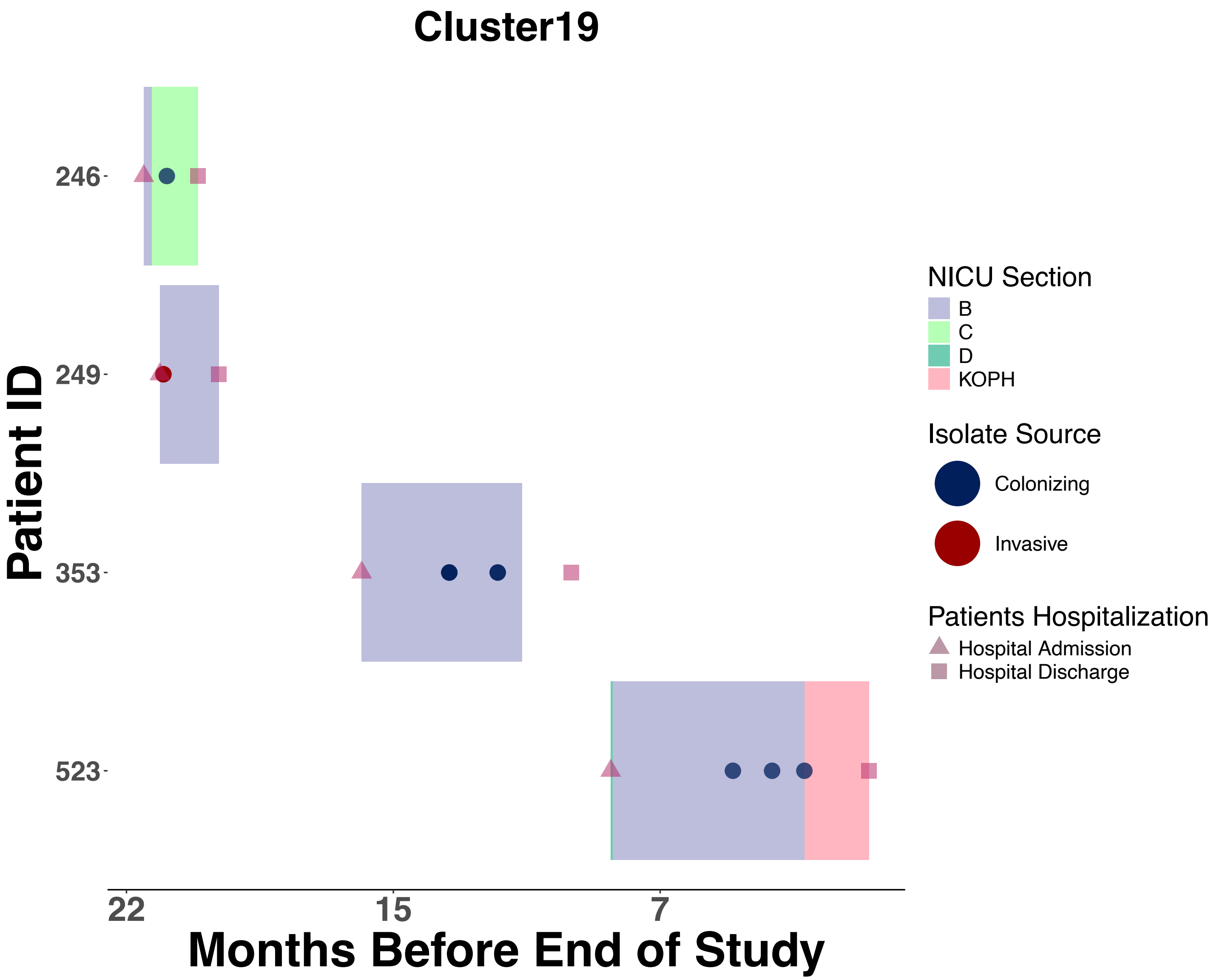
Cluster19
Patient ID
246
249
353
523
22
15
7
Months Before End of Study
NICU Section
B
C
D
KOPH
Isolate Source
Colonizing
Invasive
Patients Hospitalization
Hospital Admission
Hospital Discharge

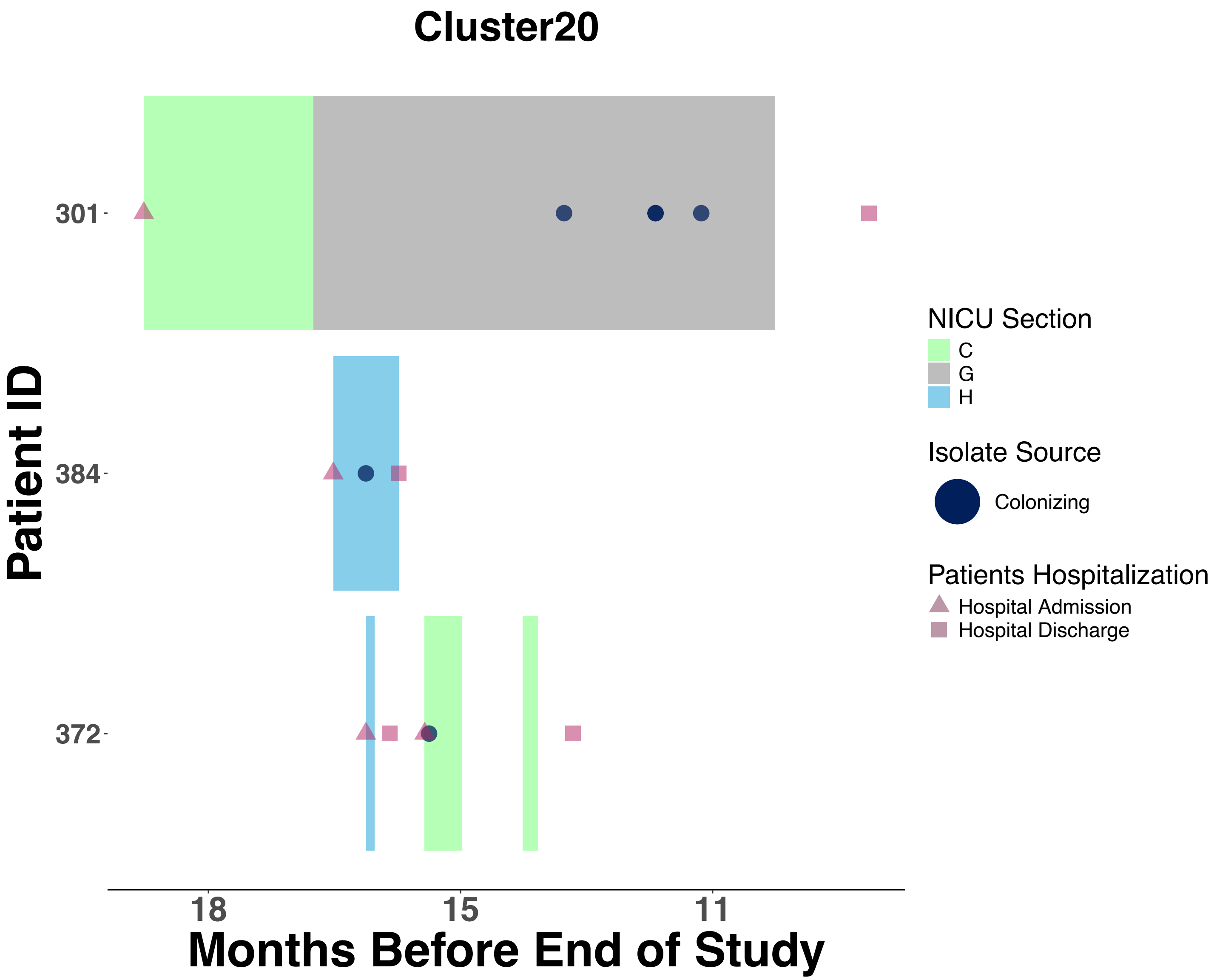
Cluster20
Patient ID
301
384
372
18
15
11
Months Before End of Study
NICU Section
C
G
H
Isolate Source
Colonizing
Patients Hospitalization
Hospital Admission
Hospital Discharge

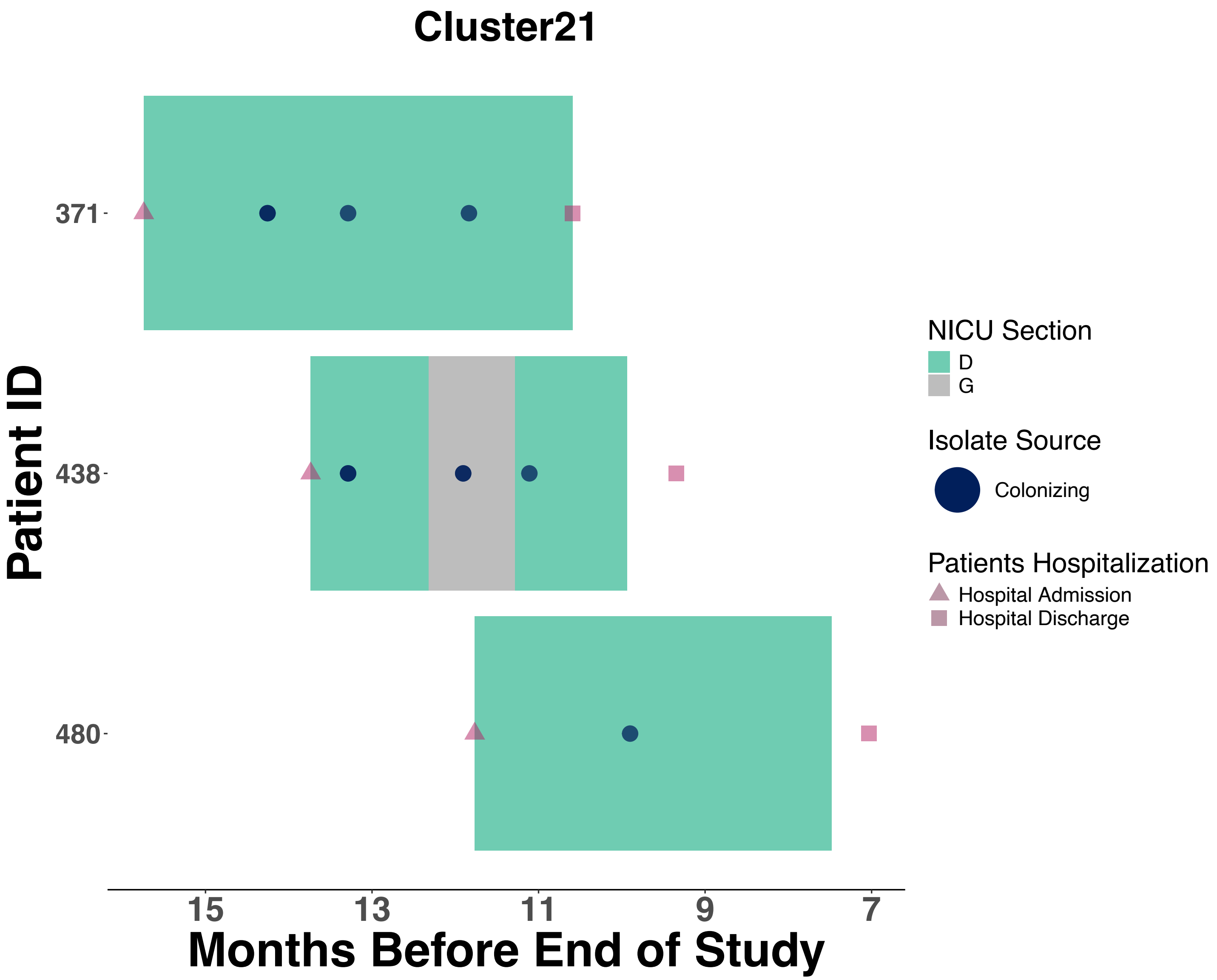
Cluster21
Patient ID
371
438
480
15
13
11
9
7
Months Before End of Study
NICU Section
D
G
Isolate Source
Colonizing
Patients Hospitalization
Hospital Admission
Hospital Discharge

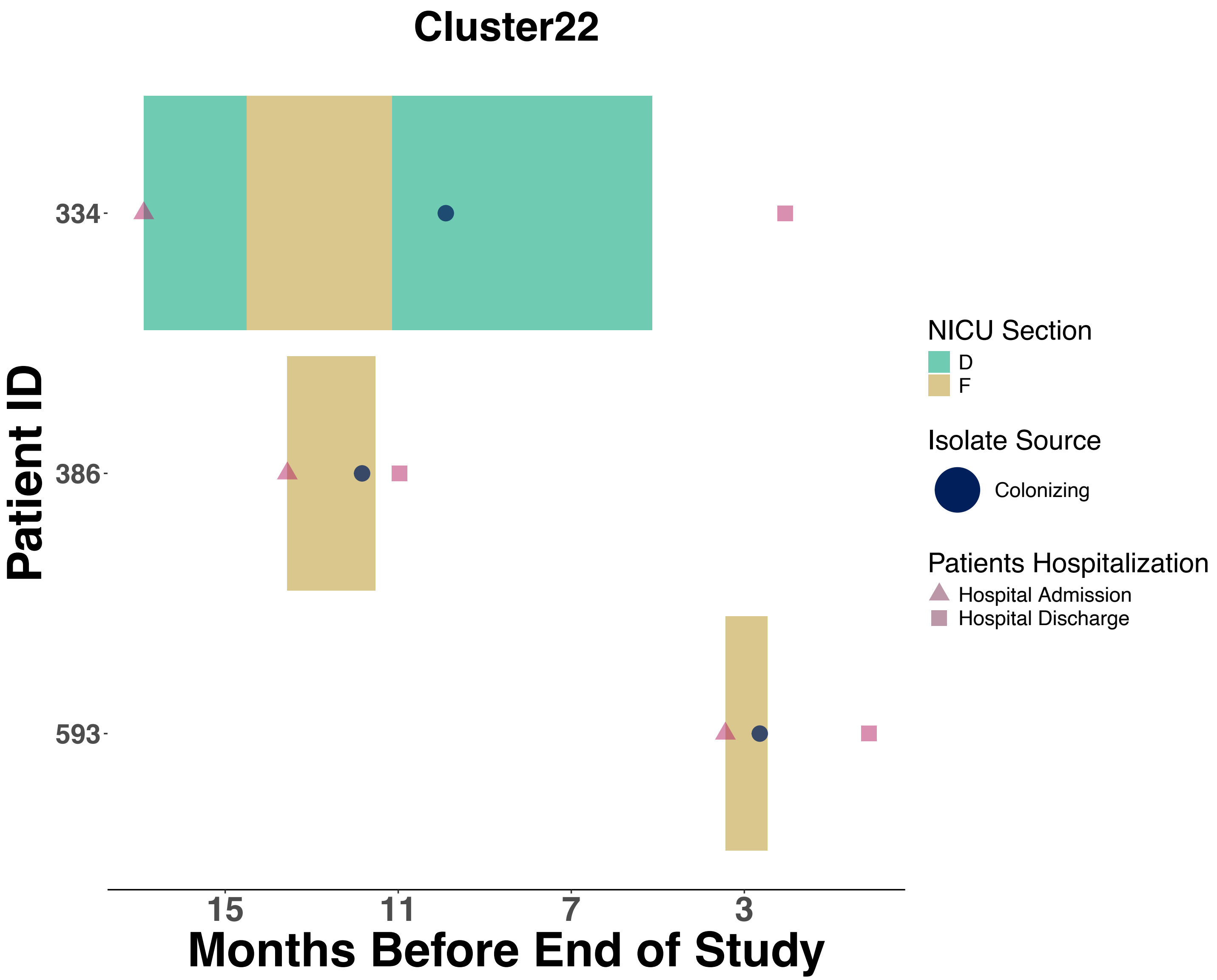
Cluster22
Patient ID
334
386
593
15
11
7
3
Months Before End of Study
NICU Section
D
F
Isolate Source
Colonizing
Patients Hospitalization
Hospital Admission
Hospital Discharge

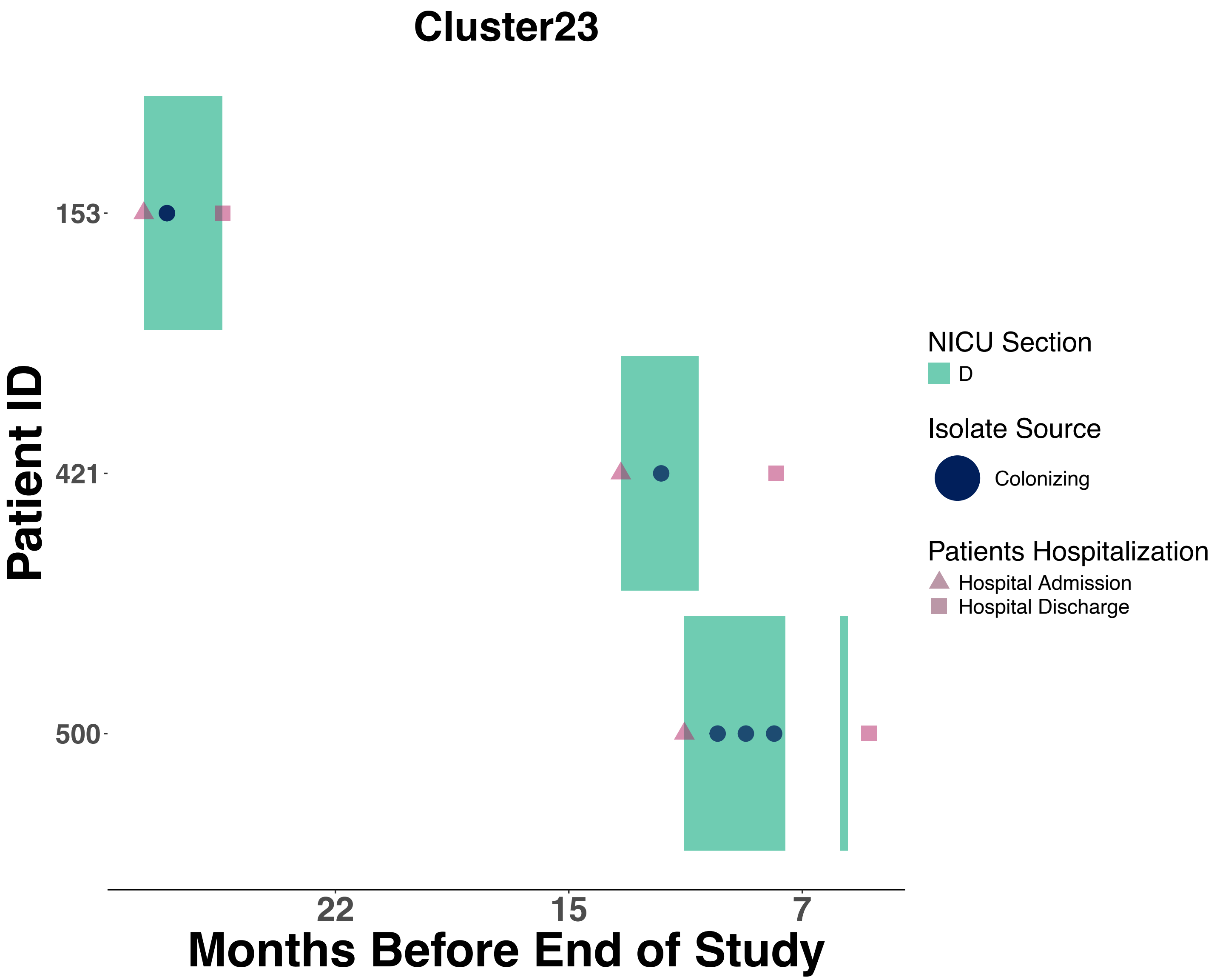
Cluster23
Patient ID
153
421
500
22
15
7
Months Before End of Study
NICU Section
D
Isolate Source
Colonizing
Patients Hospitalization
Hospital Admission
Hospital Discharge

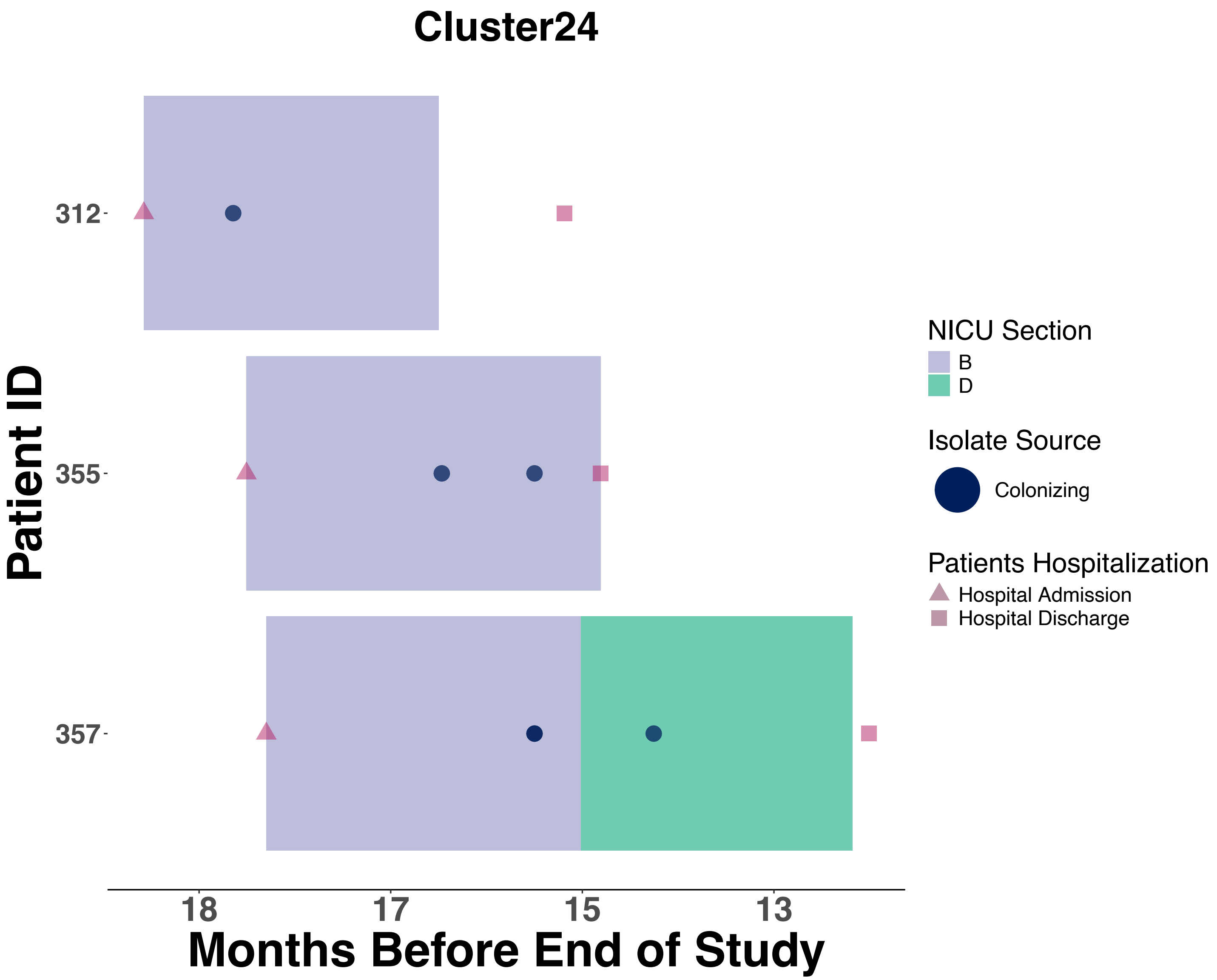
Cluster24
Patient ID
312
355
357
18
17
15
13
Months Before End of Study
NICU Section
B
D
Isolate Source
Colonizing
Patients Hospitalization
Hospital Admission
Hospital Discharge

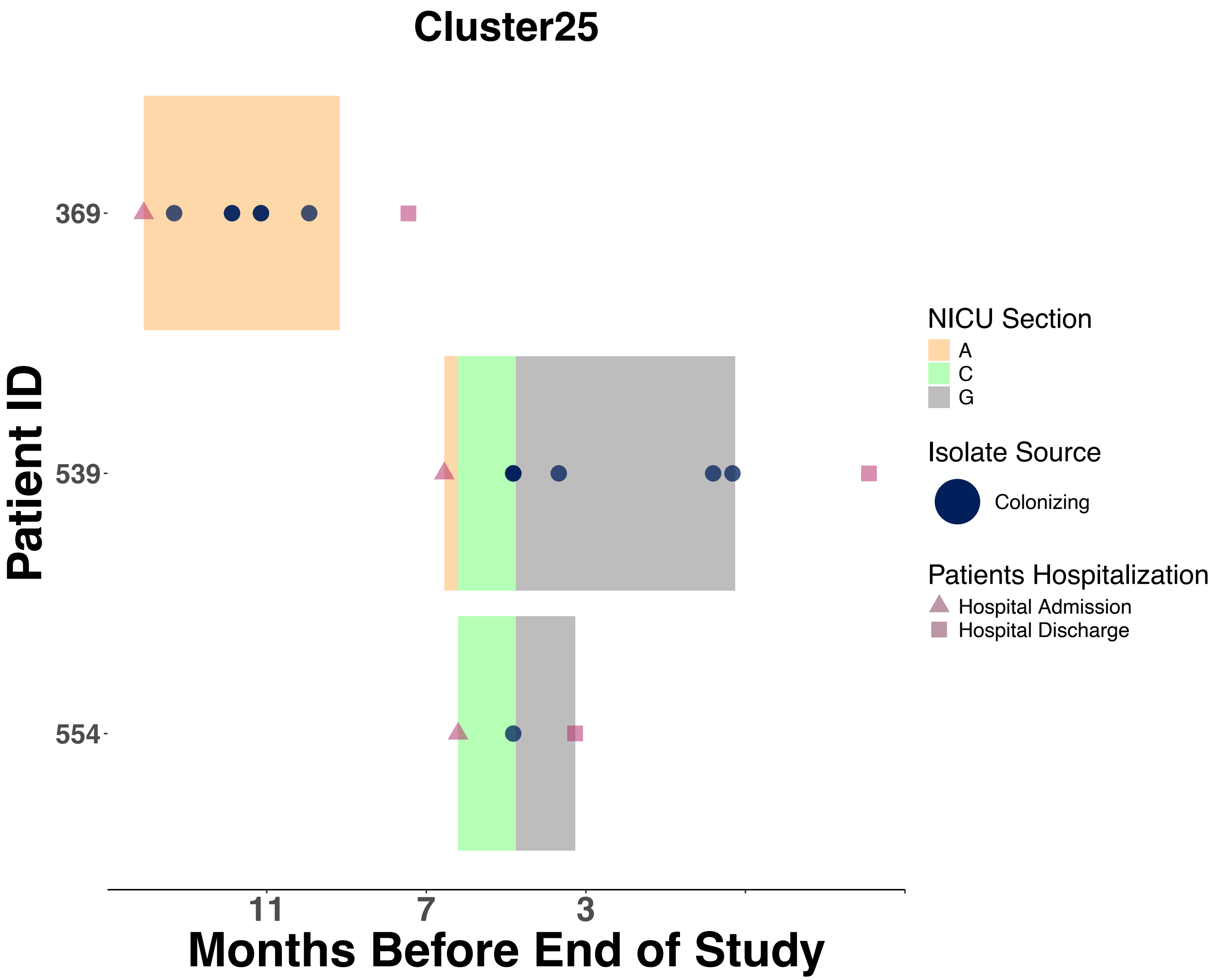
Cluster25
Patient ID
369
539
554
11
7
3
Months Before End of Study
NICU Section
A
C
G
Isolate Source
Colonizing
Patients Hospitalization
Hospital Admission
Hospital Discharge

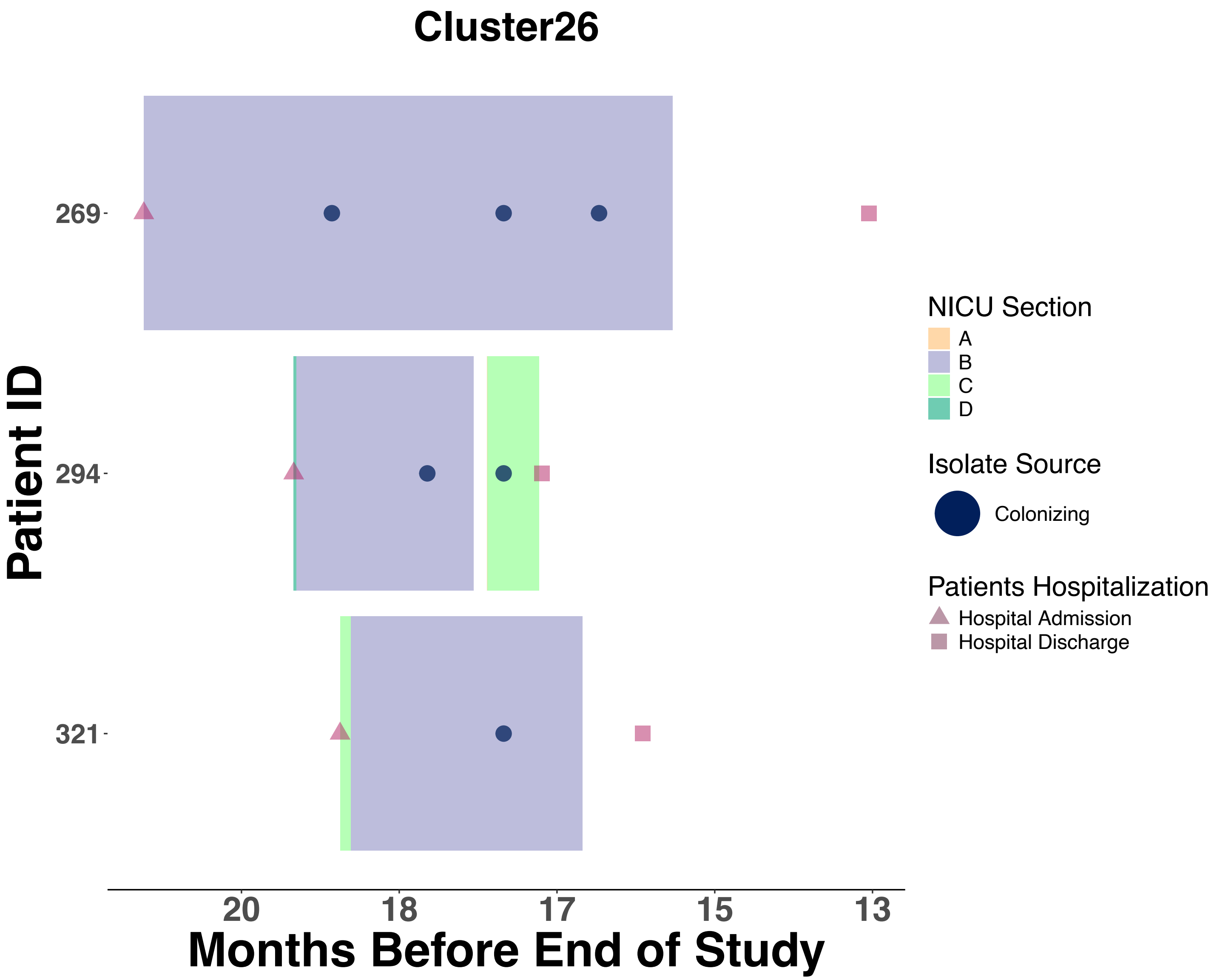

Cluster26
Patient ID
269
294
321
20
18
17
15
13
Months Before End of Study
NICU Section
A
B
C
D
Isolate Source
Colonizing
Patients Hospitalization
Hospital Admission
Hospital Discharge

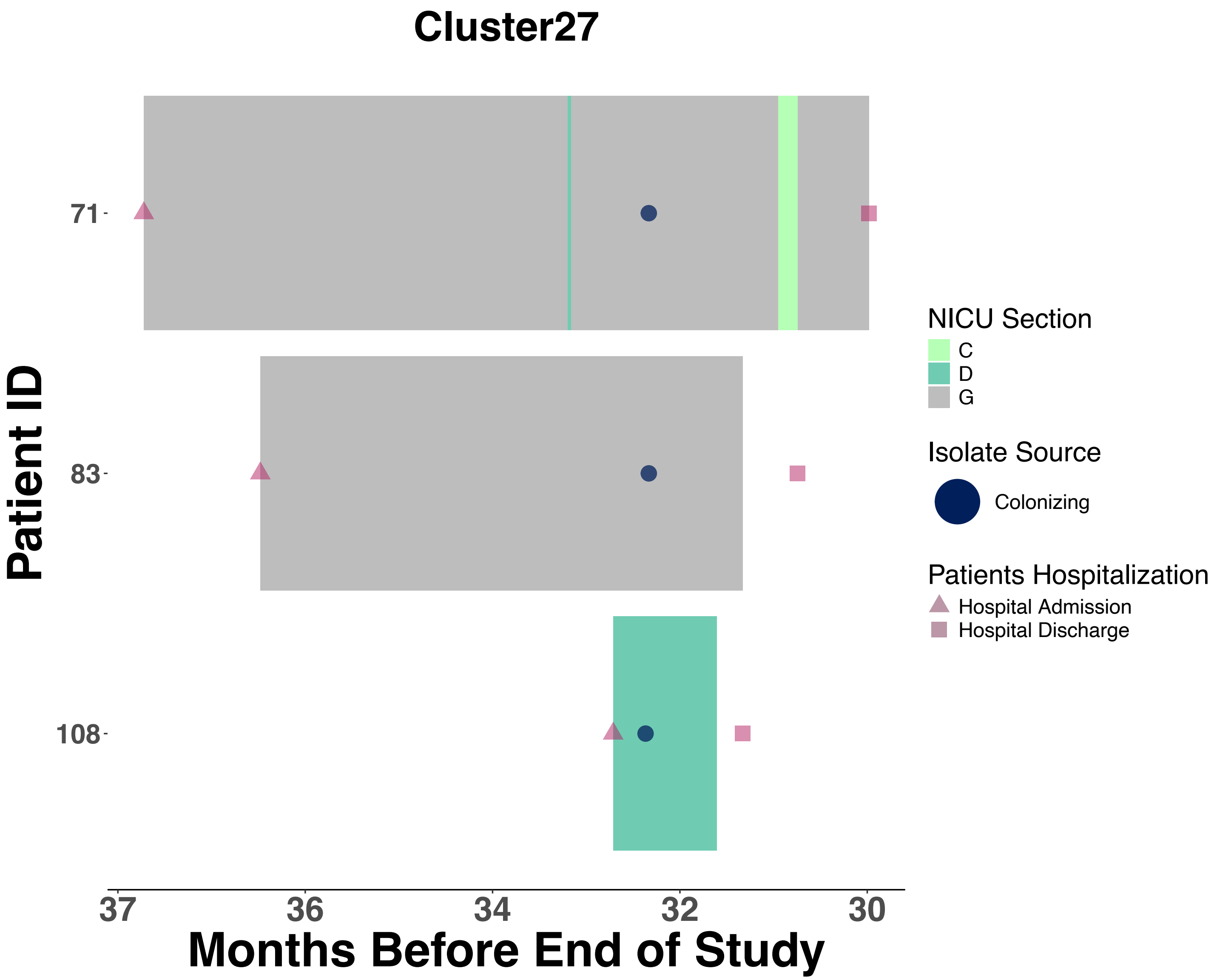
Cluster27
Patient ID
71
83
108
37
36
34
32
30
Months Before End of Study
NICU Section
C
D
G
Isolate Source
Colonizing
Patients Hospitalization
Hospital Admission
Hospital Discharge

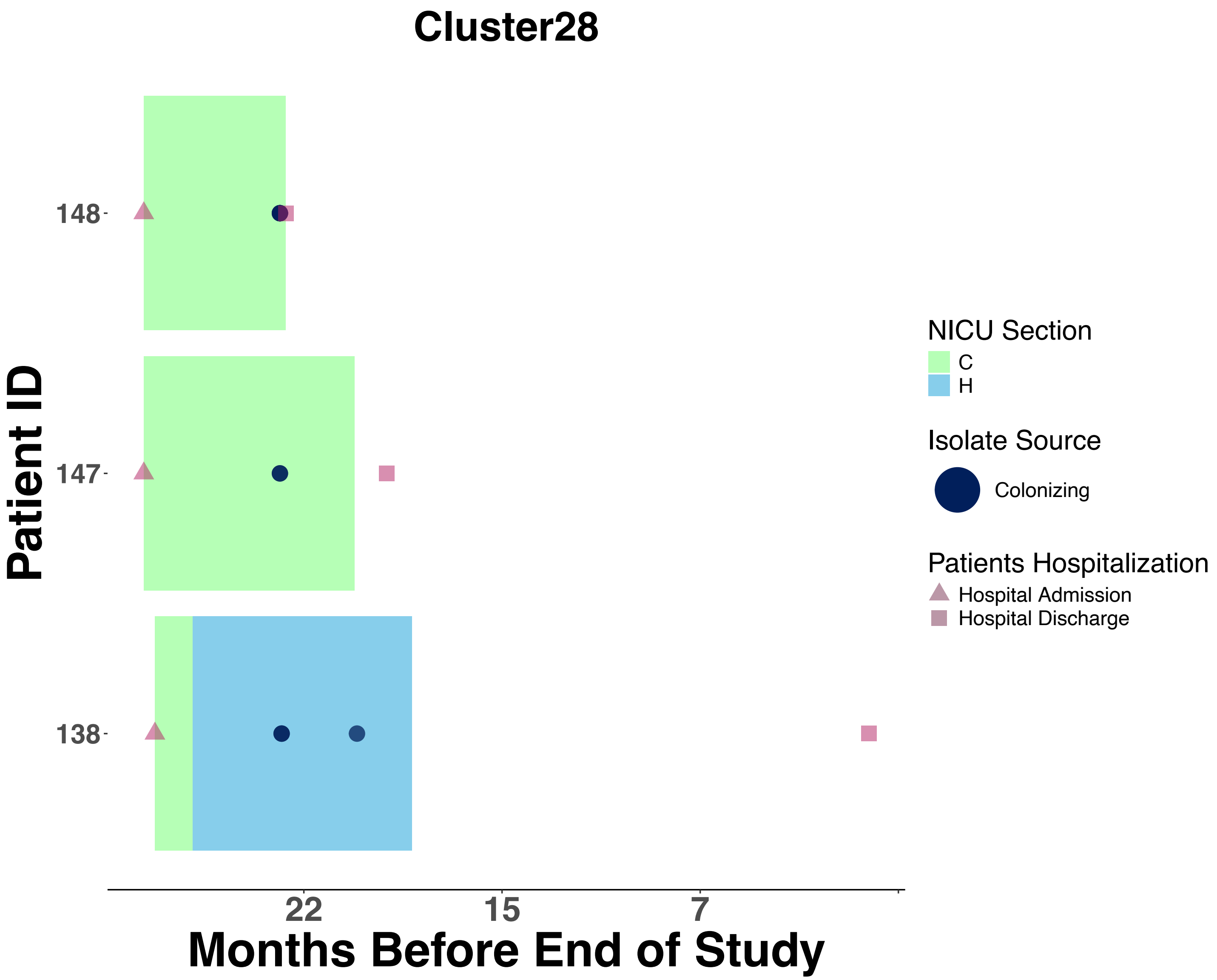

Cluster28
Patient ID
148
147
138
22
15
7
Months Before End of Study
NICU Section
C
H
Isolate Source
Colonizing
Patients Hospitalization
Hospital Admission
Hospital Discharge

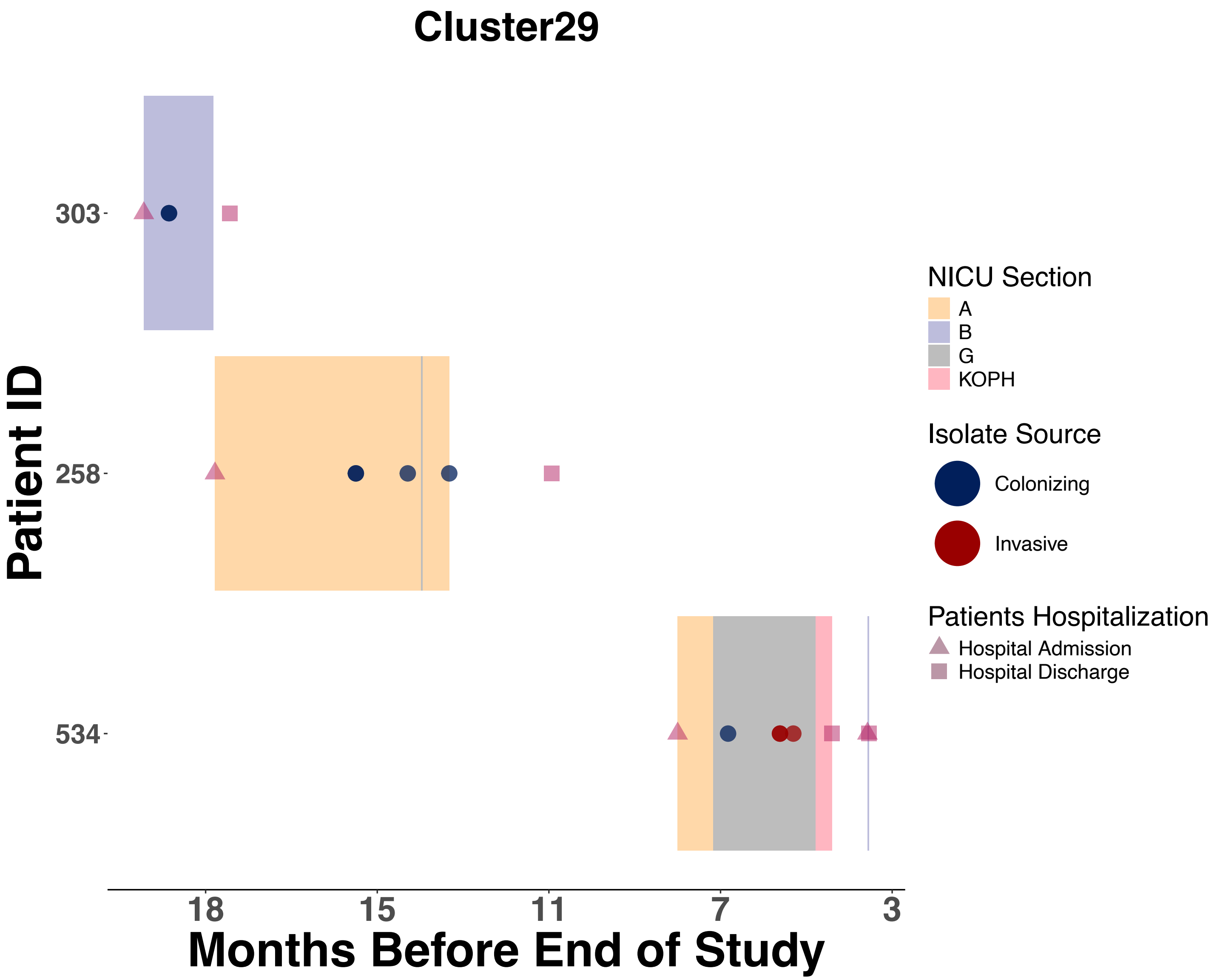

Cluster29
Patient ID
303
258
534
18
15
11
7
3
Months Before End of Study
NICU Section
A
B
G
KOPH
Isolate Source
Colonizing
Invasive
Patients Hospitalization
Hospital Admission
Hospital Discharge

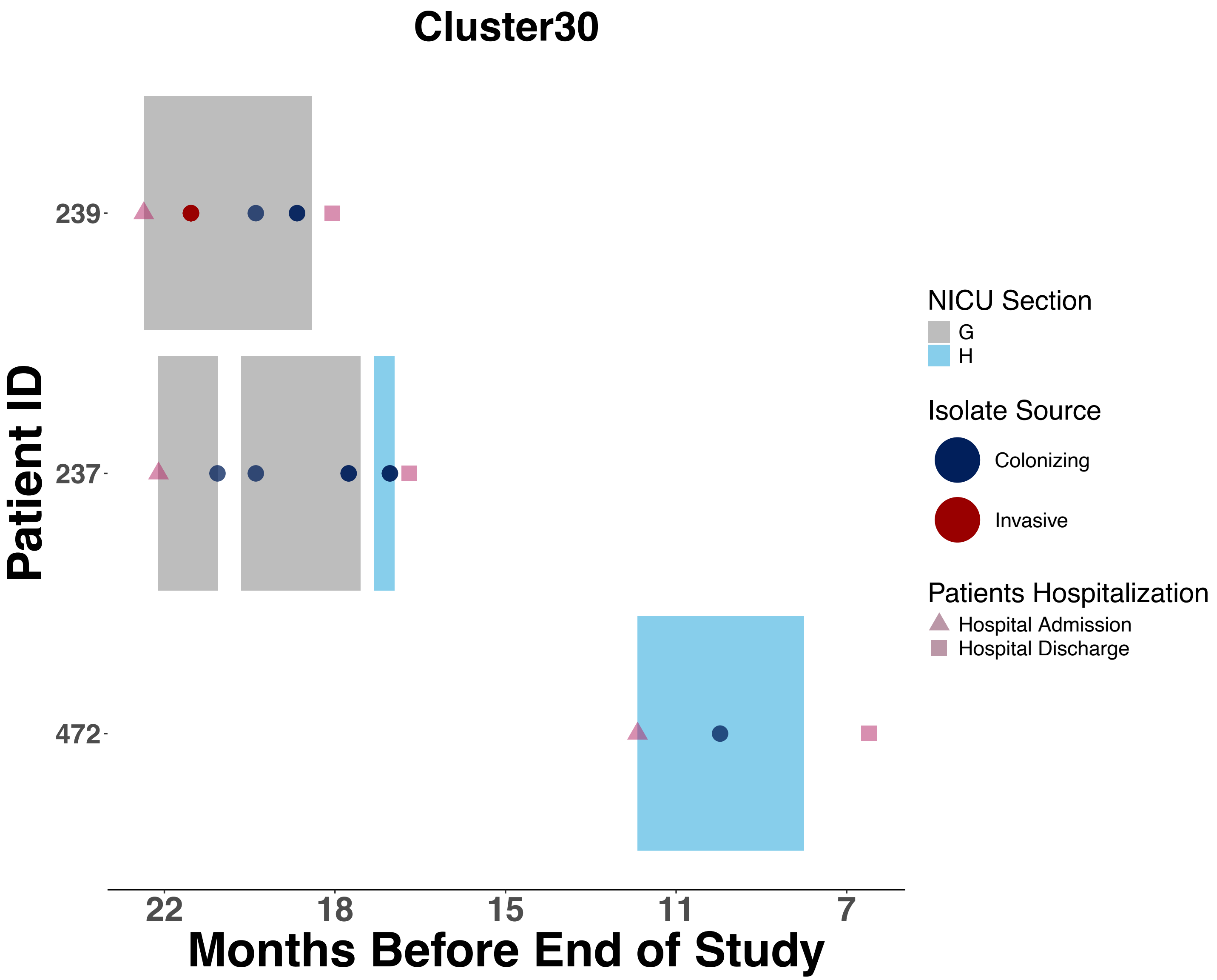

Cluster30
Patient ID
239
237
472
22
18
15
11
7
Months Before End of Study
NICU Section
G
H
Isolate Source
Colonizing
Invasive
Patients Hospitalization
Hospital Admission
Hospital Discharge

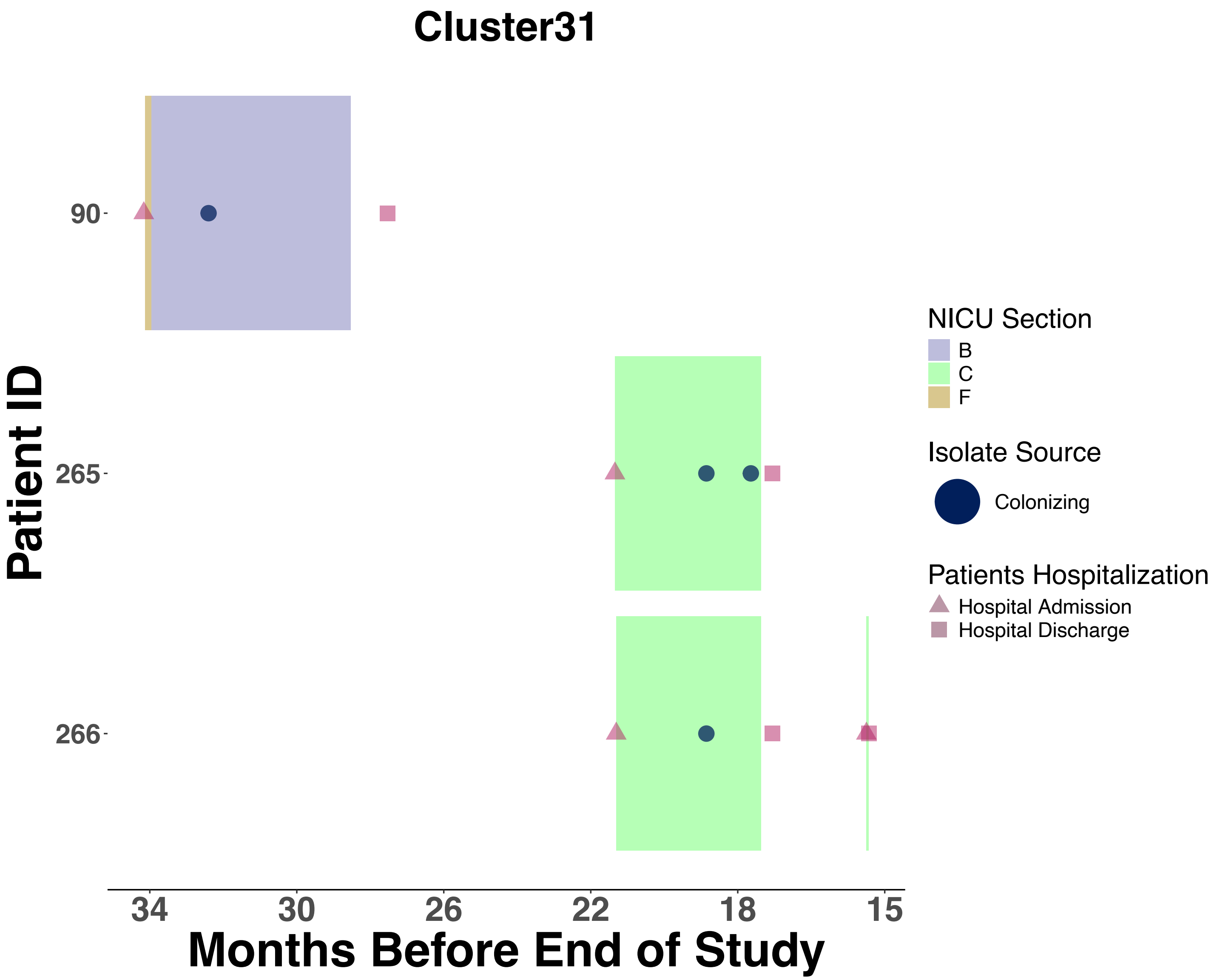
Cluster31
Patient ID
90
265
266
34
30
26
22
18
15
Months Before End of Study
NICU Section
B
C
F
Isolate Source
Colonizing
Patients Hospitalization
Hospital Admission
Hospital Discharge

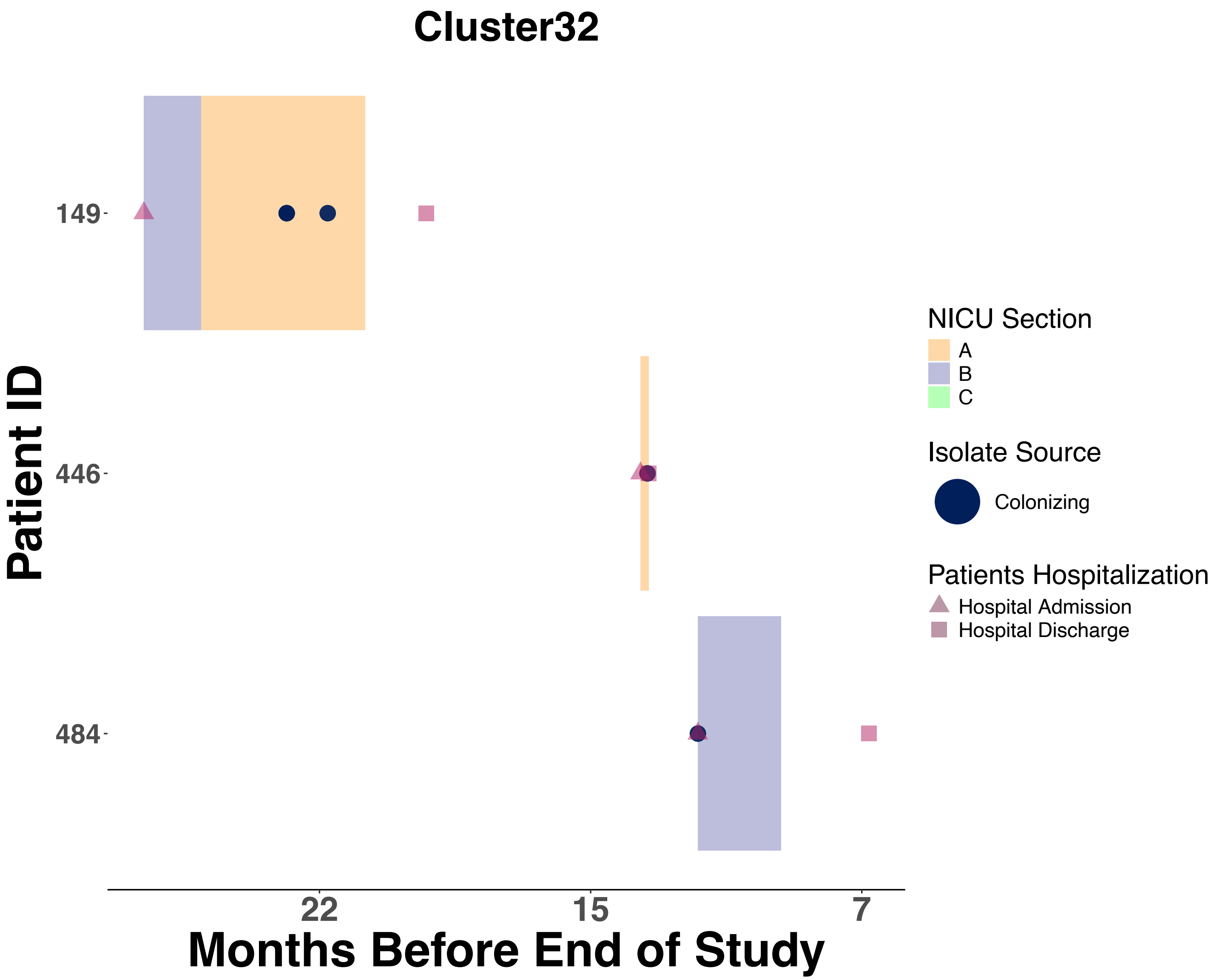
Cluster32
Patient ID
149
446
484
22
15
7
Months Before End of Study
NICU Section
A
B
C
Isolate Source
Colonizing
Patients Hospitalization
Hospital Admission
Hospital Discharge

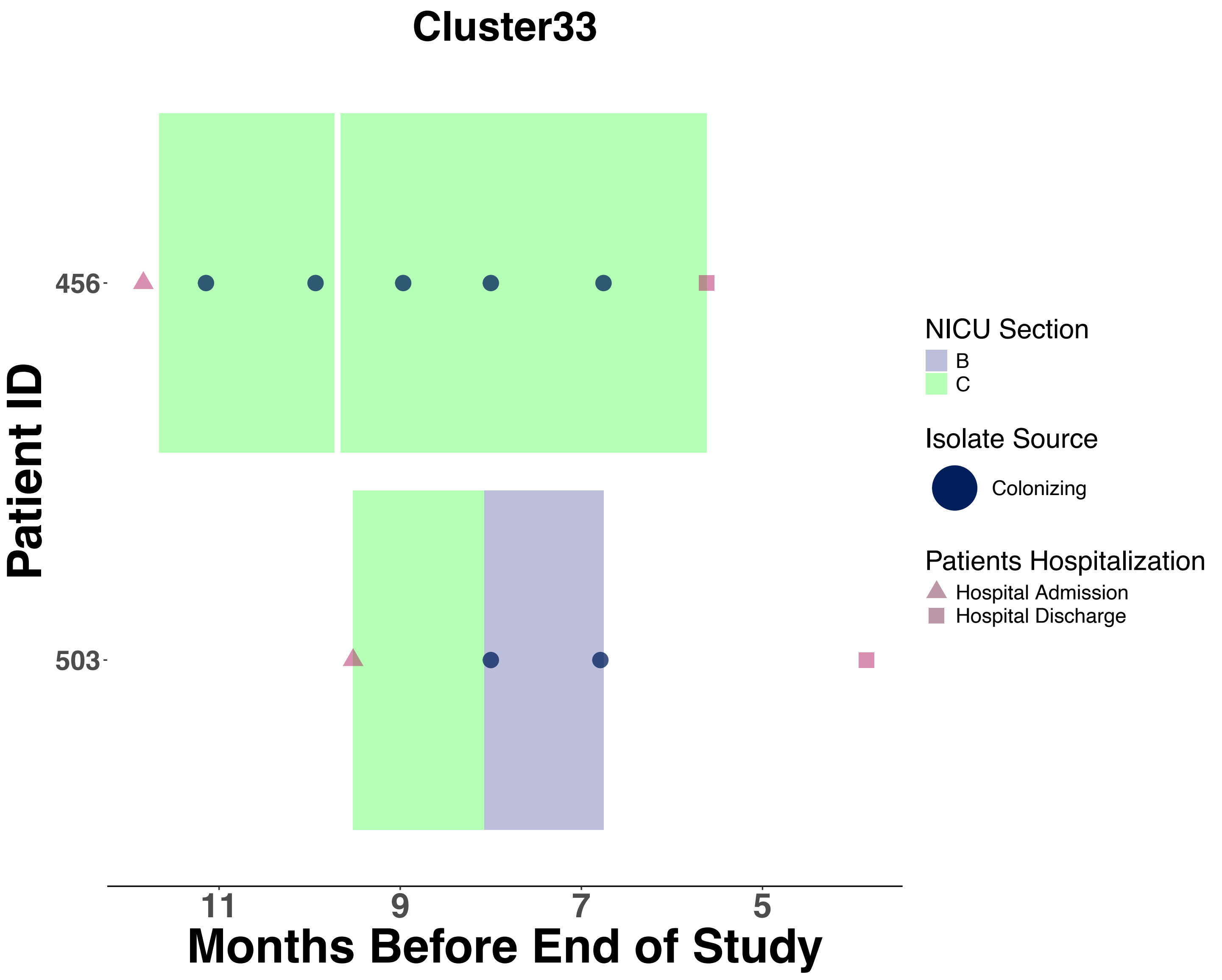
Cluster33
Patient ID
456
503
11
9
7
5
Months Before End of Study
NICU Section
B
C
Isolate Source
Colonizing
Patients Hospitalization
Hospital Admission
Hospital Discharge

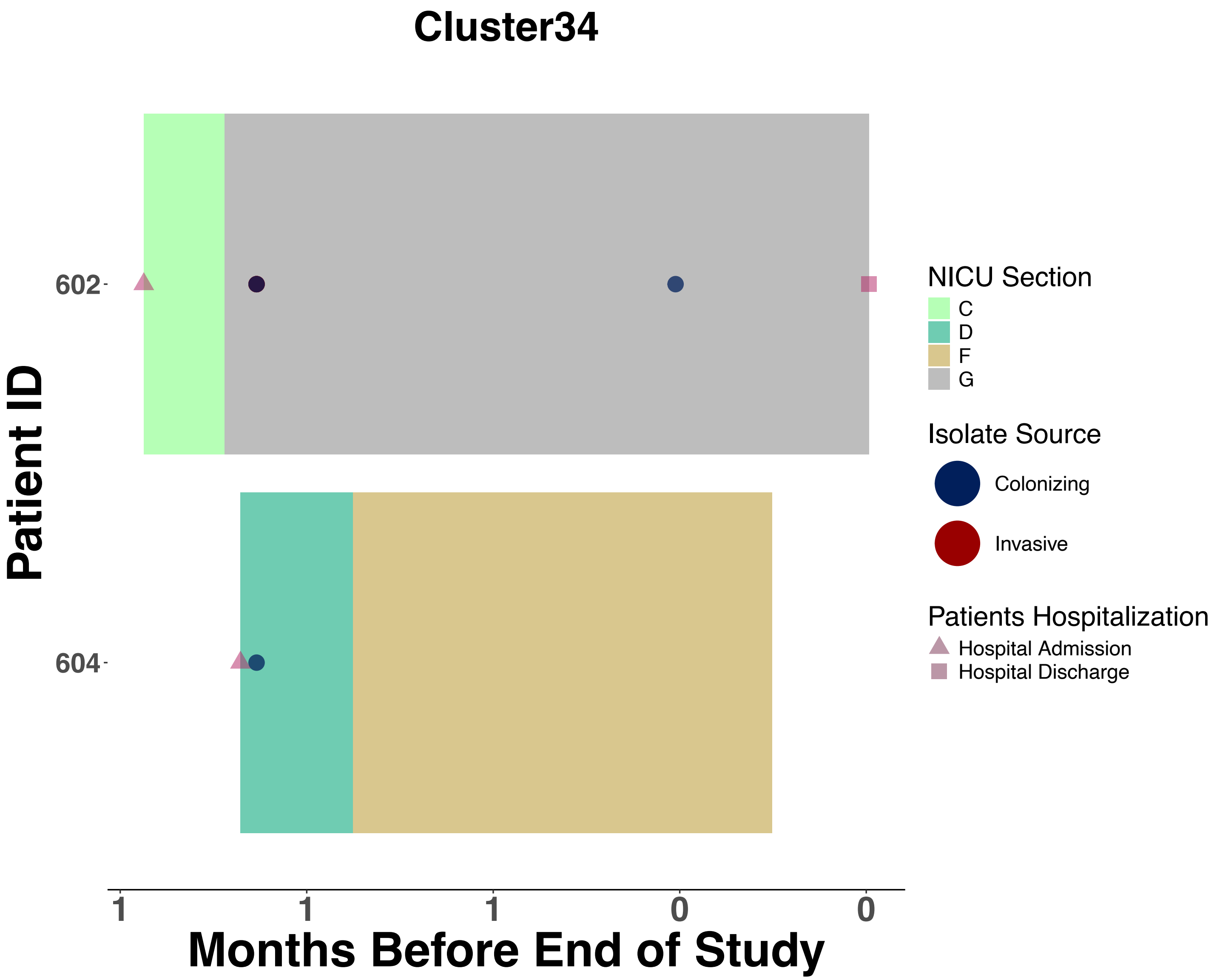

Cluster34
Patient ID
602
604
1
1
1
0
0
Months Before End of Study
NICU Section
C
D
F
G
Isolate Source
Colonizing
Invasive
Patients Hospitalization
Hospital Admission
Hospital Discharge

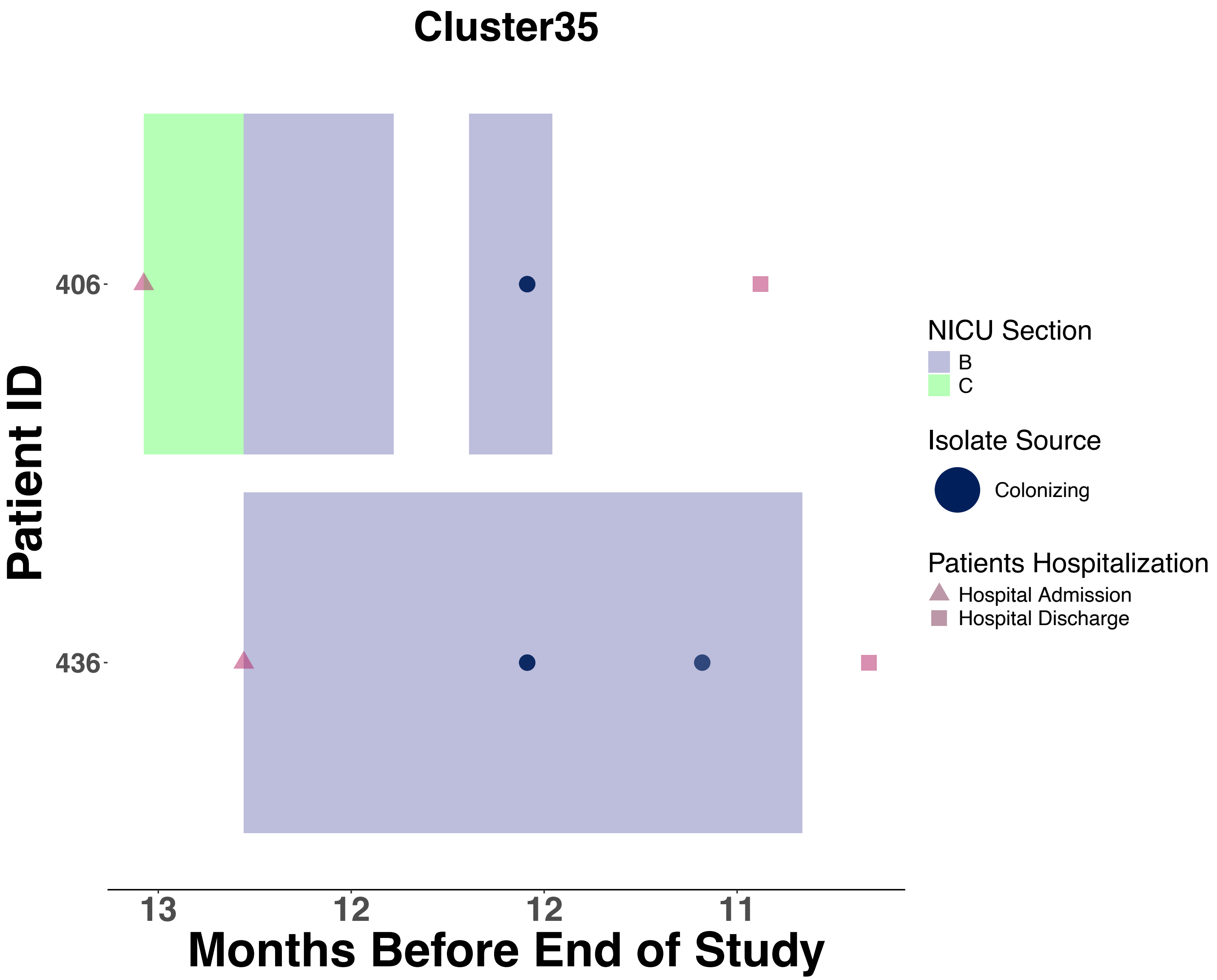

Cluster35
Patient ID
406
436
13
12
12
11
Months Before End of Study
NICU Section
B
C
Isolate Source
Colonizing
Patients Hospitalization
Hospital Admission
Hospital Discharge

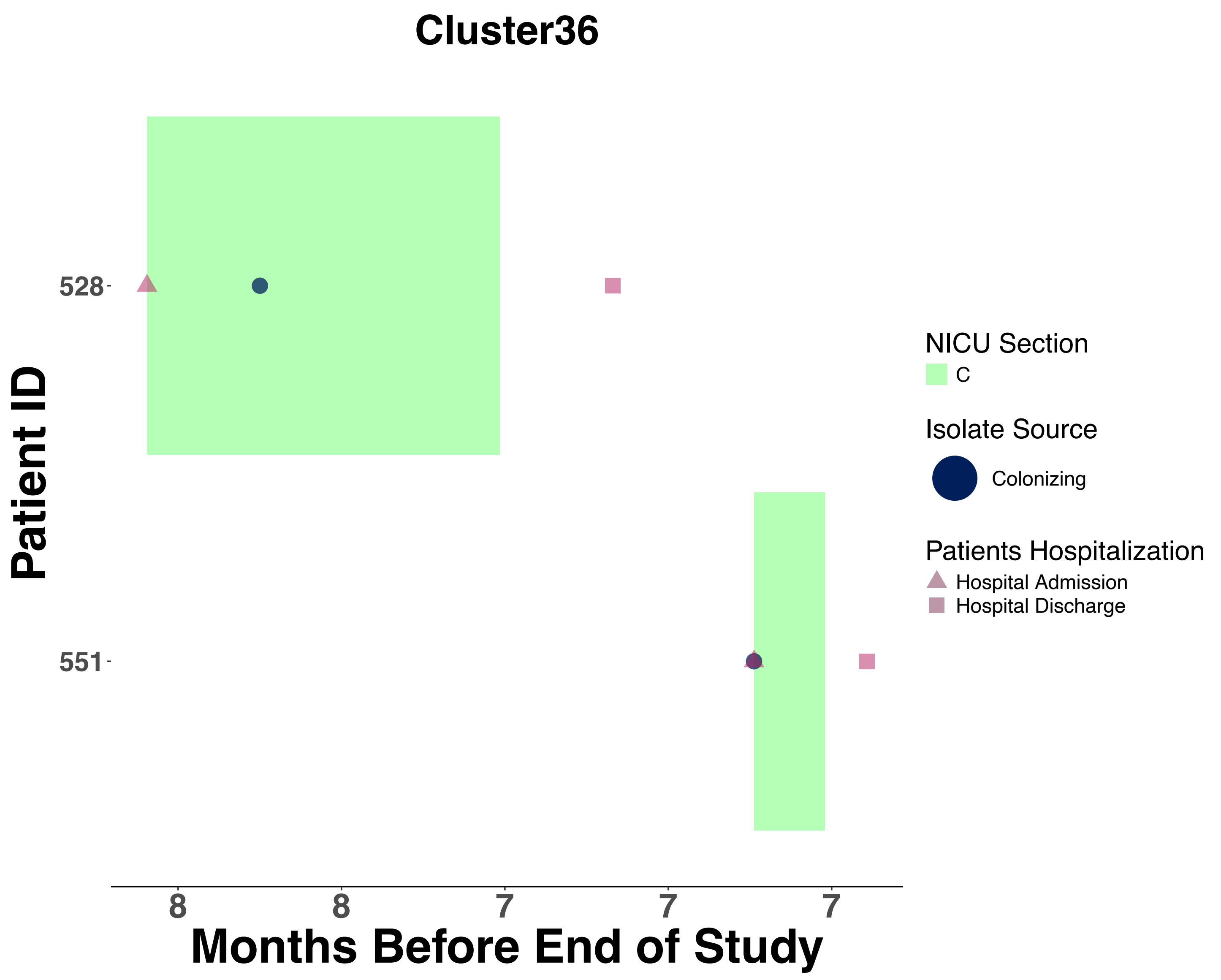
Cluster36
Patient ID
528
551
8
8
7
7
7
Months Before End of Study
NICU Section
C
Isolate Source
Colonizing
Patients Hospitalization
Hospital Admission
Hospital Discharge

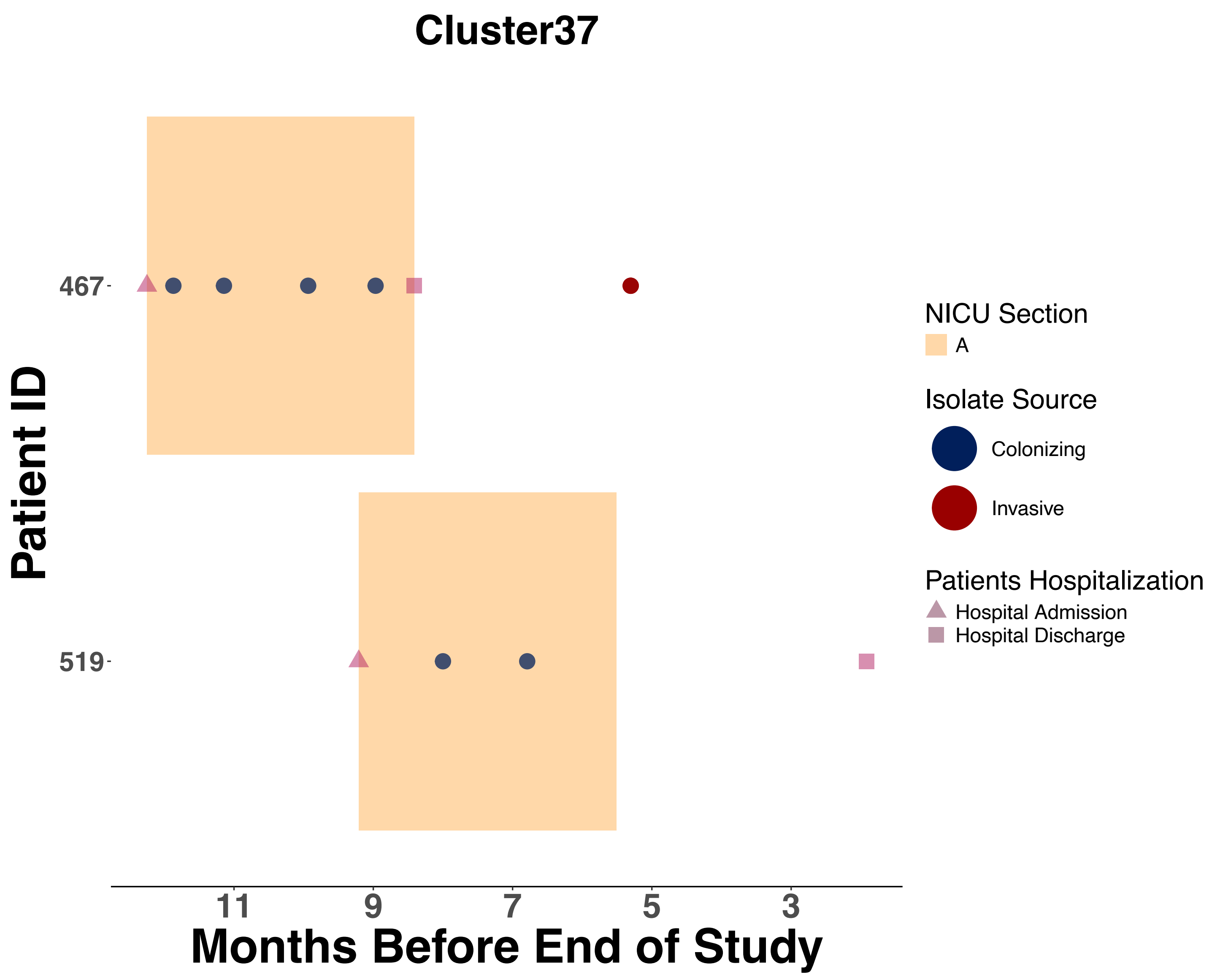

Cluster37
Patient ID
467
519
11
9
7
5
3
Months Before End of Study
NICU Section
A
Isolate Source
Colonizing
Invasive
Patients Hospitalization
Hospital Admission
Hospital Discharge

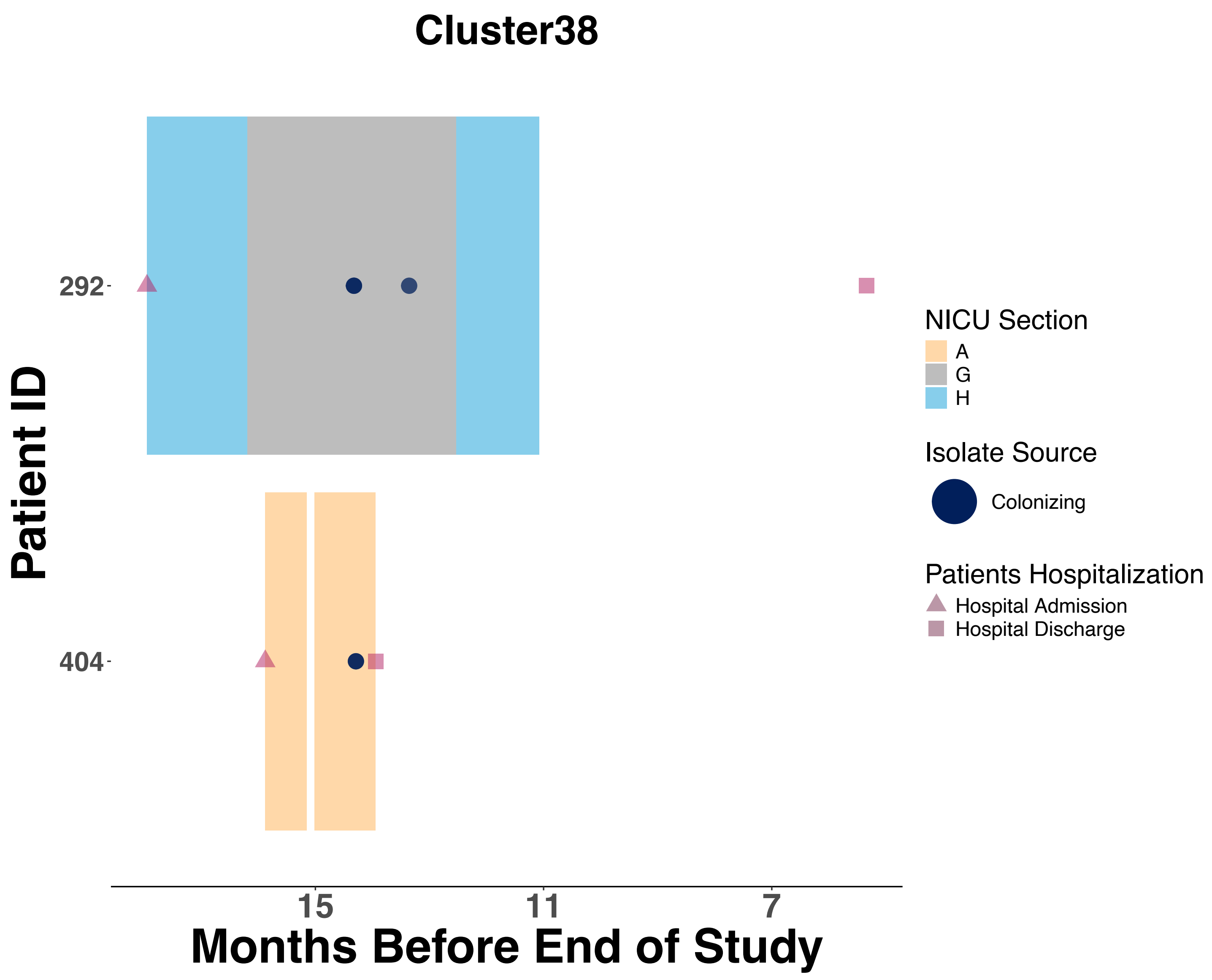
Cluster38
Patient ID
292
404
15
11
7
Months Before End of Study
NICU Section
A
G
H
Isolate Source
Colonizing
Patients Hospitalization
Hospital Admission
Hospital Discharge

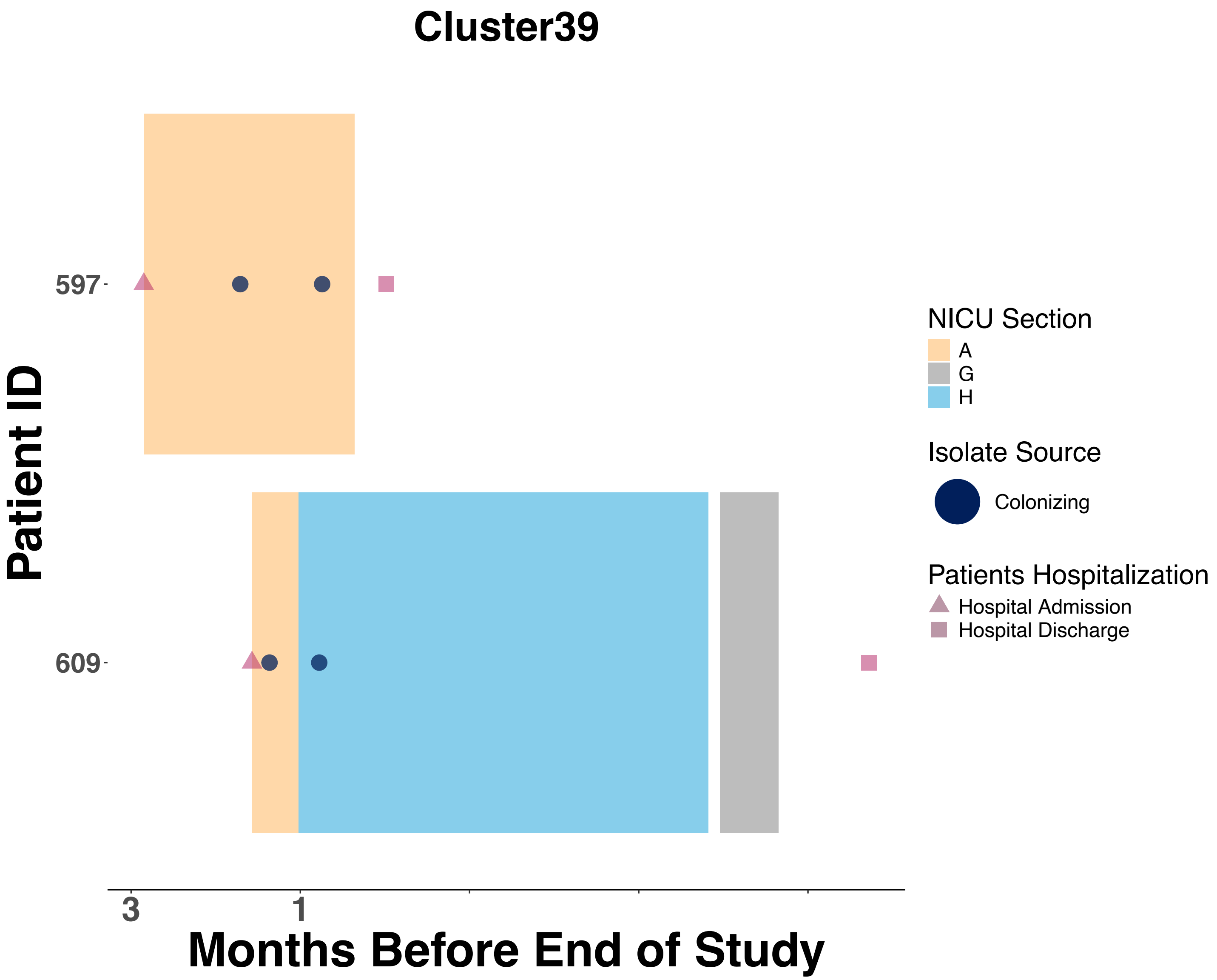
Cluster39
Patient ID
597
609
3
1
Months Before End of Study
NICU Section
A
G
H
Isolate Source
Colonizing
Patients Hospitalization
Hospital Admission
Hospital Discharge

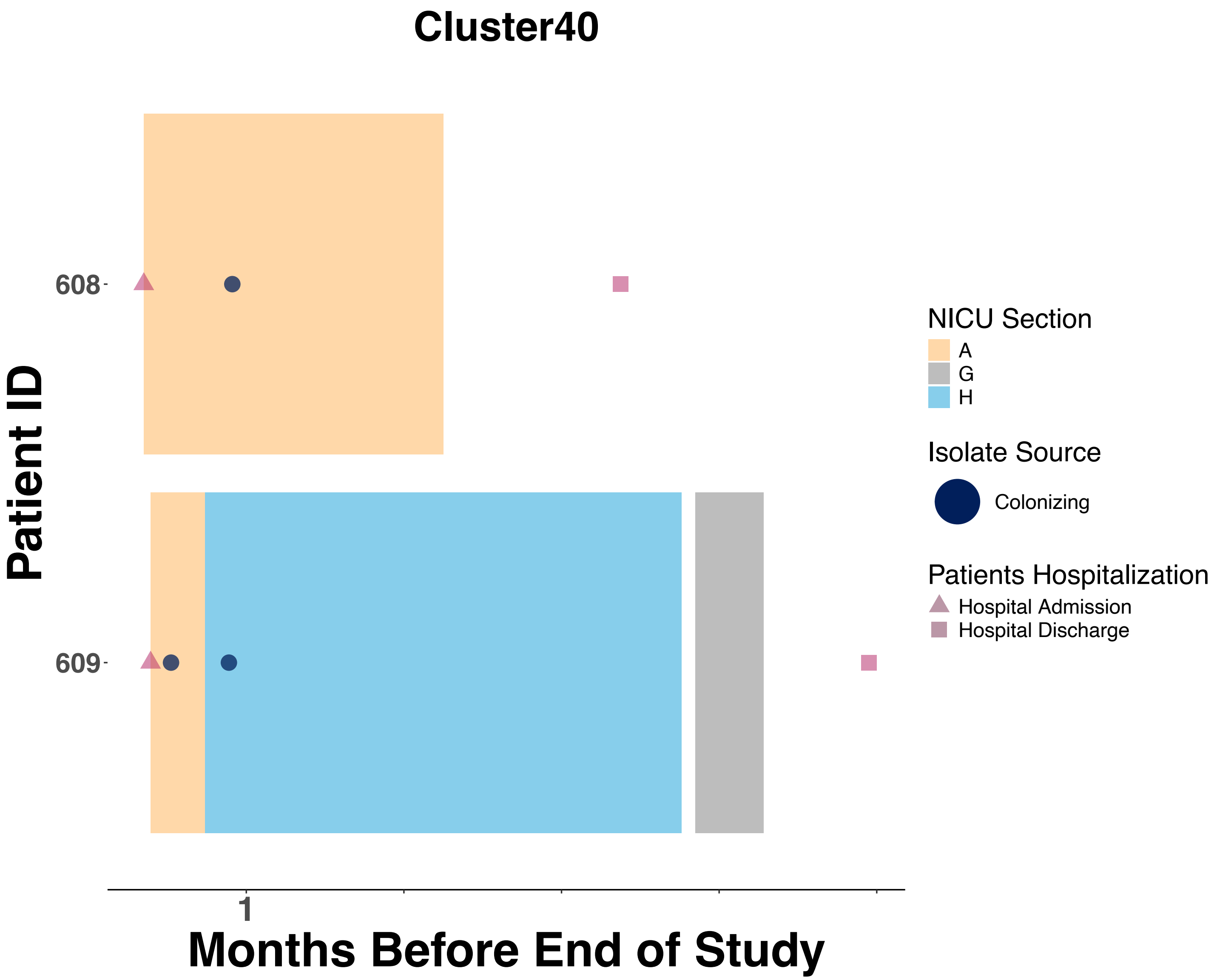

Cluster40
Patient ID
608
609
1
Months Before End of Study
NICU Section
A
G
H
Isolate Source
Colonizing
Patients Hospitalization
Hospital Admission
Hospital Discharge

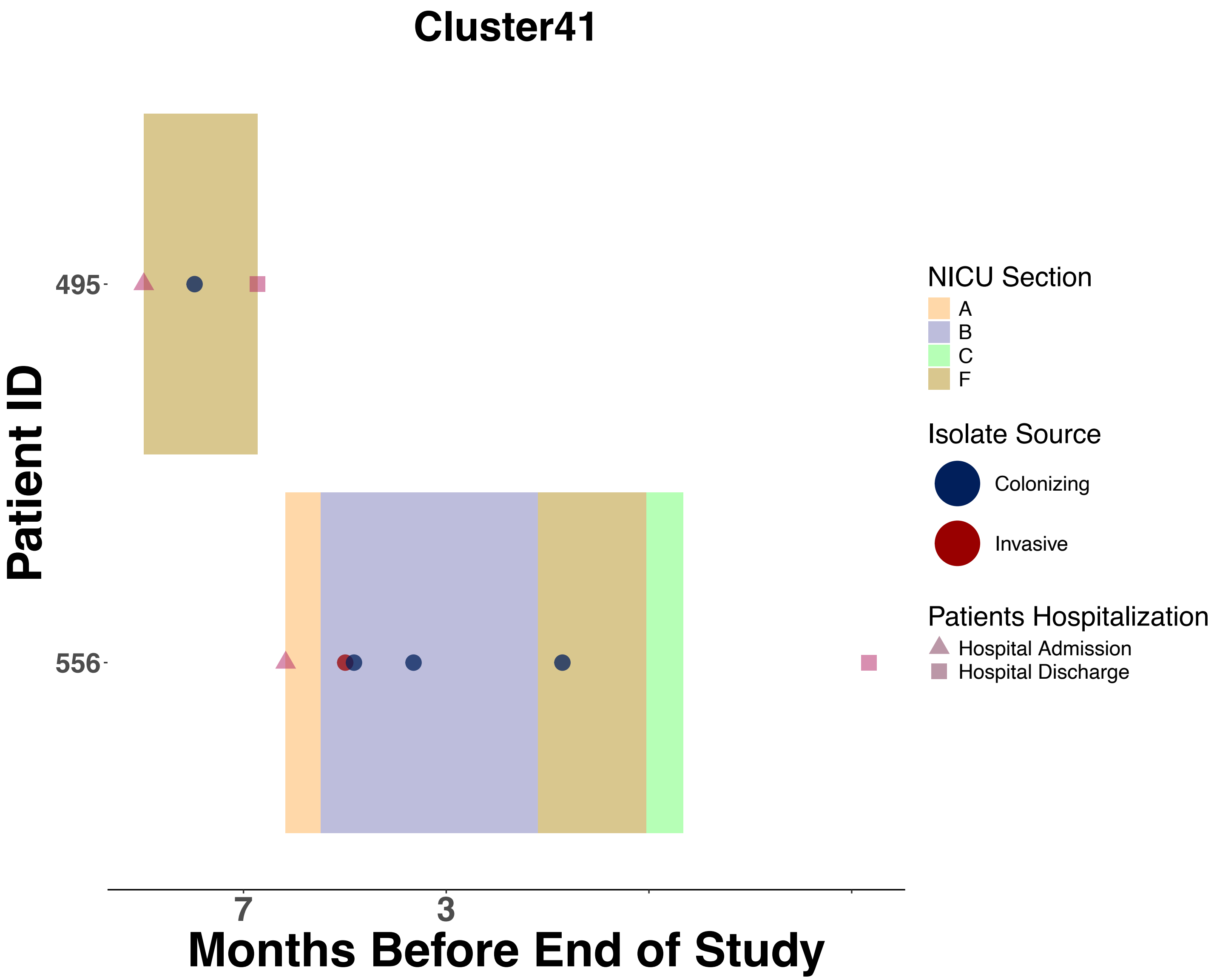

Cluster41
Patient ID
495
556
7
3
Months Before End of Study
NICU Section
A
B
C
F
Isolate Source
Colonizing
Invasive
Patients Hospitalization
Hospital Admission
Hospital Discharge

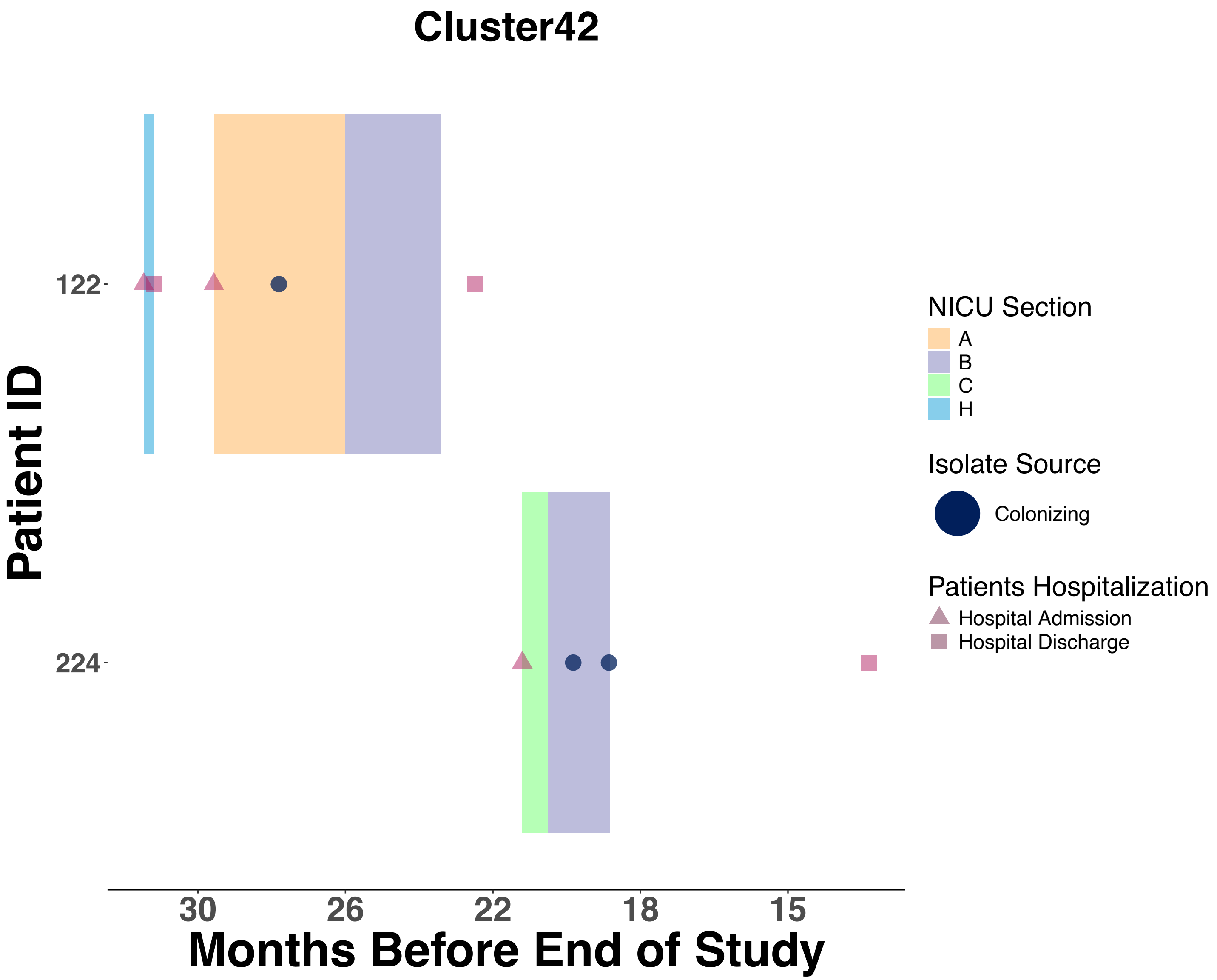

Cluster42
Patient ID
122
224
30
26
22
18
15
Months Before End of Study
NICU Section
A
B
C
H
Isolate Source
Colonizing
Patients Hospitalization
Hospital Admission
Hospital Discharge

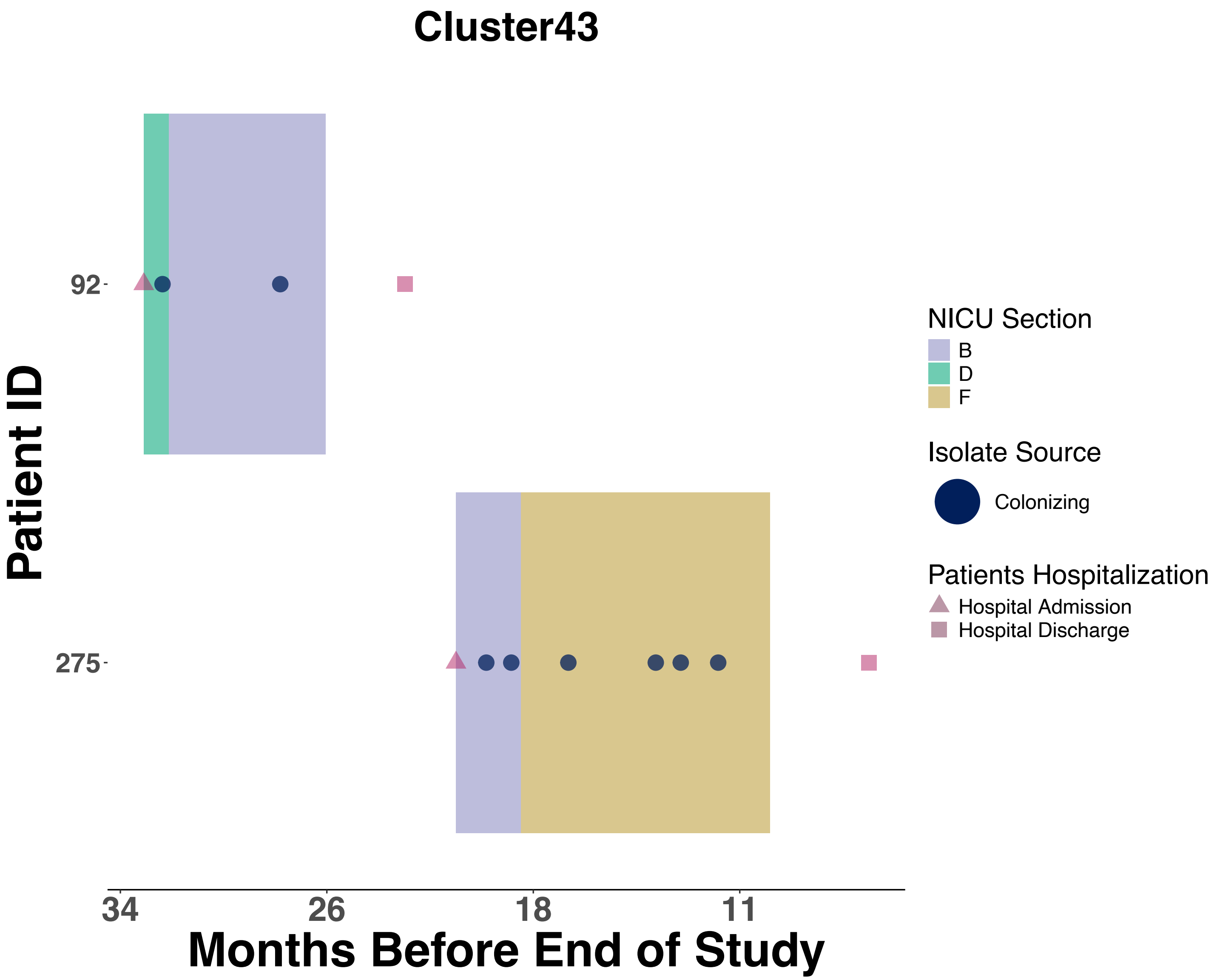
Cluster43
Patient ID
92
275
34
26
18
11
Months Before End of Study
NICU Section
B
D
F
Isolate Source
Colonizing
Patients Hospitalization
Hospital Admission
Hospital Discharge

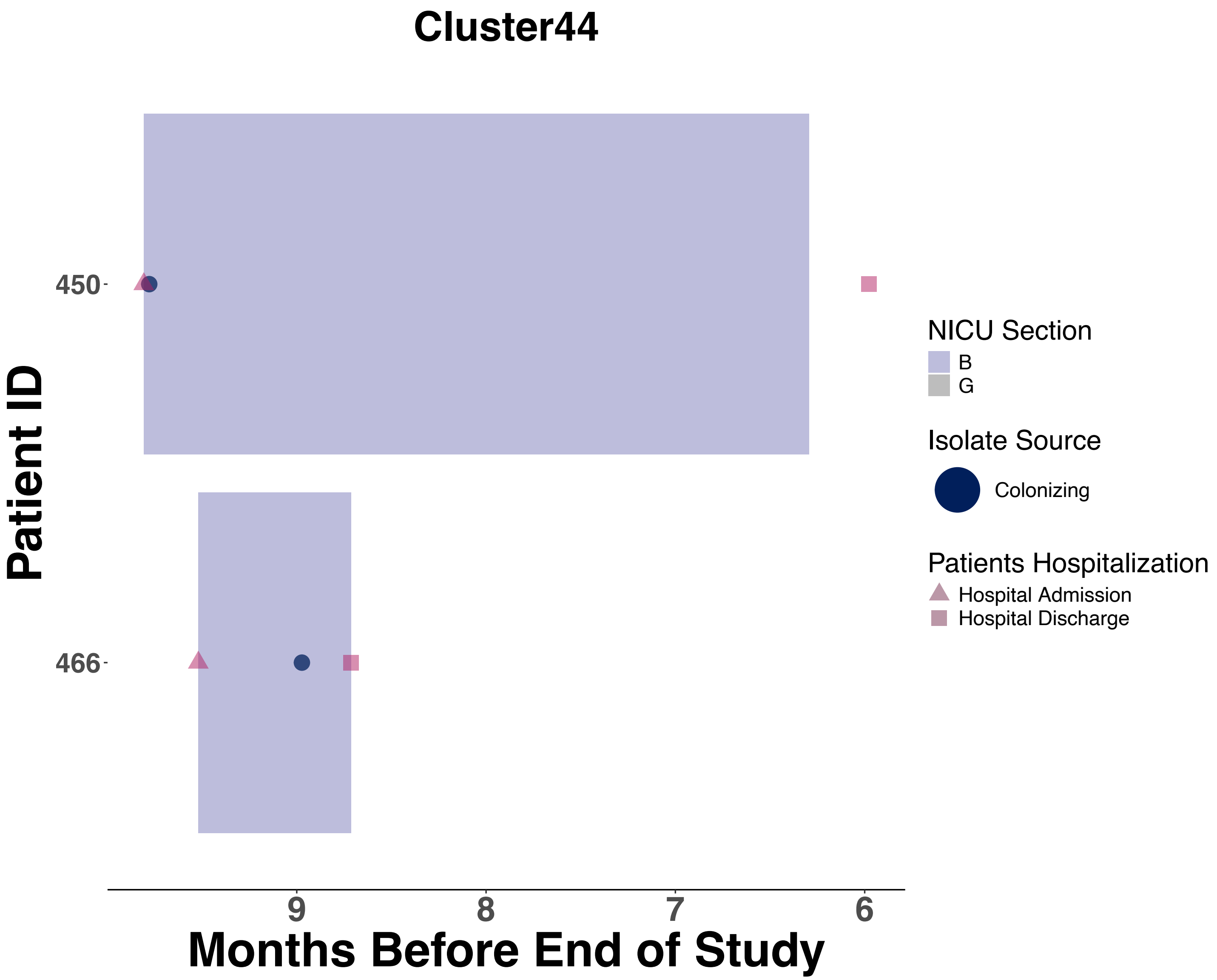

Cluster44
Patient ID
450
466
9
8
7
6
Months Before End of Study
NICU Section
B
G
Isolate Source
Colonizing
Patients Hospitalization
Hospital Admission
Hospital Discharge

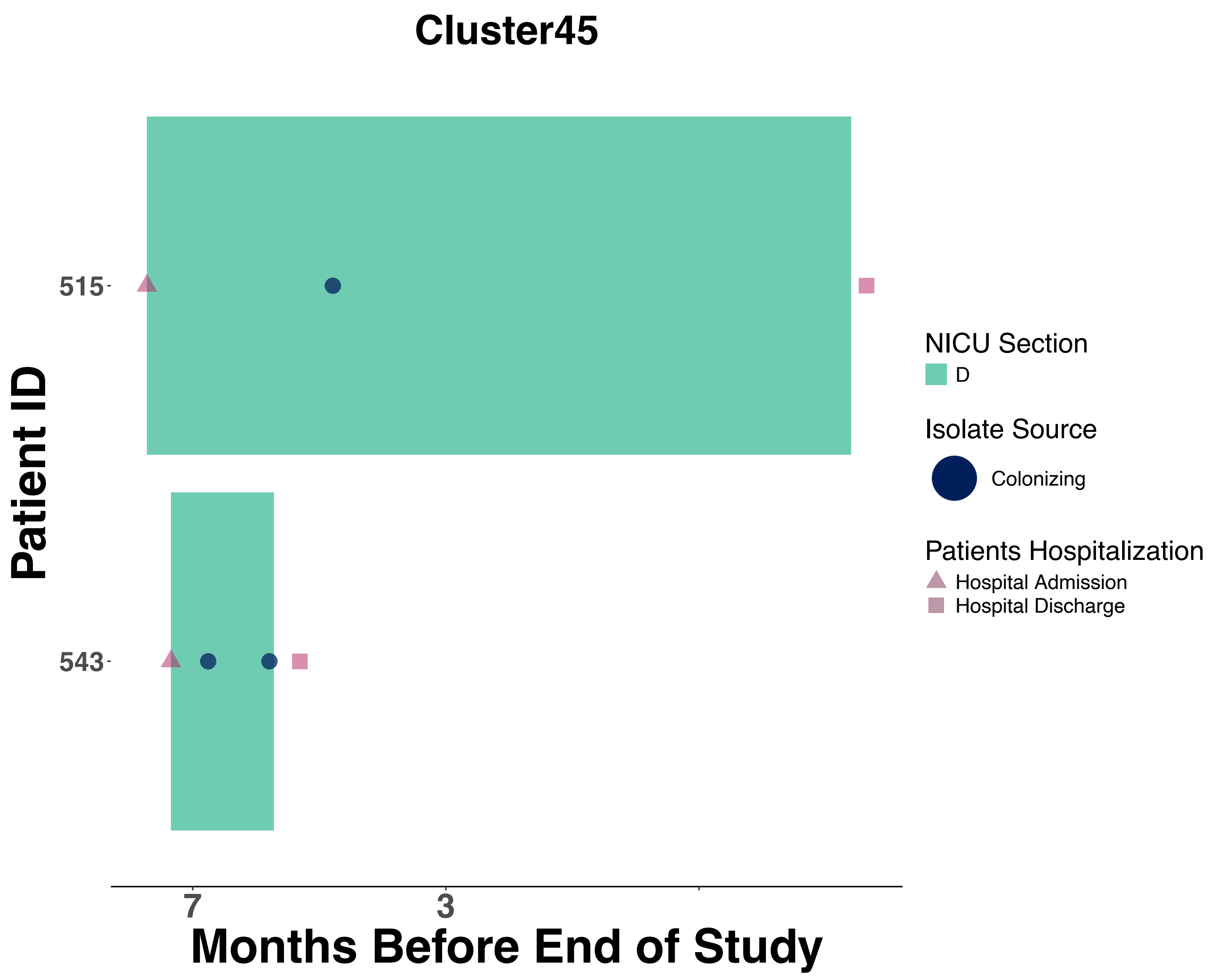

Cluster45
515
543
7
3
Months Before End of Study
Patient ID
NICU Section
D
Isolate Source
Colonizing
Patients Hospitalization
Hospital Admission
Hospital Discharge

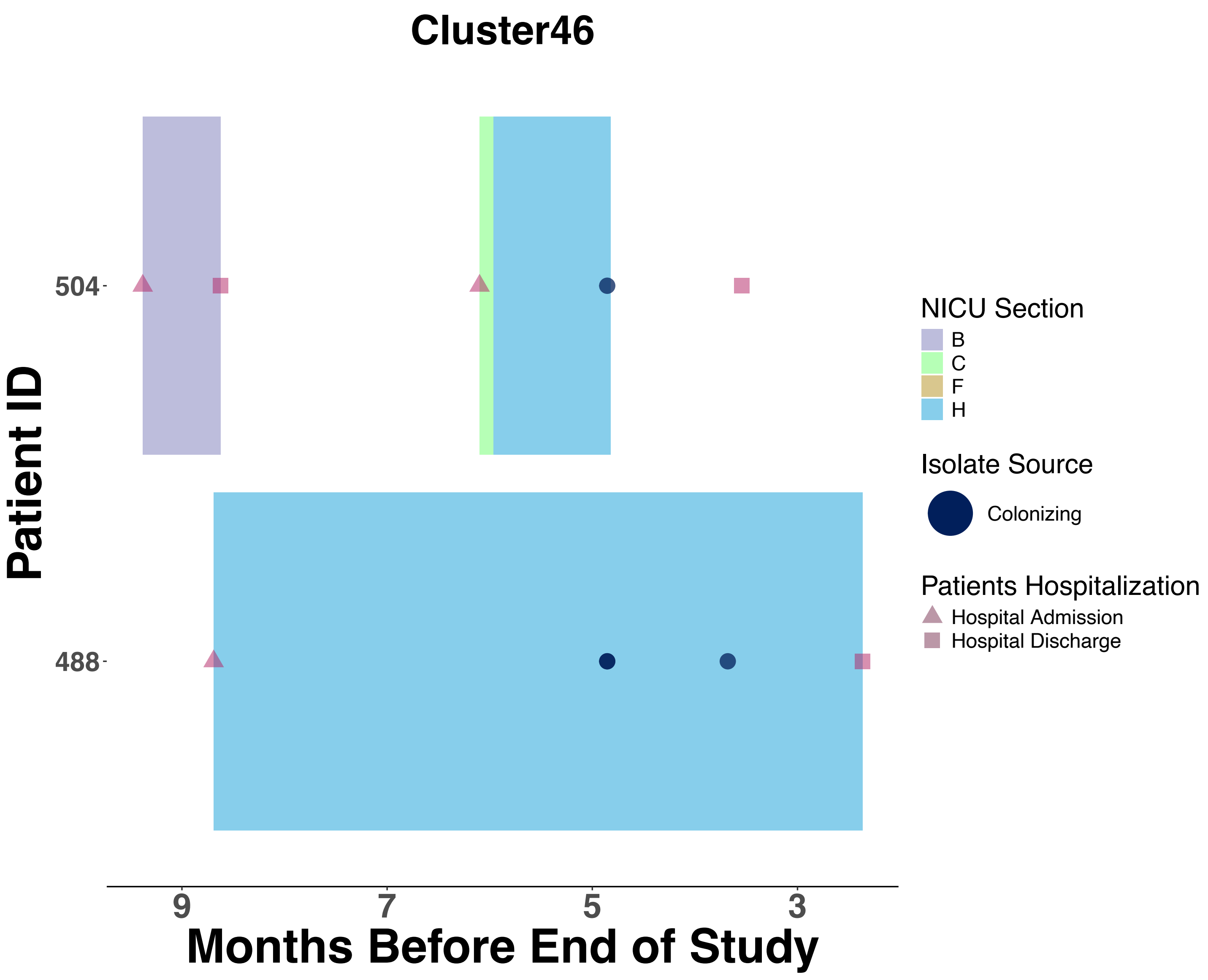

Cluster46
Patient ID
504
488
9
7
5
3
Months Before End of Study
NICU Section
B
C
F
H
Isolate Source
Colonizing
Patients Hospitalization
Hospital Admission
Hospital Discharge

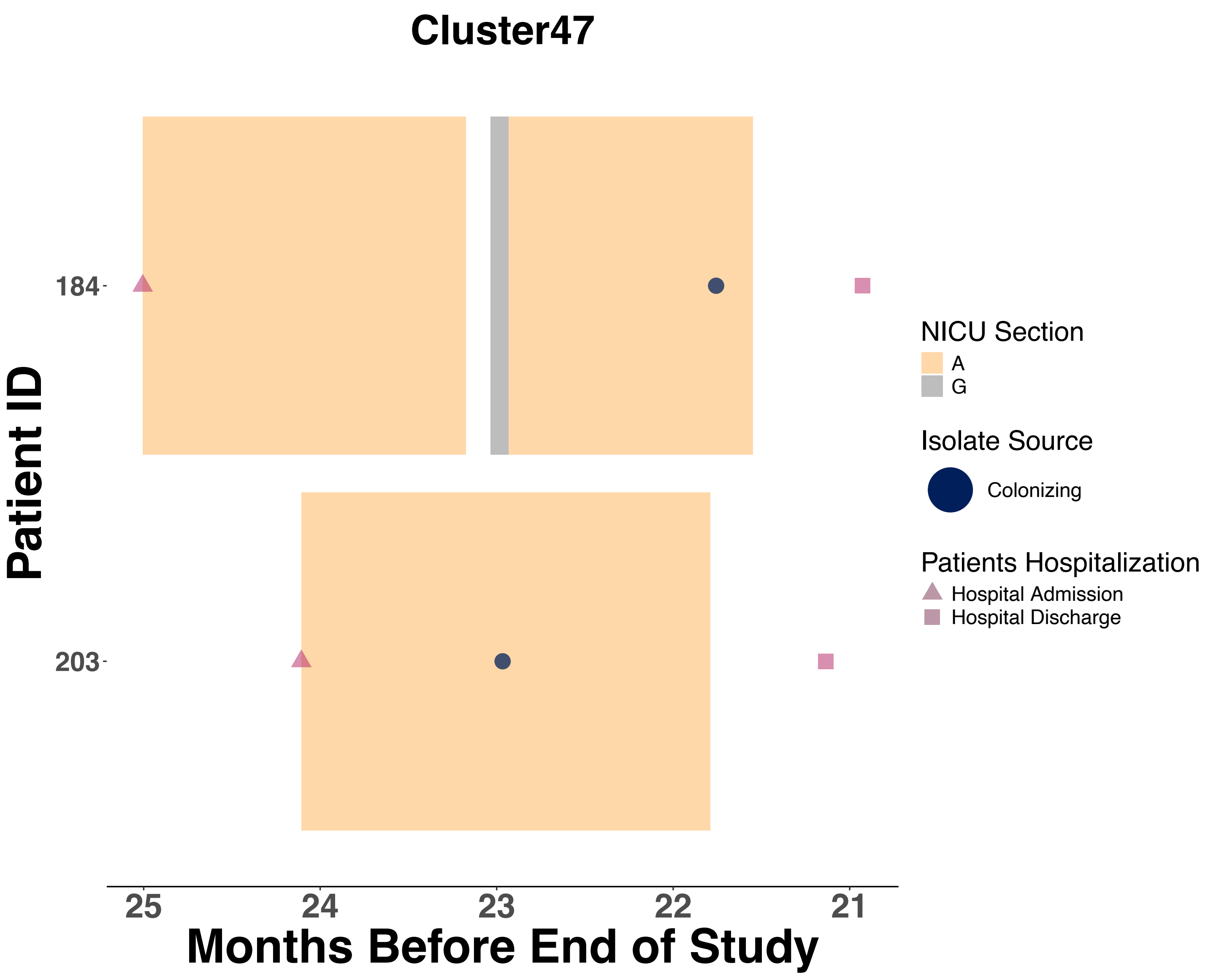
Cluster47
Patient ID
184
203
25
24
23
22
21
Months Before End of Study
NICU Section
A
G
Isolate Source
Colonizing
Patients Hospitalization
Hospital Admission
Hospital Discharge

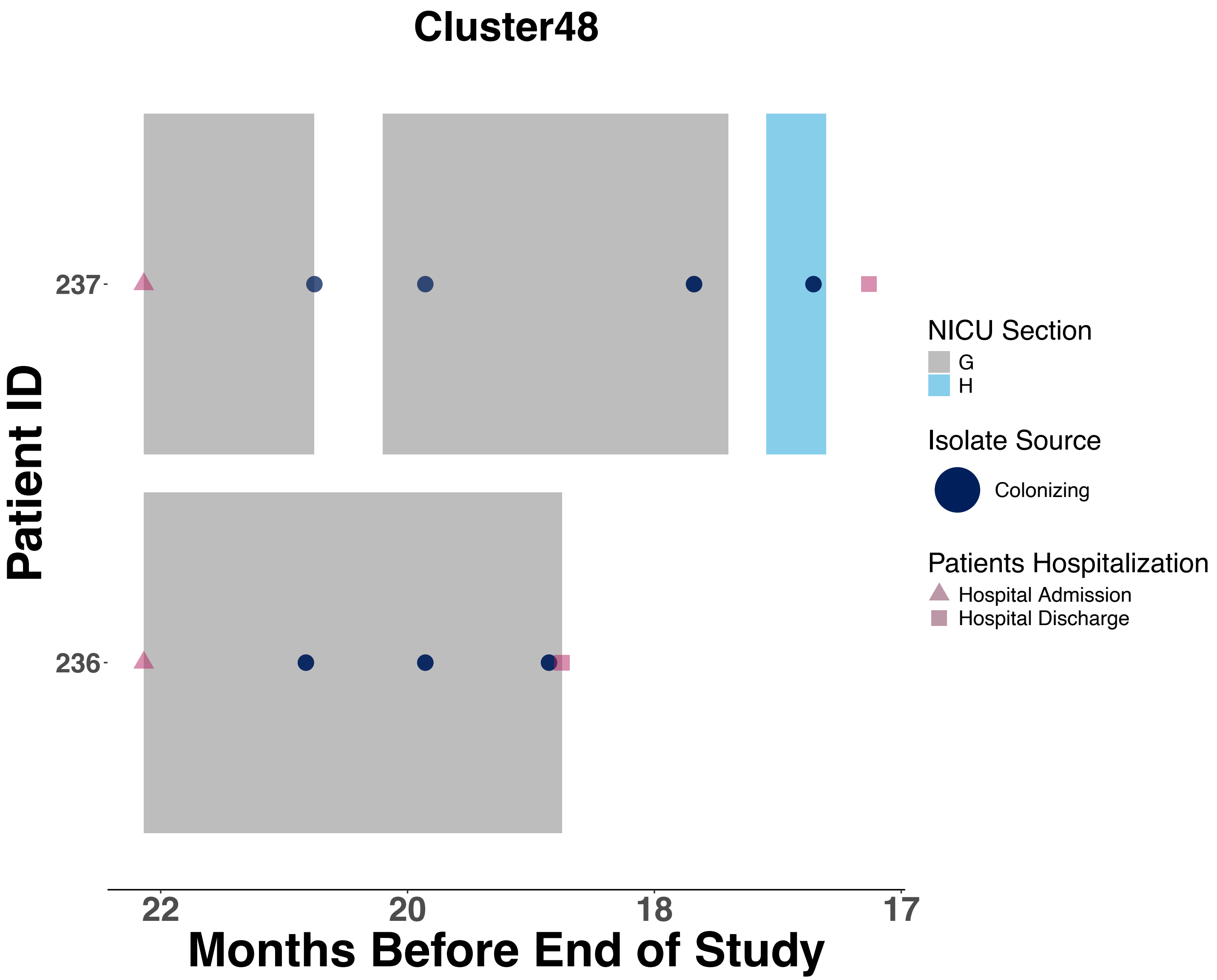

Cluster48
Patient ID
237
236
22
20
18
17
Months Before End of Study
NICU Section
G
H
Isolate Source
Colonizing
Patients Hospitalization
Hospital Admission
Hospital Discharge

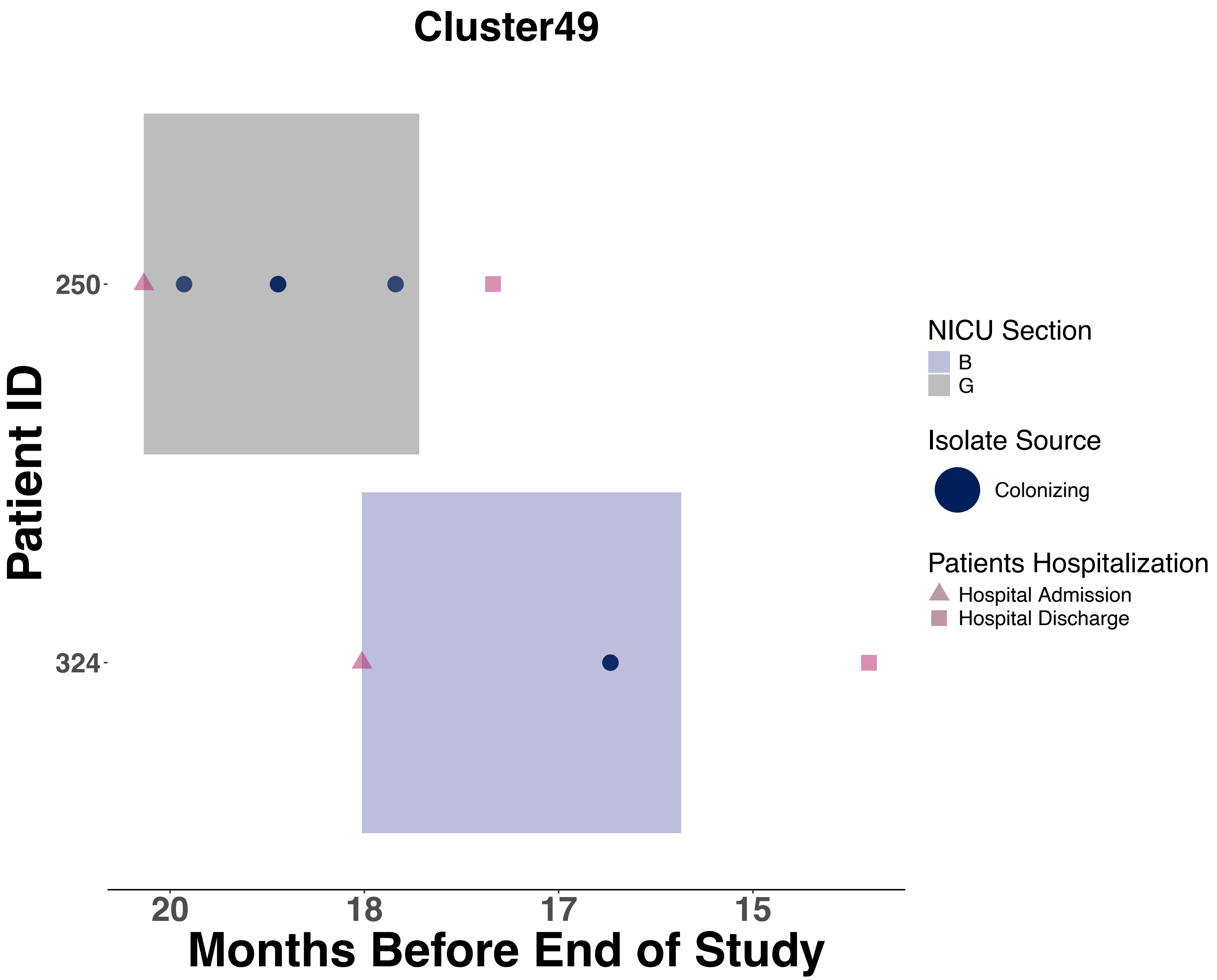
Cluster49
Patient ID
250
324
20
18
17
15
Months Before End of Study
NICU Section
B
G
Isolate Source
Colonizing
Patients Hospitalization
Hospital Admission
Hospital Discharge

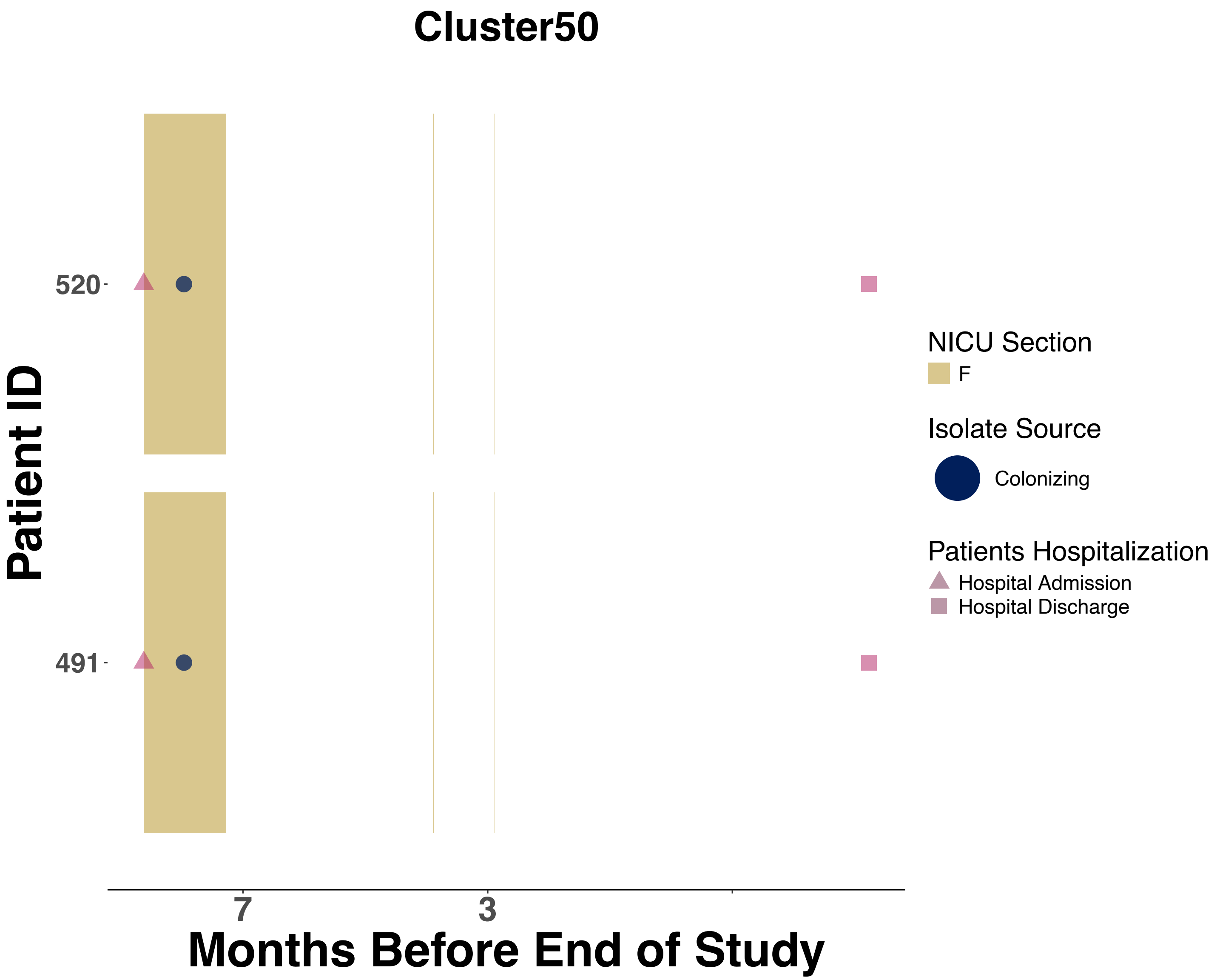
Cluster50
Patient ID
520
491
7
3
Months Before End of Study
NICU Section
F
Isolate Source
Colonizing
Patients Hospitalization
Hospital Admission
Hospital Discharge

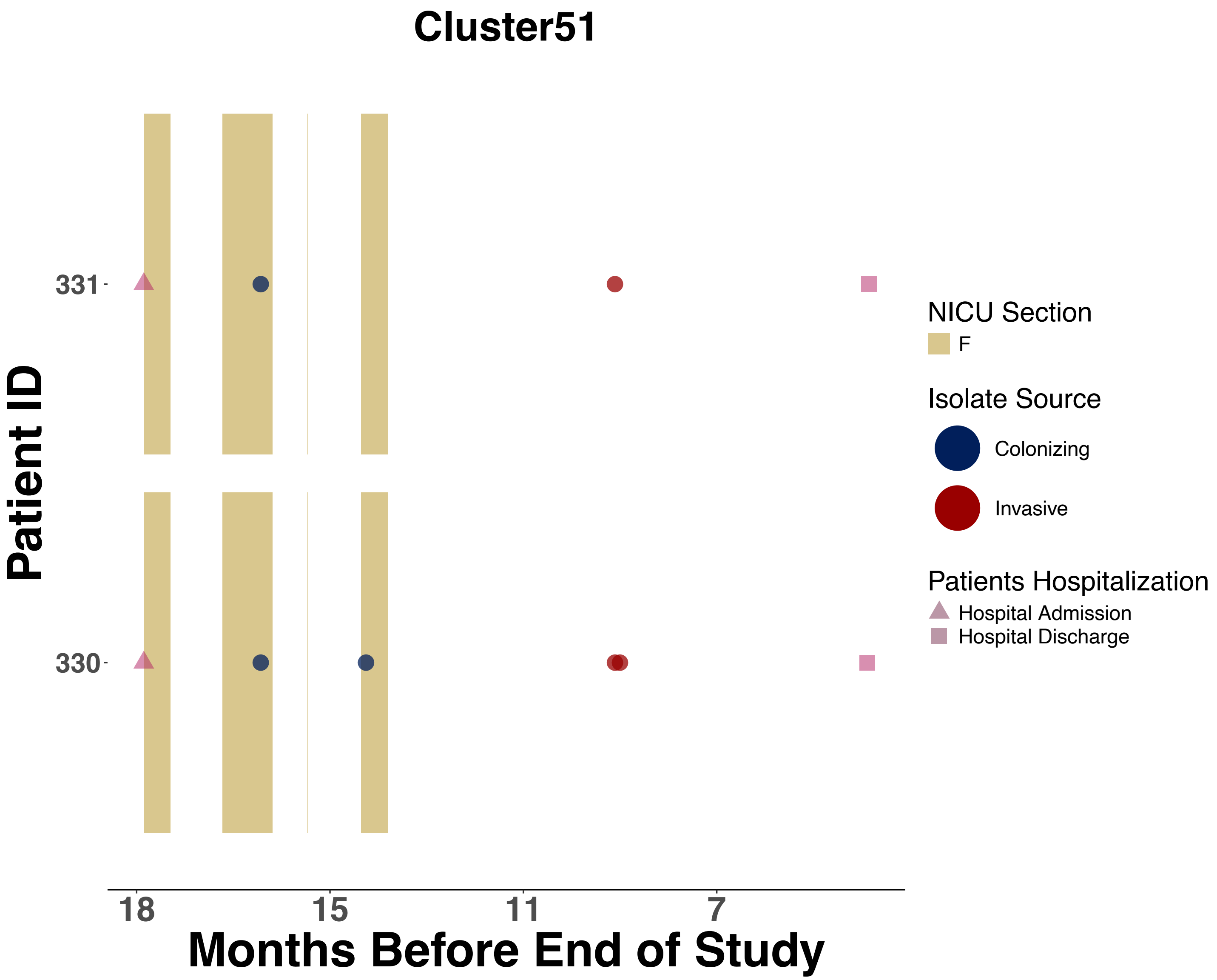
Cluster51
Patient ID
331
330
18
15
11
7
Months Before End of Study
NICU Section
F
Isolate Source
Colonizing
Invasive
Patients Hospitalization
Hospital Admission
Hospital Discharge

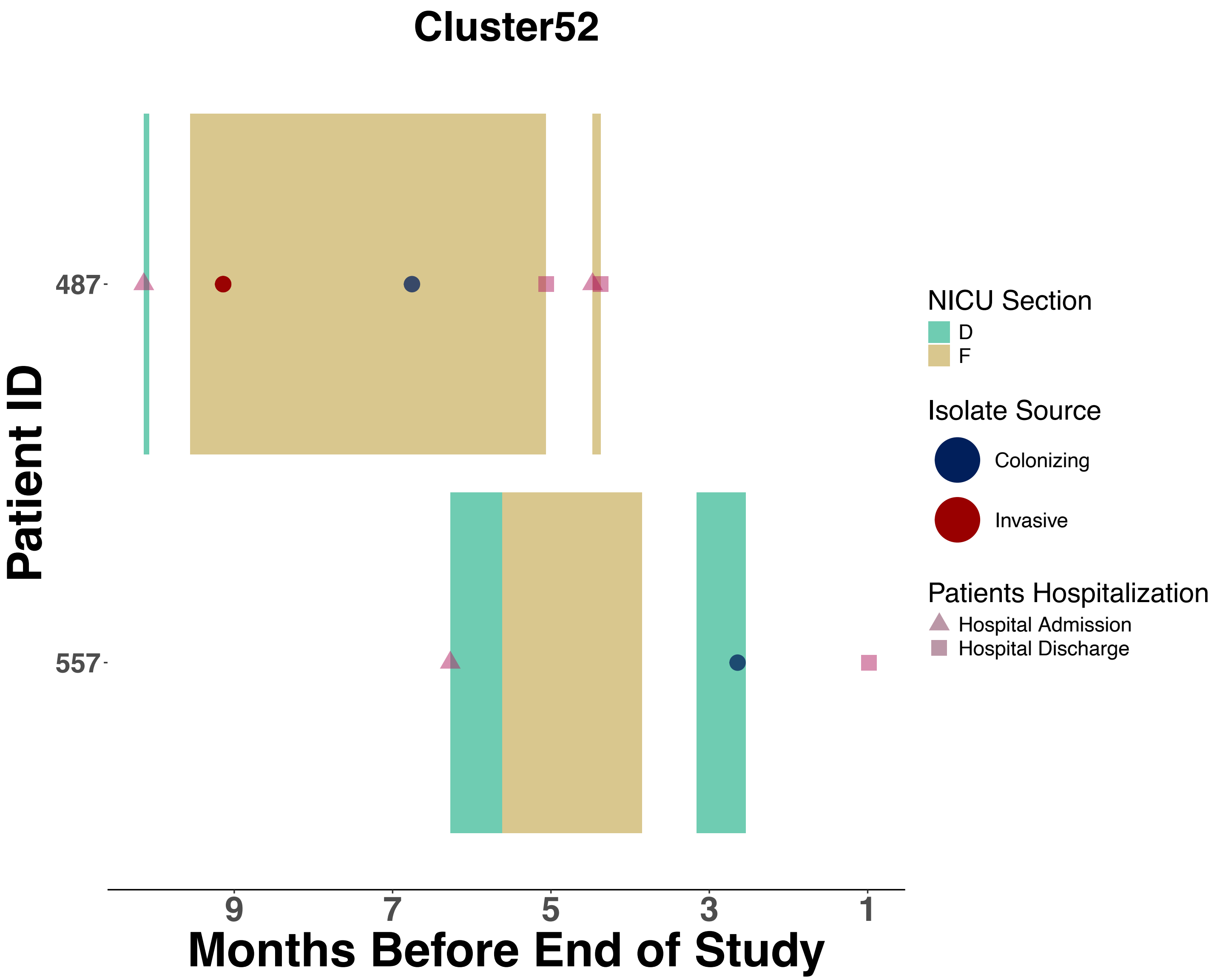

Cluster52
Patient ID
487
557
9
7
5
3
1
Months Before End of Study
NICU Section
D
F
Isolate Source
Colonizing
Invasive
Patients Hospitalization
Hospital Admission
Hospital Discharge

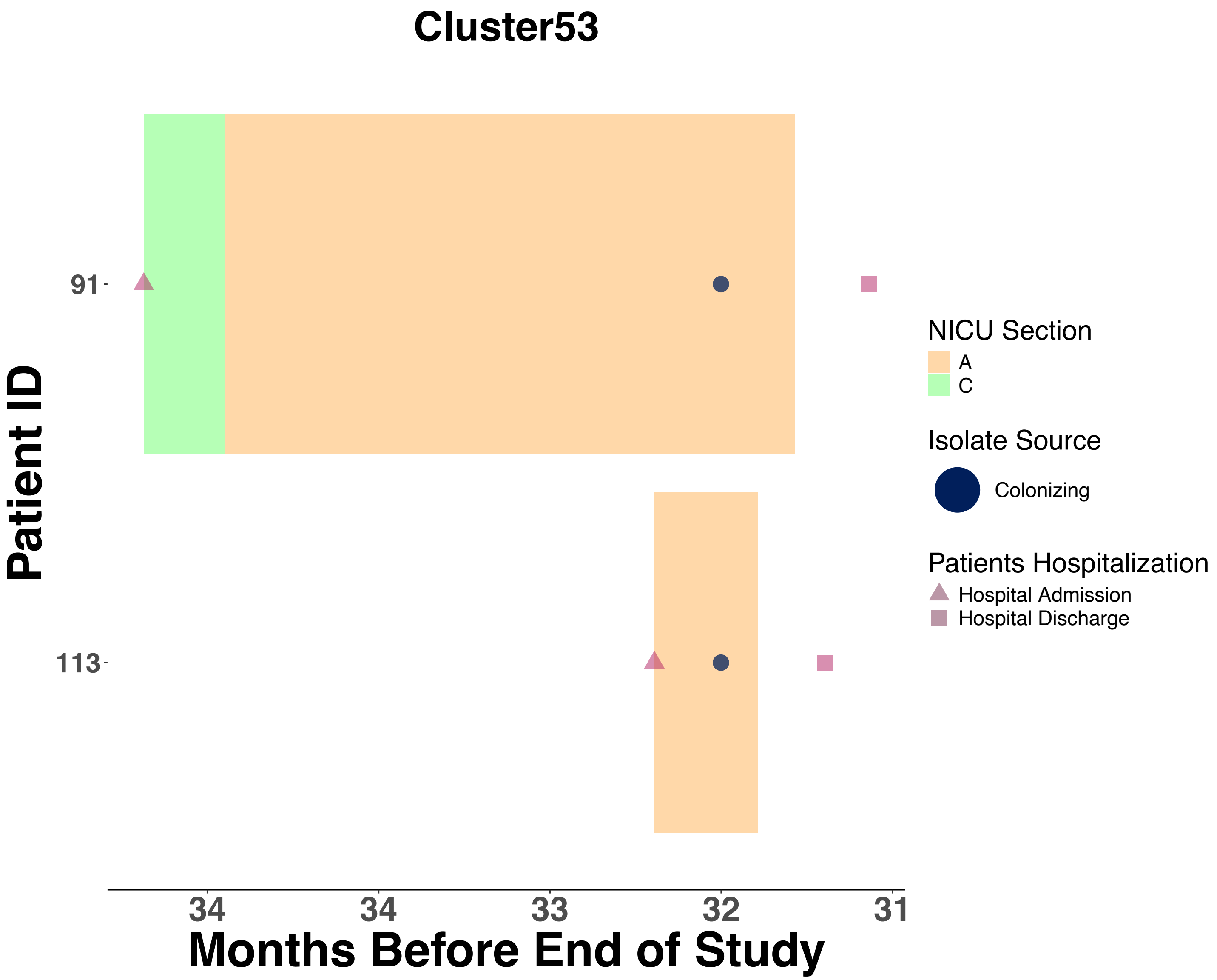
Cluster53
Patient ID
91
113
Months Before End of Study
34
34
33
32
31
NICU Section
A
C
Isolate Source
Colonizing
Patients Hospitalization
Hospital Admission
Hospital Discharge

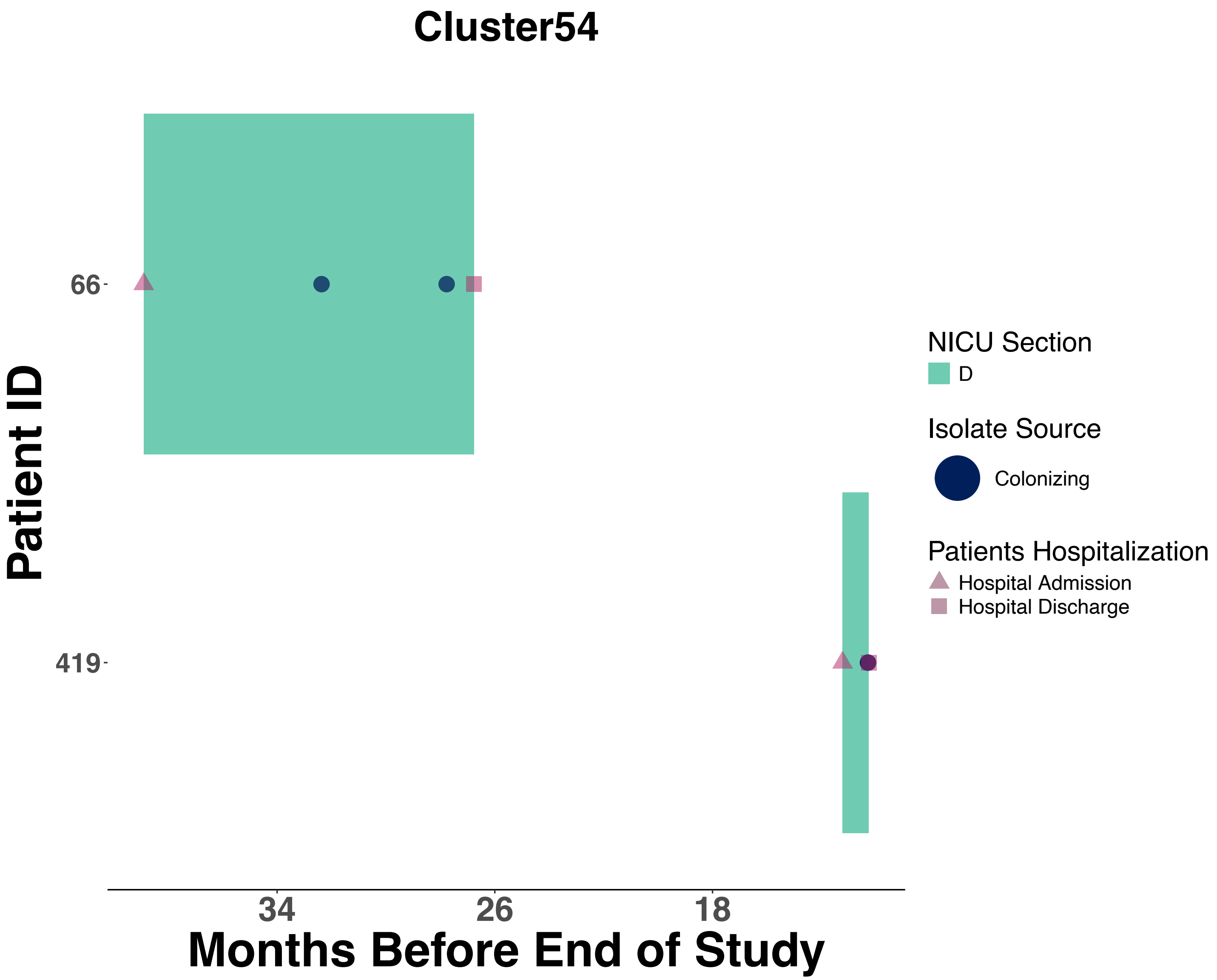

Cluster54
Patient ID
66
419
34
26
18
Months Before End of Study
NICU Section
D
Isolate Source
Colonizing
Patients Hospitalization
Hospital Admission
Hospital Discharge

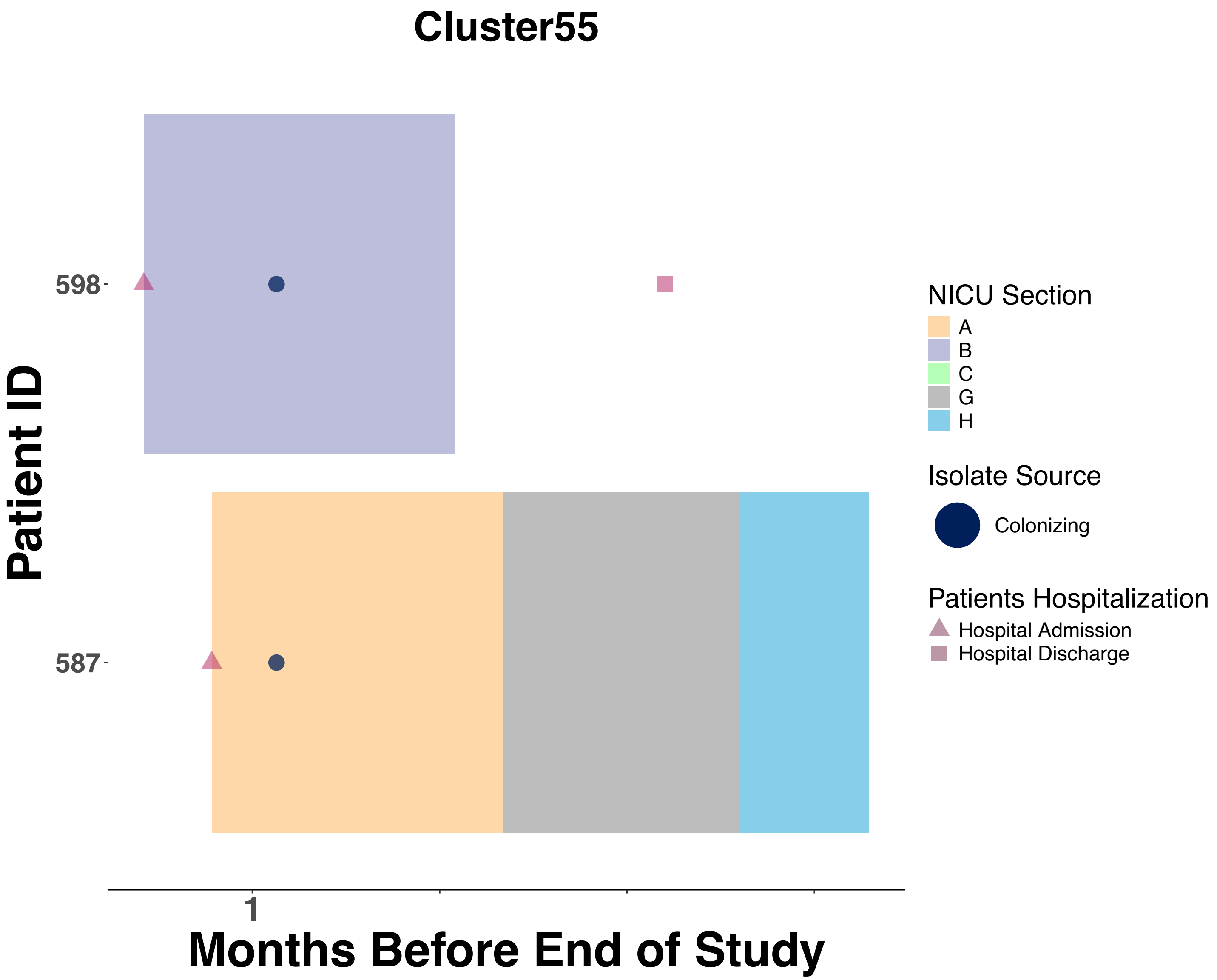
Cluster55
Patient ID
598
587
1
Months Before End of Study
NICU Section
A
B
C
G
H
Isolate Source
Colonizing
Patients Hospitalization
Hospital Admission
Hospital Discharge

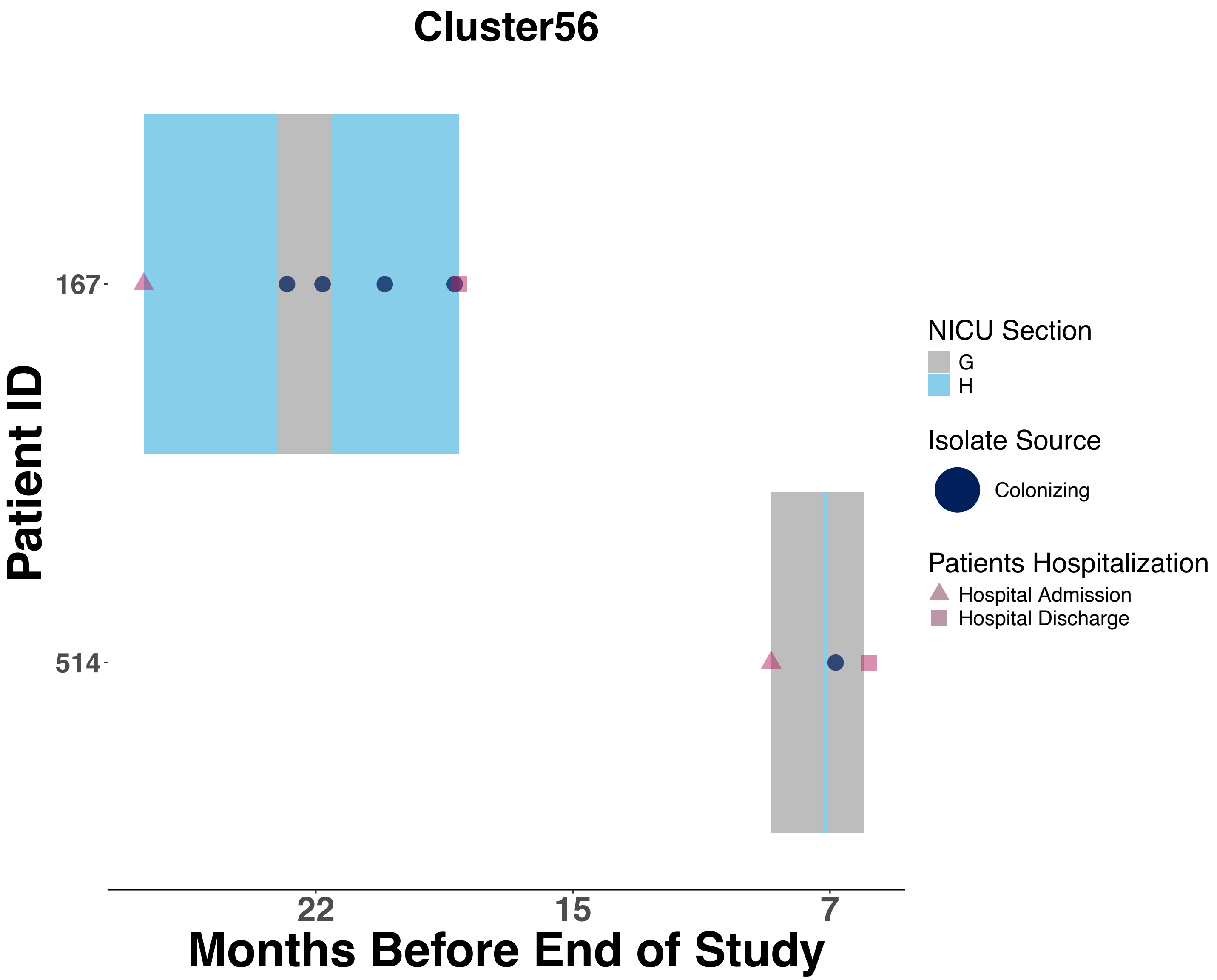

Cluster56
Patient ID
167
514
22
15
7
Months Before End of Study
NICU Section
G
H
Isolate Source
Colonizing
Patients Hospitalization
Hospital Admission
Hospital Discharge

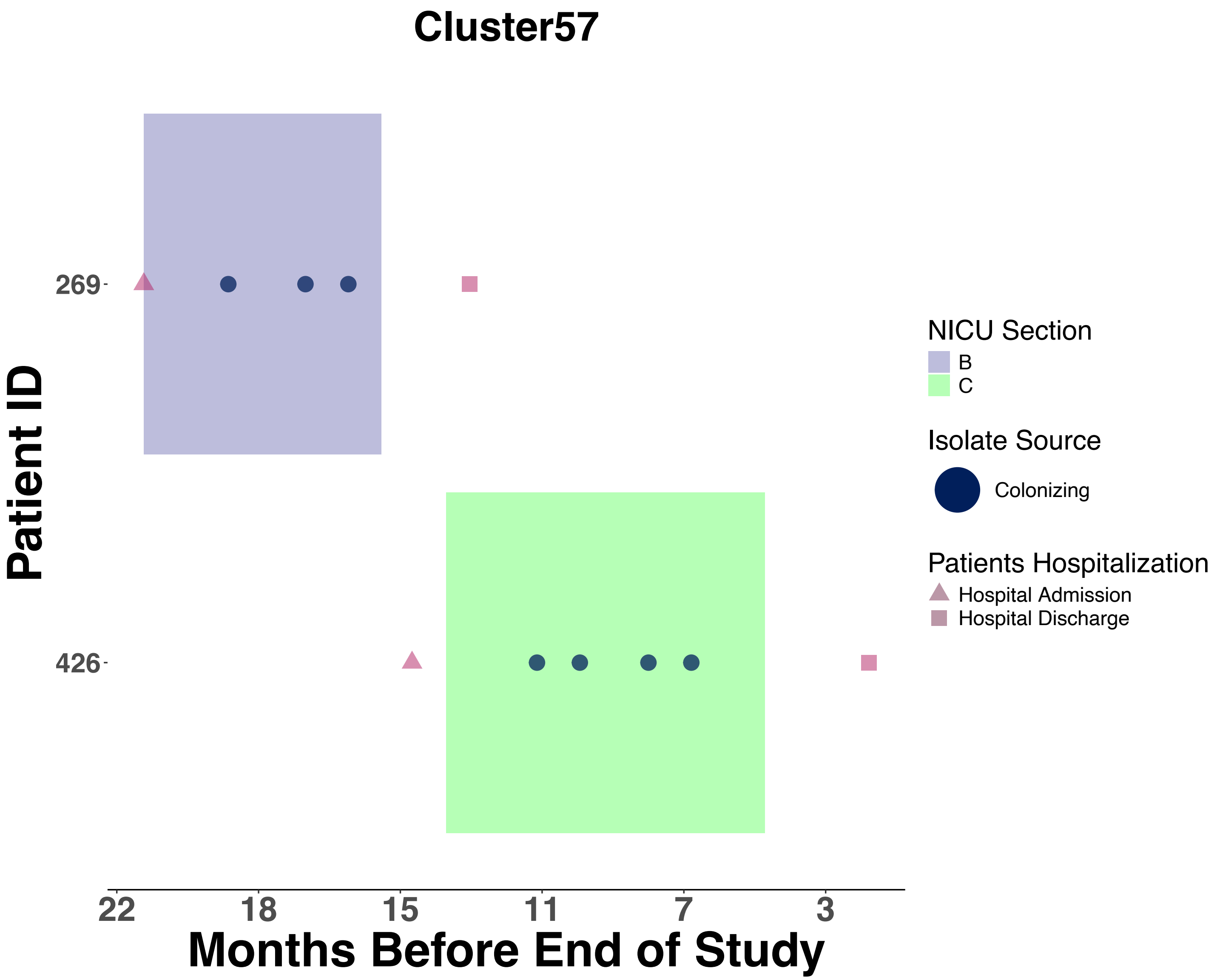
Cluster57
Patient ID
269
426
22
18
15
11
7
3
Months Before End of Study
NICU Section
B
C
Isolate Source
Colonizing
Patients Hospitalization
Hospital Admission
Hospital Discharge

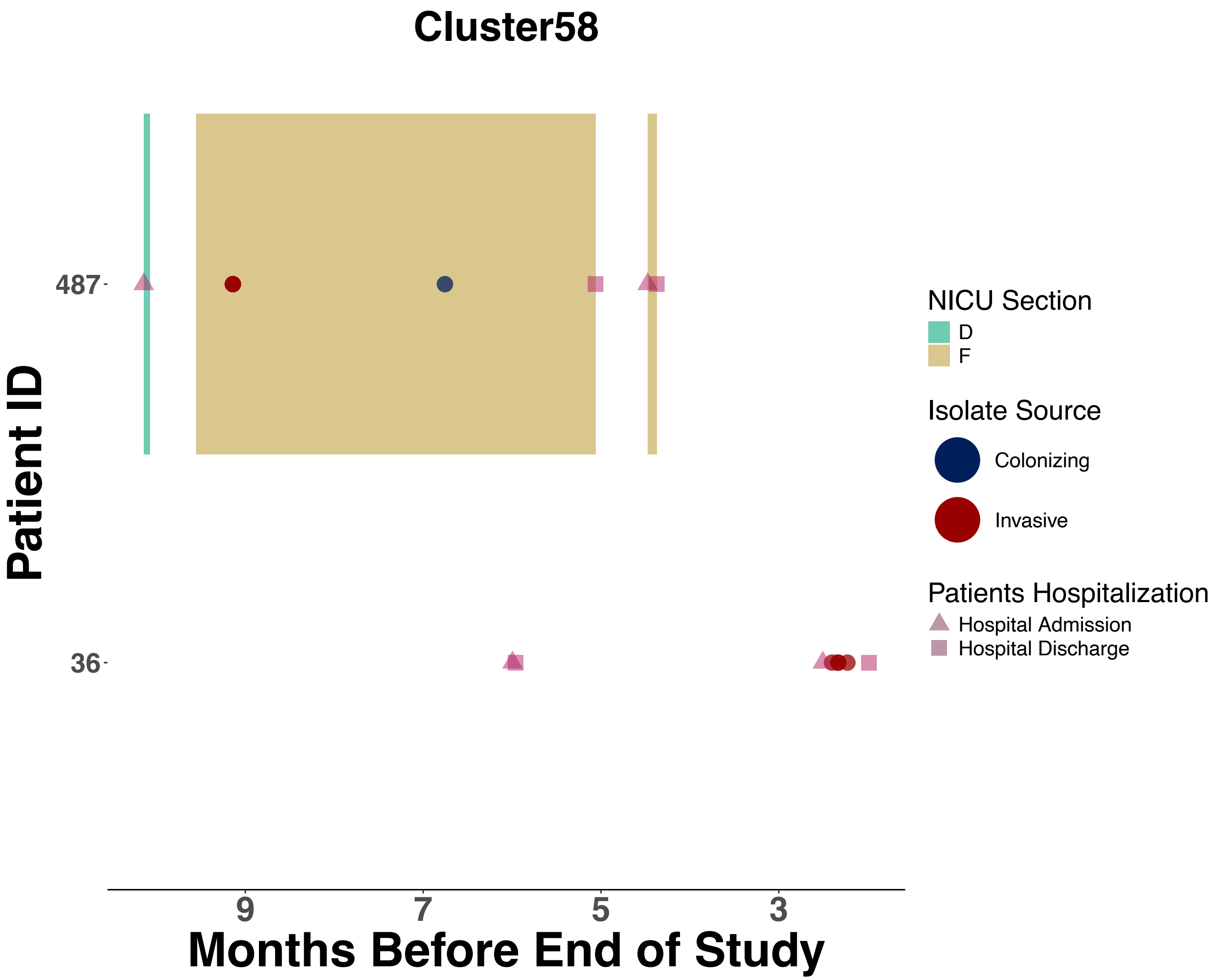

Cluster58
487
36
Patient ID
9
7
5
3
Months Before End of Study
NICU Section
D
F
Isolate Source
Colonizing
Invasive
Patients Hospitalization
Hospital Admission
Hospital Discharge

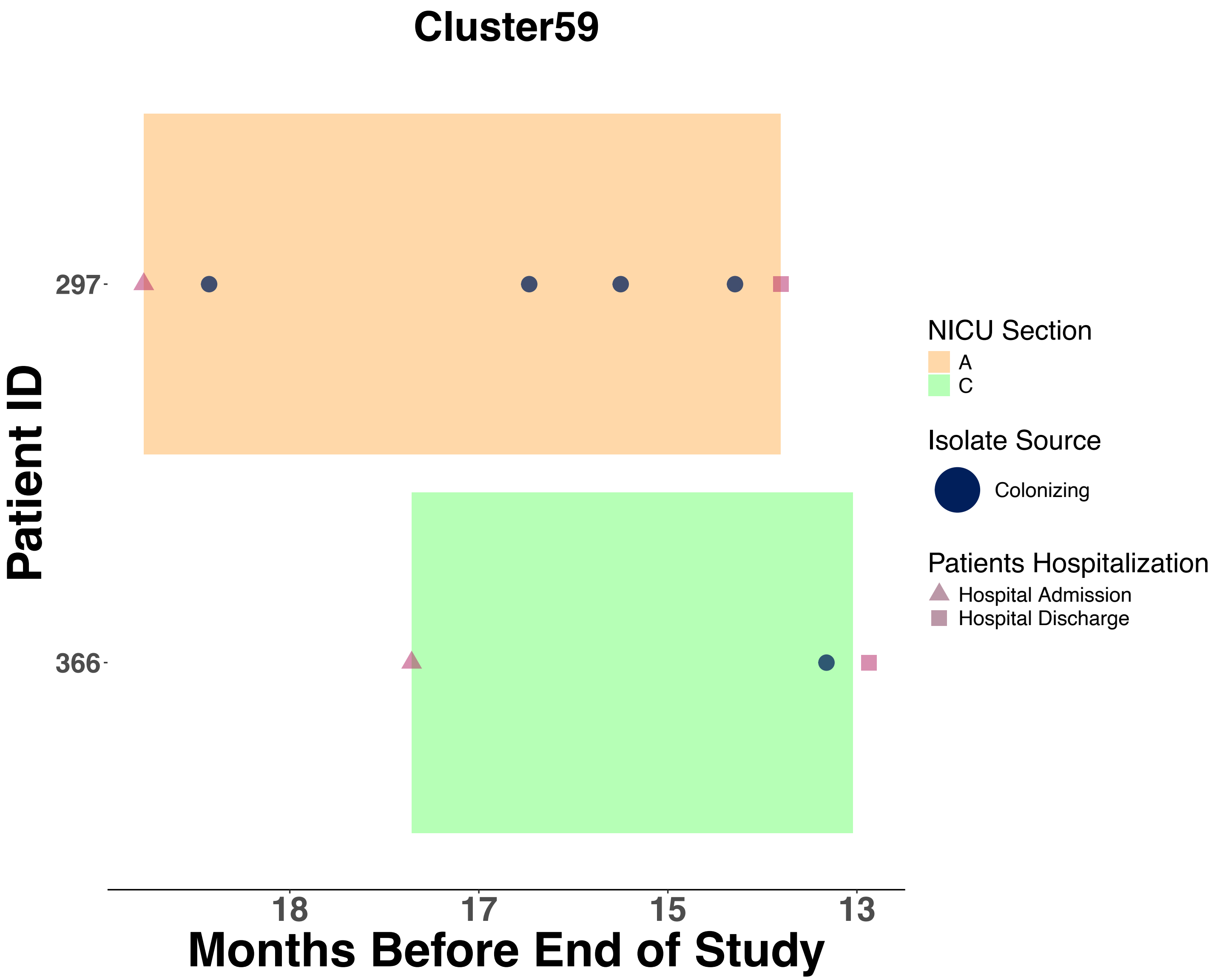

Cluster59
Patient ID
297
366
Months Before End of Study
18
17
15
13
NICU Section
A
C
Isolate Source
Colonizing
Patients Hospitalization
Hospital Admission
Hospital Discharge

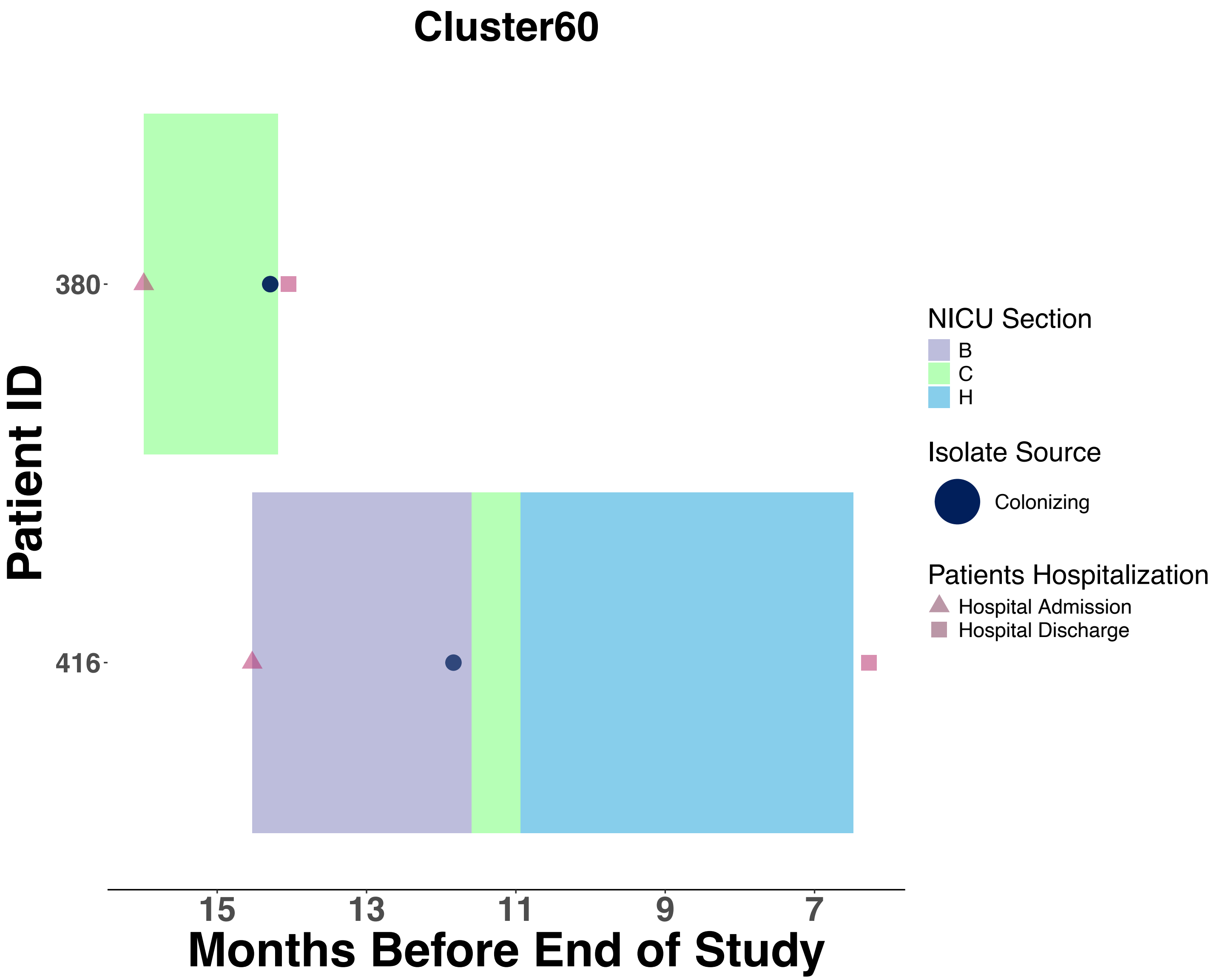

Cluster60
Patient ID
380
416
15
13
11
9
7
Months Before End of Study
NICU Section
B
C
H
Isolate Source
Colonizing
Patients Hospitalization
Hospital Admission
Hospital Discharge

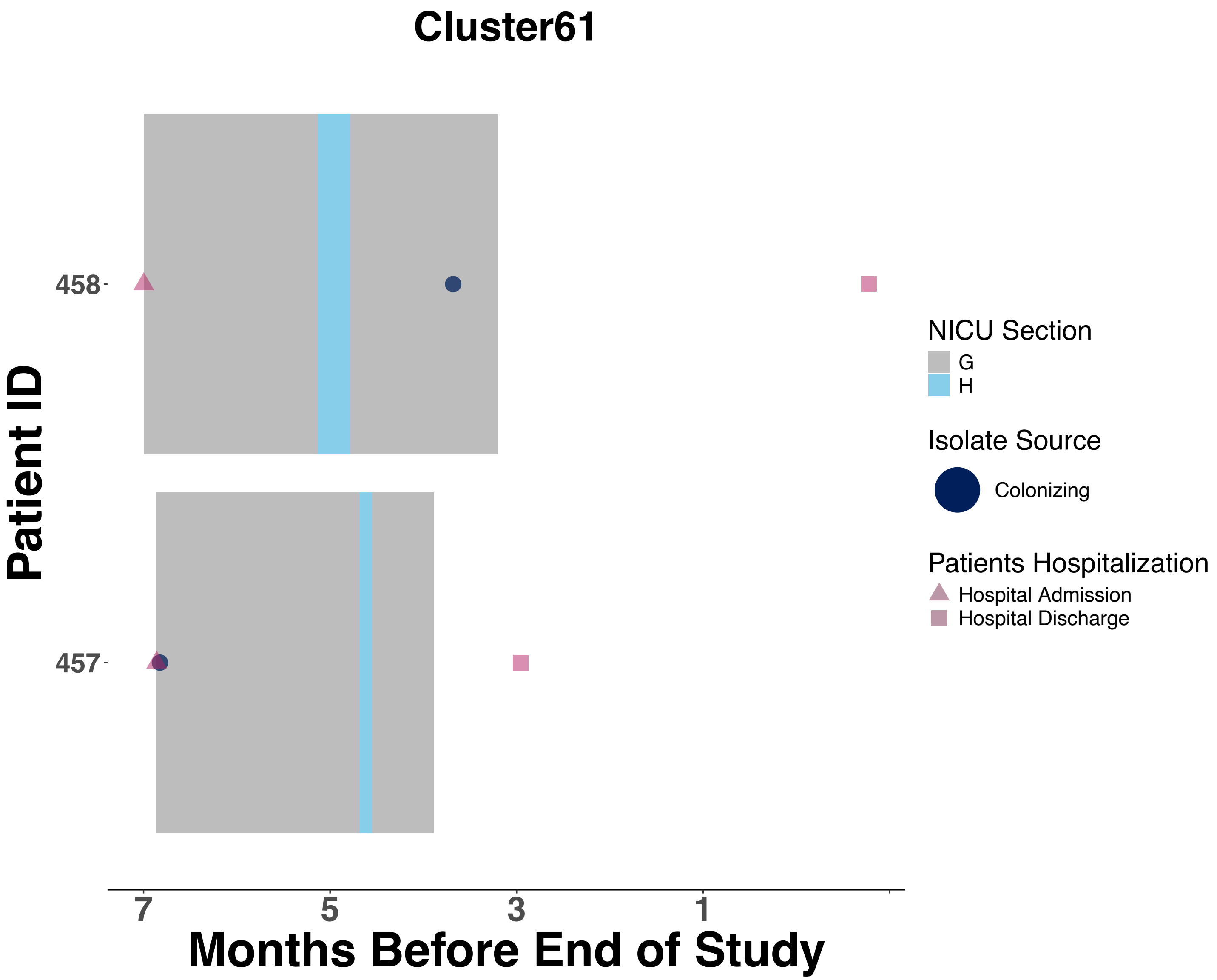

Cluster61
Patient ID
458
457
7
5
3
1
Months Before End of Study
NICU Section
G
H
Isolate Source
Colonizing
Patients Hospitalization
Hospital Admission
Hospital Discharge

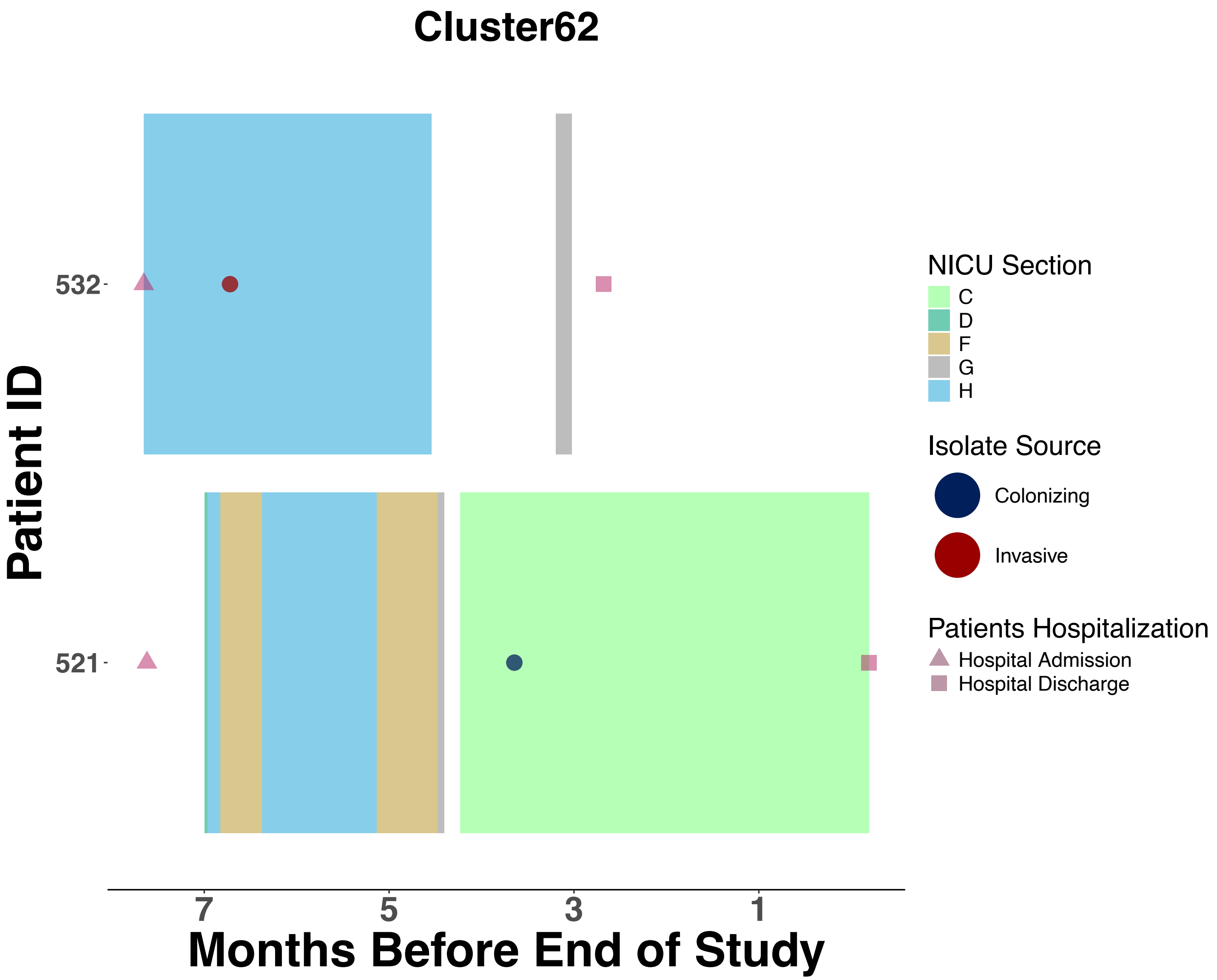
Cluster62
Patient ID
532
521
7
5
3
1
Months Before End of Study
NICU Section
C
D
F
G
H
Isolate Source
Colonizing
Invasive
Patients Hospitalization
Hospital Admission
Hospital Discharge

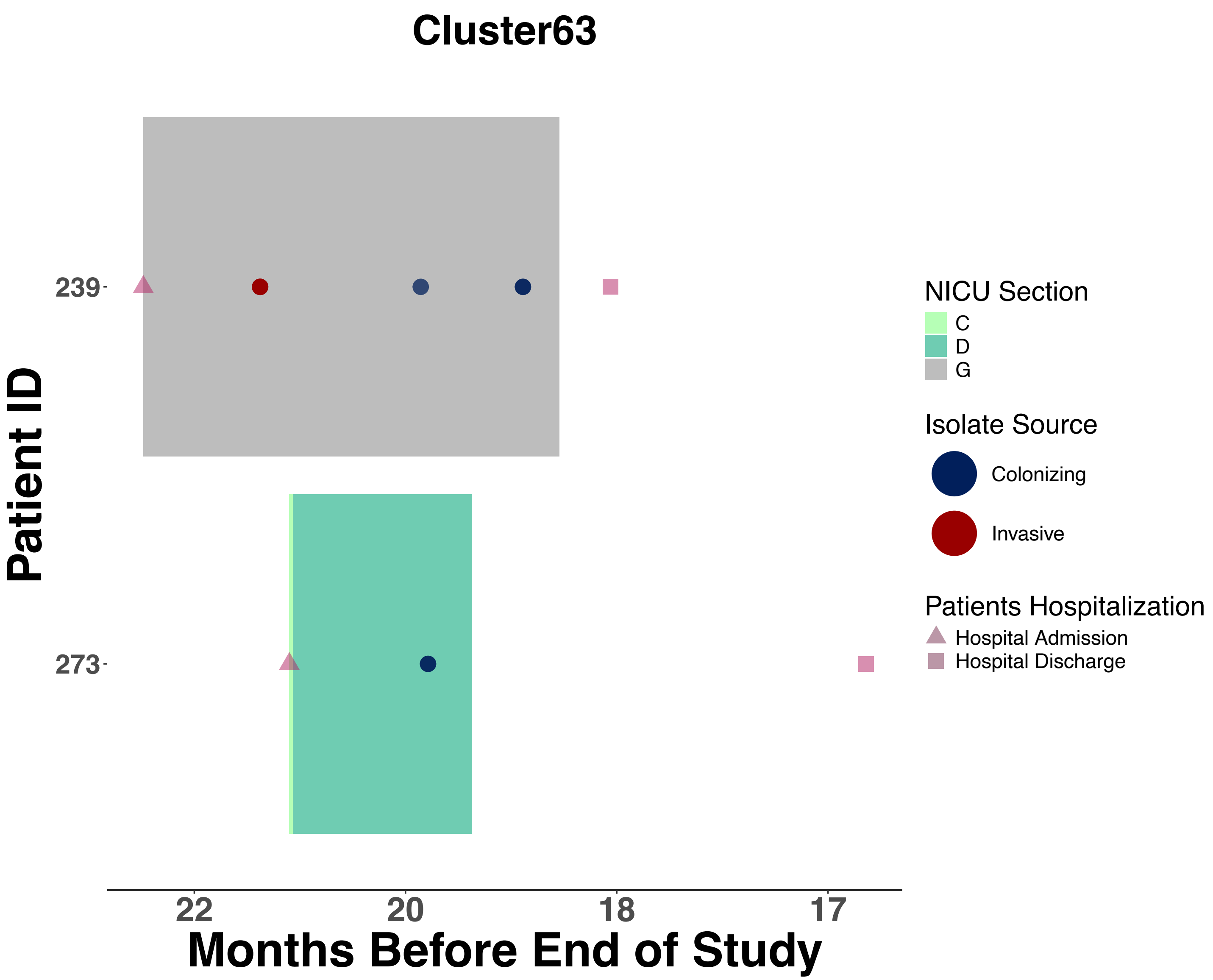

Cluster63
Patient ID
239
273
22
20
18
17
Months Before End of Study
NICU Section
C
D
G
Isolate Source
Colonizing
Invasive
Patients Hospitalization
Hospital Admission
Hospital Discharge

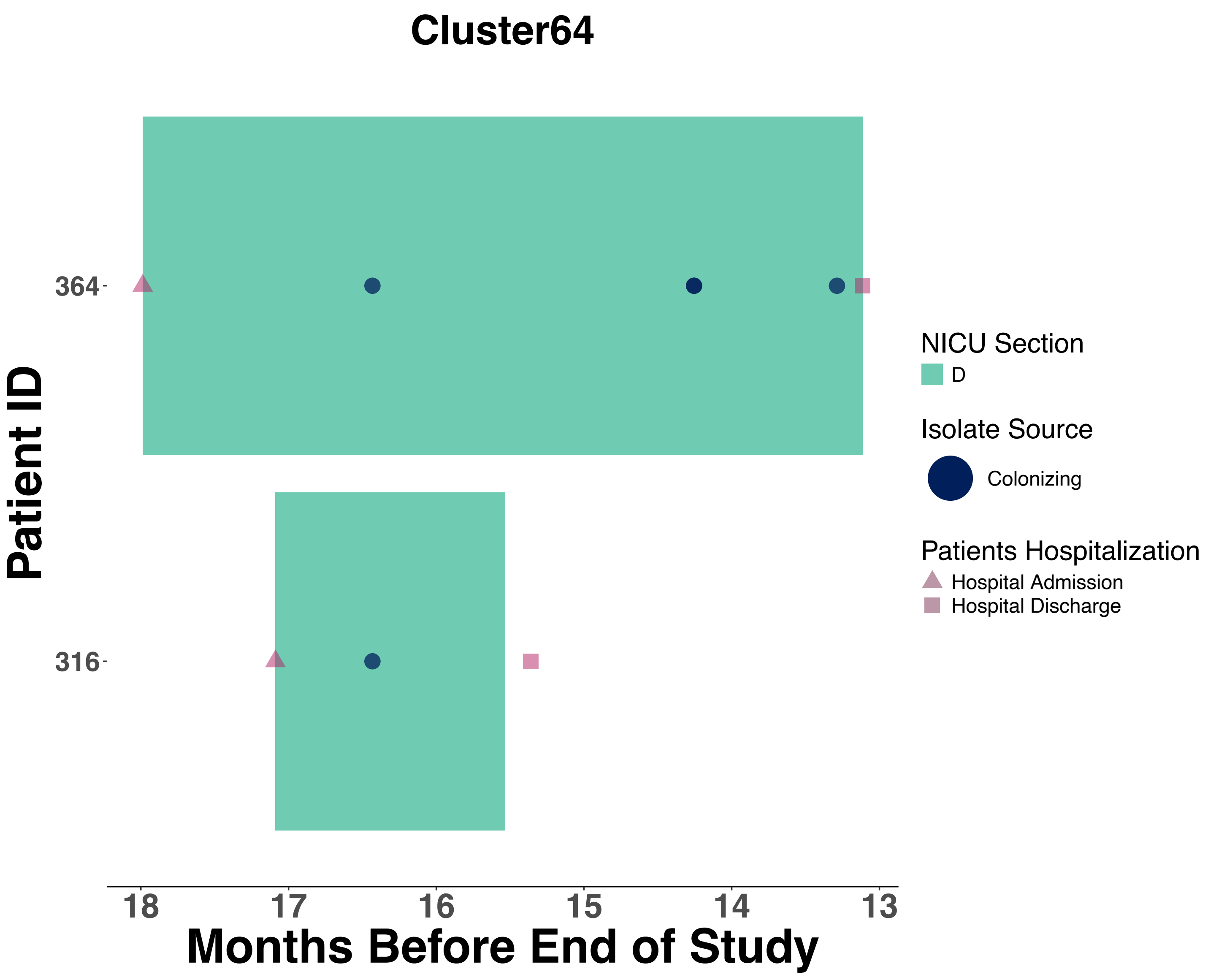

Cluster64
Patient ID
364
316
18
17
16
15
14
13
Months Before End of Study
NICU Section
D
Isolate Source
Colonizing
Patients Hospitalization
Hospital Admission
Hospital Discharge

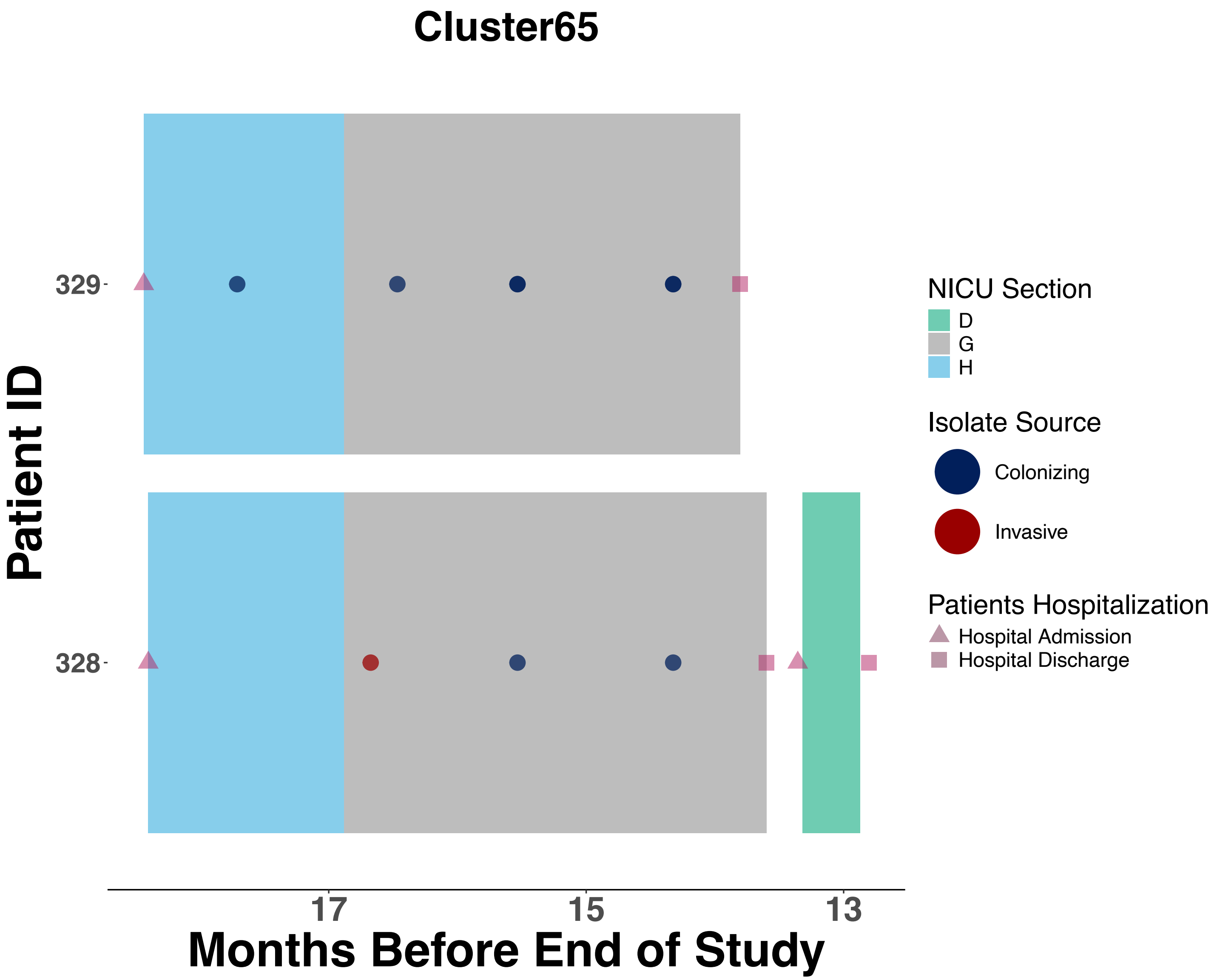

Cluster65
Patient ID
329
328
17
15
13
Months Before End of Study
NICU Section
D
G
H
Isolate Source
Colonizing
Invasive
Patients Hospitalization
Hospital Admission
Hospital Discharge

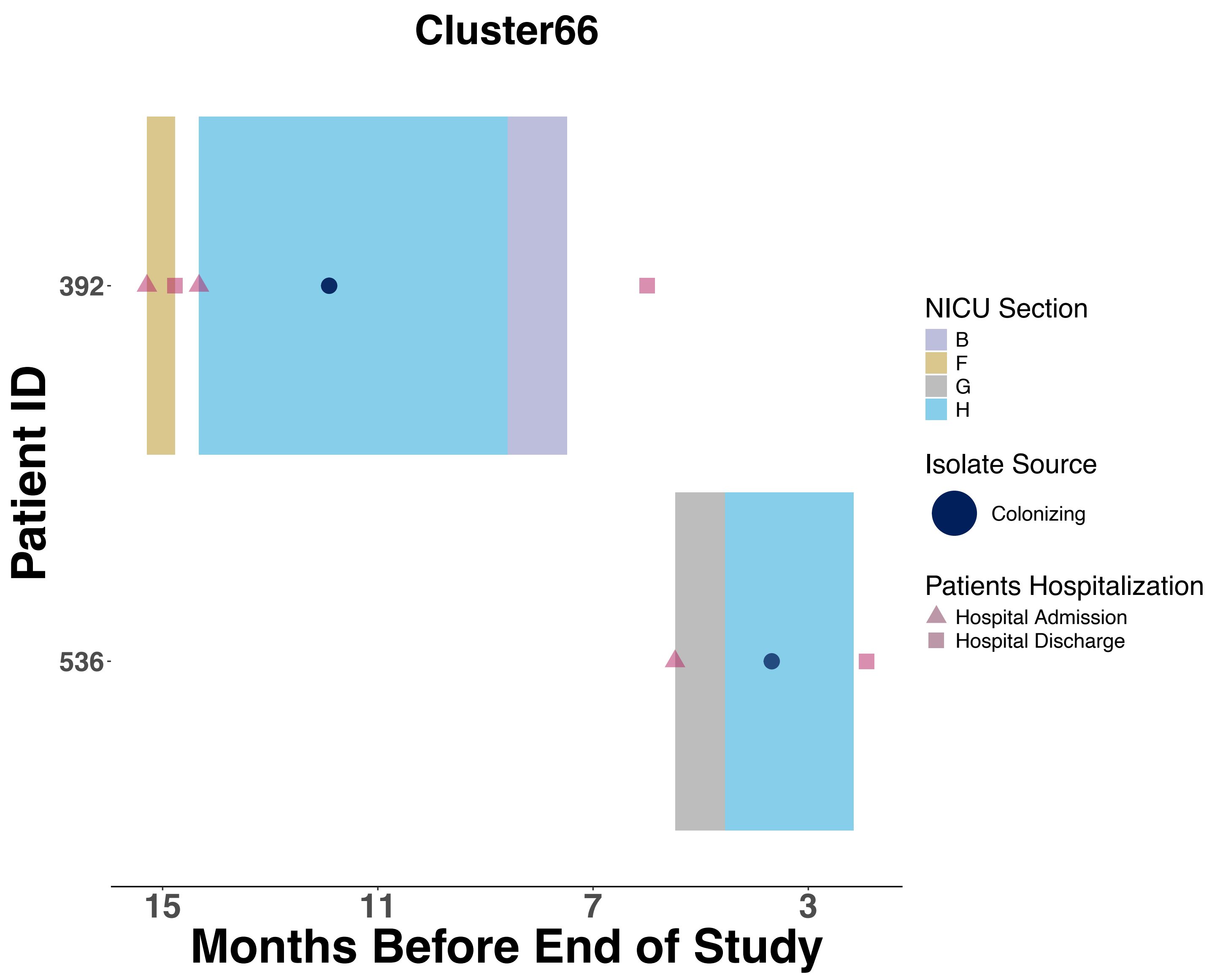

Cluster66
Patient ID
392
536
15
11
7
3
Months Before End of Study
NICU Section
B
F
G
H
Isolate Source
Colonizing
Patients Hospitalization
Hospital Admission
Hospital Discharge

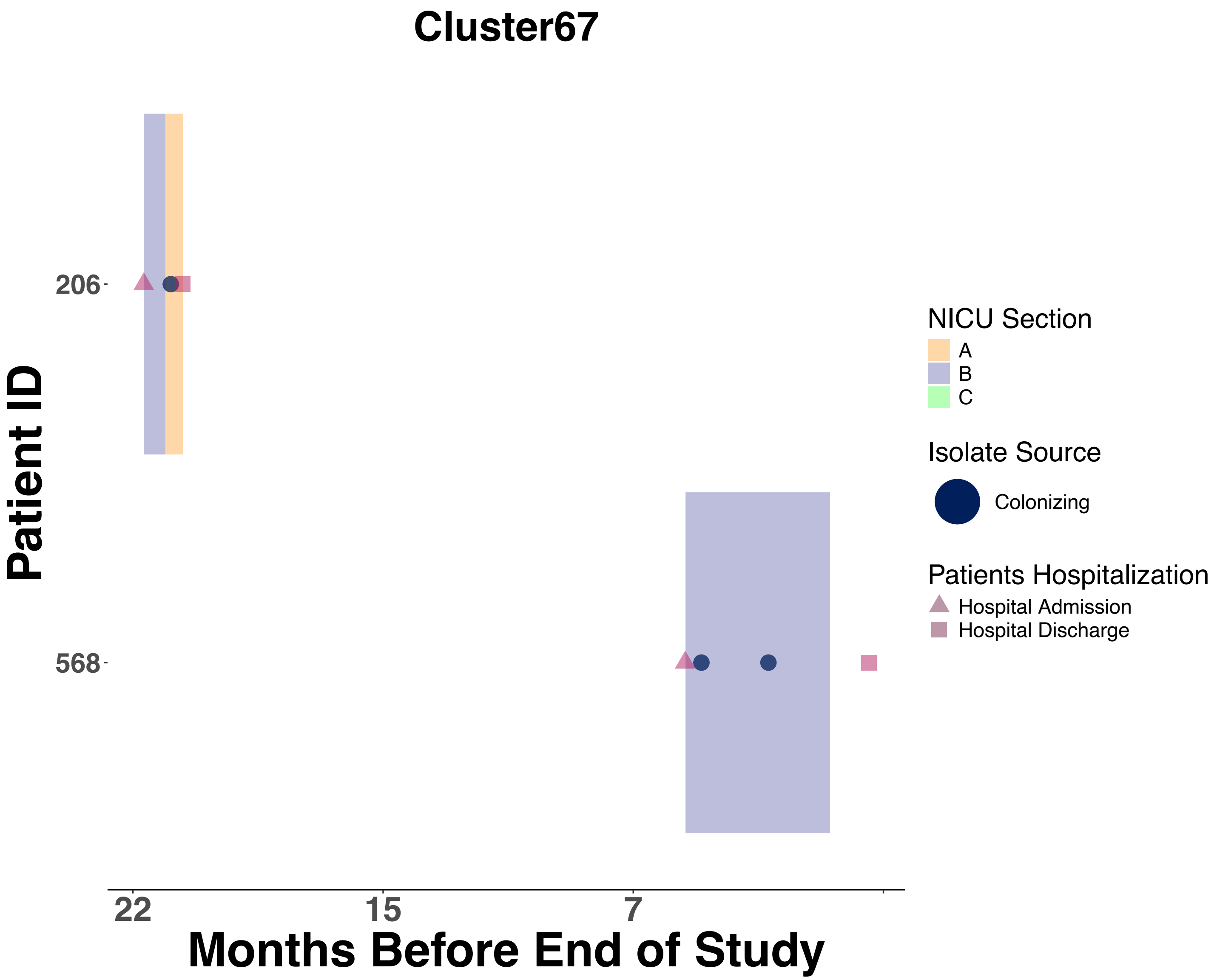
Cluster67
Patient ID
206
568
22
15
7
Months Before End of Study
NICU Section
A
B
C
Isolate Source
Colonizing
Patients Hospitalization
Hospital Admission
Hospital Discharge

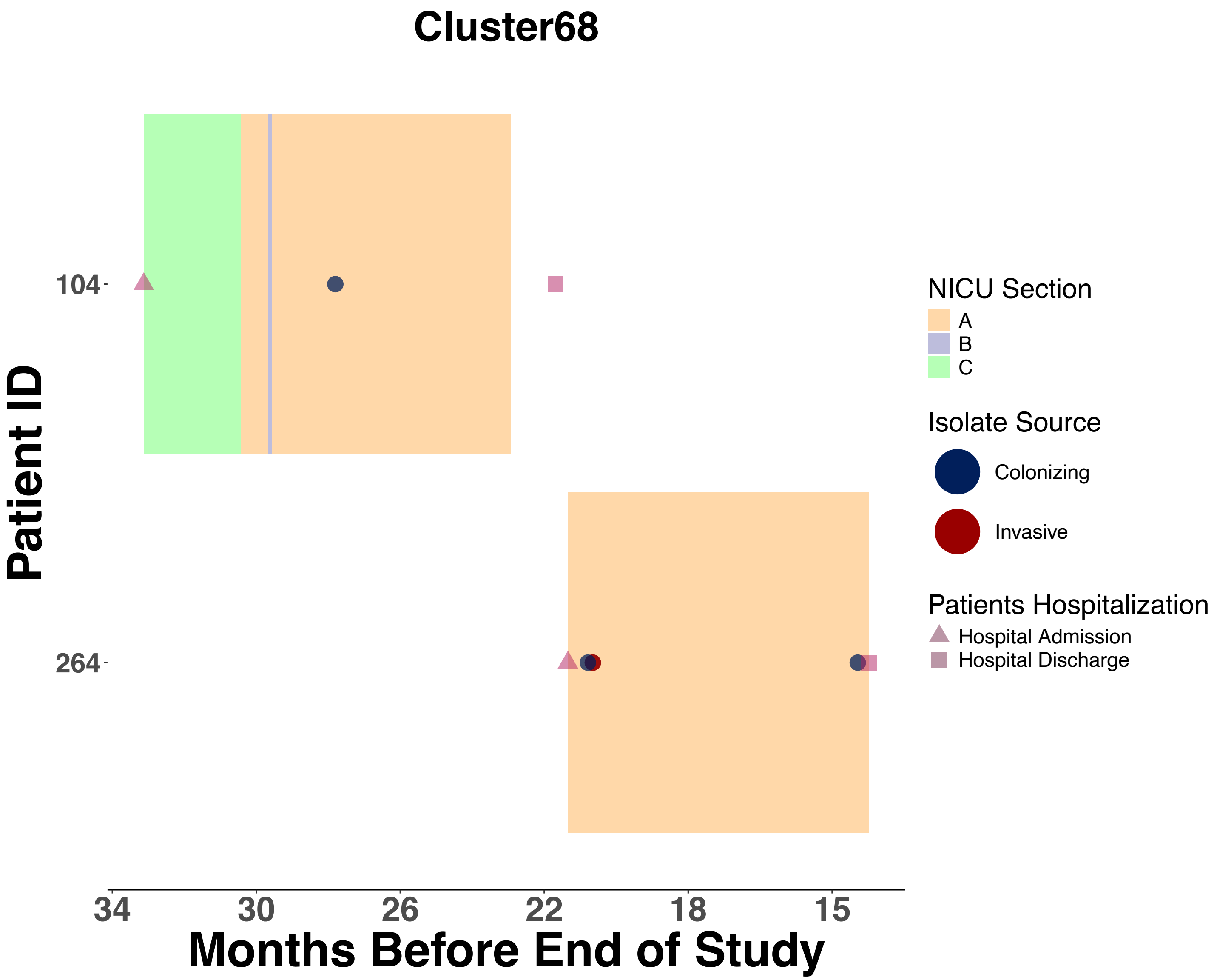
Cluster68
Patient ID
104
264
34
30
26
22
18
15
Months Before End of Study
NICU Section
A
B
C
Isolate Source
Colonizing
Invasive
Patients Hospitalization
Hospital Admission
Hospital Discharge

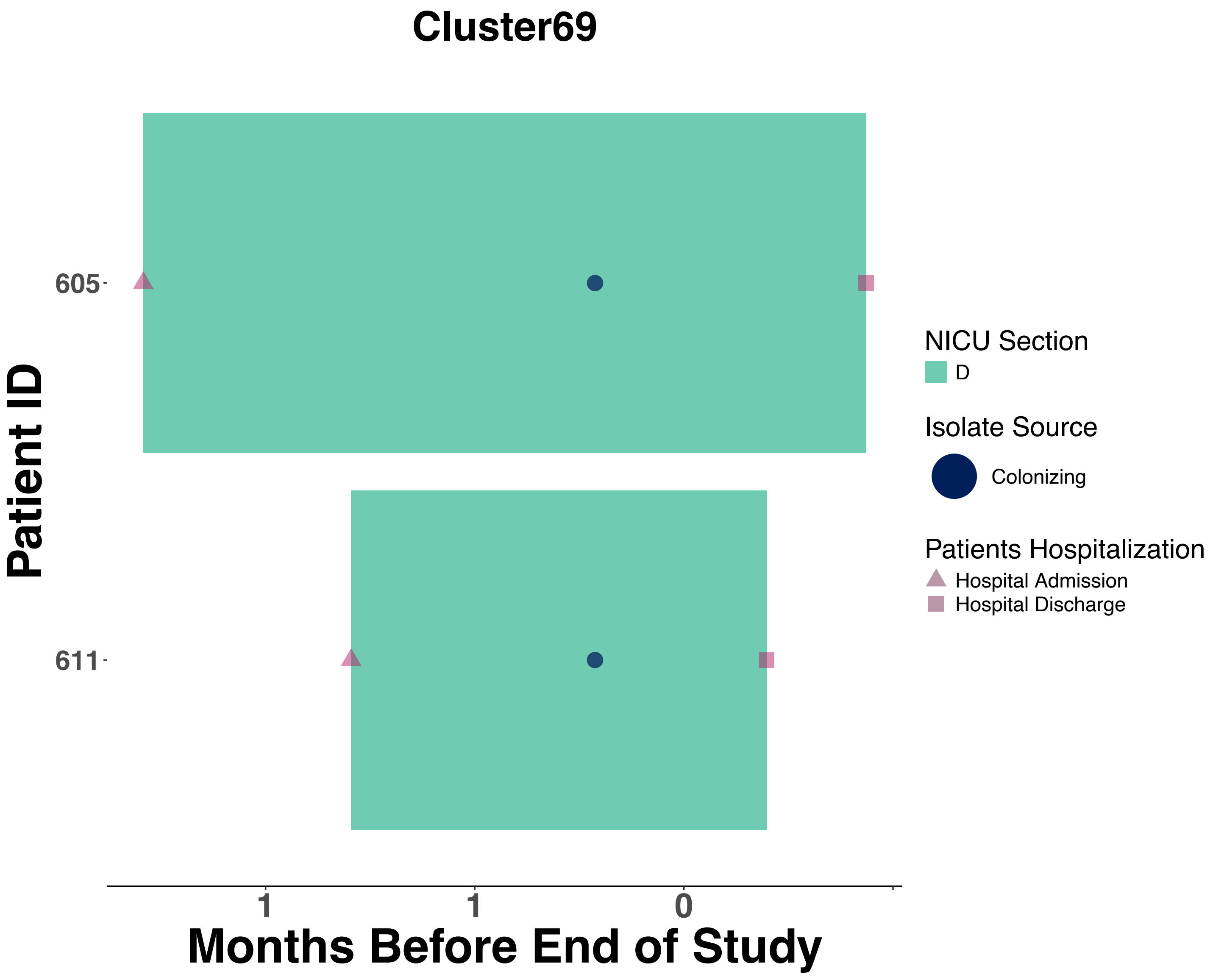
Cluster69
Patient ID
605
611
1
1
0
Months Before End of Study
NICU Section
D
Isolate Source
Colonizing
Patients Hospitalization
Hospital Admission
Hospital Discharge

Figure S5:  Temporal analysis for each identified transmission cluster by treatment team assignments

Each horizontal bar represents a single patient's NICU stay, with colored rectangles indicating corresponding treatment team assignments over time. Admission dates are marked with triangles and discharge dates with squares. X-axis (Months Before End of Study) represents the number of months prior to completion of study period.

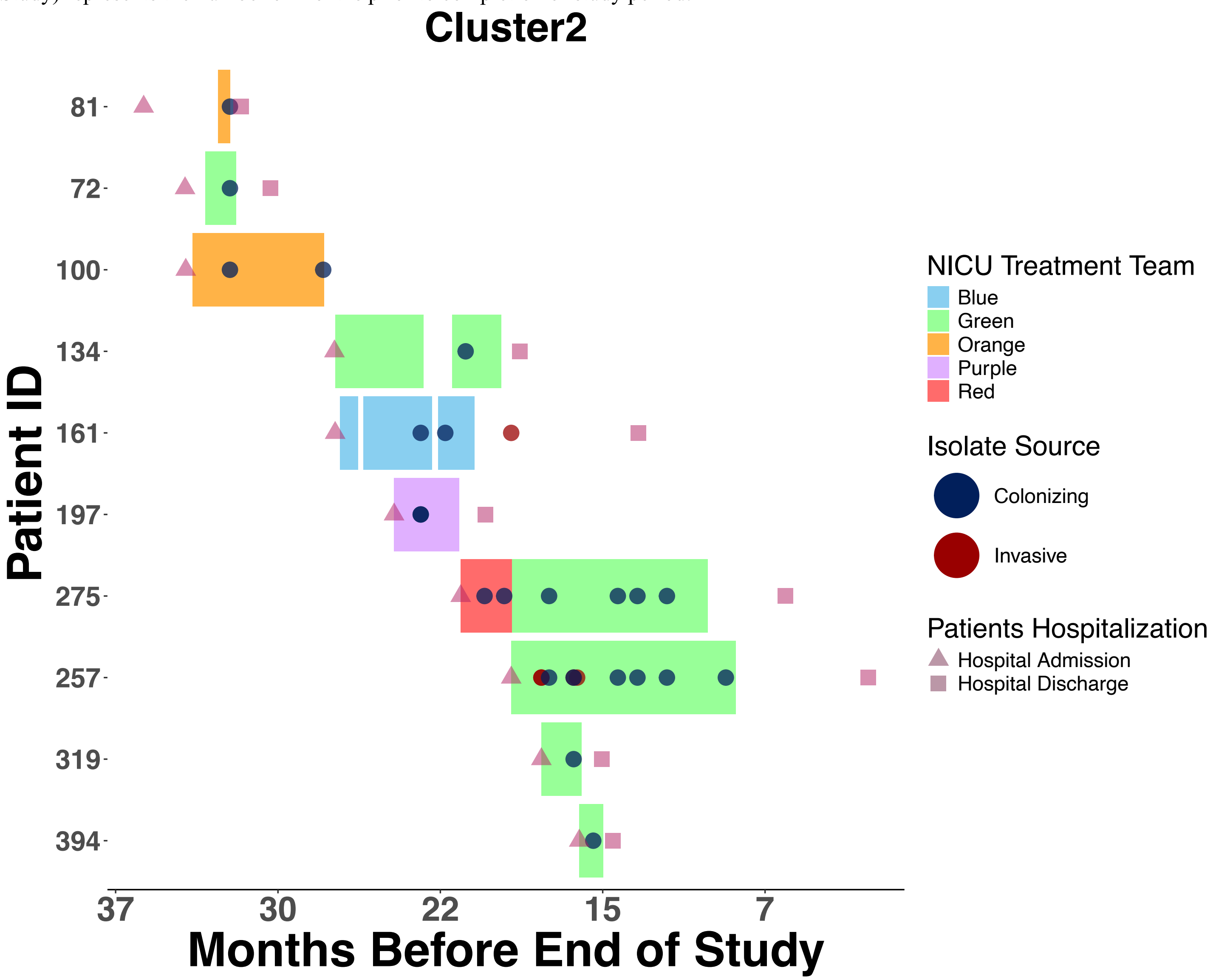

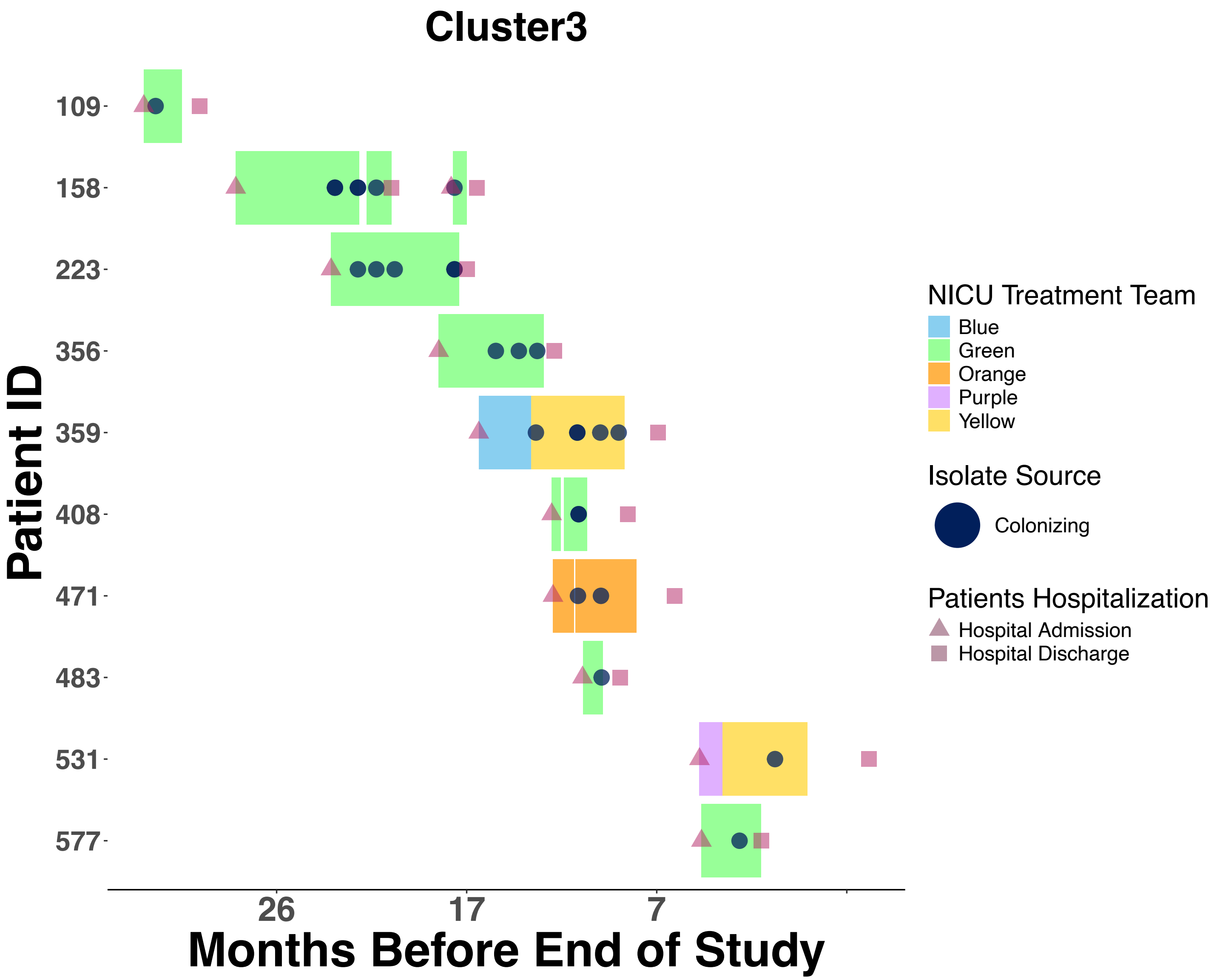

Cluster3
Patient ID
109
158
223
356
359
408
471
483
531
577
26
17
7
Months Before End of Study
NICU Treatment Team
Blue
Green
Orange
Purple
Yellow
Isolate Source
Colonizing
Patients Hospitalization
Hospital Admission
Hospital Discharge

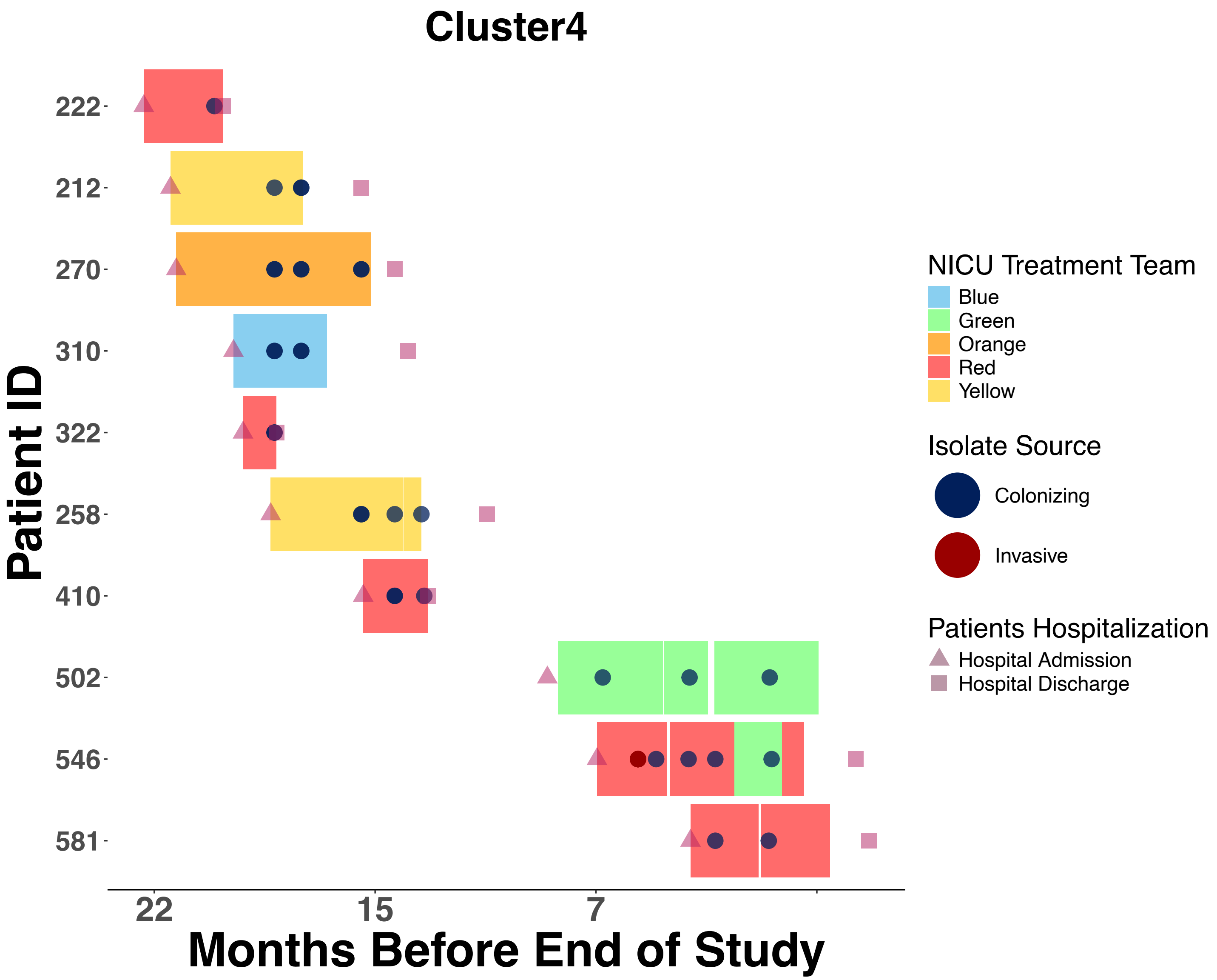

Cluster4
Patient ID
222
212
270
310
322
258
410
502
546
581
22
15
7
Months Before End of Study
NICU Treatment Team
Blue
Green
Orange
Red
Yellow
Isolate Source
Colonizing
Invasive
Patients Hospitalization
Hospital Admission
Hospital Discharge

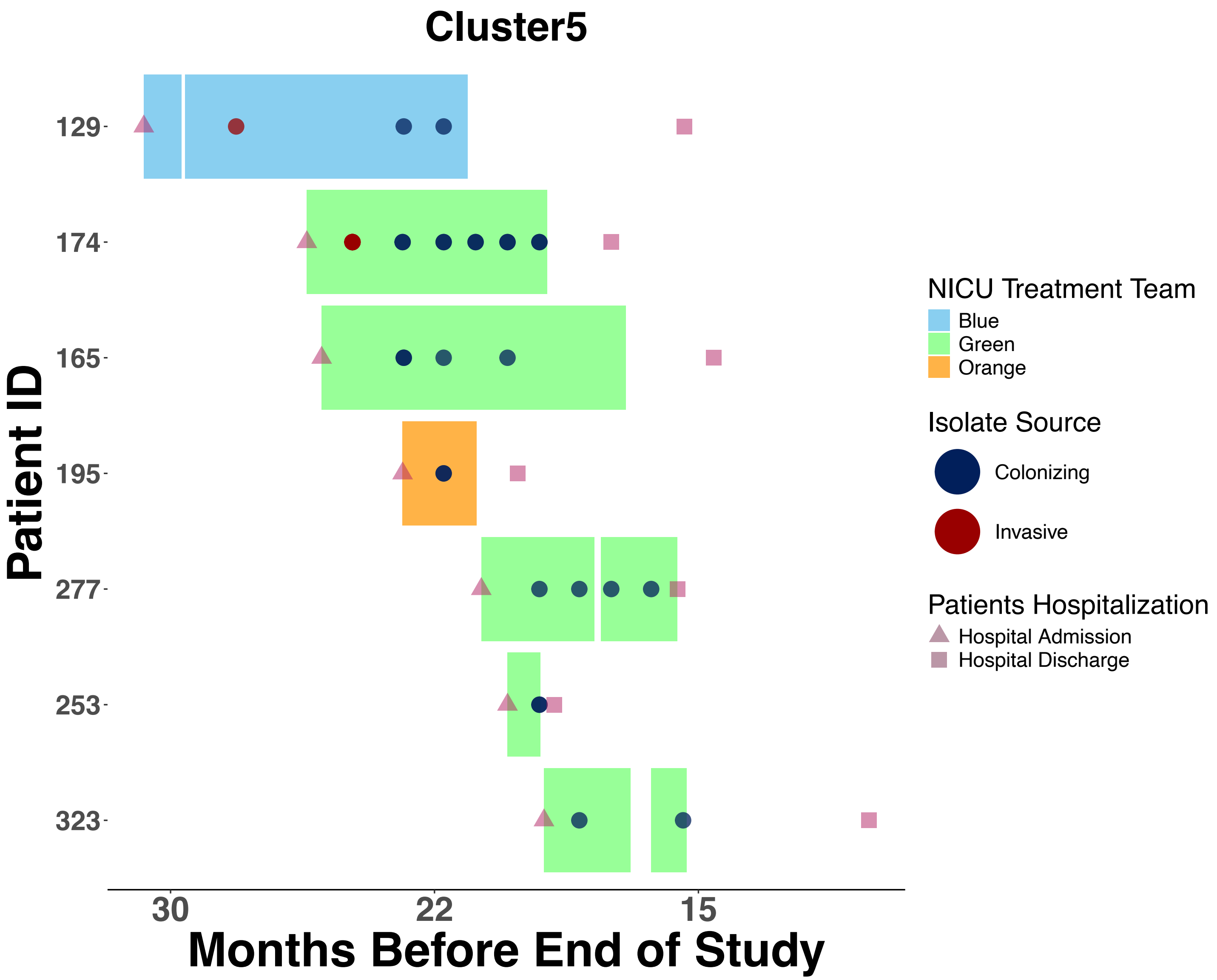
Cluster5
Patient ID
129
174
165
195
277
253
323
30
22
15
Months Before End of Study
NICU Treatment Team
Blue
Green
Orange
Isolate Source
Colonizing
Invasive
Patients Hospitalization
Hospital Admission
Hospital Discharge

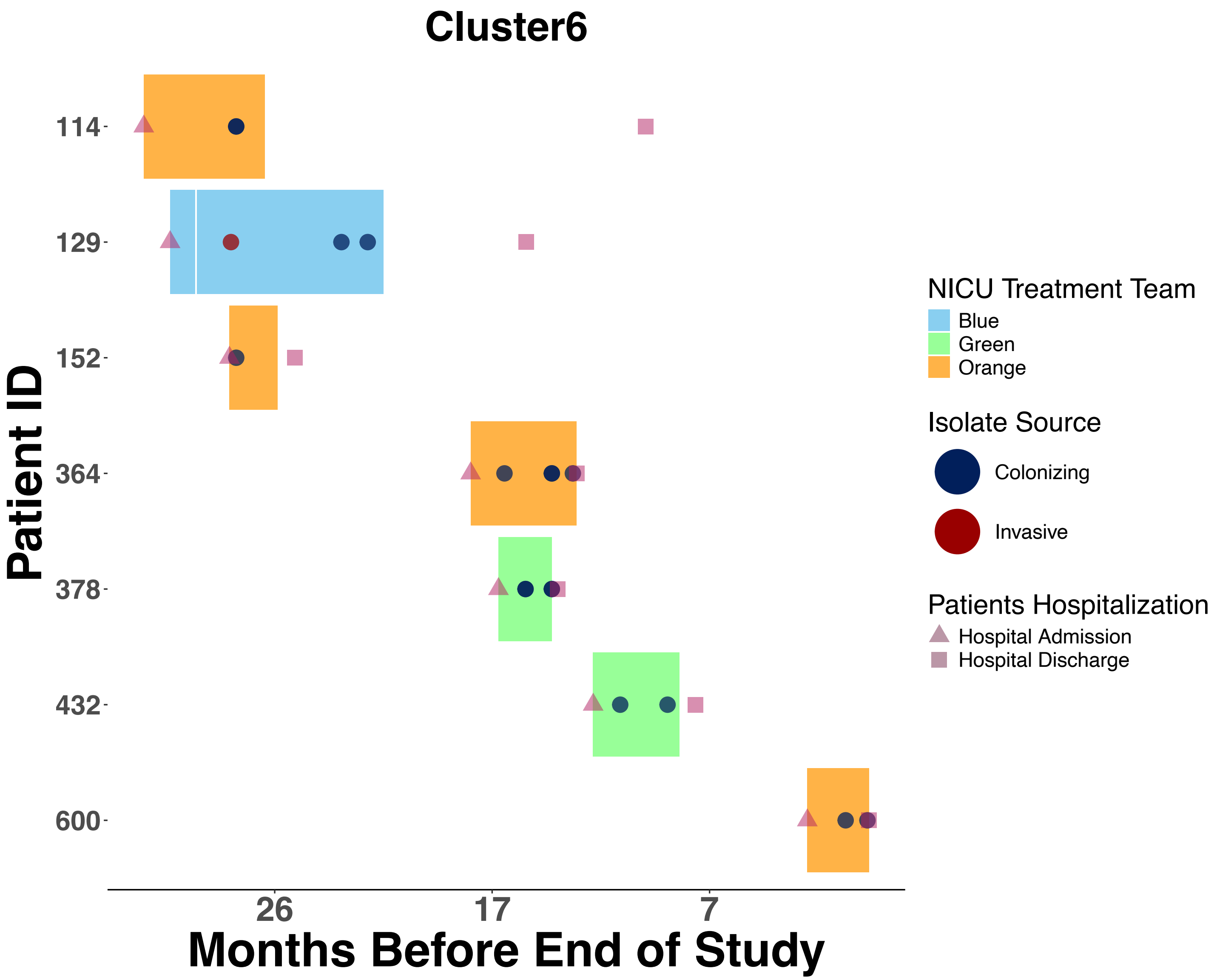
Cluster6
Patient ID
114
129
152
364
378
432
600
26
17
7
Months Before End of Study
NICU Treatment Team
Blue
Green
Orange
Isolate Source
Colonizing
Invasive
Patients Hospitalization
Hospital Admission
Hospital Discharge

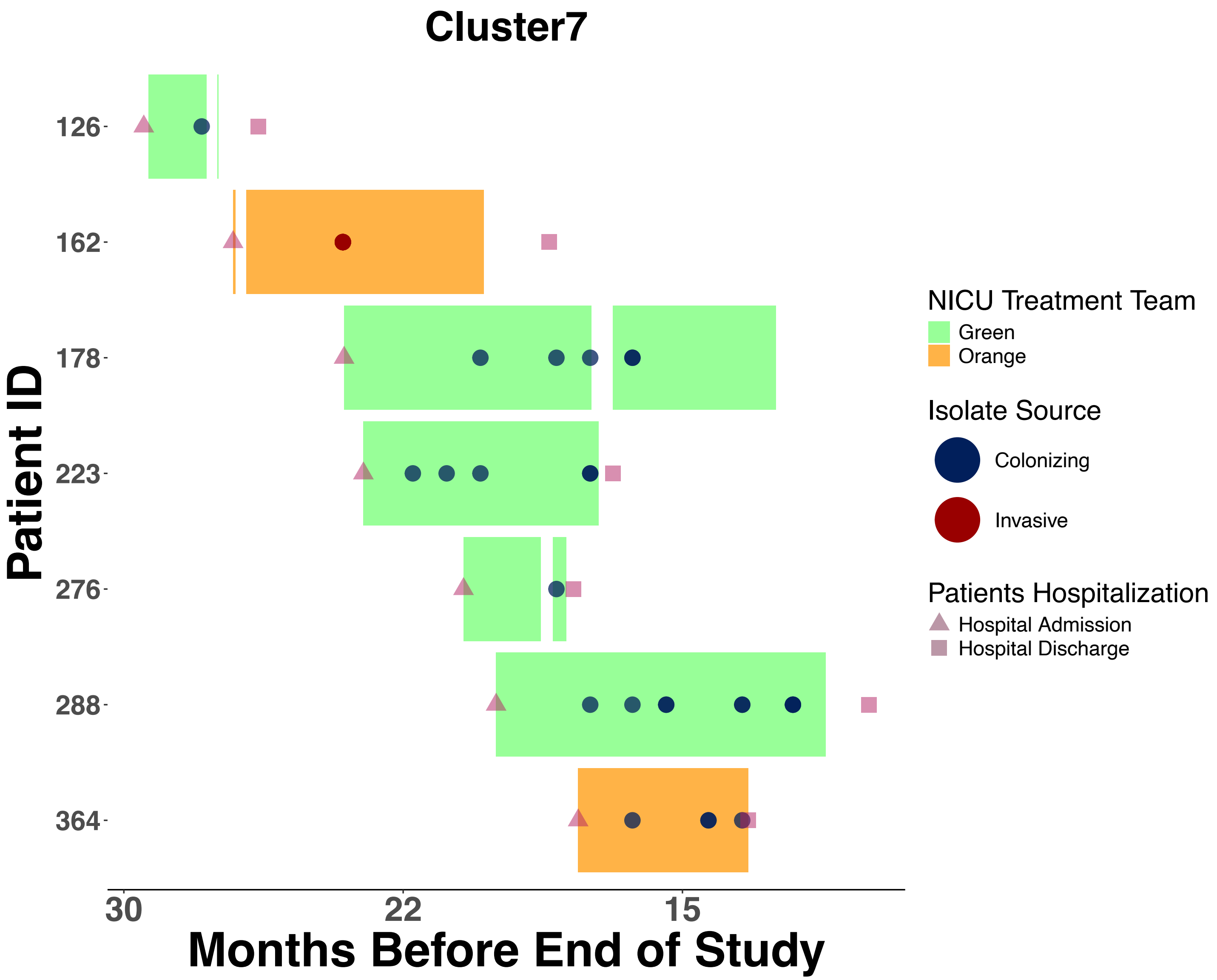

Cluster7
Patient ID
126
162
178
223
276
288
364
30
22
15
Months Before End of Study
NICU Treatment Team
Green
Orange
Isolate Source
Colonizing
Invasive
Patients Hospitalization
Hospital Admission
Hospital Discharge

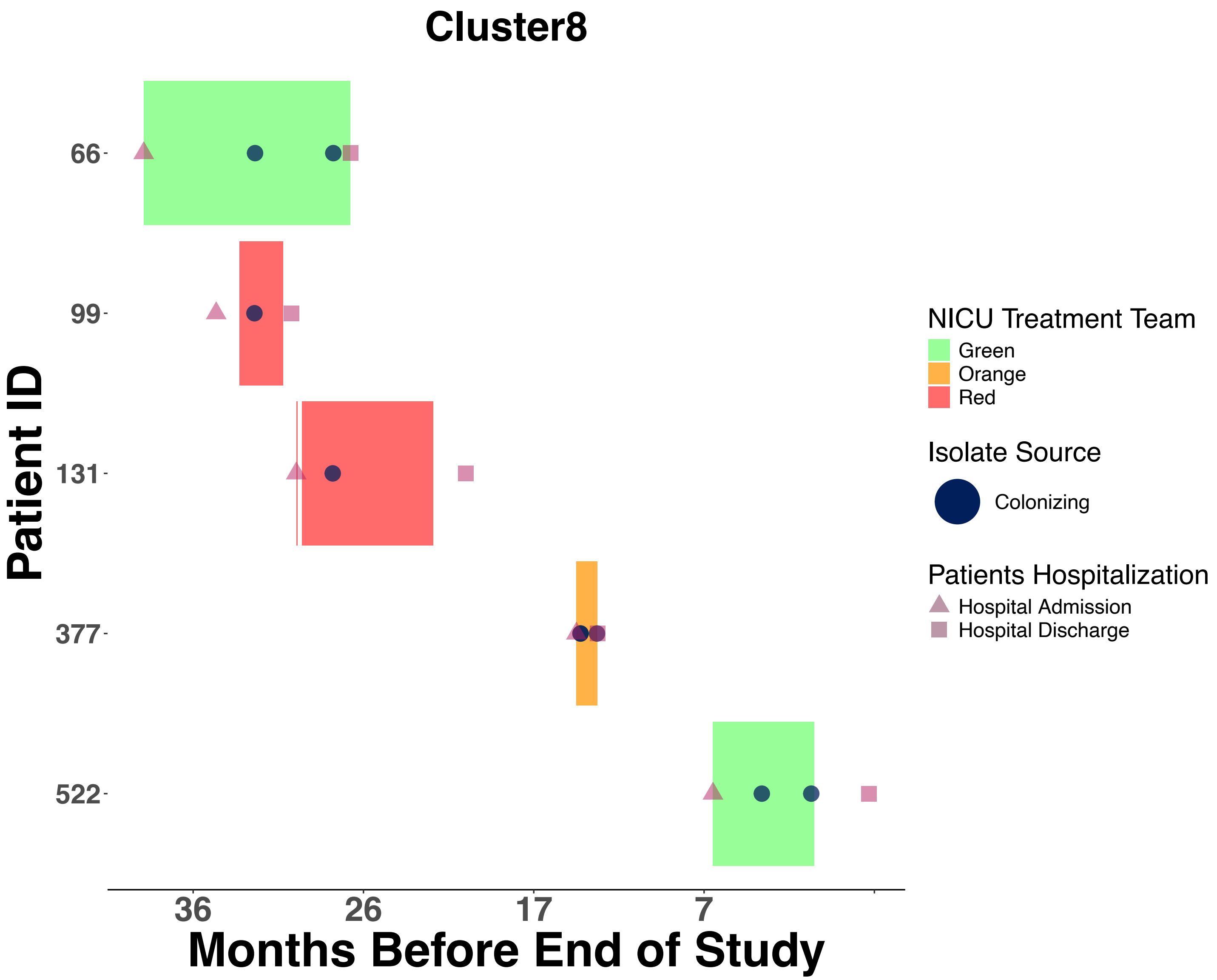

Cluster8
Patient ID
66
99
131
377
522
36
26
17
7
Months Before End of Study
NICU Treatment Team
Green
Orange
Red
Isolate Source
Colonizing
Patients Hospitalization
Hospital Admission
Hospital Discharge

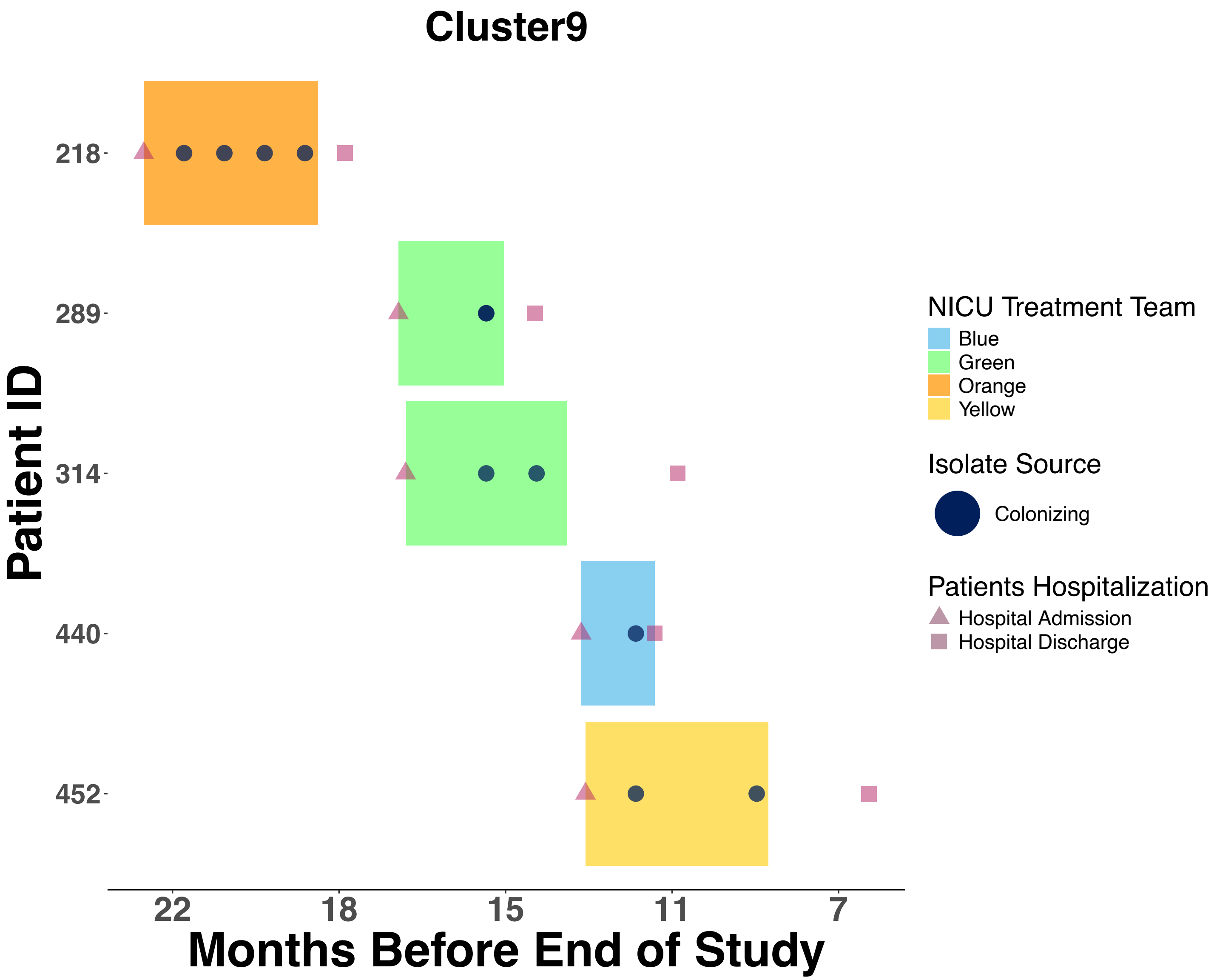
Cluster9
Patient ID
218
289
314
440
452
22
18
15
11
7
Months Before End of Study
NICU Treatment Team
Blue
Green
Orange
Yellow
Isolate Source
Colonizing
Patients Hospitalization
Hospital Admission
Hospital Discharge

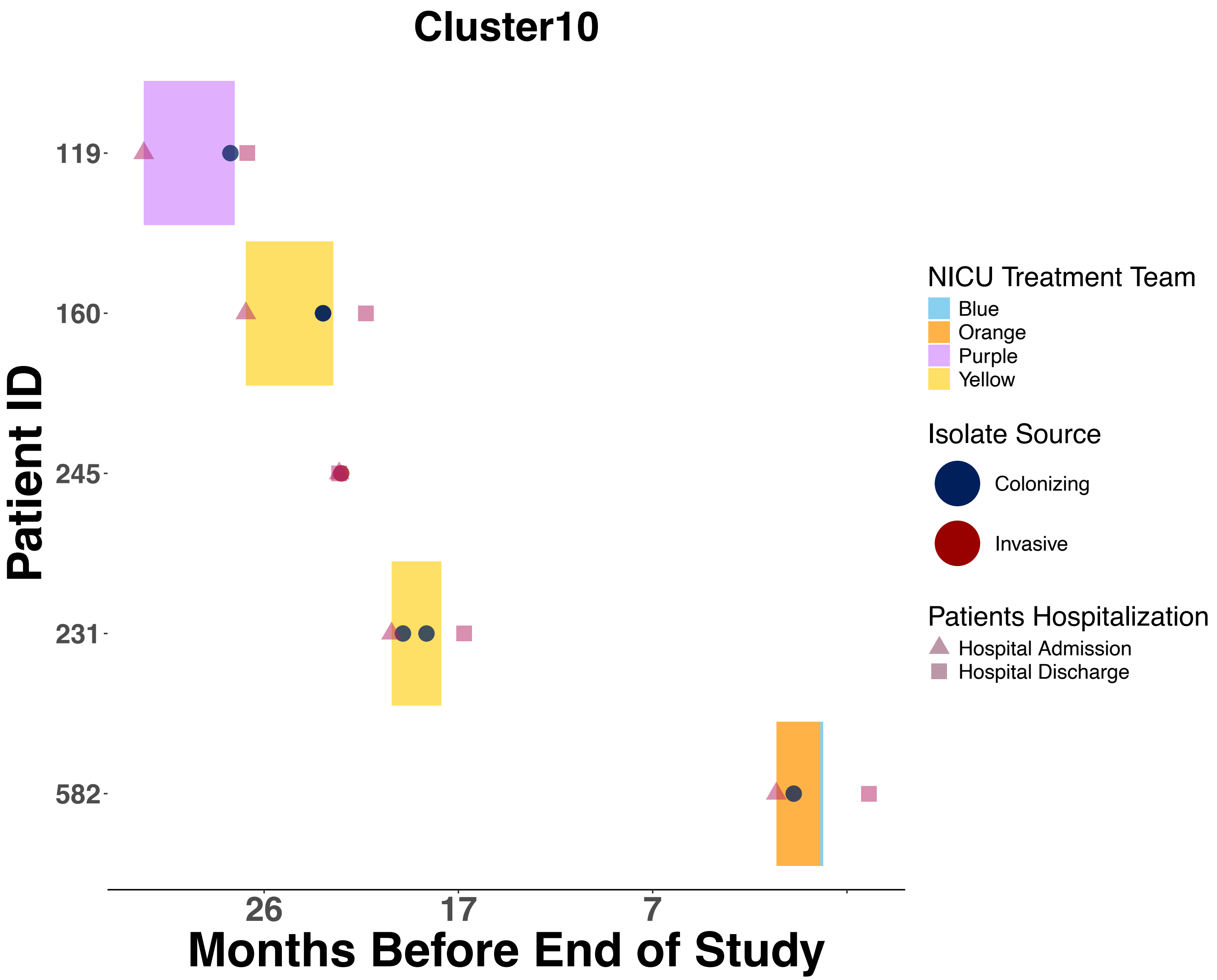
Cluster10
Patient ID
119
160
245
231
582
26
17
7
Months Before End of Study
NICU Treatment Team
Blue
Orange
Purple
Yellow
Isolate Source
Colonizing
Invasive
Patients Hospitalization
Hospital Admission
Hospital Discharge

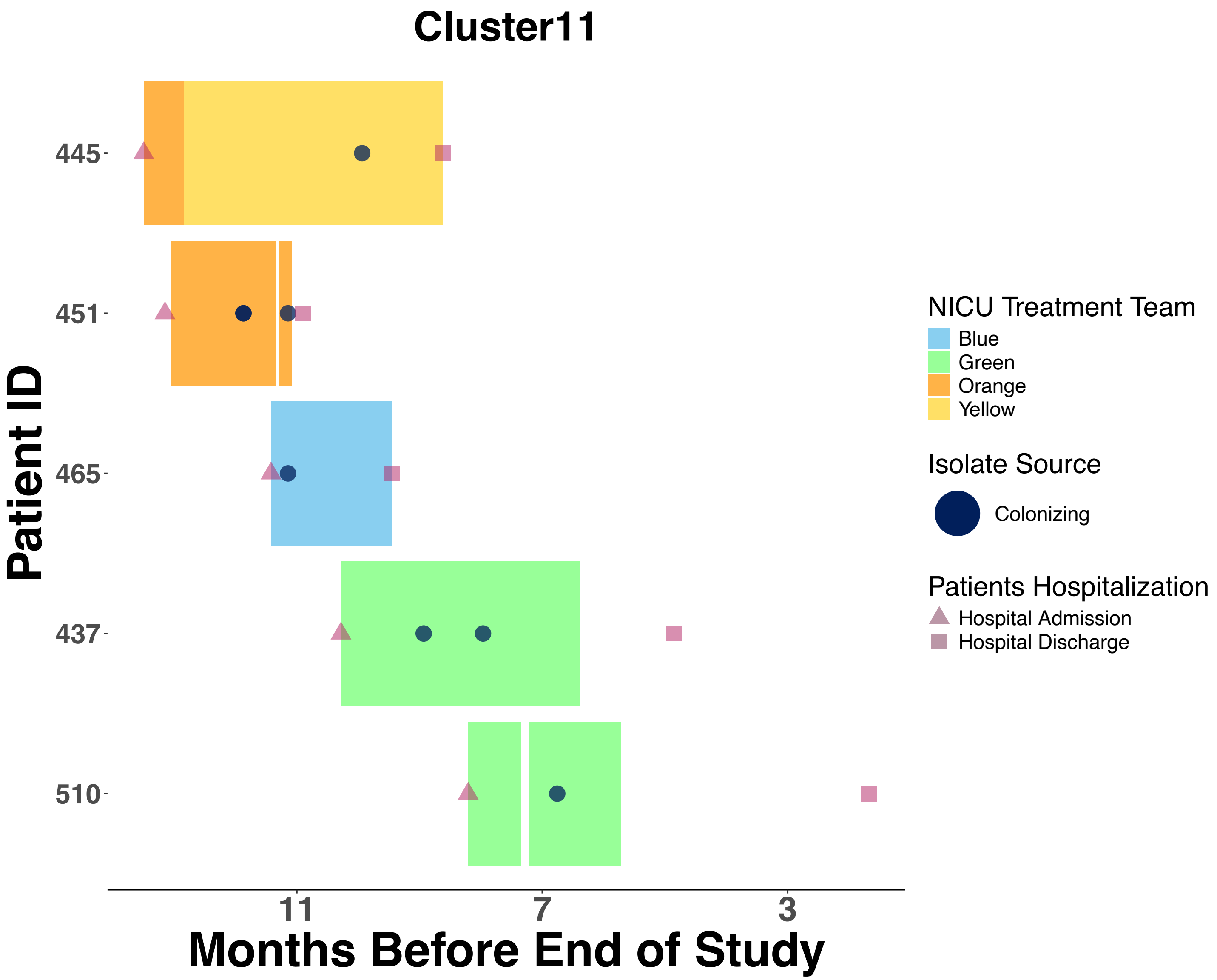

Cluster11
Patient ID
445
451
465
437
510
11
7
3
Months Before End of Study
NICU Treatment Team
Blue
Green
Orange
Yellow
Isolate Source
Colonizing
Patients Hospitalization
Hospital Admission
Hospital Discharge

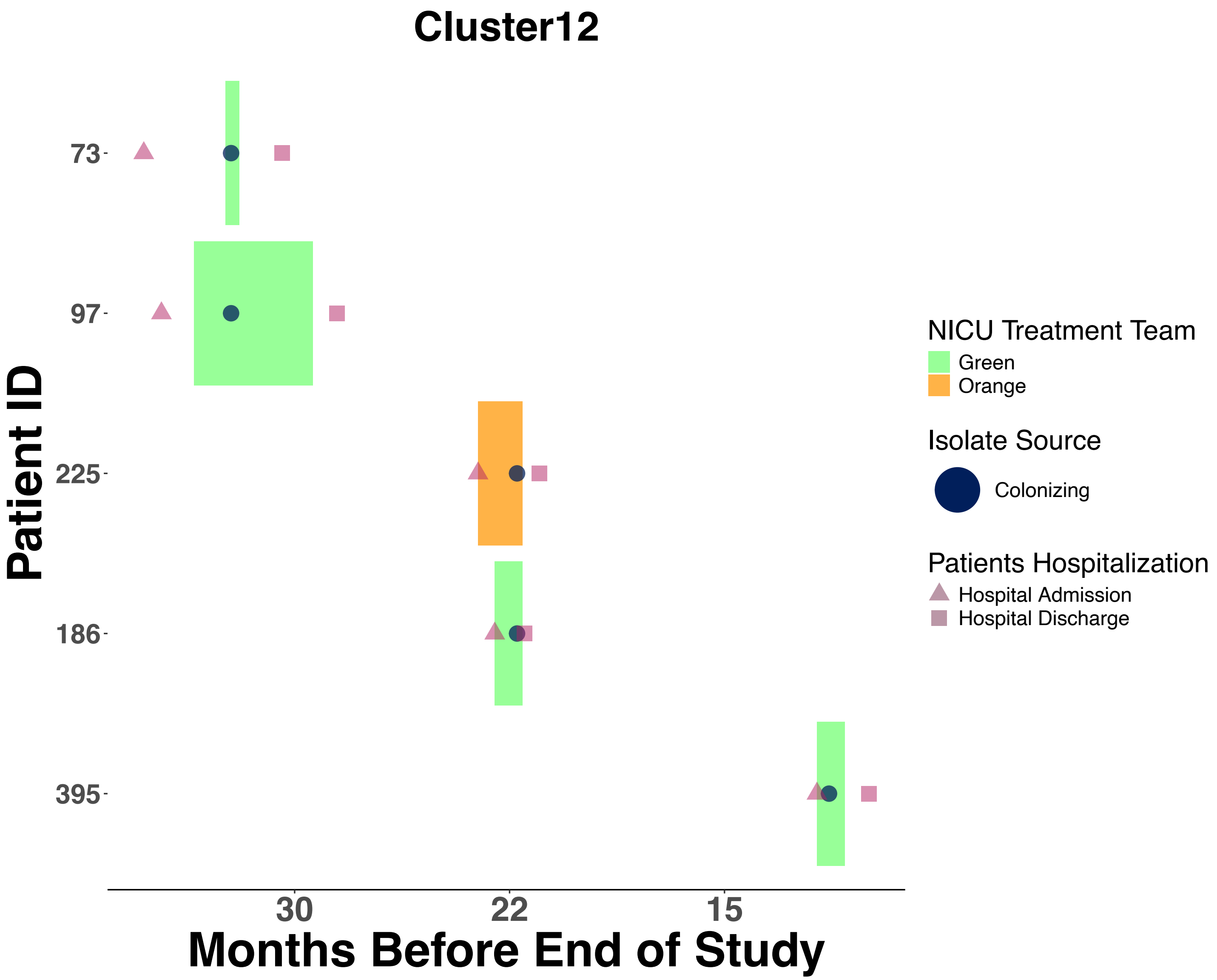
Cluster12
Patient ID
73
97
225
186
395
30
22
15
Months Before End of Study
NICU Treatment Team
Green
Orange
Isolate Source
Colonizing
Patients Hospitalization
Hospital Admission
Hospital Discharge

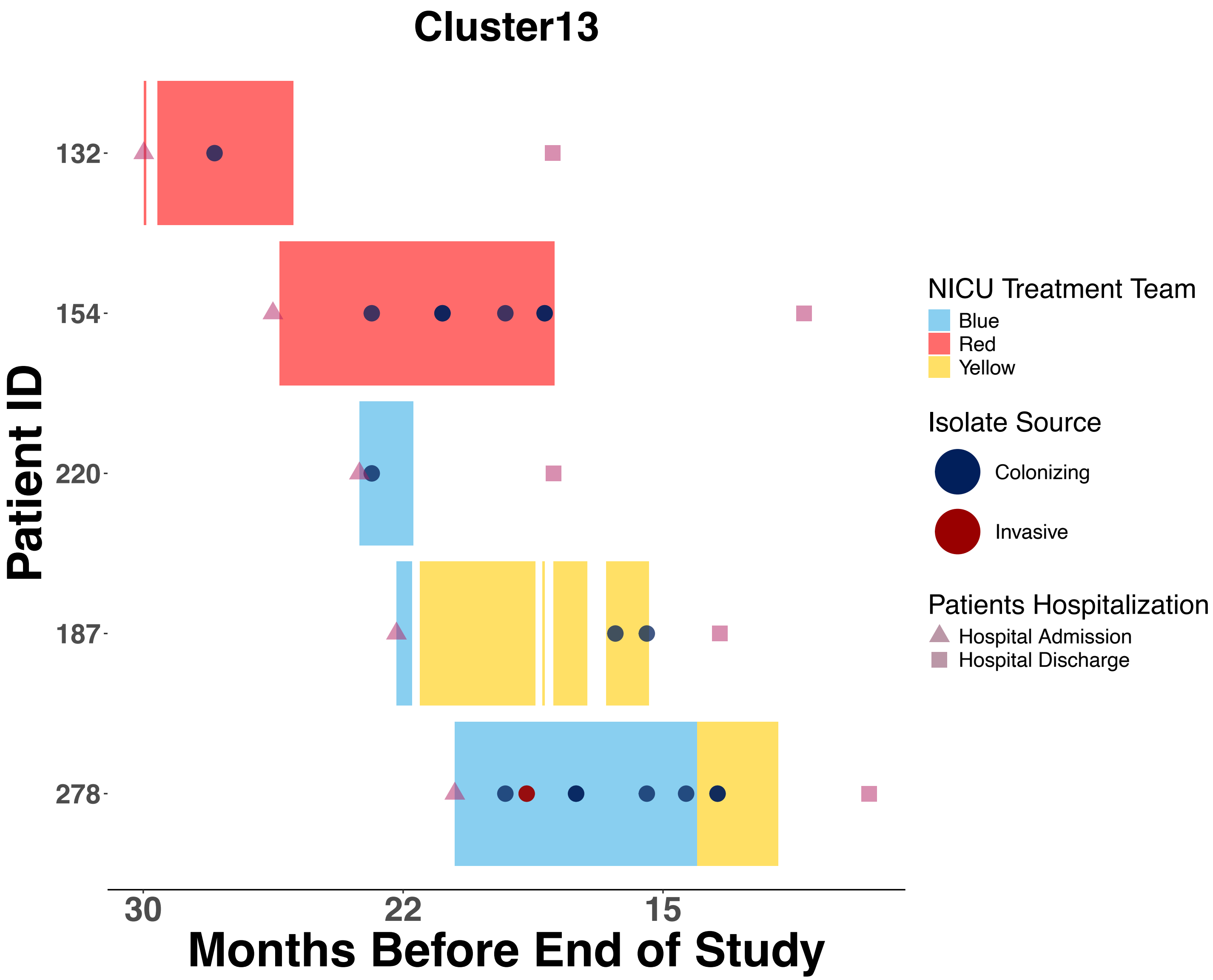

Cluster13
Patient ID
132
154
220
187
278
30
22
15
Months Before End of Study
NICU Treatment Team
Blue
Red
Yellow
Isolate Source
Colonizing
Invasive
Patients Hospitalization
Hospital Admission
Hospital Discharge

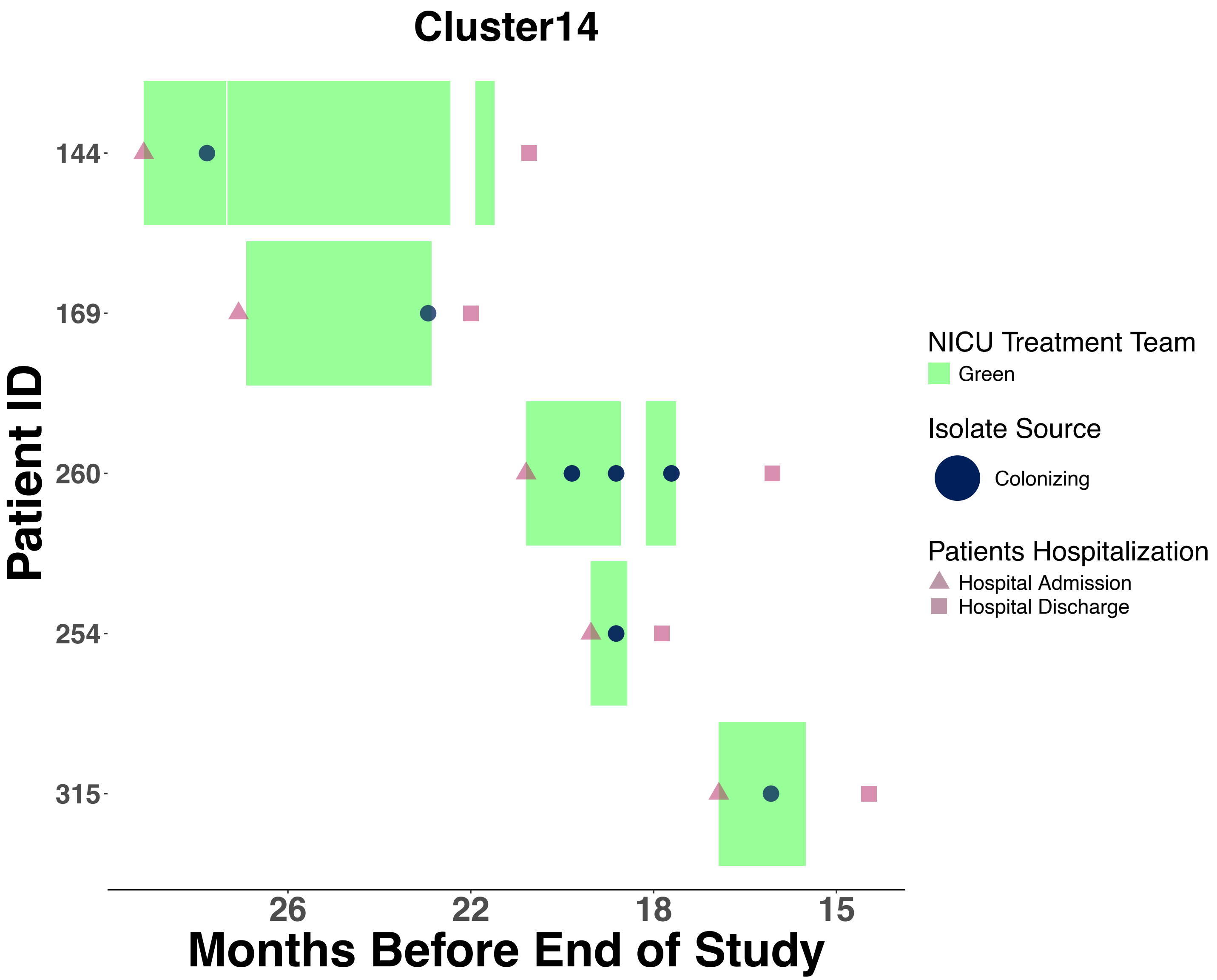
Cluster14
Patient ID
144
169
260
254
315
26
22
18
15
Months Before End of Study
NICU Treatment Team
Green
Isolate Source
Colonizing
Patients Hospitalization
Hospital Admission
Hospital Discharge

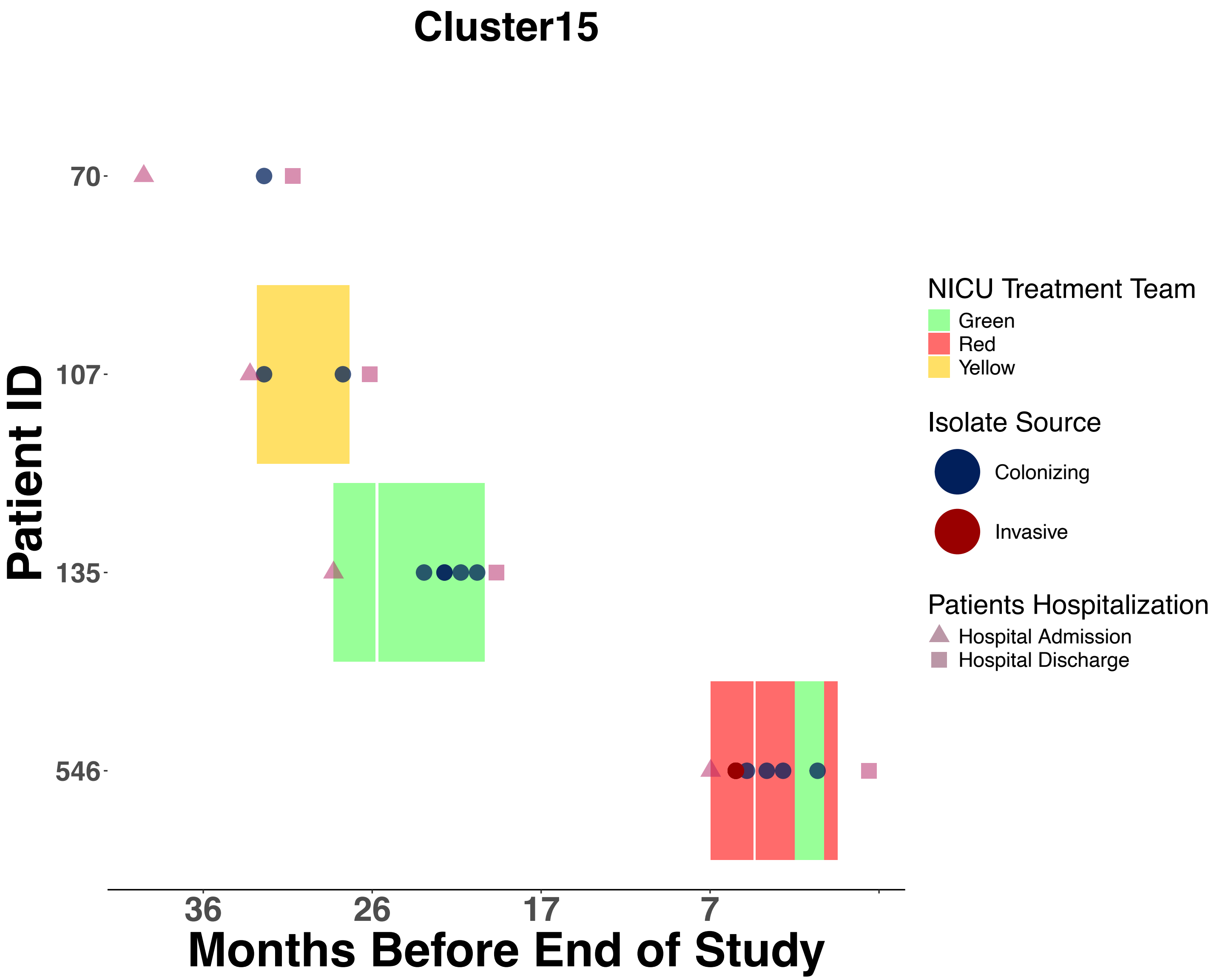
Cluster15
Patient ID
70
107
135
546
36
26
17
7
Months Before End of Study
NICU Treatment Team
Green
Red
Yellow
Isolate Source
Colonizing
Invasive
Patients Hospitalization
Hospital Admission
Hospital Discharge

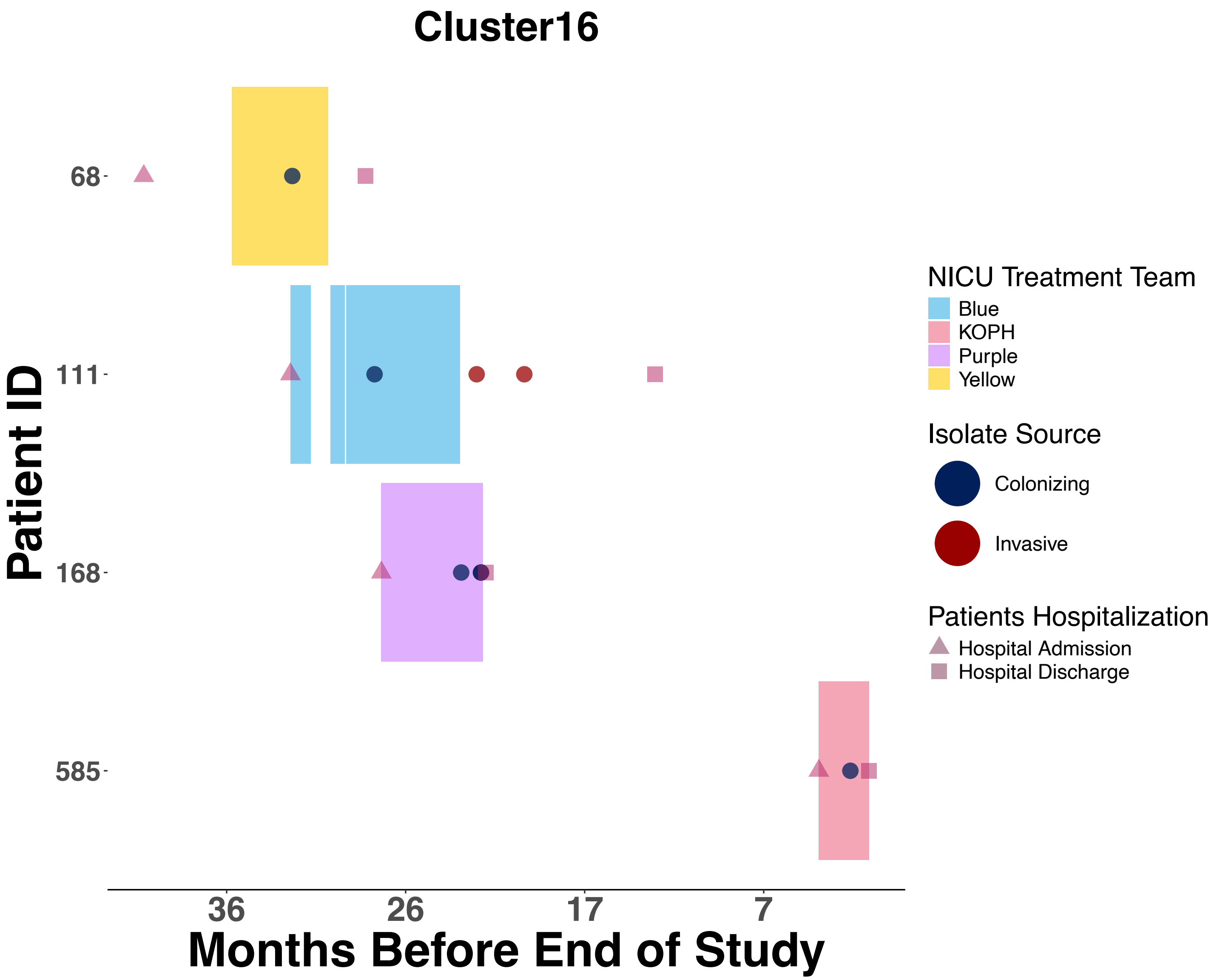

Cluster16
Patient ID
68
111
168
585
36
26
17
7
Months Before End of Study
NICU Treatment Team
Blue
KOPH
Purple
Yellow
Isolate Source
Colonizing
Invasive
Patients Hospitalization
Hospital Admission
Hospital Discharge

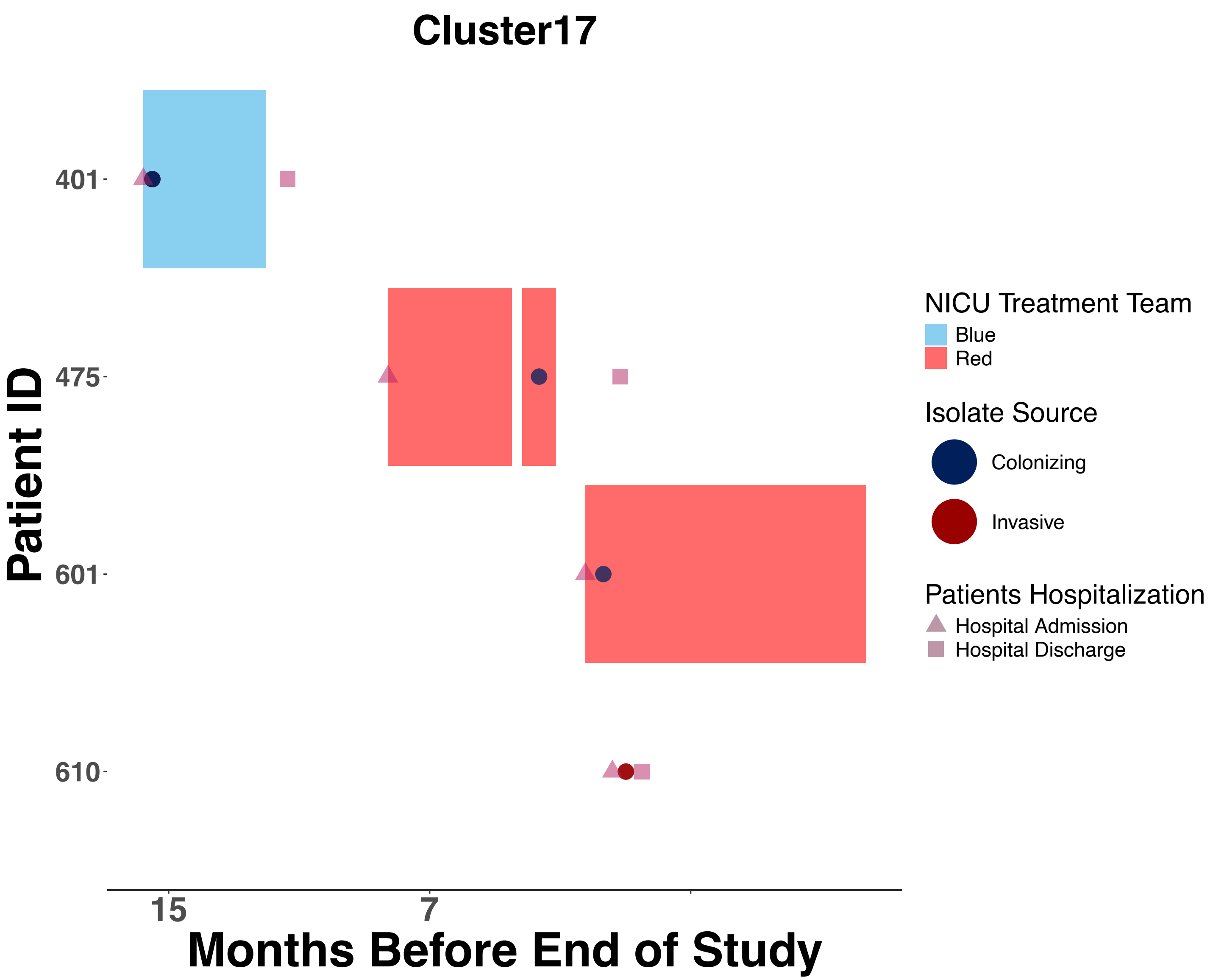

Cluster17
Patient ID
401
475
601
610
15
7
Months Before End of Study
NICU Treatment Team
Blue
Red
Isolate Source
Colonizing
Invasive
Patients Hospitalization
Hospital Admission
Hospital Discharge

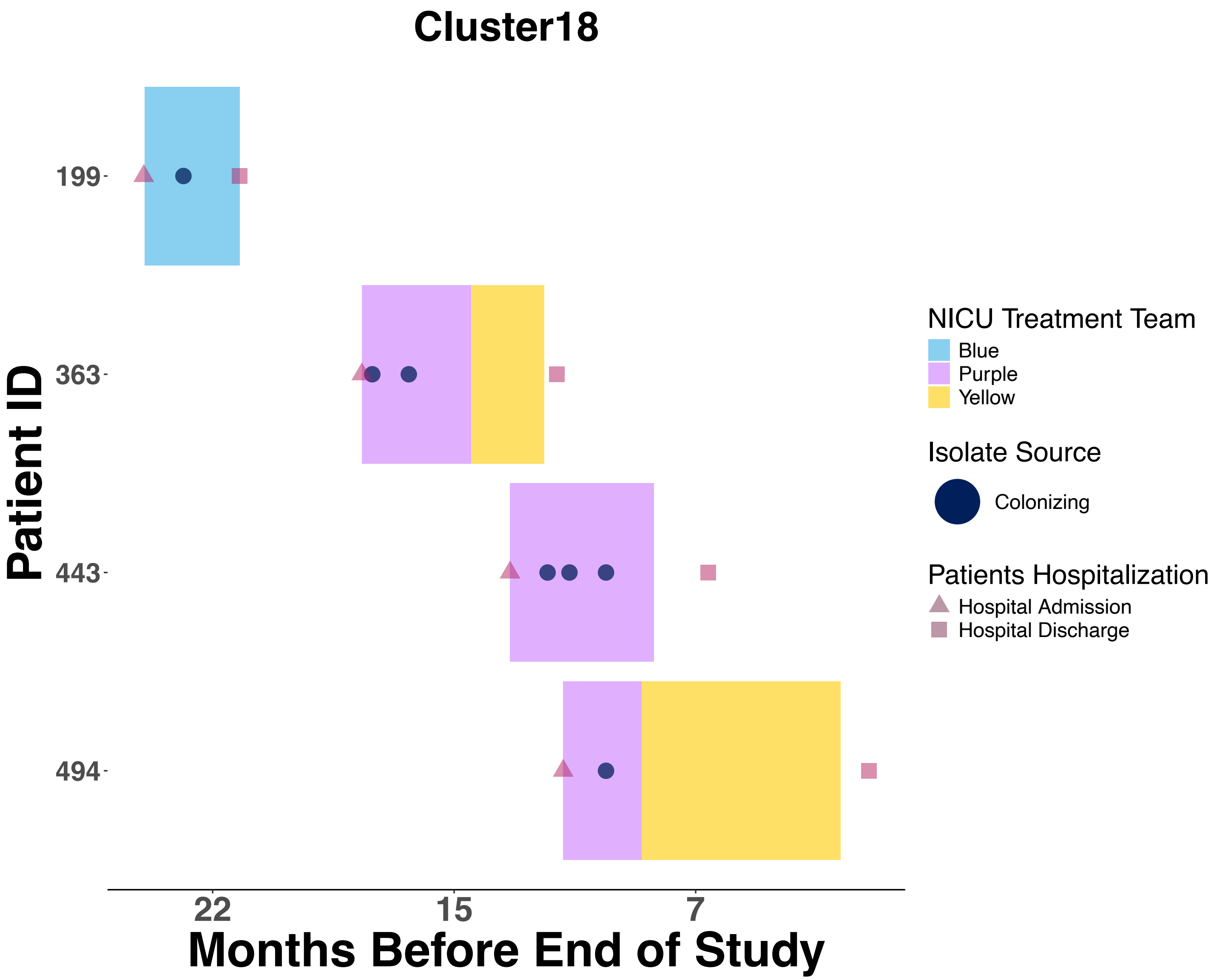
Cluster18
Patient ID
199
363
443
494
22
15
7
Months Before End of Study
NICU Treatment Team
Blue
Purple
Yellow
Isolate Source
Colonizing
Patients Hospitalization
Hospital Admission
Hospital Discharge

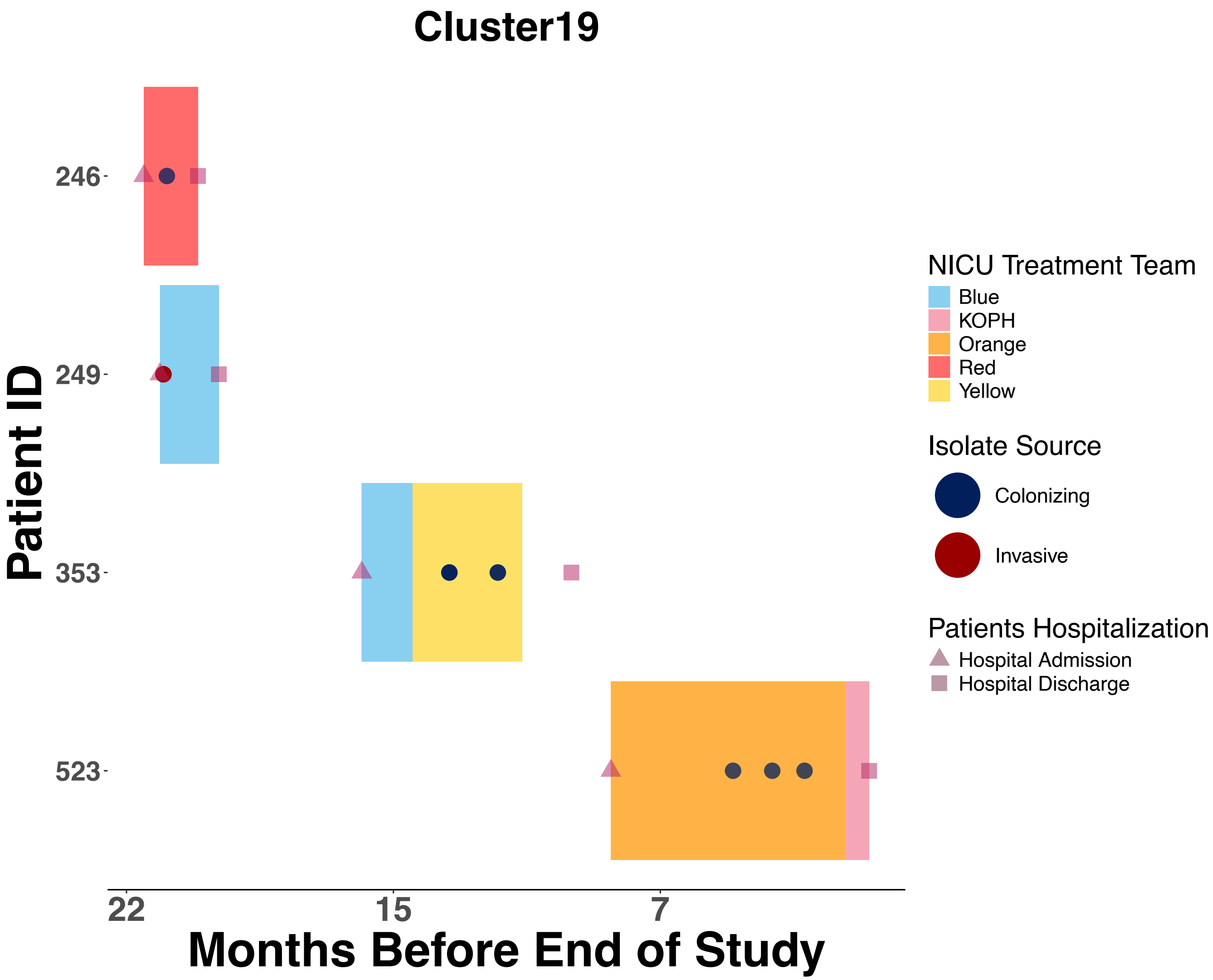

Cluster19
Patient ID
246
249
353
523
22
15
7
Months Before End of Study
NICU Treatment Team
Blue
KOPH
Orange
Red
Yellow
Isolate Source
Colonizing
Invasive
Patients Hospitalization
Hospital Admission
Hospital Discharge

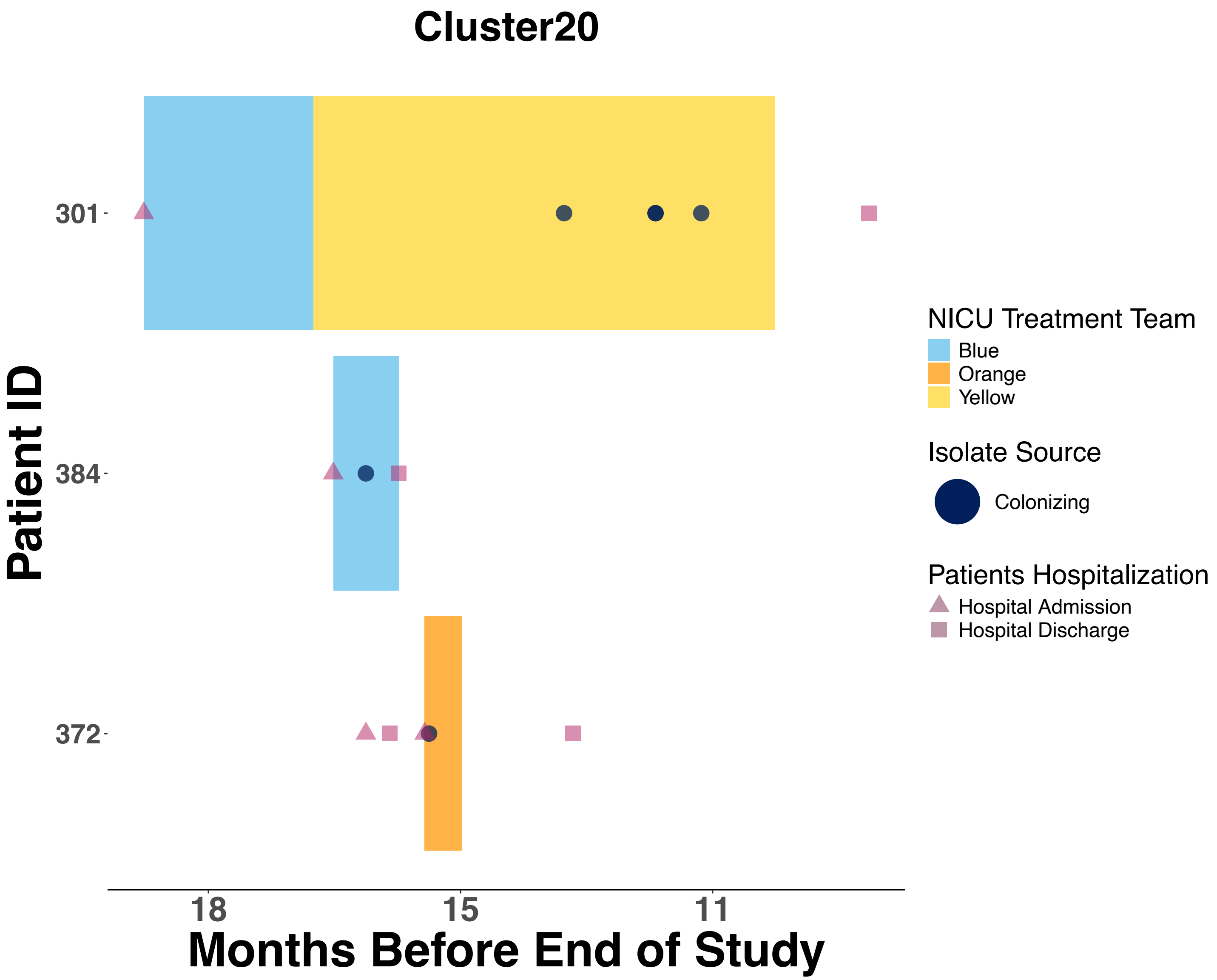

Cluster20
Patient ID
301
384
372
18
15
11
Months Before End of Study
NICU Treatment Team
Blue
Orange
Yellow
Isolate Source
Colonizing
Patients Hospitalization
Hospital Admission
Hospital Discharge

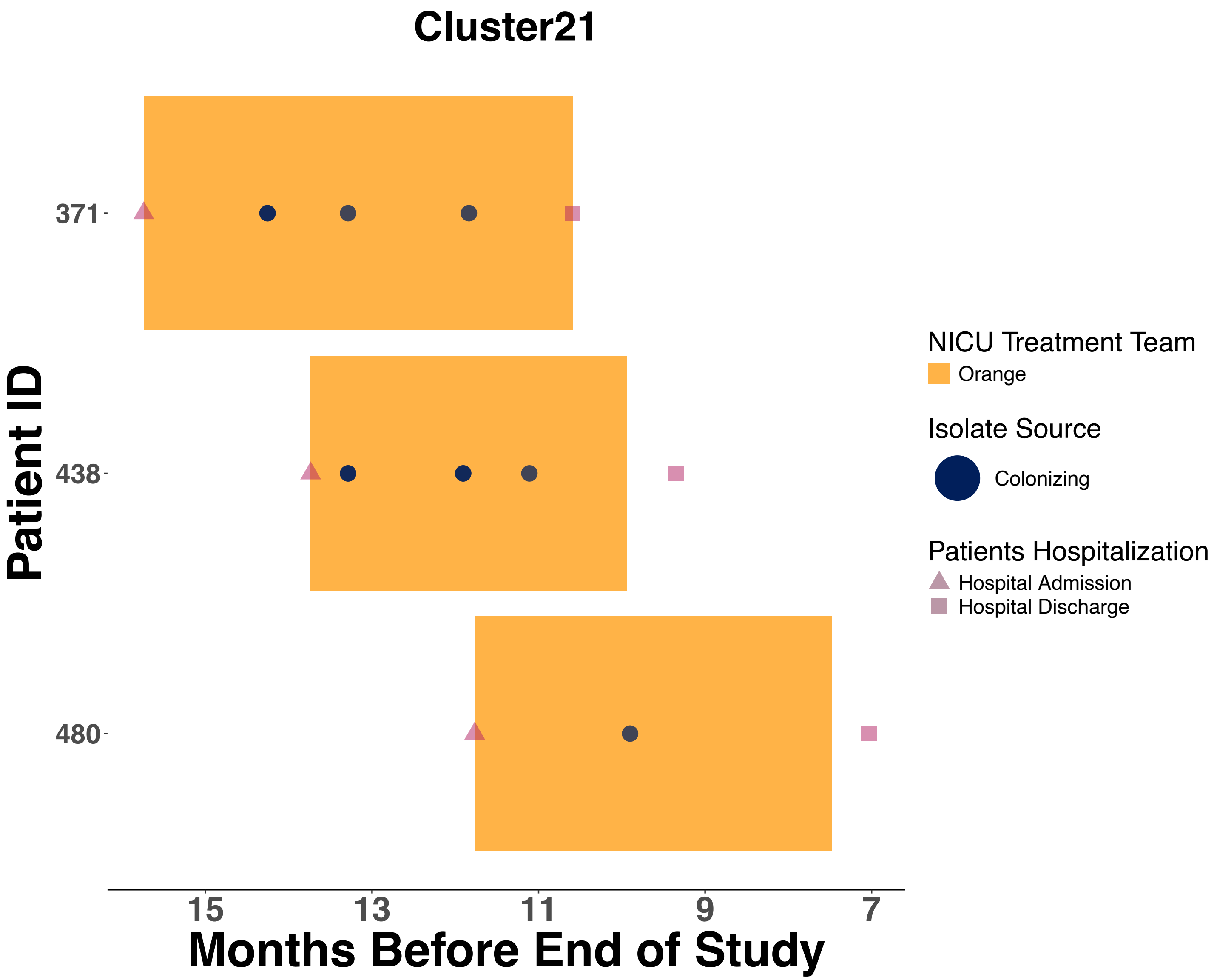

Cluster21
Patient ID
371
438
480
15
13
11
9
7
Months Before End of Study
NICU Treatment Team
Orange
Isolate Source
Colonizing
Patients Hospitalization
Hospital Admission
Hospital Discharge

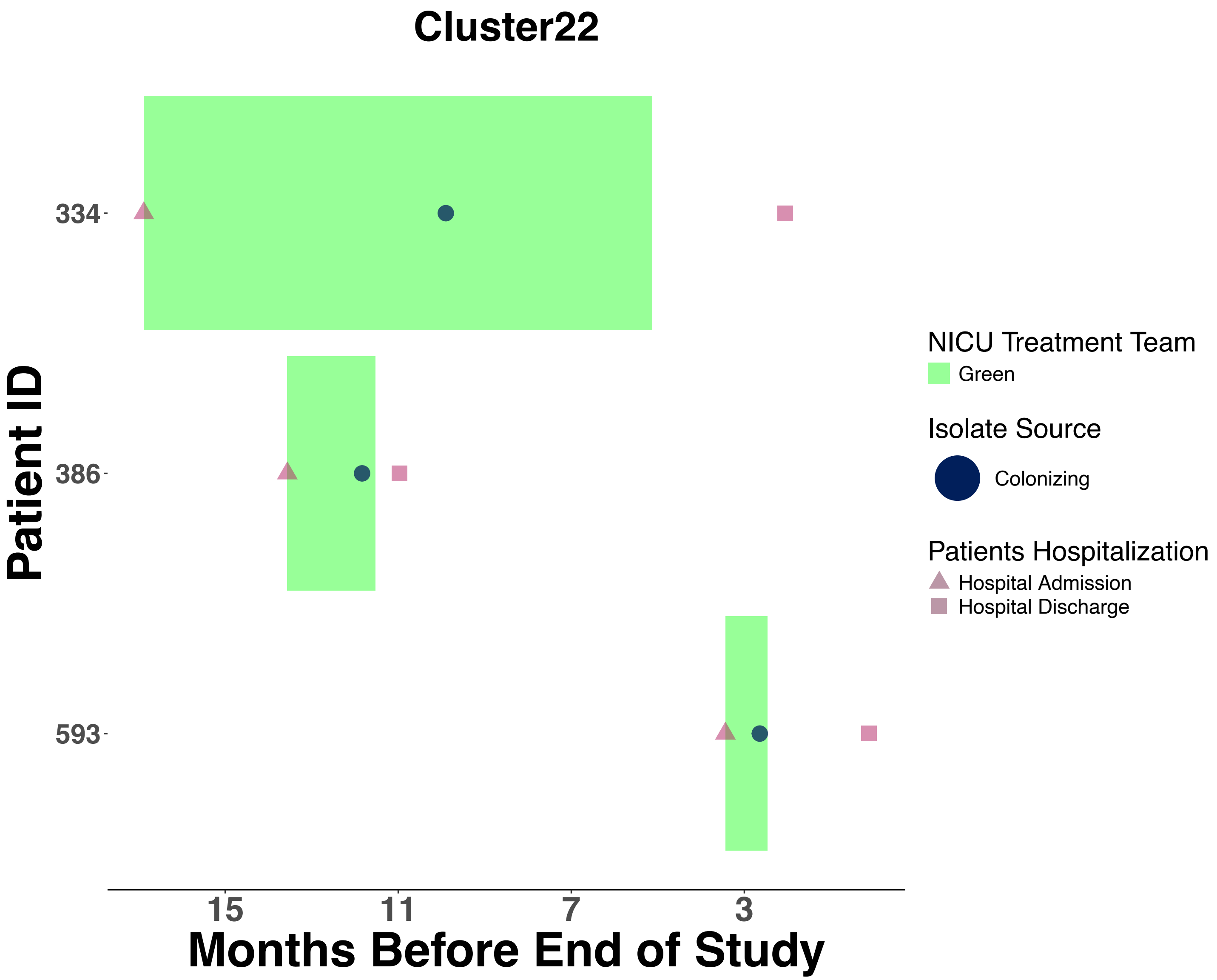

Cluster22
Patient ID
334
386
593
15
11
7
3
Months Before End of Study
NICU Treatment Team
Green
Isolate Source
Colonizing
Patients Hospitalization
Hospital Admission
Hospital Discharge

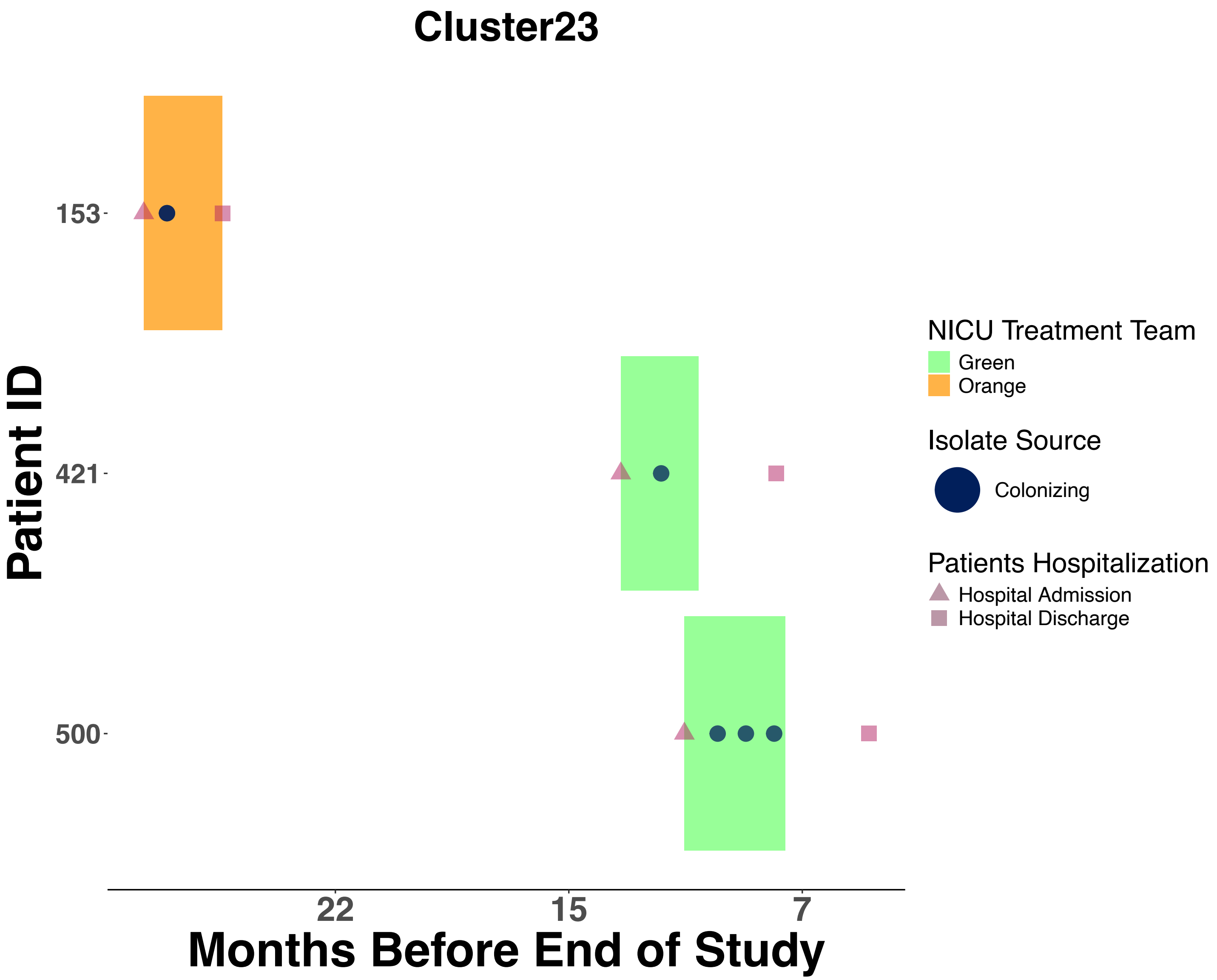

Cluster23
Patient ID
153
421
500
22
15
7
Months Before End of Study
NICU Treatment Team
Green
Orange
Isolate Source
Colonizing
Patients Hospitalization
Hospital Admission
Hospital Discharge

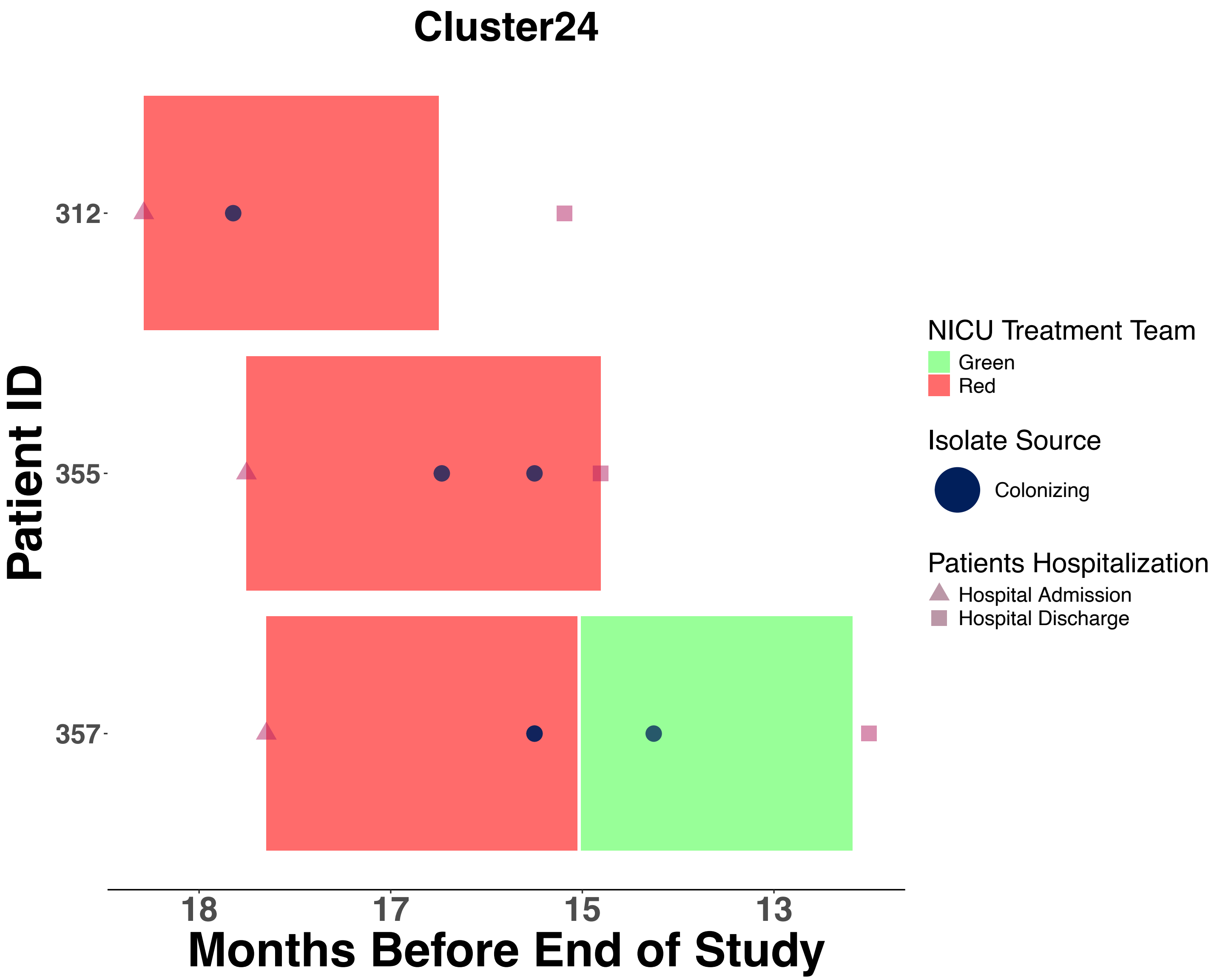
Cluster24
Patient ID
312
355
357
18
17
15
13
Months Before End of Study
NICU Treatment Team
Green
Red
Isolate Source
Colonizing
Patients Hospitalization
Hospital Admission
Hospital Discharge

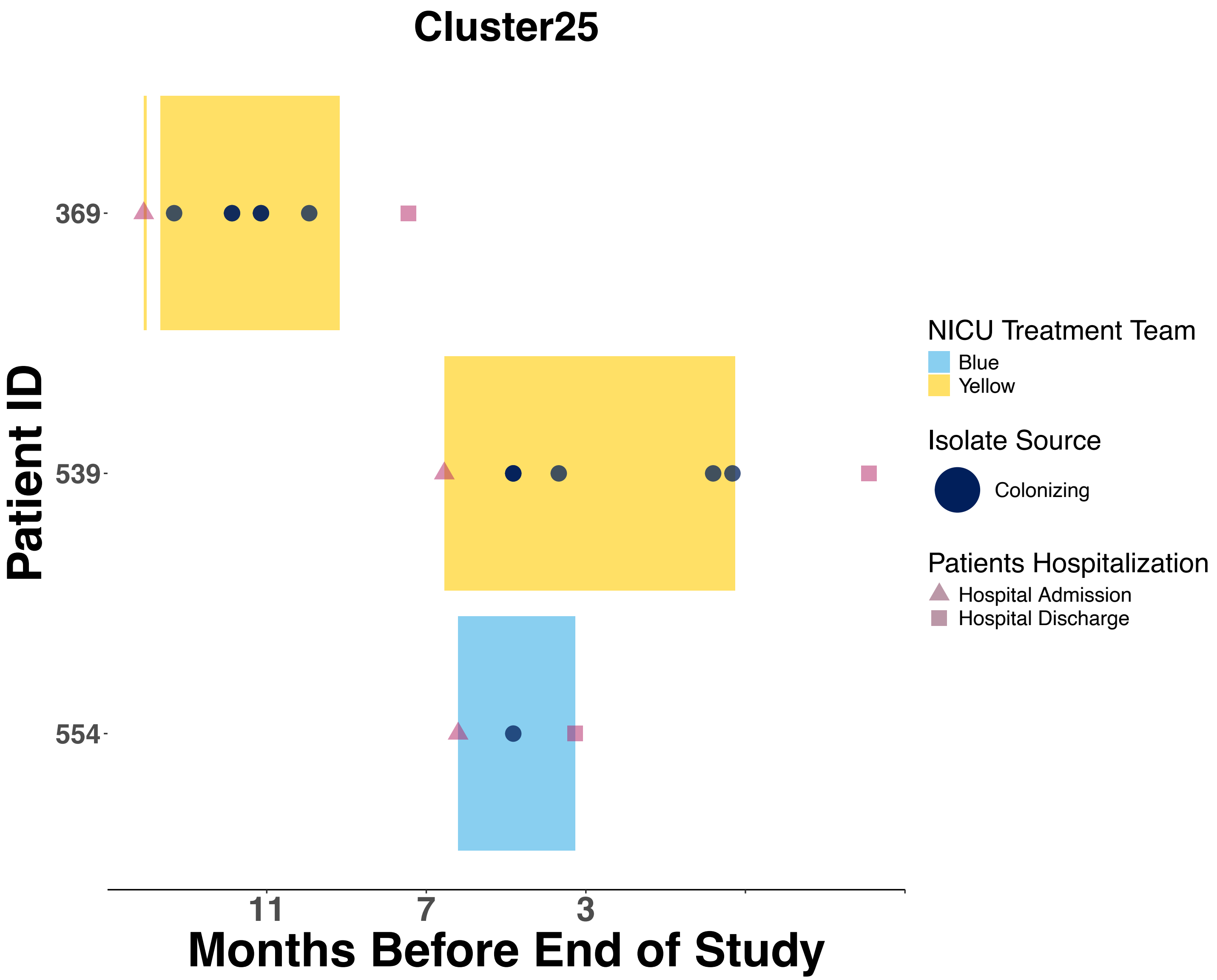
Cluster25
369
539
554
Patient ID
11
7
3
Months Before End of Study
NICU Treatment Team
Blue
Yellow
Isolate Source
Colonizing
Patients Hospitalization
Hospital Admission
Hospital Discharge

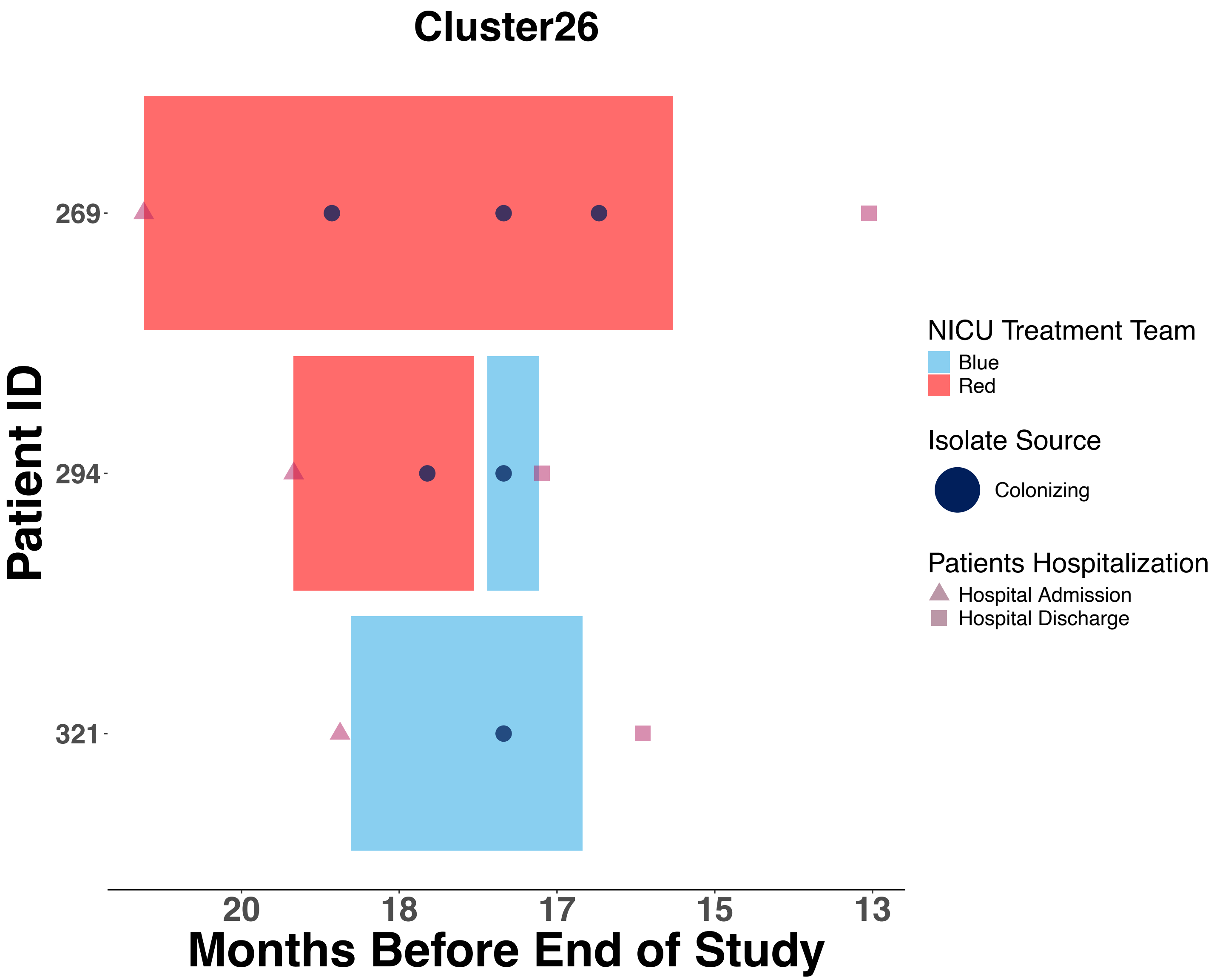
Cluster26
Patient ID
269
294
321
20
18
17
15
13
Months Before End of Study
NICU Treatment Team
Blue
Red
Isolate Source
Colonizing
Patients Hospitalization
Hospital Admission
Hospital Discharge

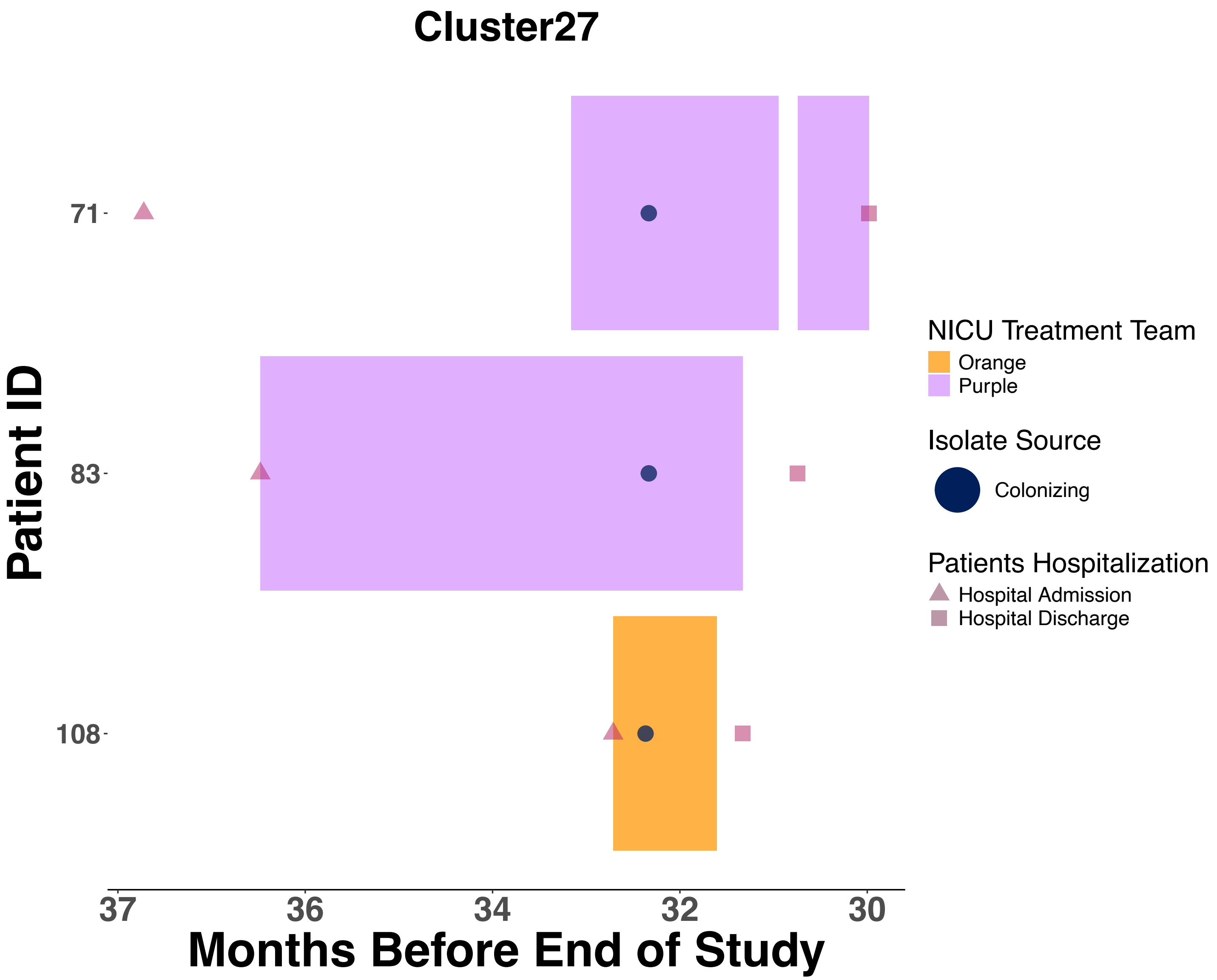
Cluster27
Patient ID
71
83
108
37
36
34
32
30
Months Before End of Study
NICU Treatment Team
Orange
Purple
Isolate Source
Colonizing
Patients Hospitalization
Hospital Admission
Hospital Discharge

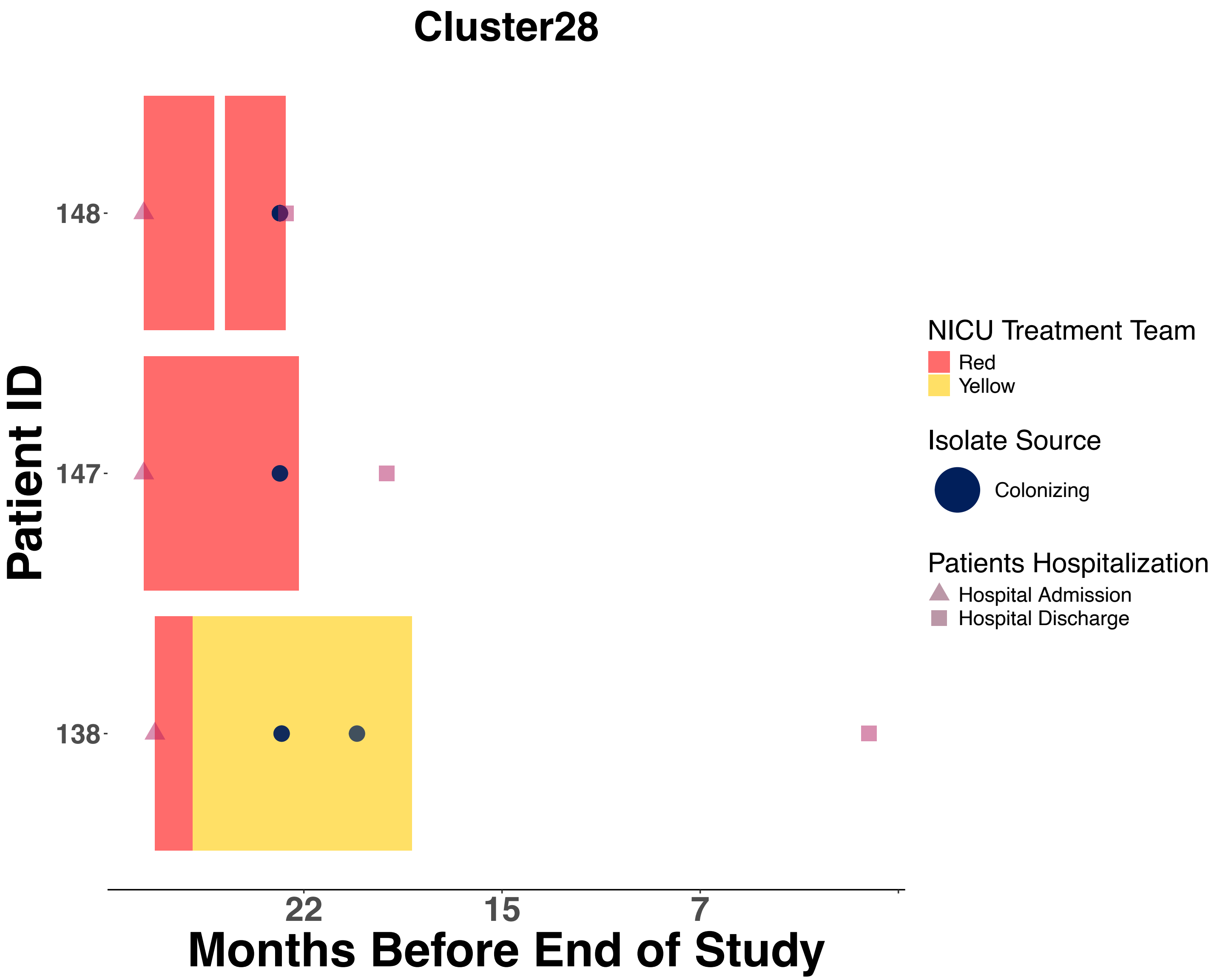
Cluster28
Patient ID
148
147
138
22
15
7
Months Before End of Study
NICU Treatment Team
Red
Yellow
Isolate Source
Colonizing
Patients Hospitalization
Hospital Admission
Hospital Discharge

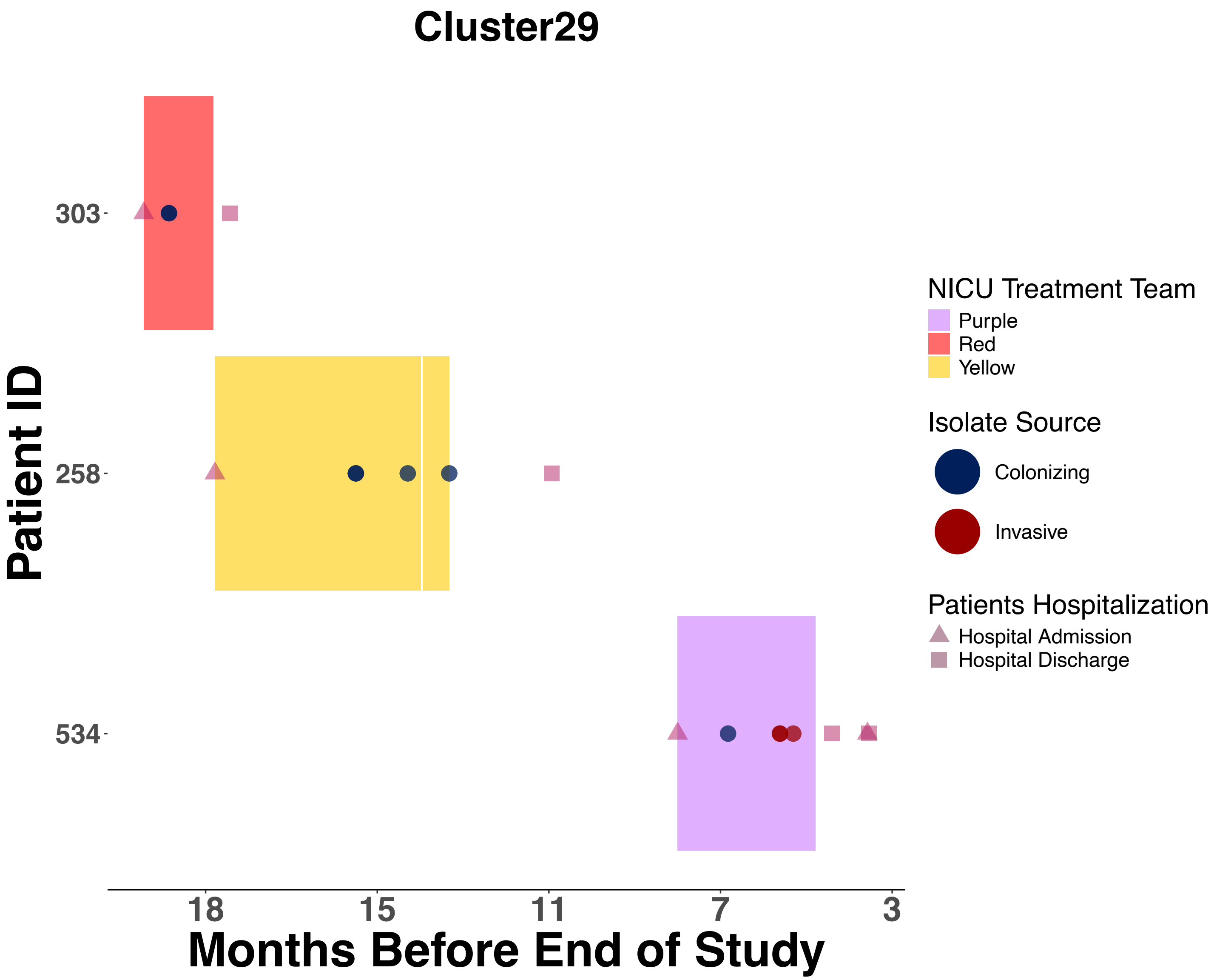

Cluster29
303
258
534
Patient ID
18
15
11
7
3
Months Before End of Study
NICU Treatment Team
Purple
Red
Yellow
Isolate Source
Colonizing
Invasive
Patients Hospitalization
Hospital Admission
Hospital Discharge

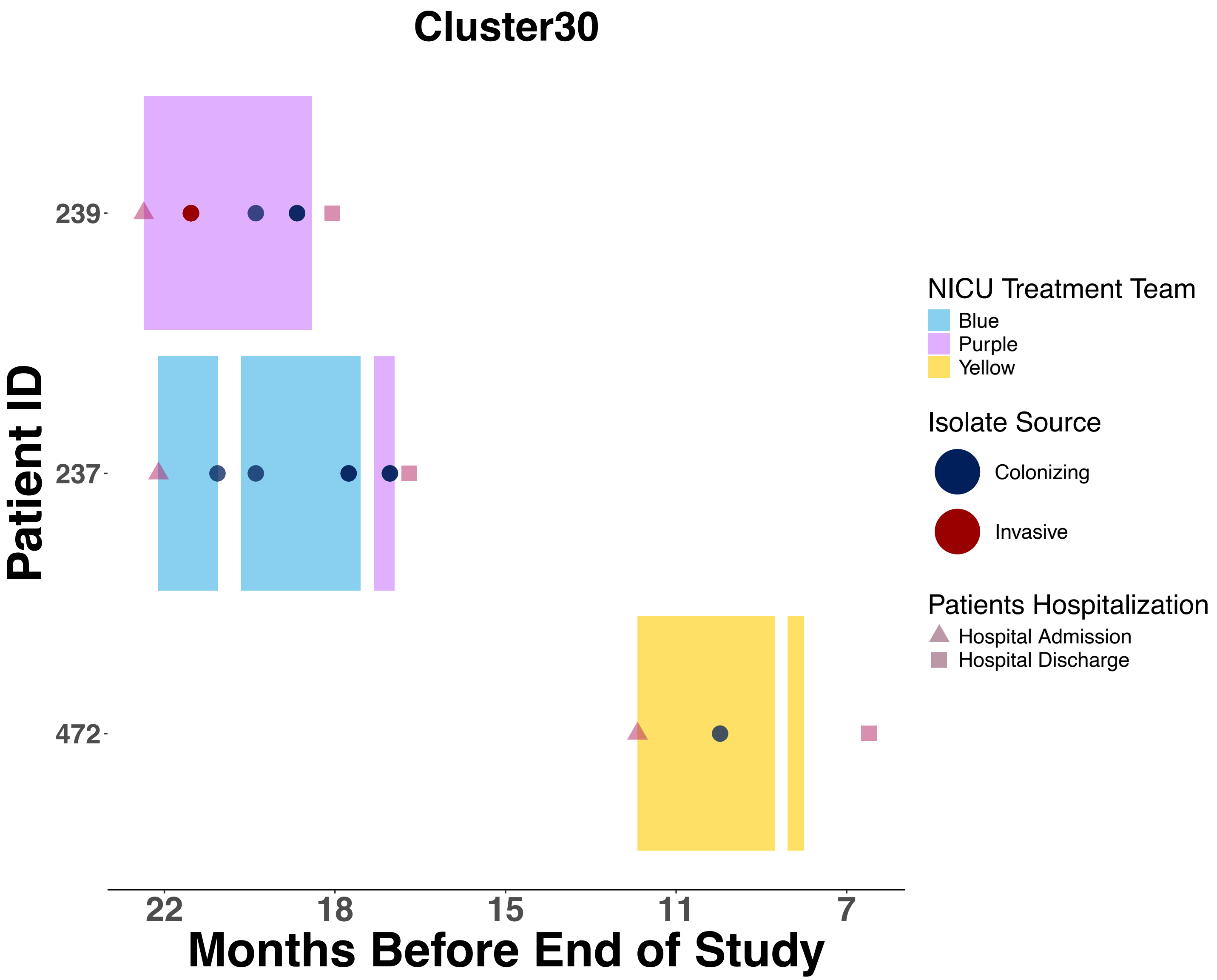
Cluster30
Patient ID
239
237
472
22
18
15
11
7
Months Before End of Study
NICU Treatment Team
Blue
Purple
Yellow
Isolate Source
Colonizing
Invasive
Patients Hospitalization
Hospital Admission
Hospital Discharge

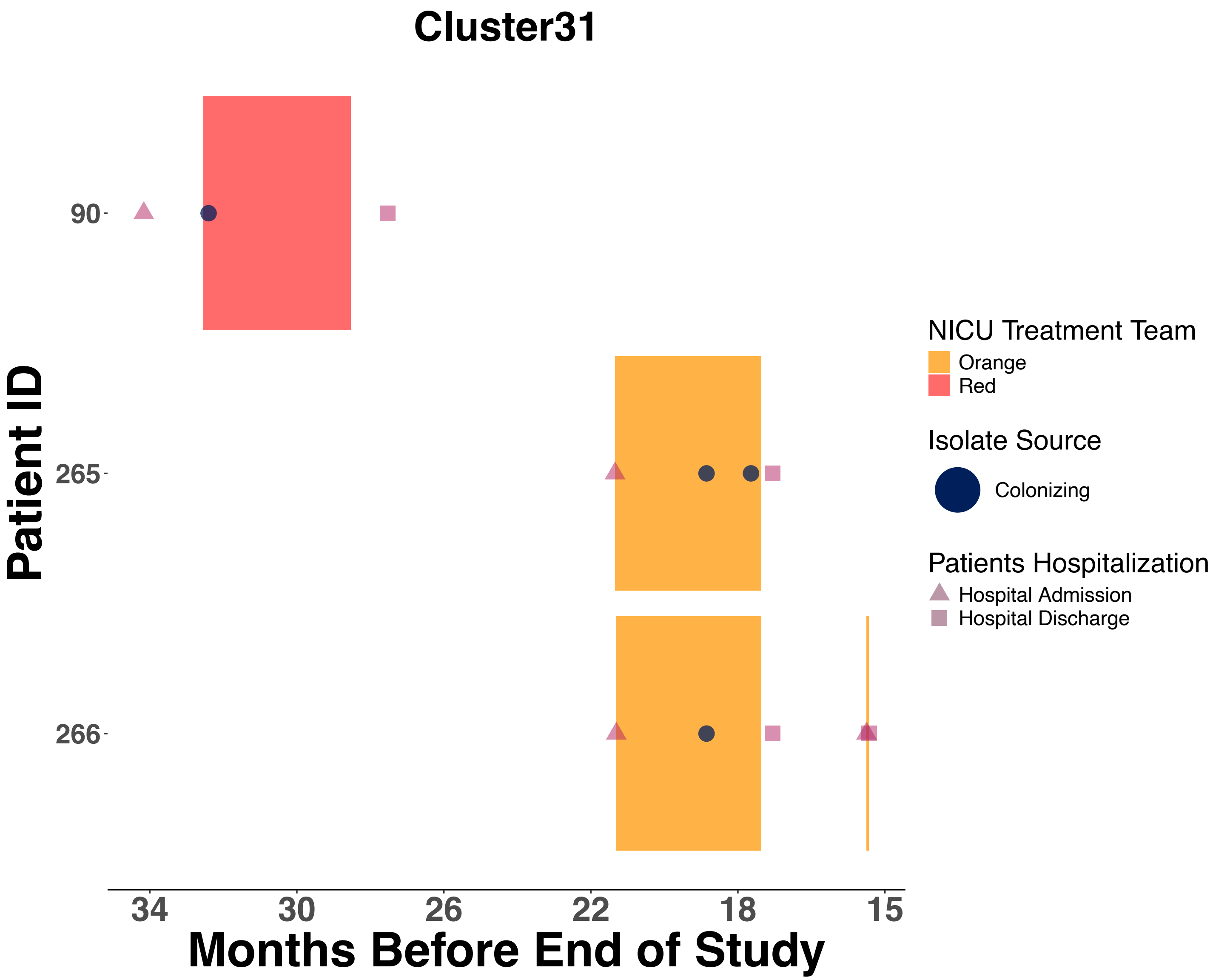

Cluster31
Patient ID
90
265
266
34
30
26
22
18
15
Months Before End of Study
NICU Treatment Team
Orange
Red
Isolate Source
Colonizing
Patients Hospitalization
Hospital Admission
Hospital Discharge

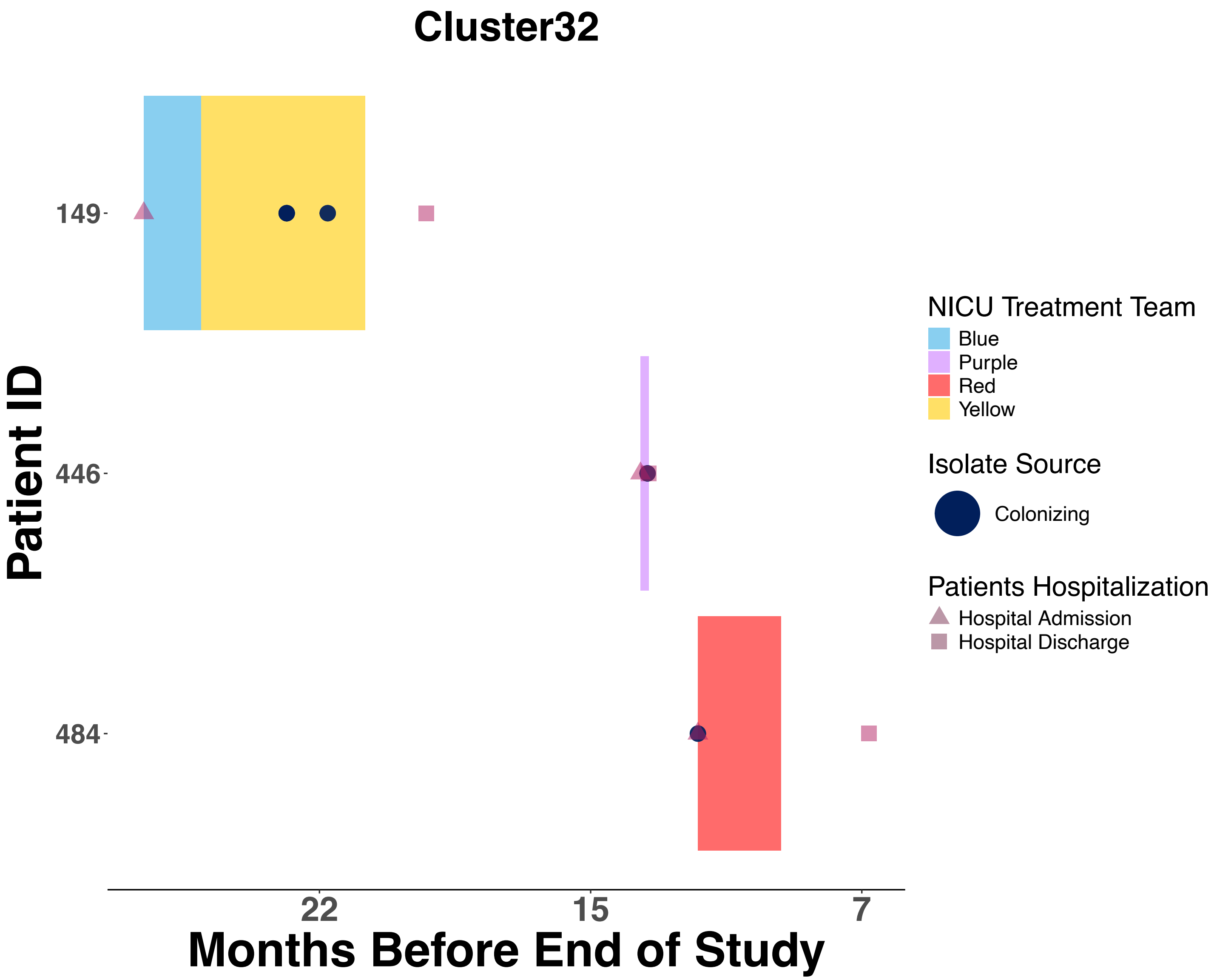
Cluster32
Patient ID
149
446
484
22
15
7
Months Before End of Study
NICU Treatment Team
Blue
Purple
Red
Yellow
Isolate Source
Colonizing
Patients Hospitalization
Hospital Admission
Hospital Discharge

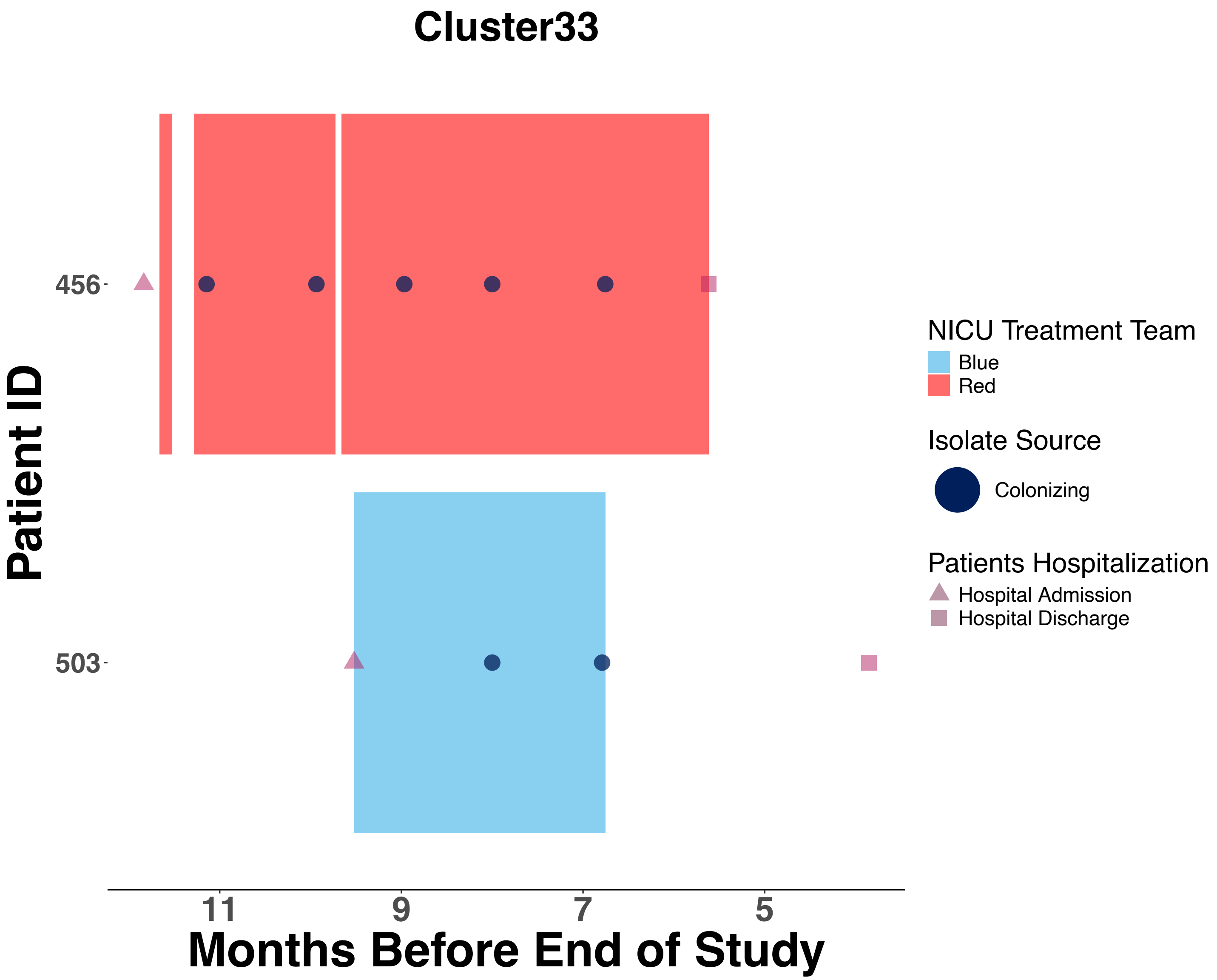
Cluster33
Patient ID
456
503
11
9
7
5
Months Before End of Study
NICU Treatment Team
Blue
Red
Isolate Source
Colonizing
Patients Hospitalization
Hospital Admission
Hospital Discharge

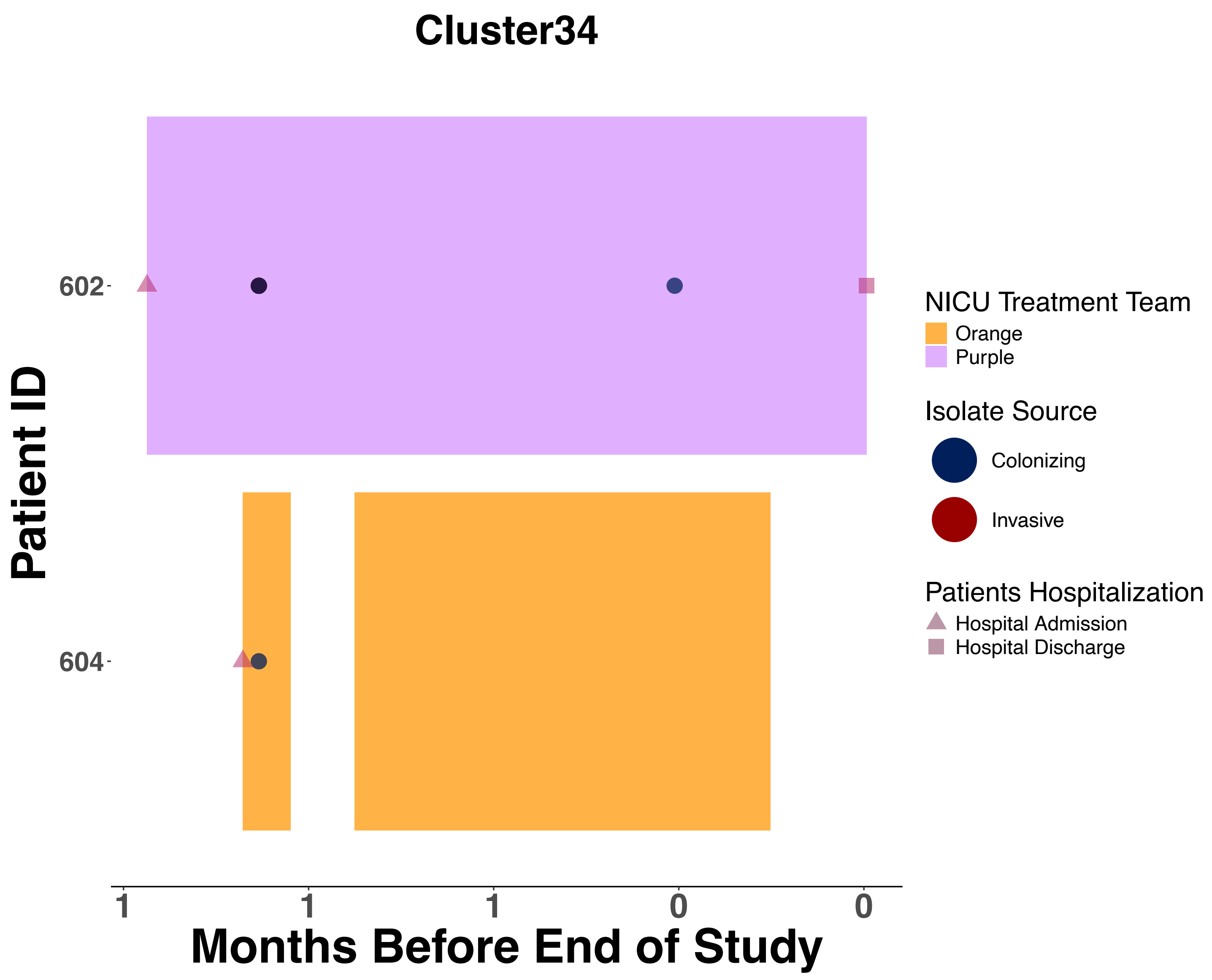

Cluster34
Patient ID
602
604
1
1
1
0
0
Months Before End of Study
NICU Treatment Team
Orange
Purple
Isolate Source
Colonizing
Invasive
Patients Hospitalization
Hospital Admission
Hospital Discharge

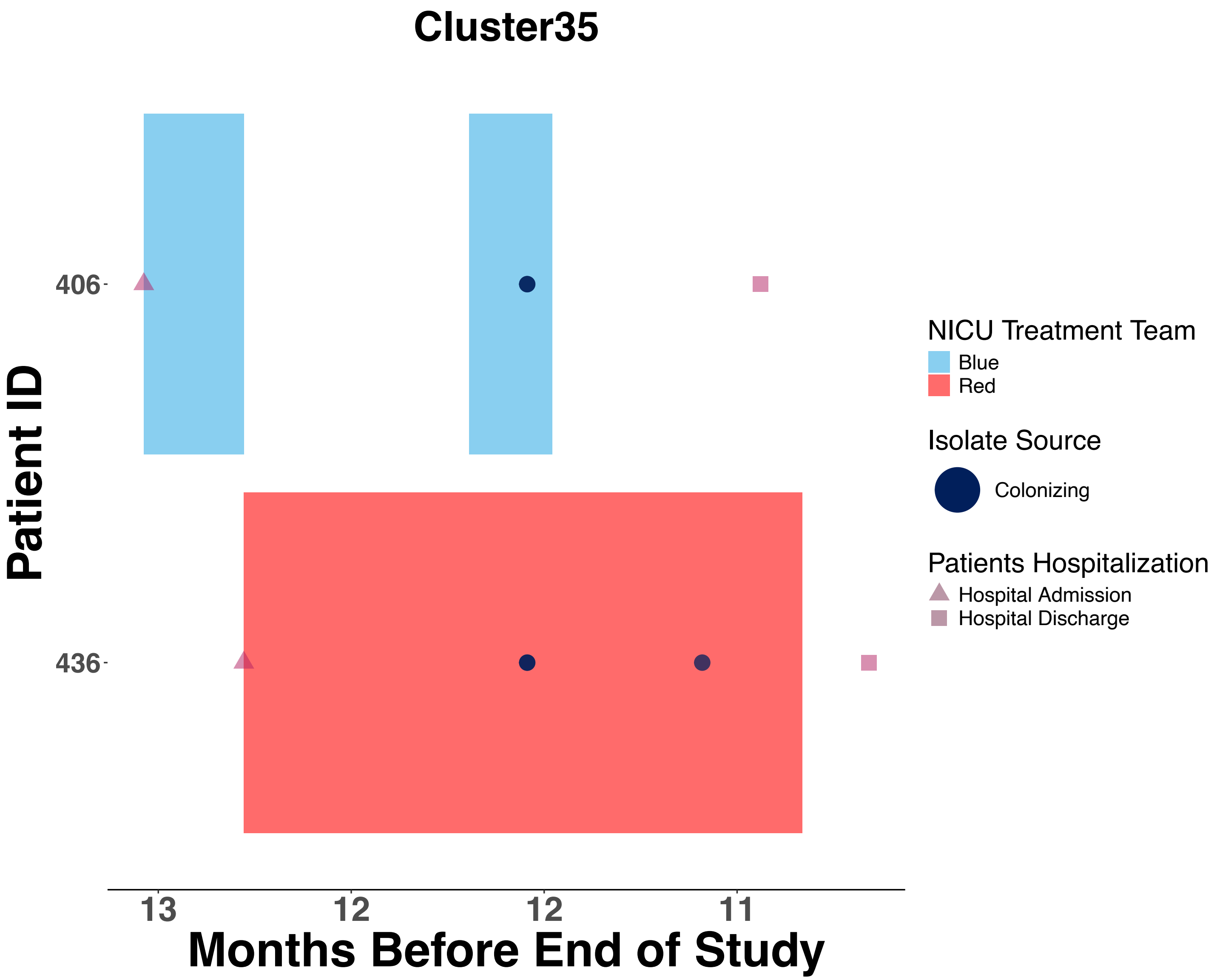

Cluster35
Patient ID
406
436
13
12
12
11
Months Before End of Study
NICU Treatment Team
Blue
Red
Isolate Source
Colonizing
Patients Hospitalization
Hospital Admission
Hospital Discharge

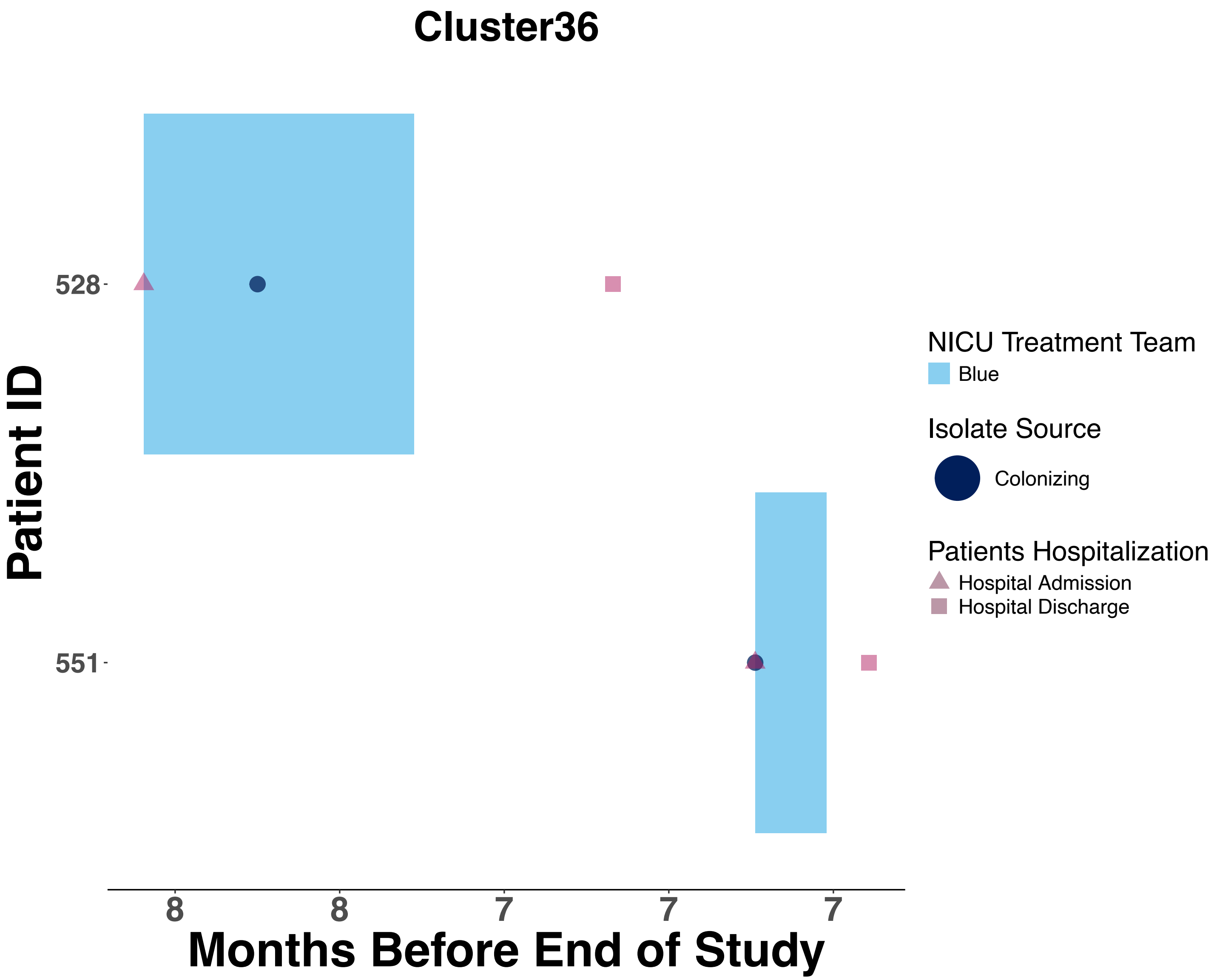

Cluster36
Patient ID
528
551
Months Before End of Study
8
8
7
7
7
NICU Treatment Team
Blue
Isolate Source
Colonizing
Patients Hospitalization
Hospital Admission
Hospital Discharge

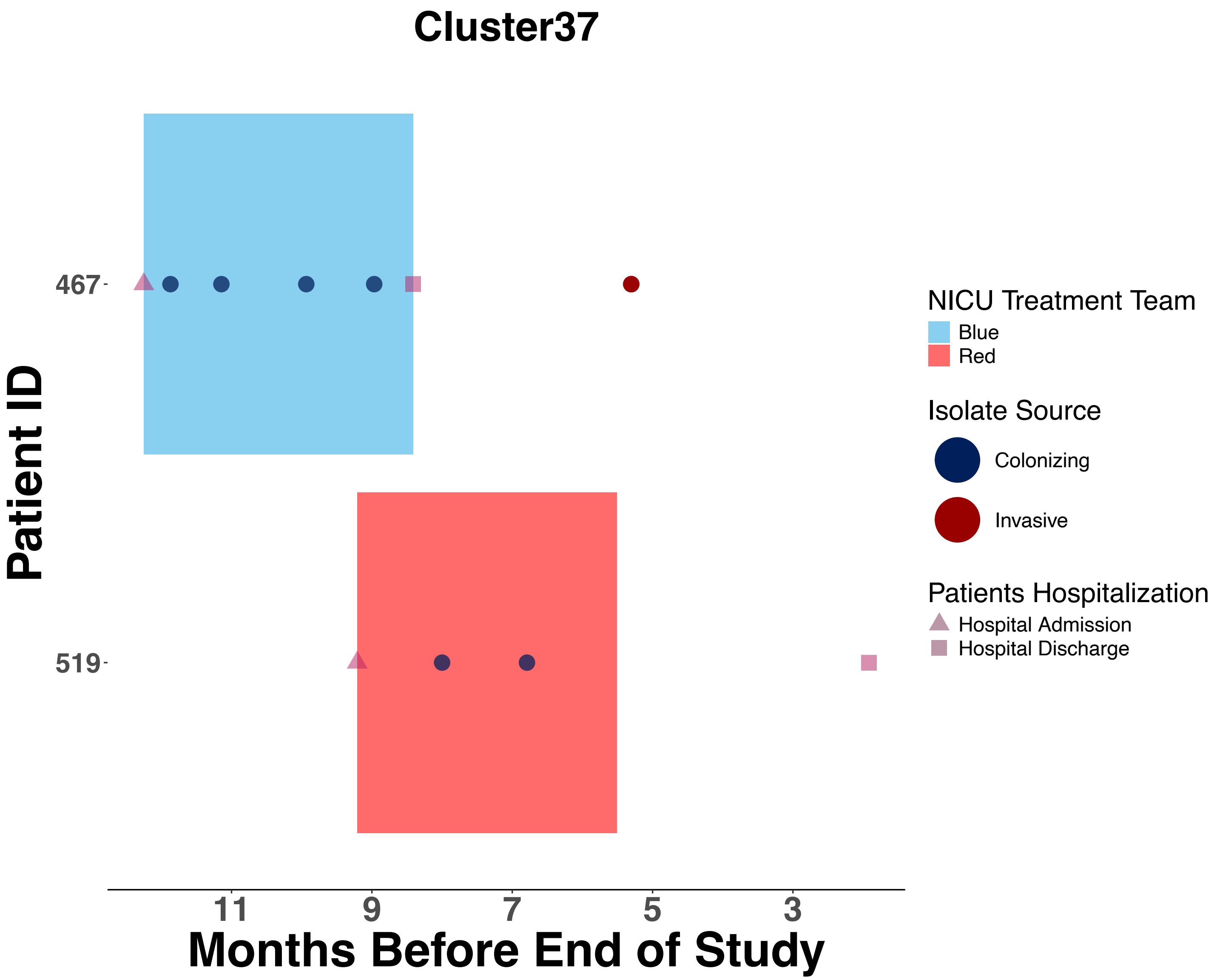

Cluster37
467
519
Patient ID
11
9
7
5
3
Months Before End of Study
NICU Treatment Team
Blue
Red
Isolate Source
Colonizing
Invasive
Patients Hospitalization
Hospital Admission
Hospital Discharge

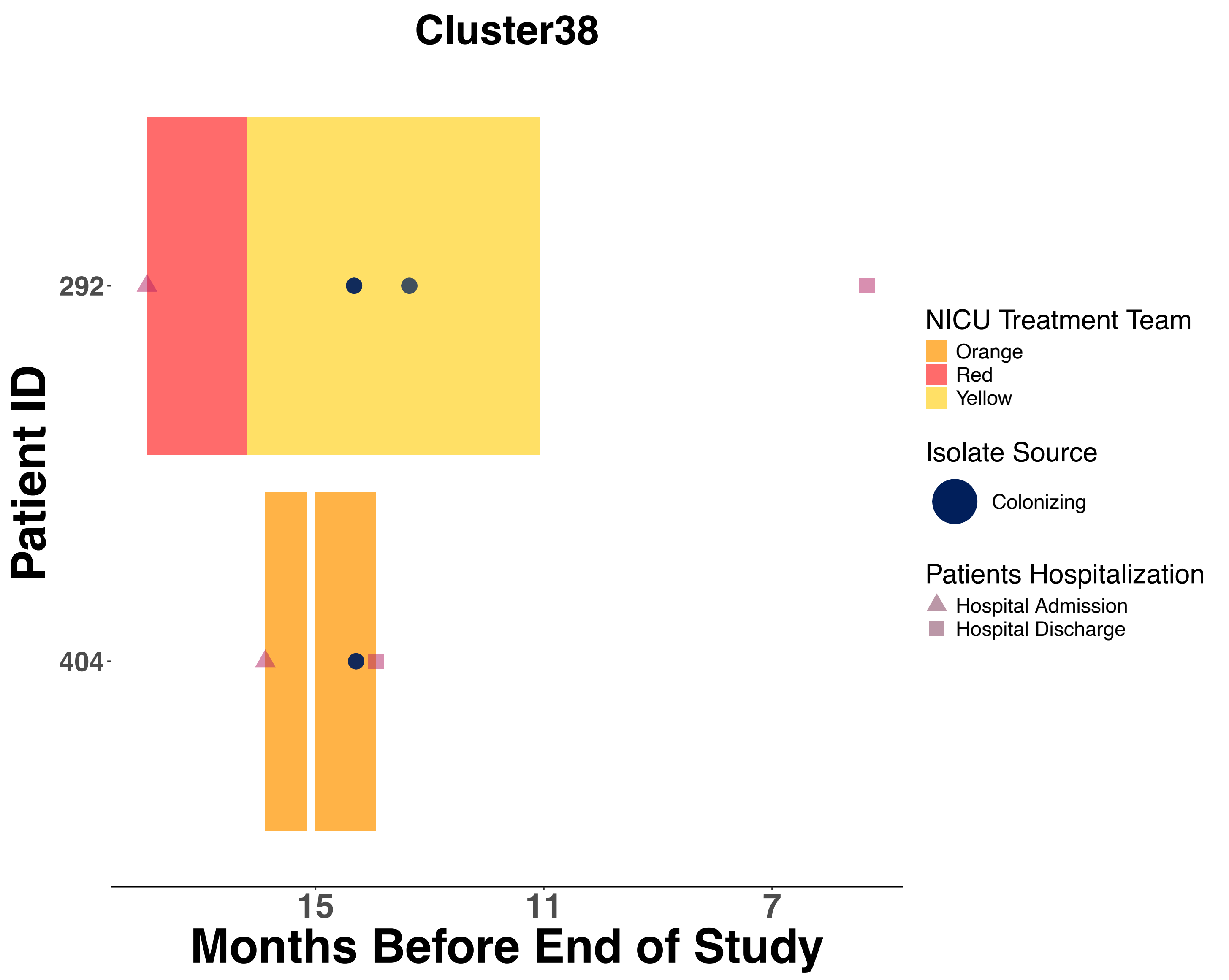
Cluster38
Patient ID
292
404
15
11
7
Months Before End of Study
NICU Treatment Team
Orange
Red
Yellow
Isolate Source
Colonizing
Patients Hospitalization
Hospital Admission
Hospital Discharge

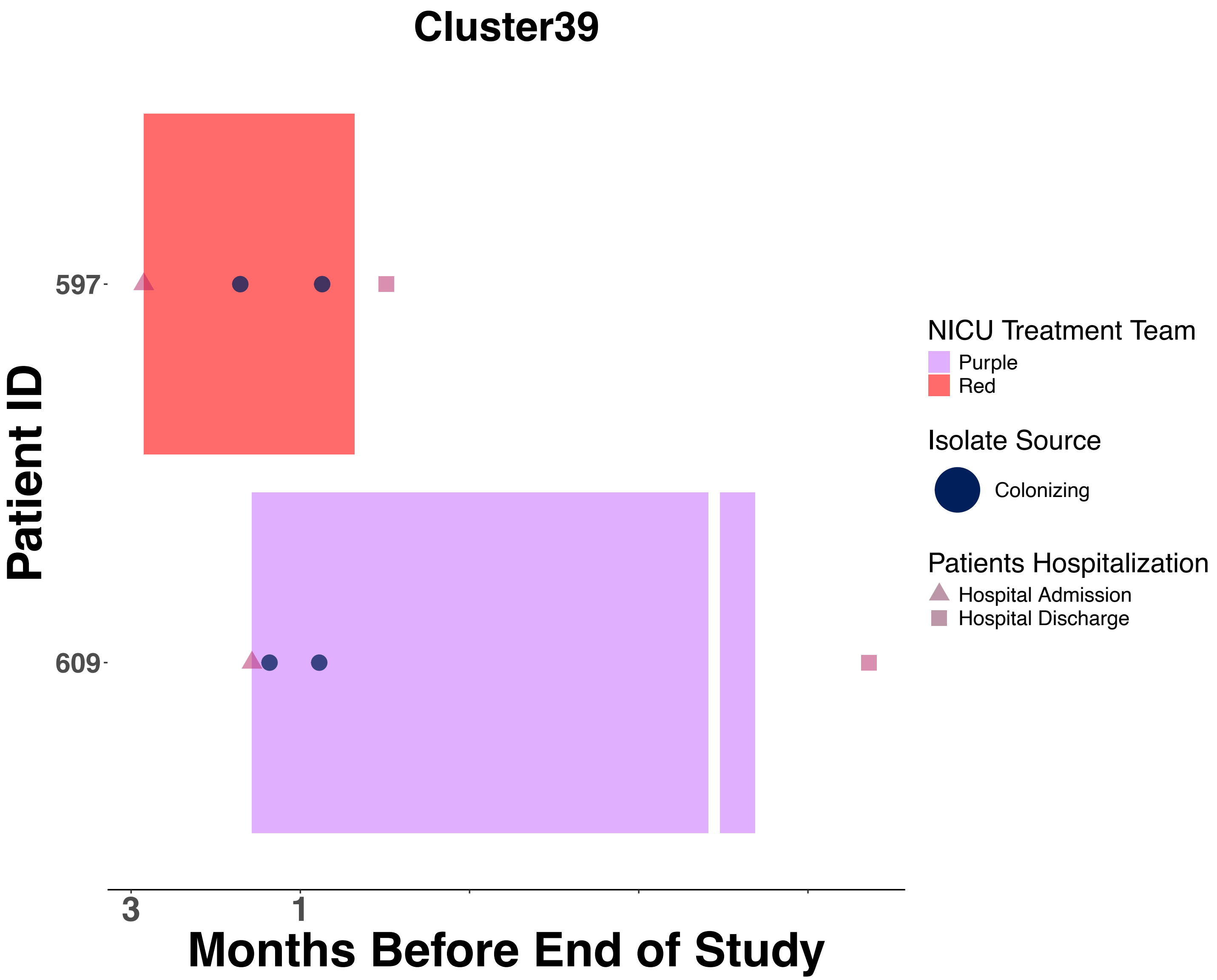
Cluster39
Patient ID
597
609
3
1
Months Before End of Study
NICU Treatment Team
Purple
Red
Isolate Source
Colonizing
Patients Hospitalization
Hospital Admission
Hospital Discharge

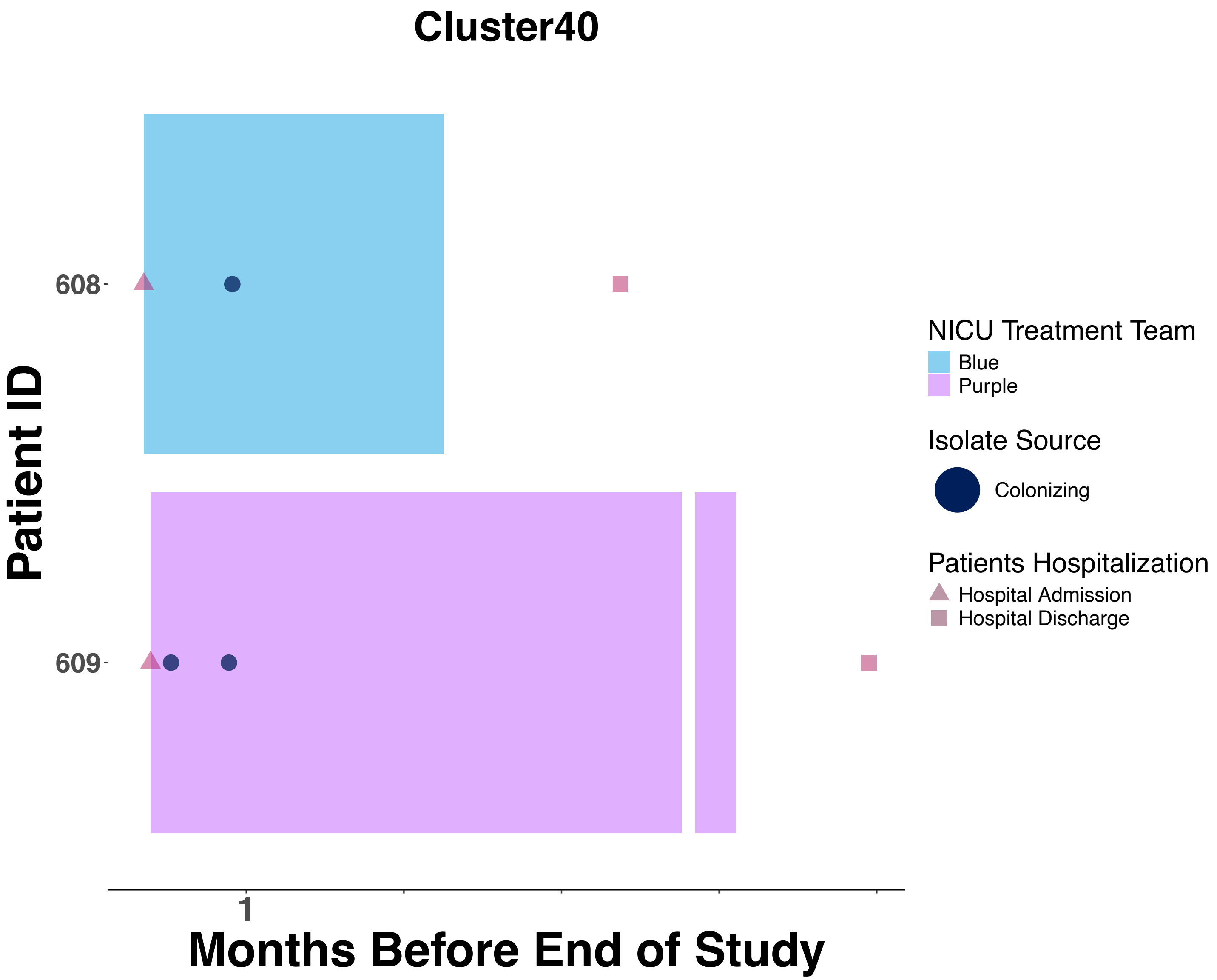

Cluster40
Patient ID
608
609
1
Months Before End of Study
NICU Treatment Team
Blue
Purple
Isolate Source
Colonizing
Patients Hospitalization
Hospital Admission
Hospital Discharge

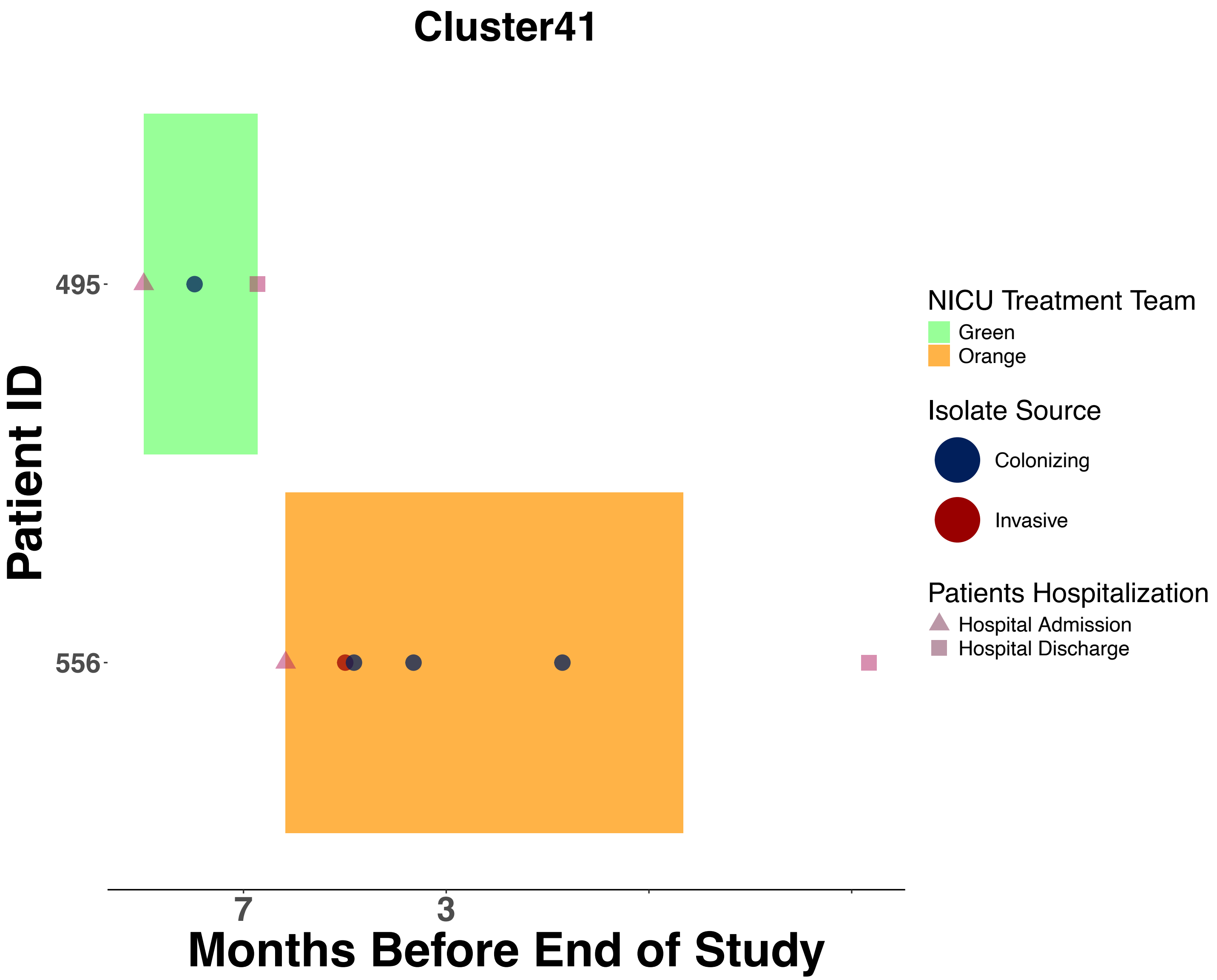
Cluster41
Patient ID
495
556
7
3
Months Before End of Study
NICU Treatment Team
Green
Orange
Isolate Source
Colonizing
Invasive
Patients Hospitalization
Hospital Admission
Hospital Discharge

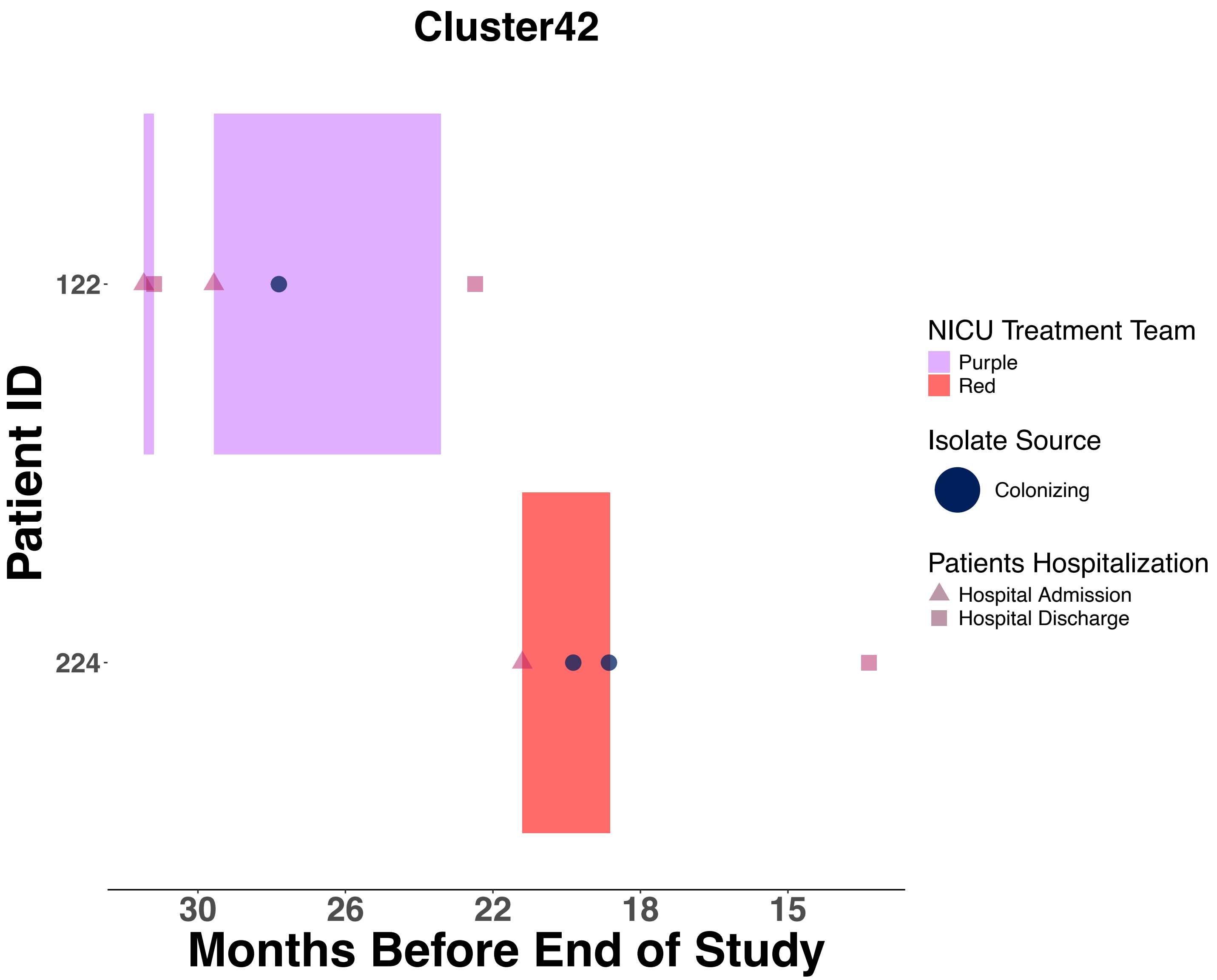
Cluster42
Patient ID
122
224
30
26
22
18
15
Months Before End of Study
NICU Treatment Team
Purple
Red
Isolate Source
Colonizing
Patients Hospitalization
Hospital Admission
Hospital Discharge

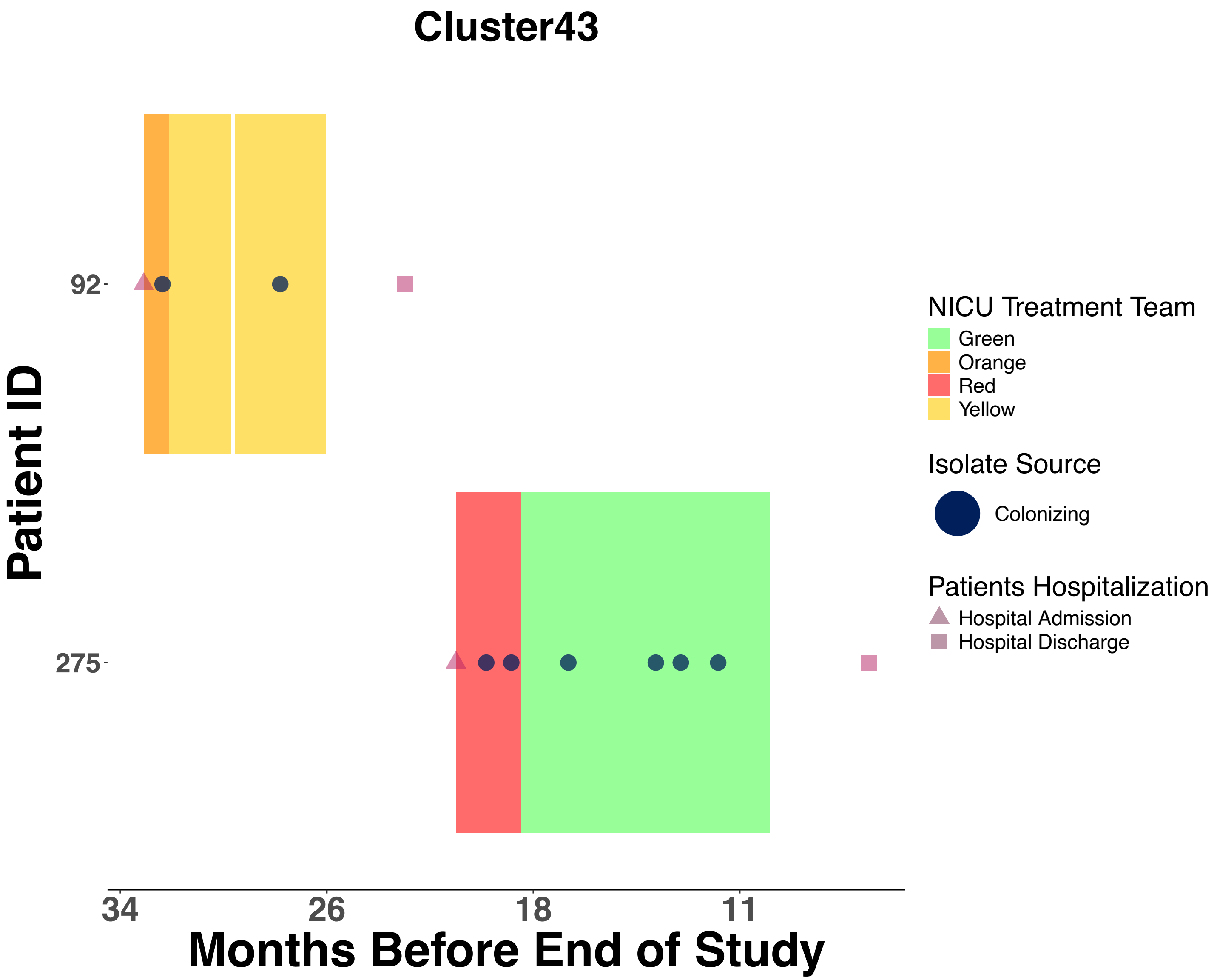

Cluster43
Patient ID
92
275
34
26
18
11
Months Before End of Study
NICU Treatment Team
Green
Orange
Red
Yellow
Isolate Source
Colonizing
Patients Hospitalization
Hospital Admission
Hospital Discharge

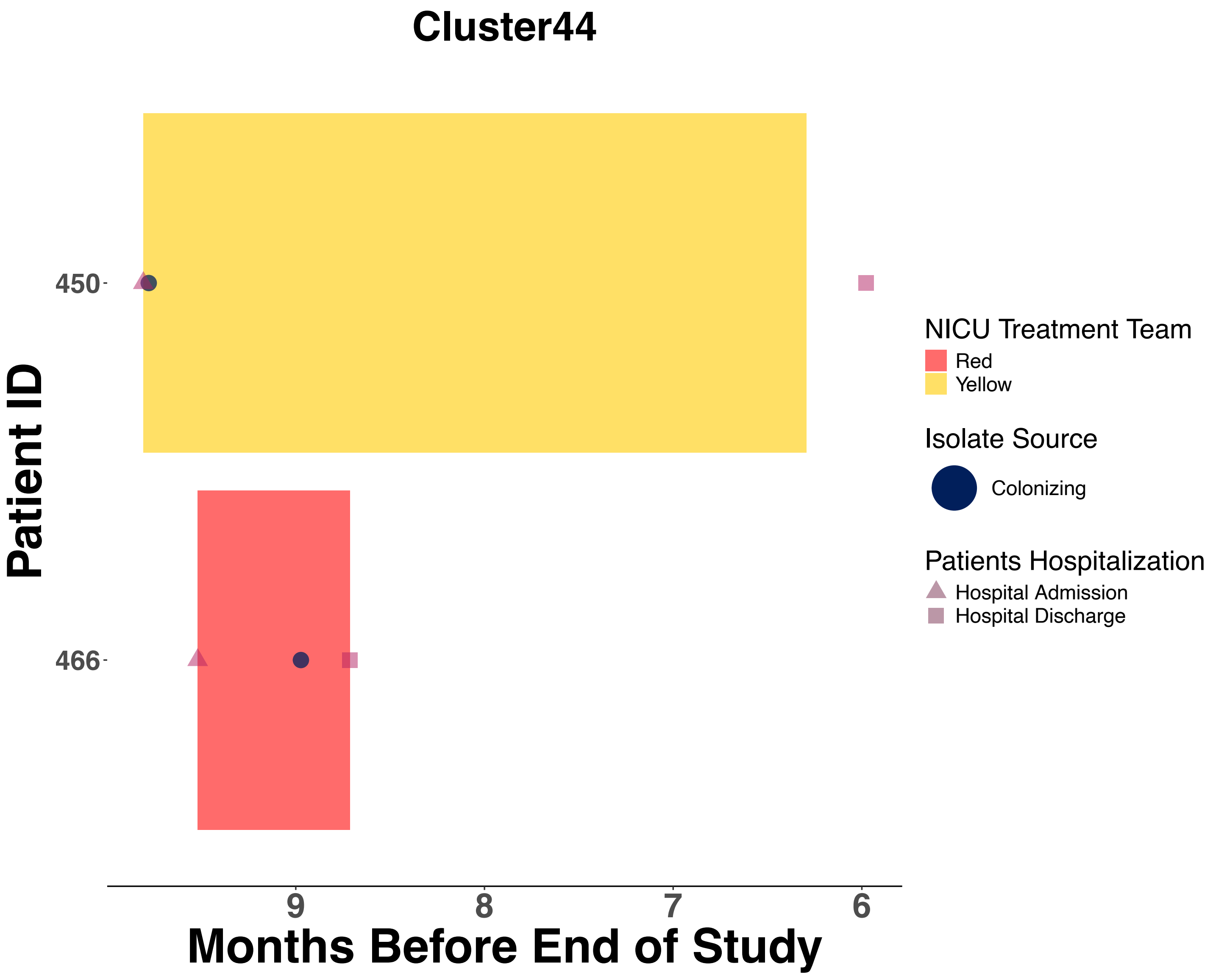

Cluster44
450
466
Patient ID
9
8
7
6
Months Before End of Study
NICU Treatment Team
Red
Yellow
Isolate Source
Colonizing
Patients Hospitalization
Hospital Admission
Hospital Discharge

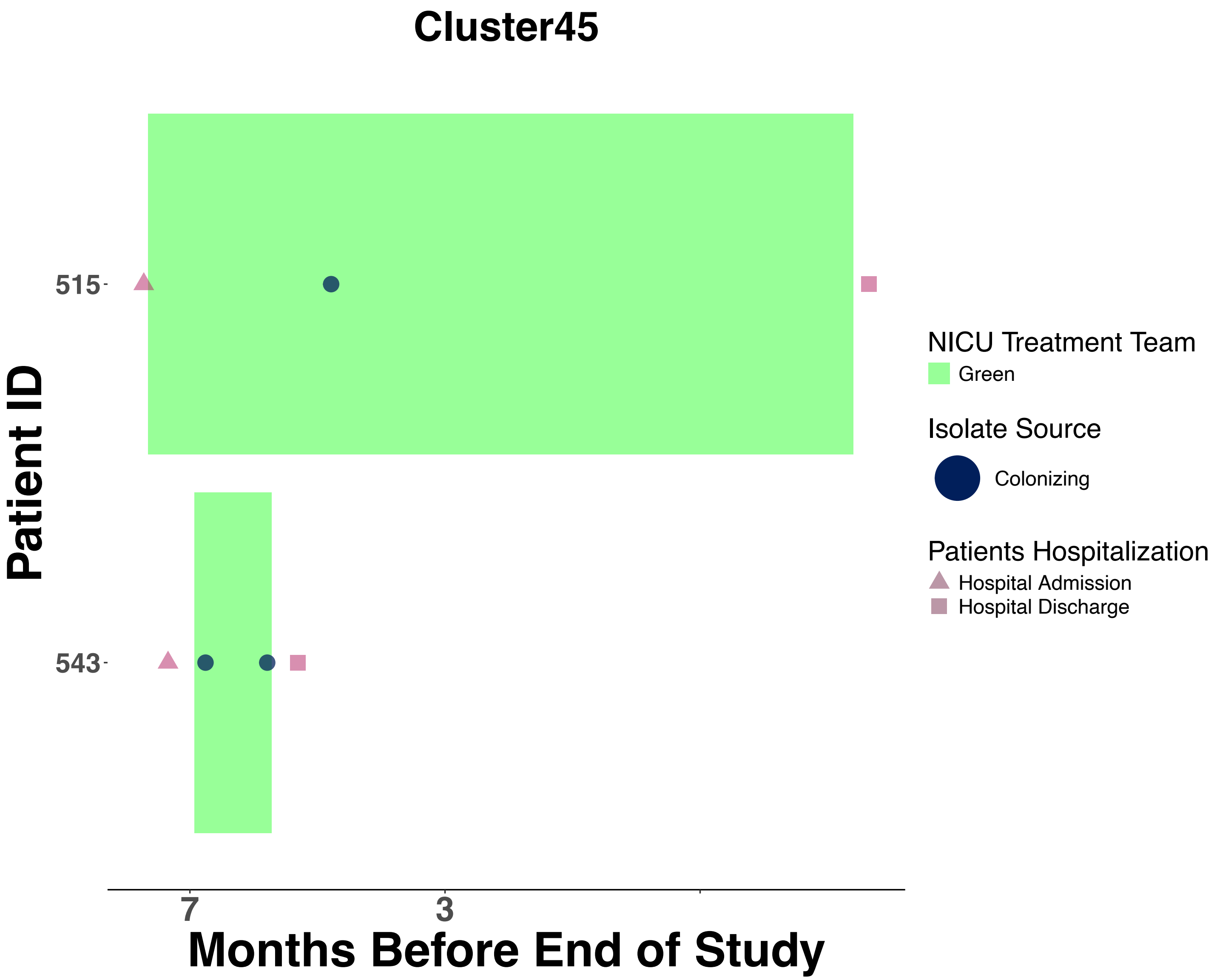
Cluster45
Patient ID
515
543
7
3
Months Before End of Study
NICU Treatment Team
Green
Isolate Source
Colonizing
Patients Hospitalization
Hospital Admission
Hospital Discharge

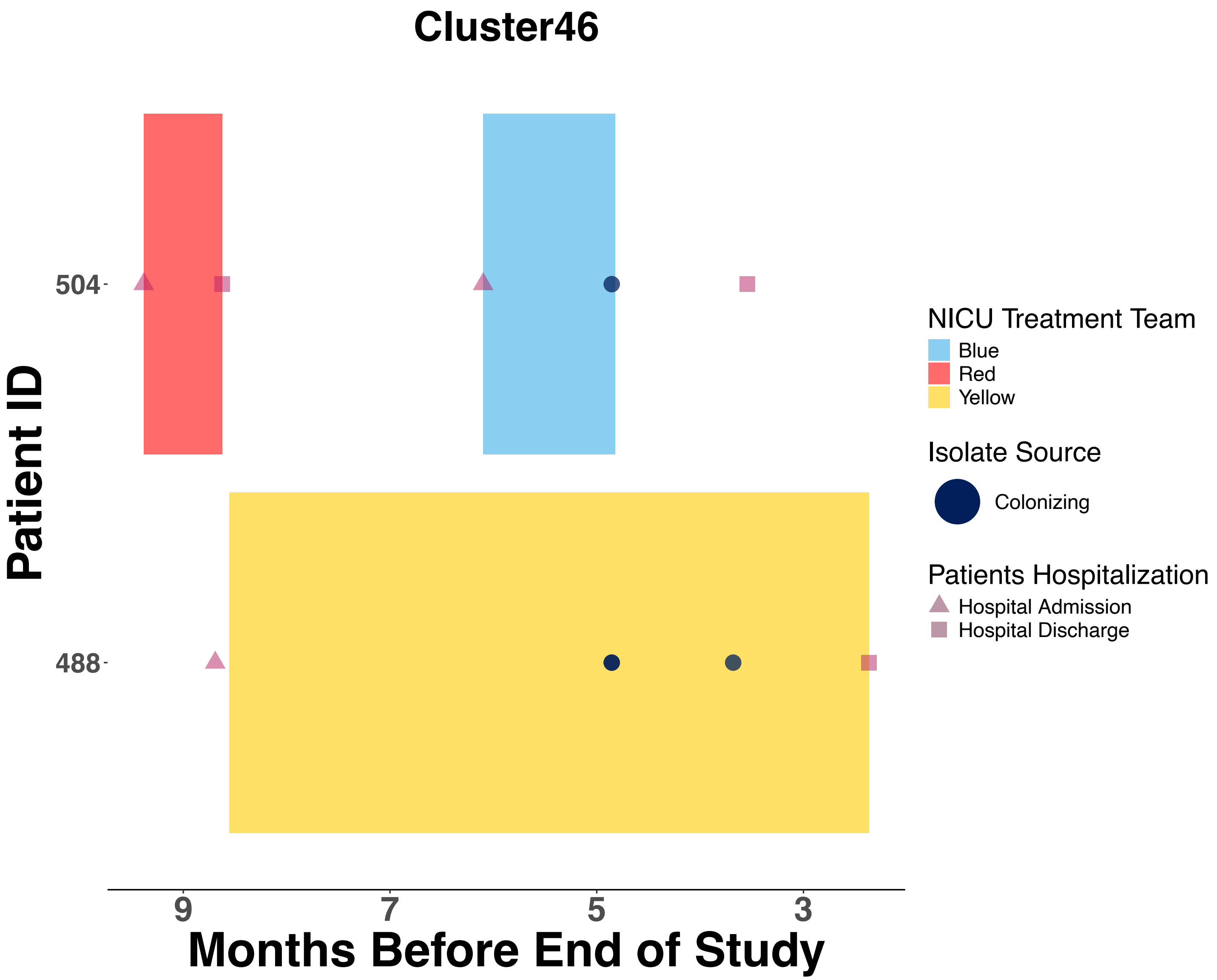

Cluster46
504
488
Patient ID
9
7
5
3
Months Before End of Study
NICU Treatment Team
Blue
Red
Yellow
Isolate Source
Colonizing
Patients Hospitalization
Hospital Admission
Hospital Discharge

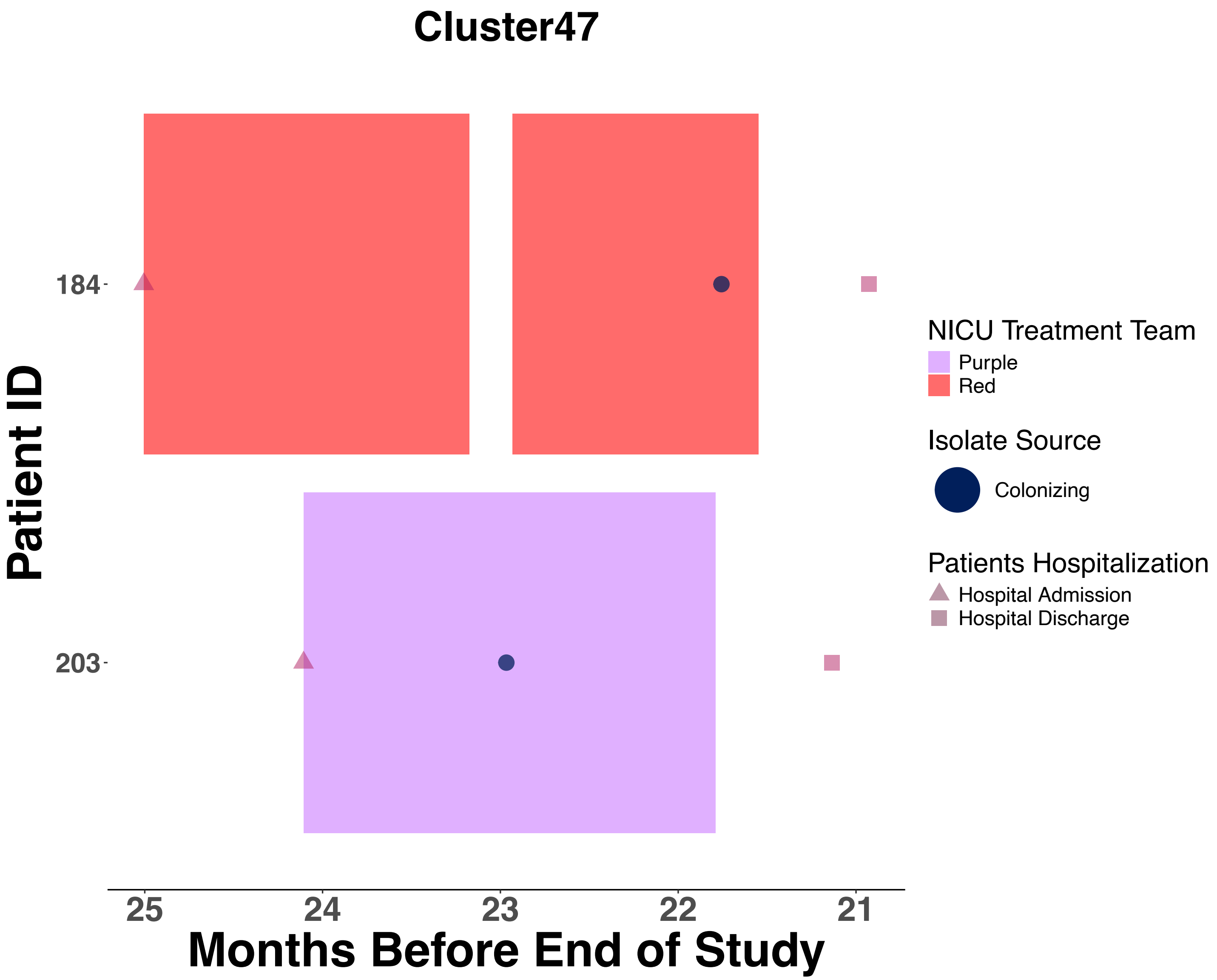
Cluster47
Patient ID
184
203
25
24
23
22
21
Months Before End of Study
NICU Treatment Team
Purple
Red
Isolate Source
Colonizing
Patients Hospitalization
Hospital Admission
Hospital Discharge

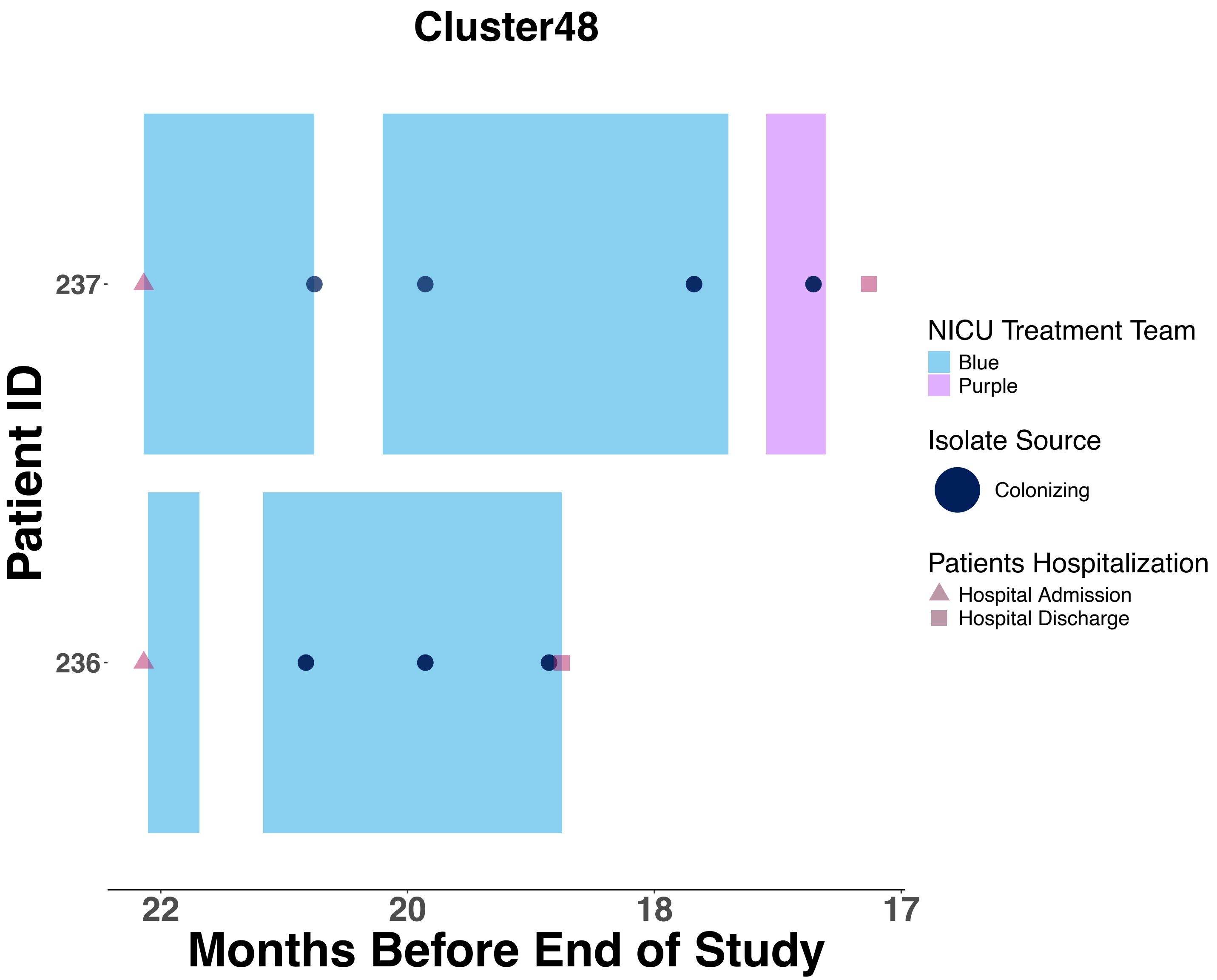
Cluster48
Patient ID
237
236
22
20
18
17
Months Before End of Study
NICU Treatment Team
Blue
Purple
Isolate Source
Colonizing
Patients Hospitalization
Hospital Admission
Hospital Discharge

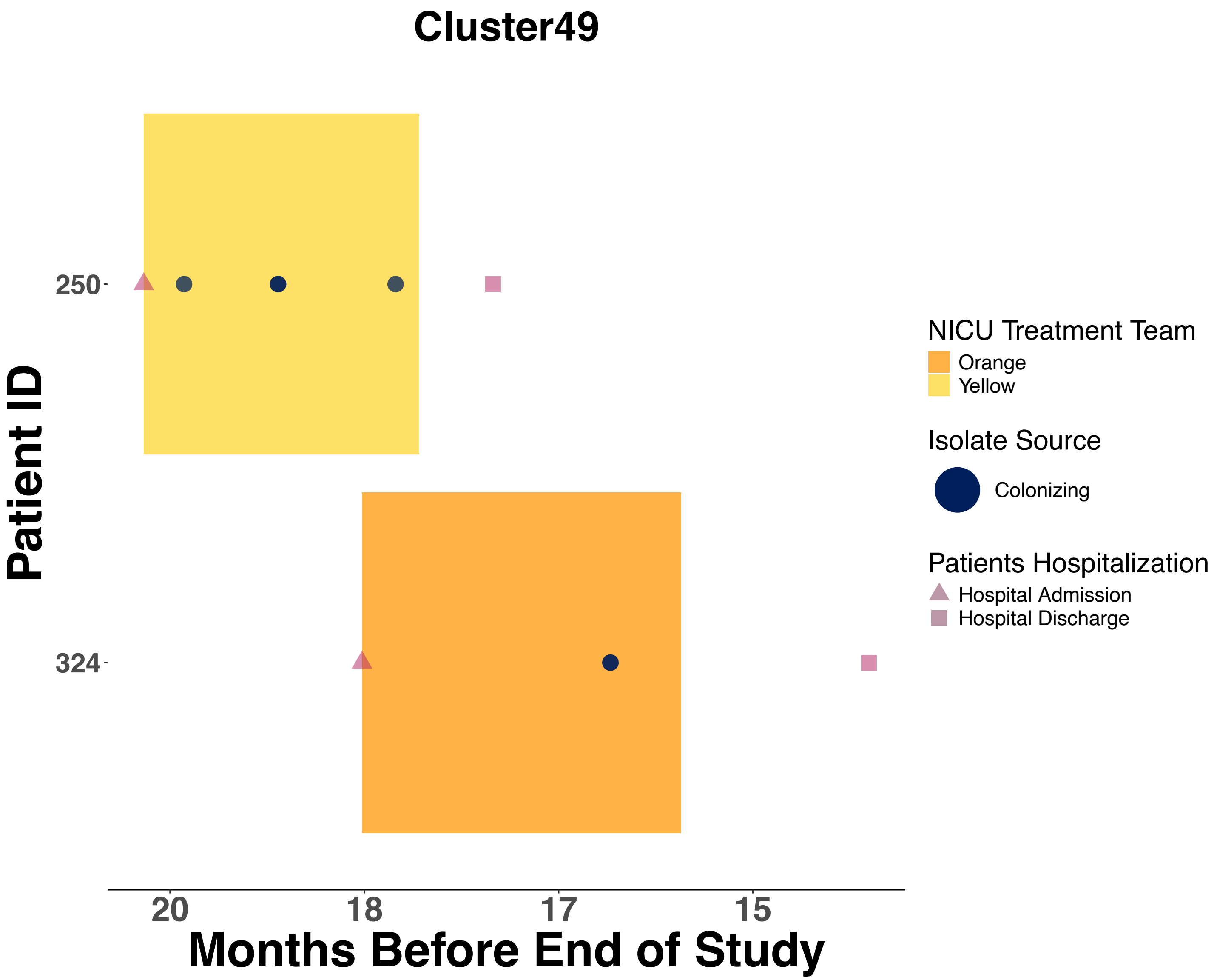

Cluster49
Patient ID
250
324
20
18
17
15
Months Before End of Study
NICU Treatment Team
Orange
Yellow
Isolate Source
Colonizing
Patients Hospitalization
Hospital Admission
Hospital Discharge

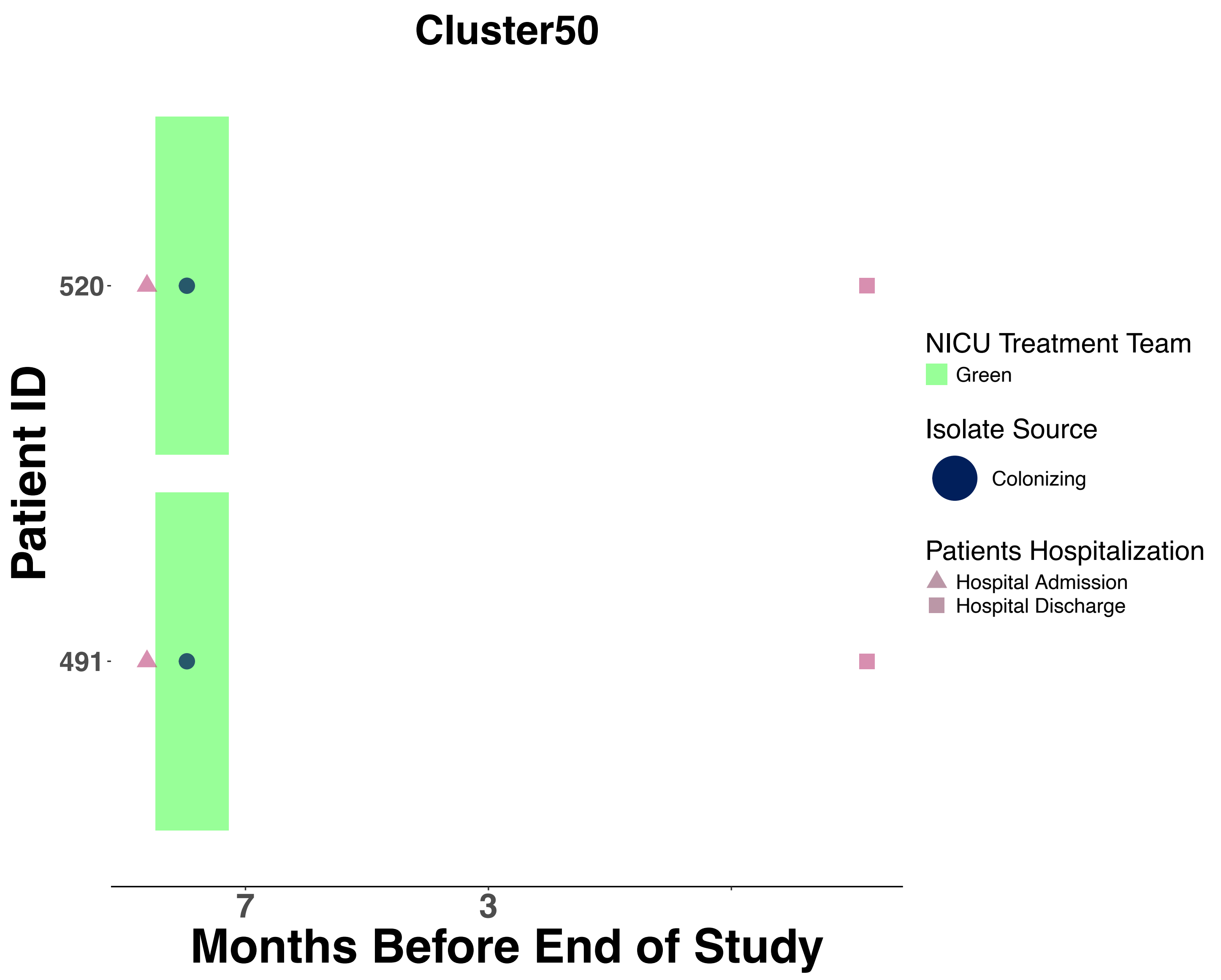
Cluster50
Patient ID
520
491
7
3
Months Before End of Study
NICU Treatment Team
Green
Isolate Source
Colonizing
Patients Hospitalization
Hospital Admission
Hospital Discharge

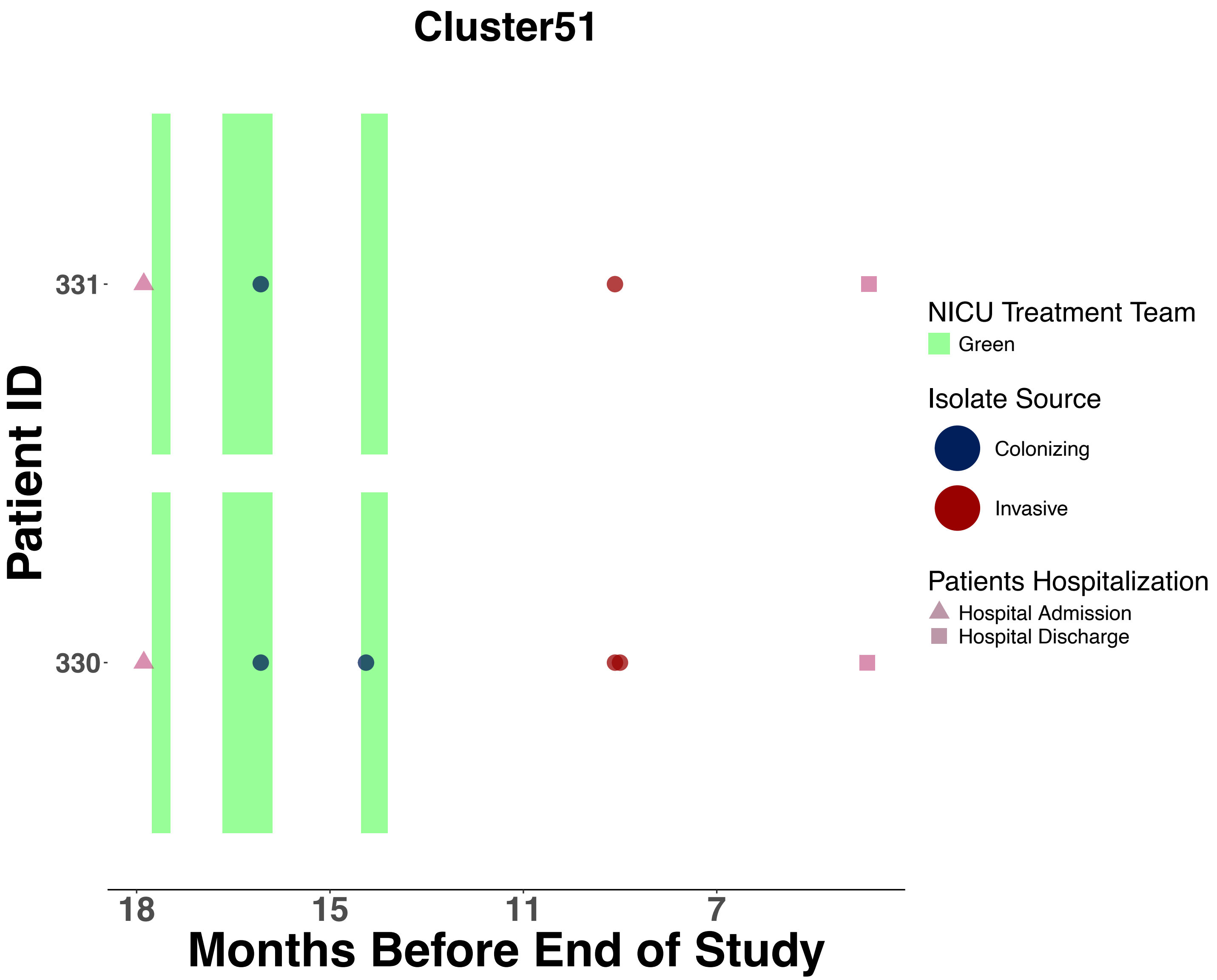

Cluster51
Patient ID
331
330
18
15
11
7
Months Before End of Study
NICU Treatment Team
Green
Isolate Source
Colonizing
Invasive
Patients Hospitalization
Hospital Admission
Hospital Discharge

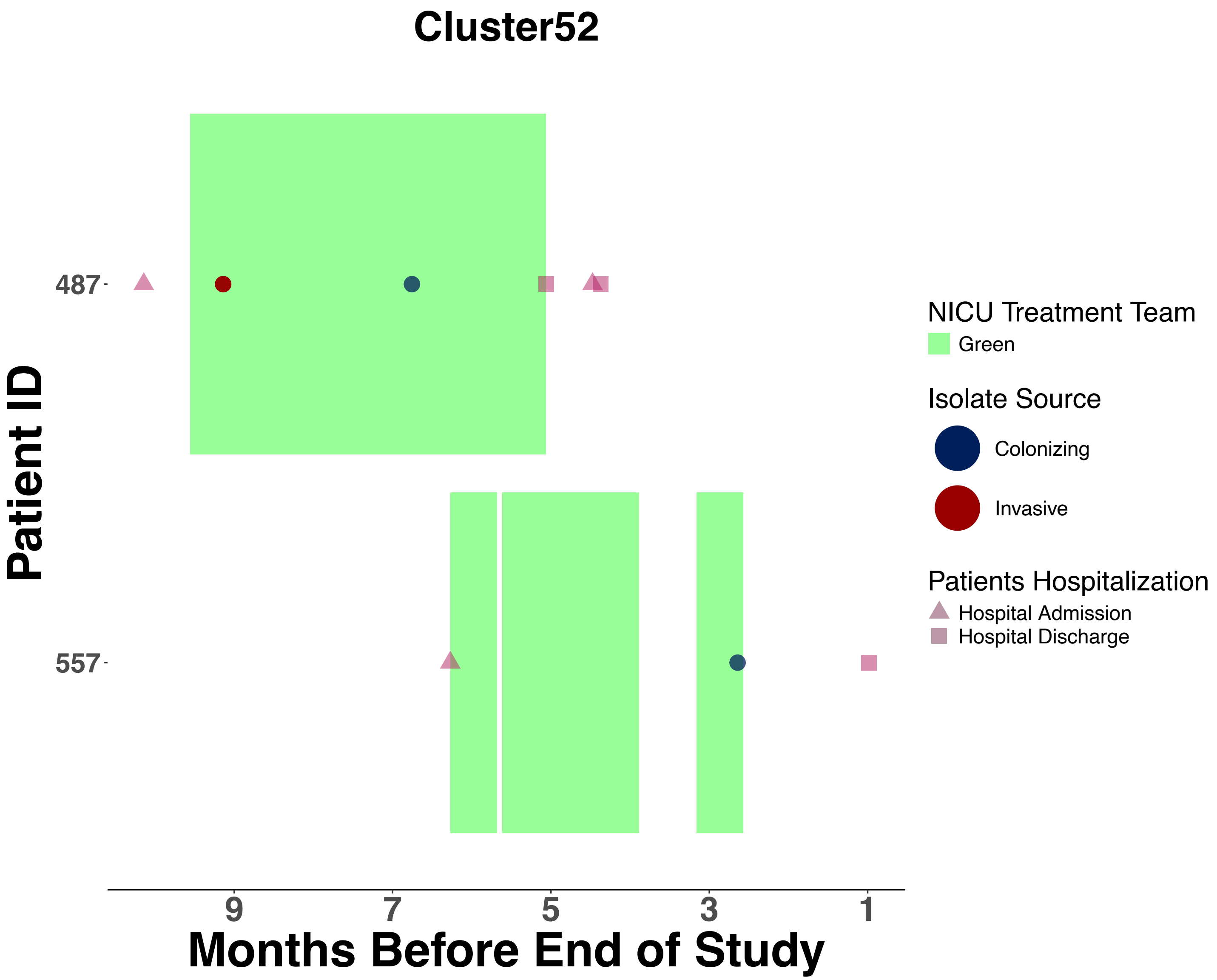
Cluster52
Patient ID
487
557
9
7
5
3
1
Months Before End of Study
NICU Treatment Team
Green
Isolate Source
Colonizing
Invasive
Patients Hospitalization
Hospital Admission
Hospital Discharge

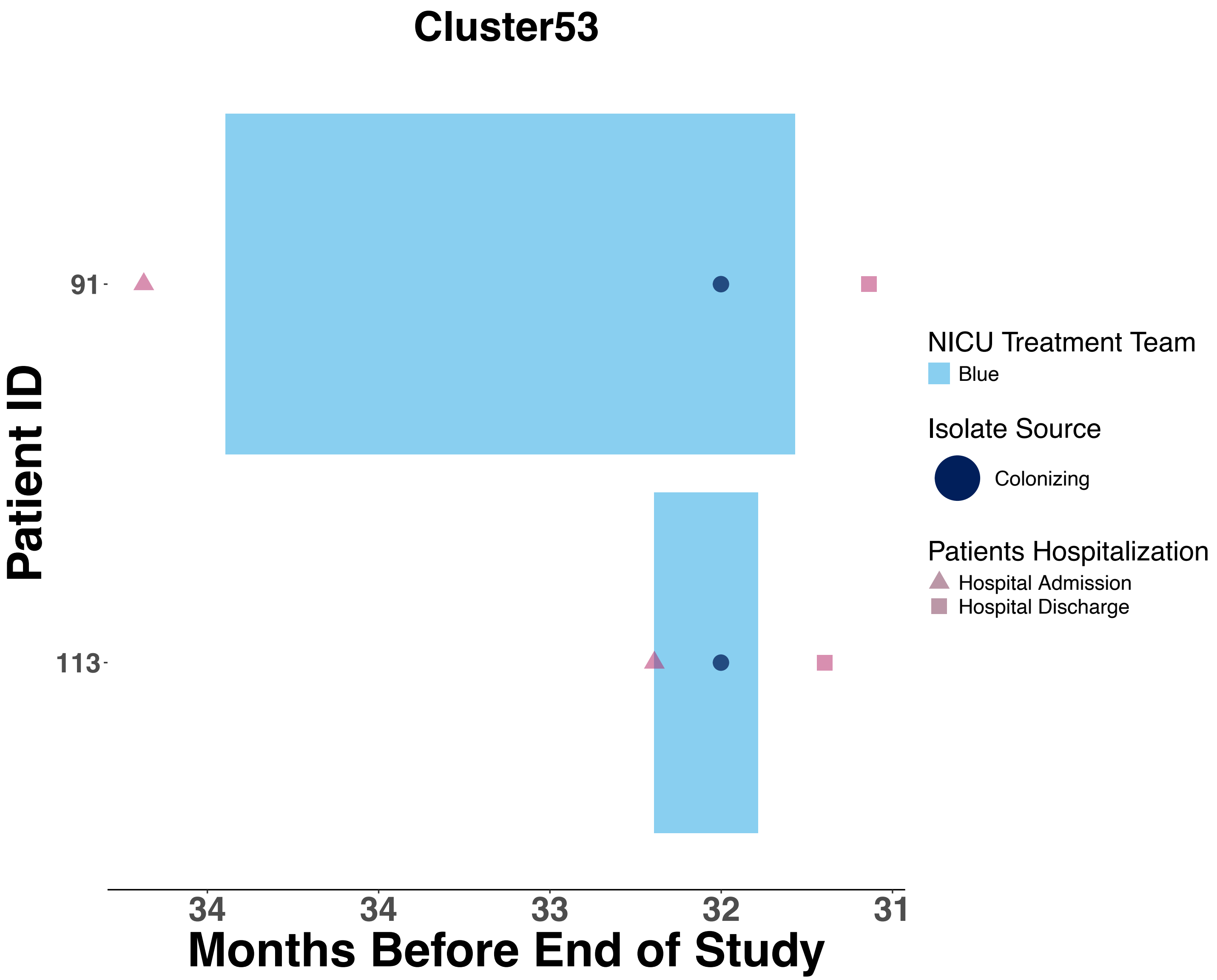

Cluster53
Patient ID
91
113
34
34
33
32
31
Months Before End of Study
NICU Treatment Team
Blue
Isolate Source
Colonizing
Patients Hospitalization
Hospital Admission
Hospital Discharge

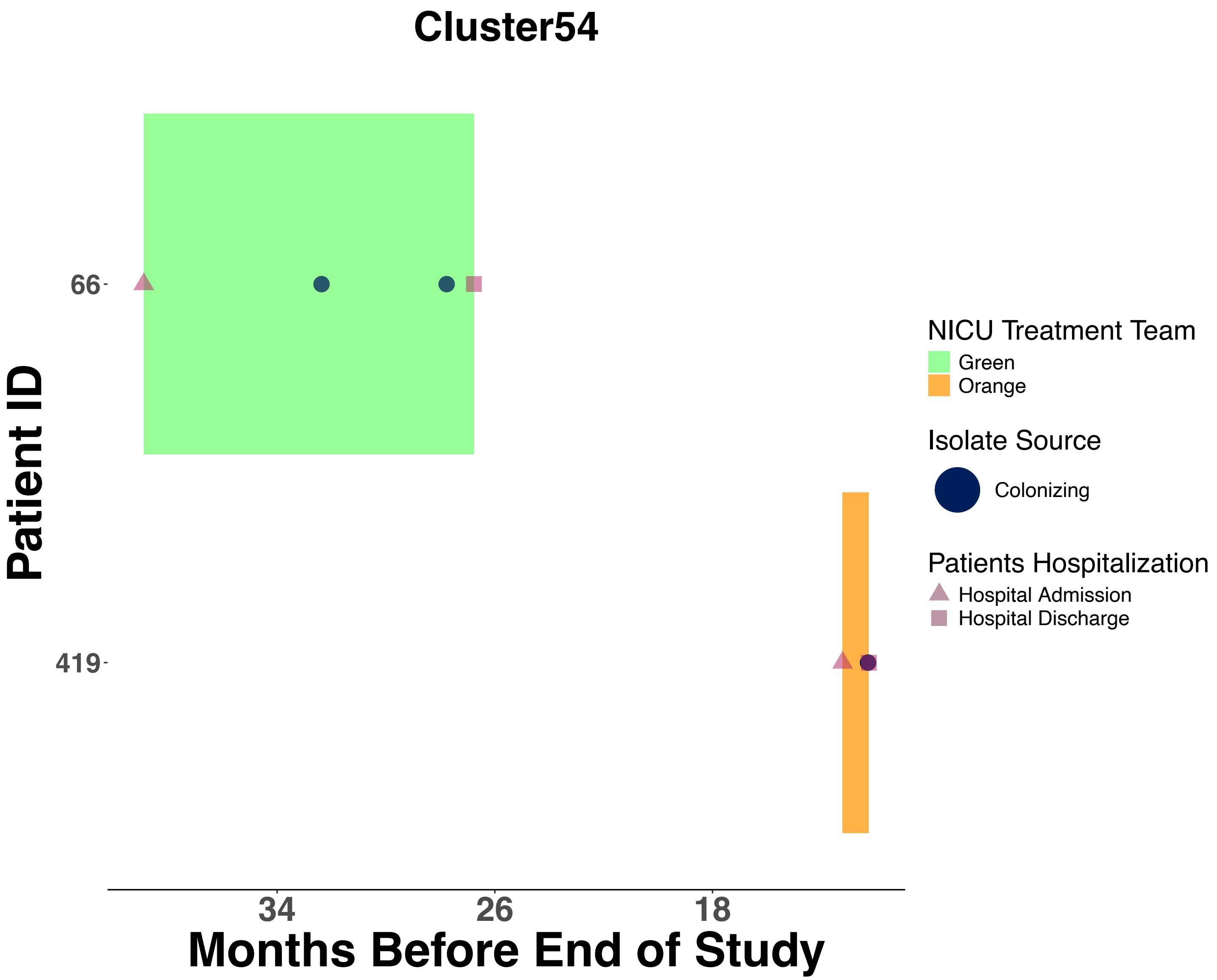
Cluster54
Patient ID
66
419
34
26
18
Months Before End of Study
NICU Treatment Team
Green
Orange
Isolate Source
Colonizing
Patients Hospitalization
Hospital Admission
Hospital Discharge

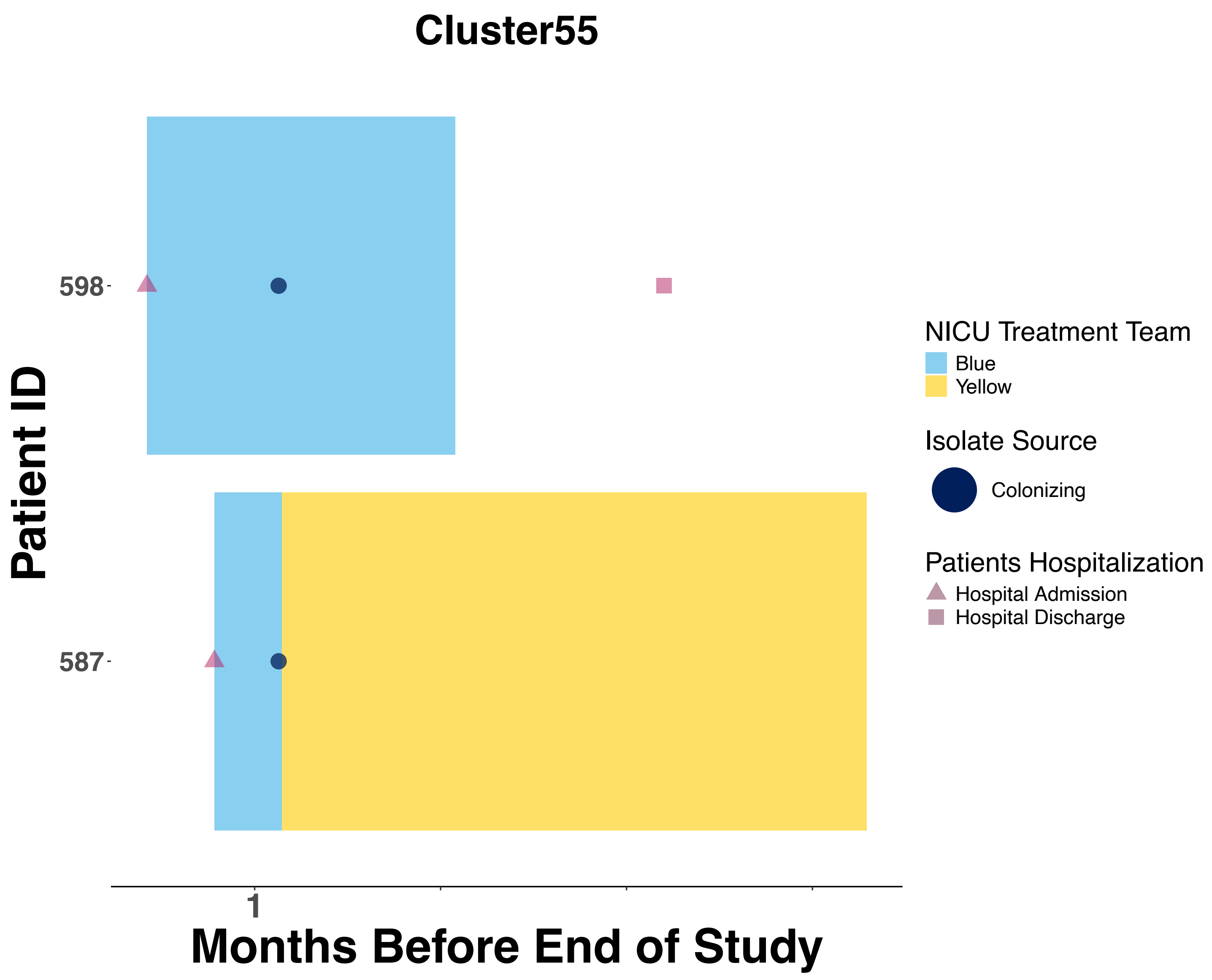

Cluster55
Patient ID
598
587
1
Months Before End of Study
NICU Treatment Team
Blue
Yellow
Isolate Source
Colonizing
Patients Hospitalization
Hospital Admission
Hospital Discharge

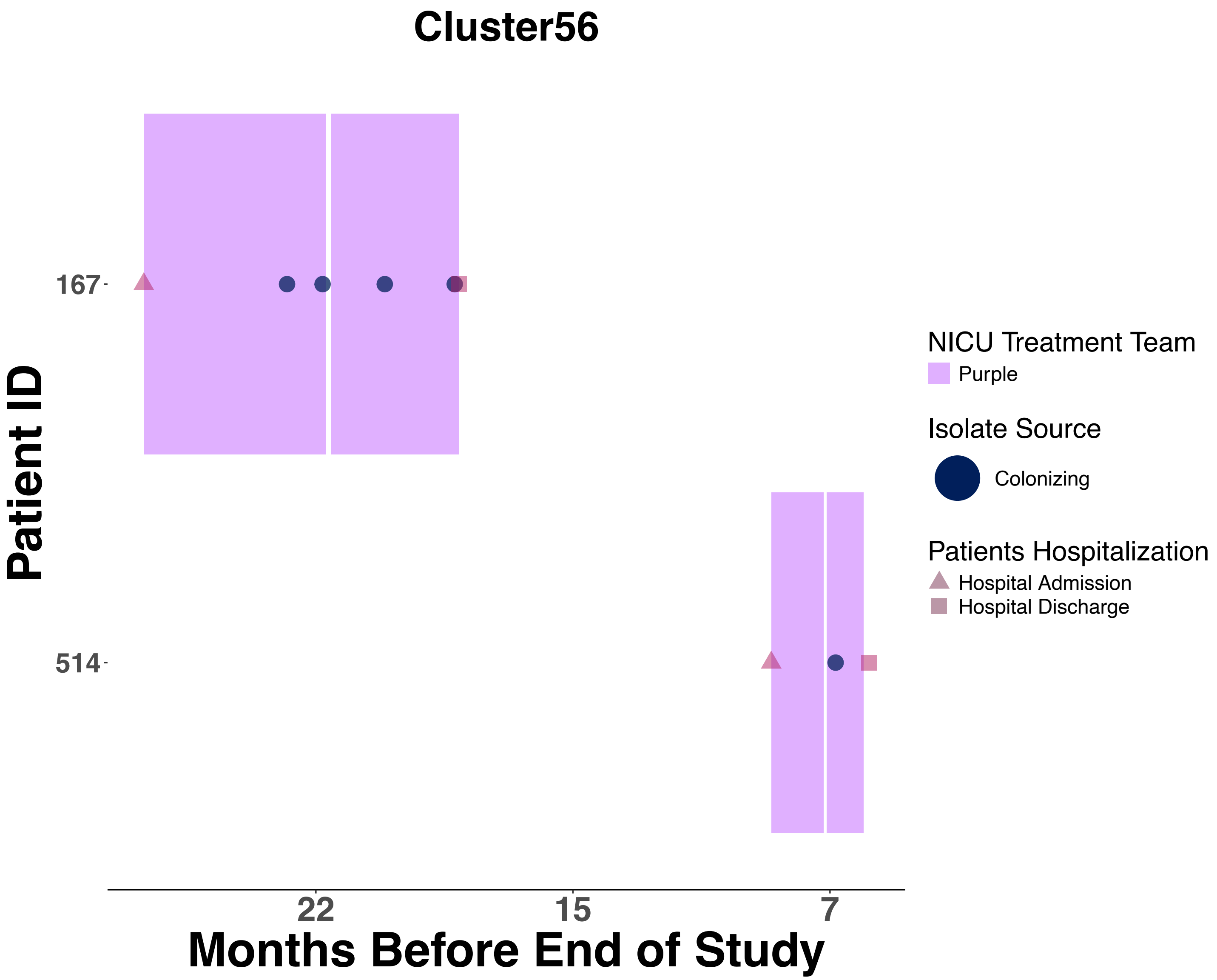

Cluster56
Patient ID
167
514
22
15
7
Months Before End of Study
NICU Treatment Team
Purple
Isolate Source
Colonizing
Patients Hospitalization
Hospital Admission
Hospital Discharge

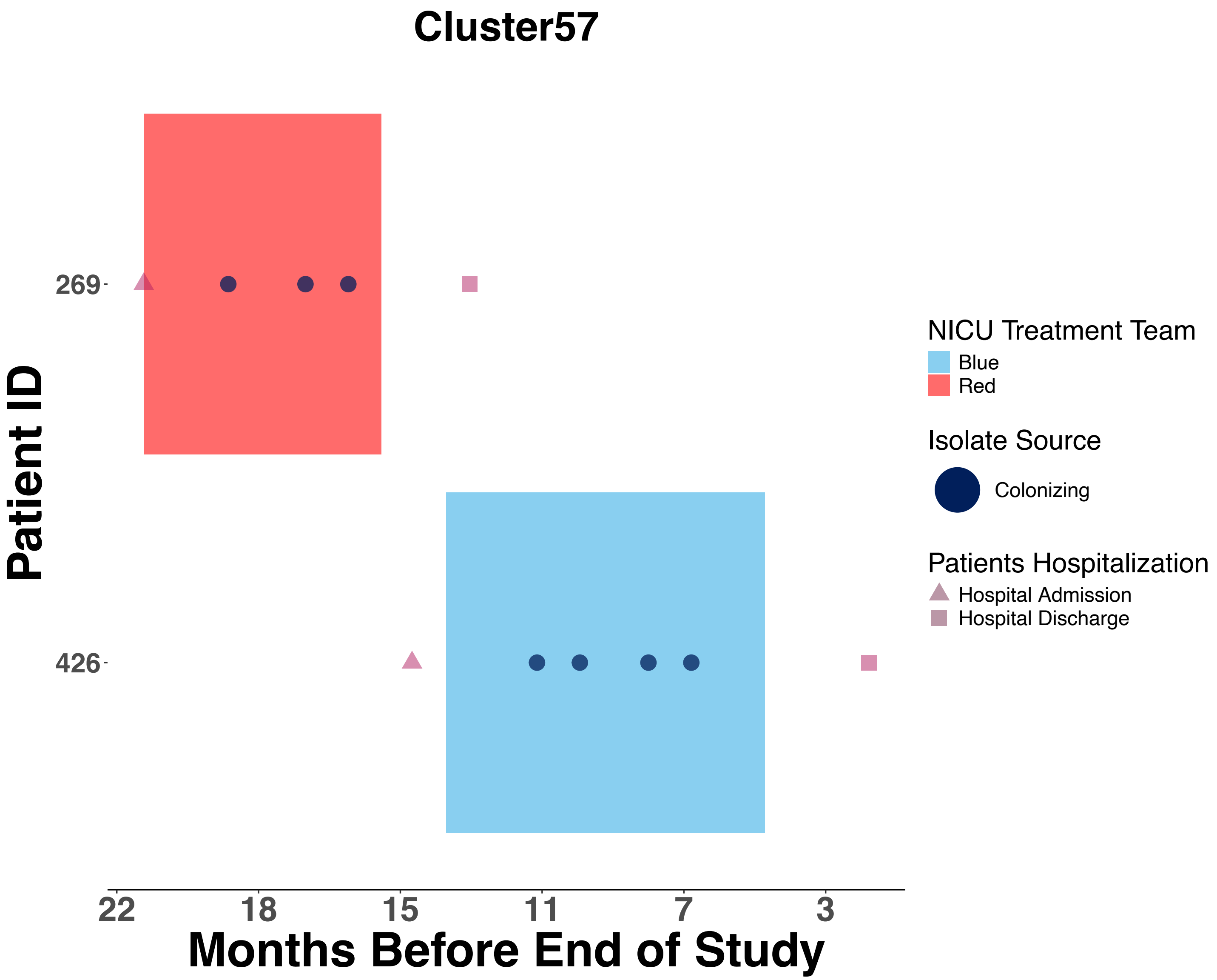

Cluster57
Patient ID
269
426
Months Before End of Study
22
18
15
11
7
3
NICU Treatment Team
Blue
Red
Isolate Source
Colonizing
Patients Hospitalization
Hospital Admission
Hospital Discharge

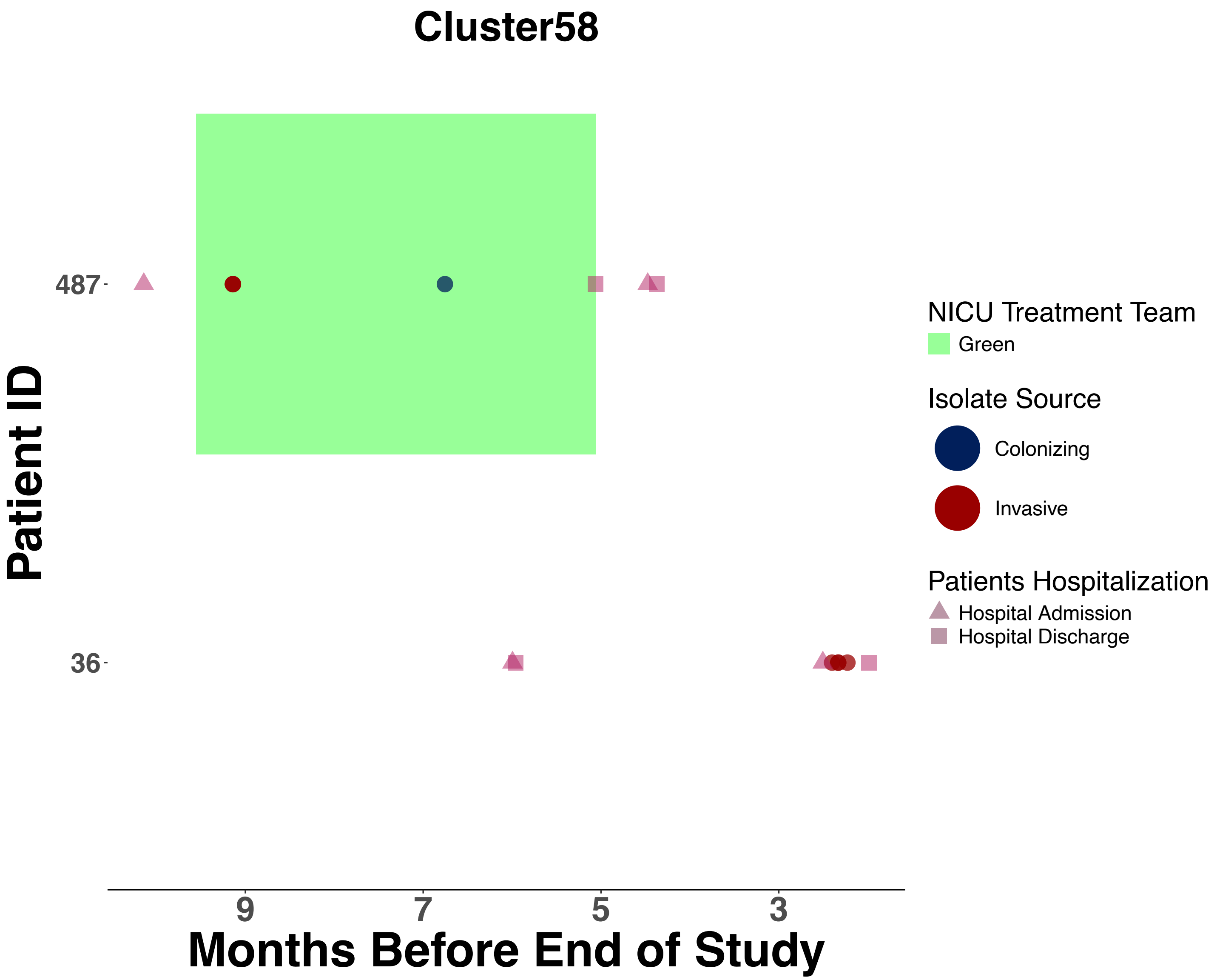

Cluster58
Patient ID
487
36
9
7
5
3
Months Before End of Study
NICU Treatment Team
Green
Isolate Source
Colonizing
Invasive
Patients Hospitalization
Hospital Admission
Hospital Discharge

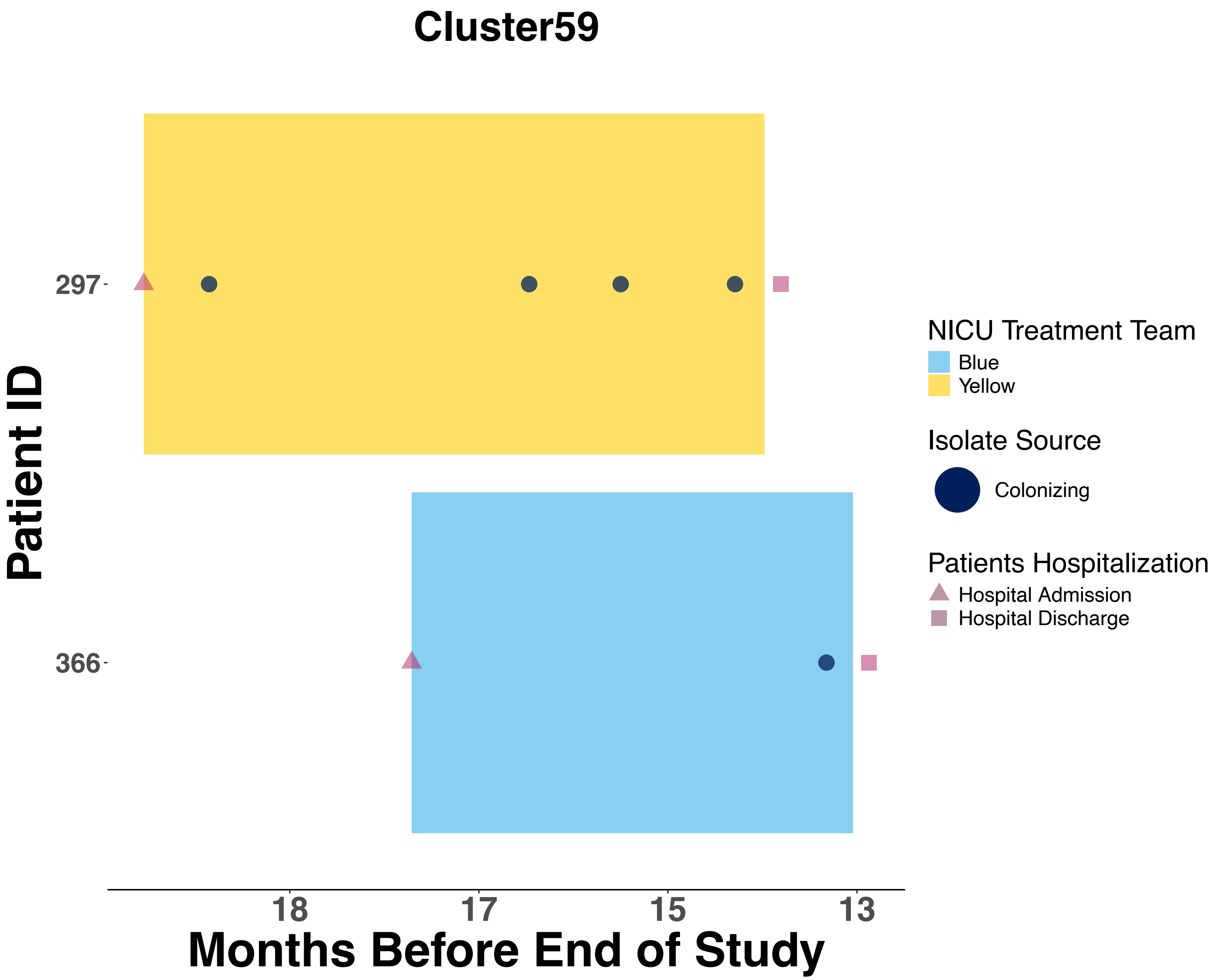

Cluster59
Patient ID
297
366
18
17
15
13
Months Before End of Study
NICU Treatment Team
Blue
Yellow
Isolate Source
Colonizing
Patients Hospitalization
Hospital Admission
Hospital Discharge

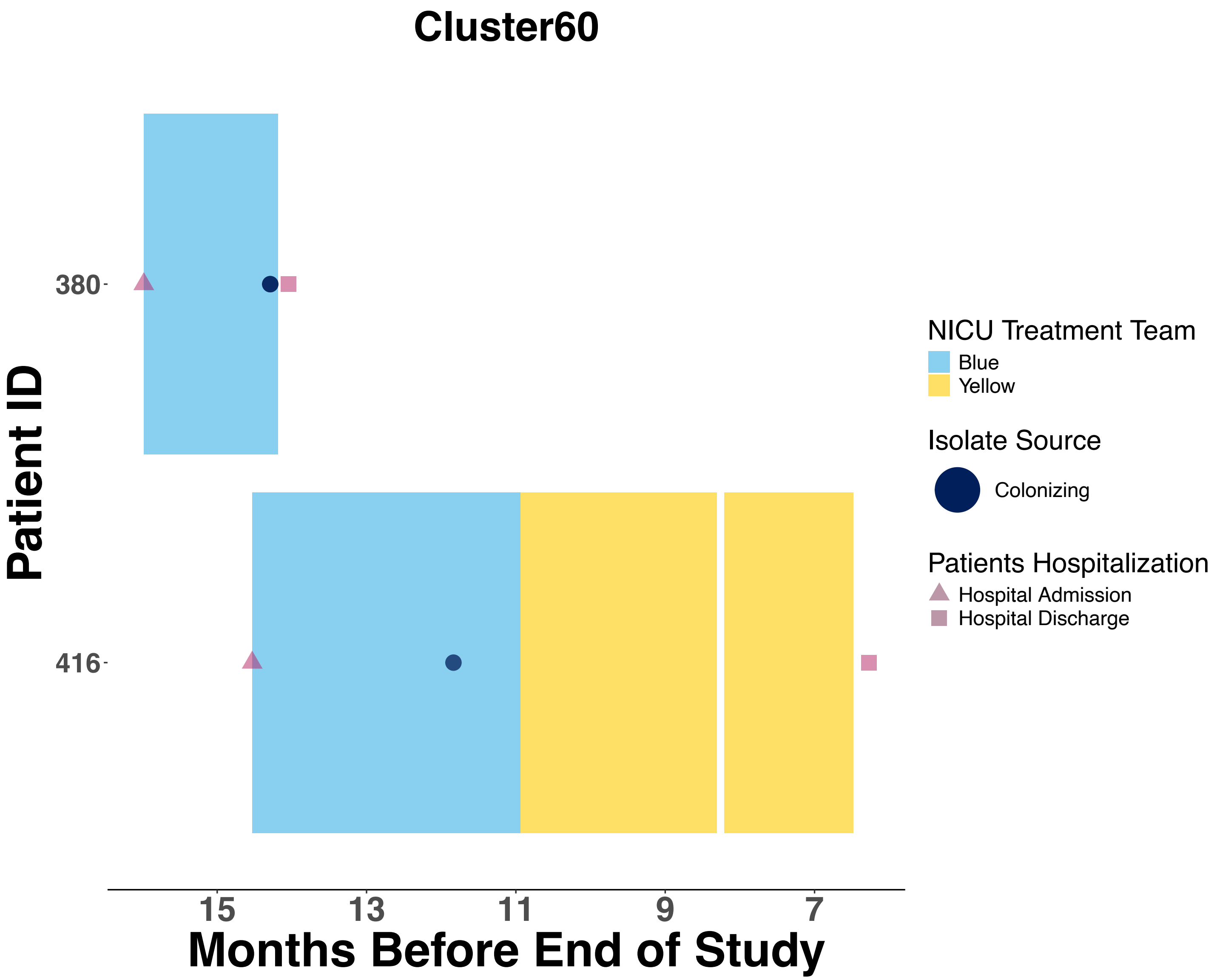

Cluster60
Patient ID
380
416
15
13
11
9
7
Months Before End of Study
NICU Treatment Team
Blue
Yellow
Isolate Source
Colonizing
Patients Hospitalization
Hospital Admission
Hospital Discharge

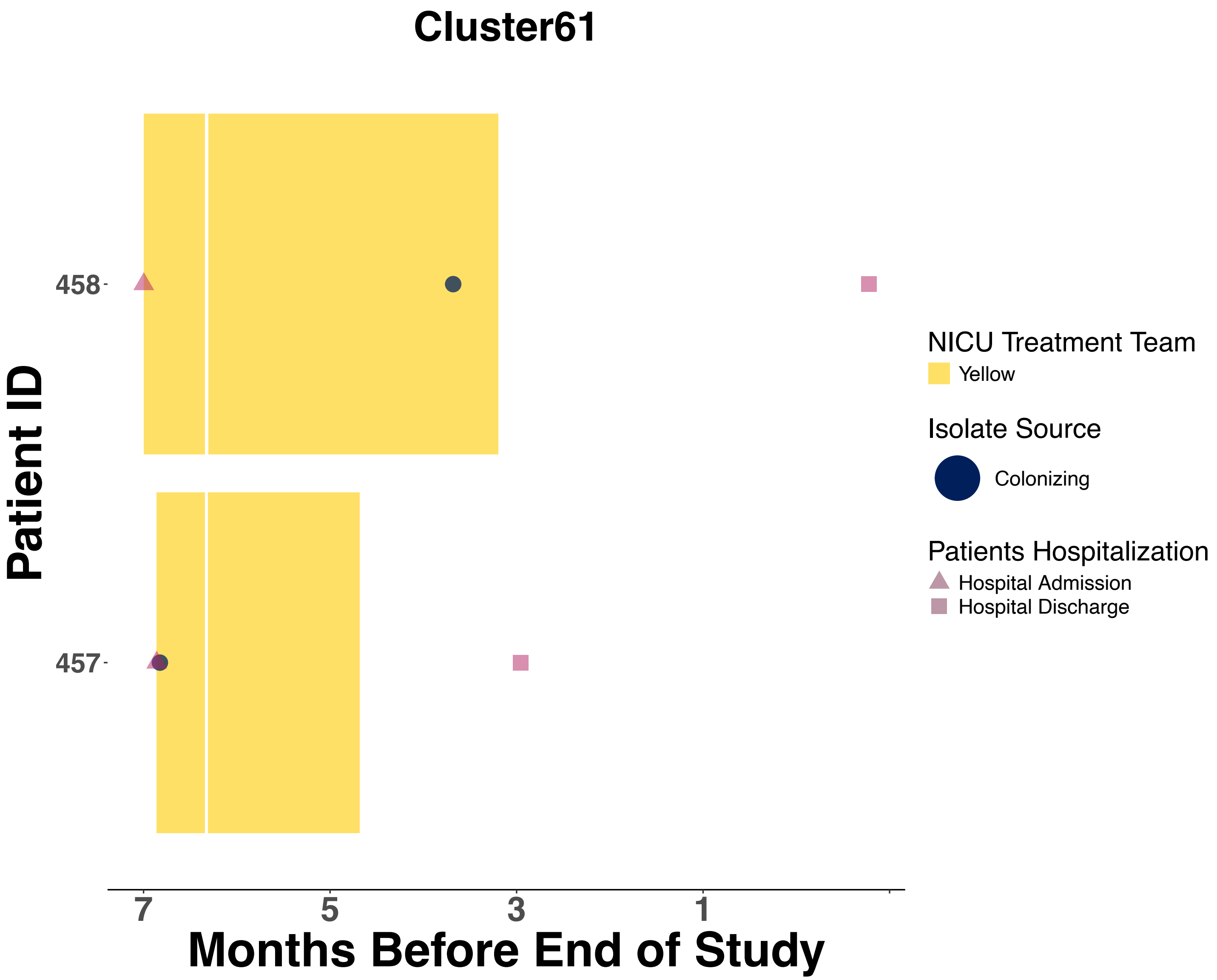

Cluster61
Patient ID
458
457
7
5
3
1
Months Before End of Study
NICU Treatment Team
Yellow
Isolate Source
Colonizing
Patients Hospitalization
Hospital Admission
Hospital Discharge

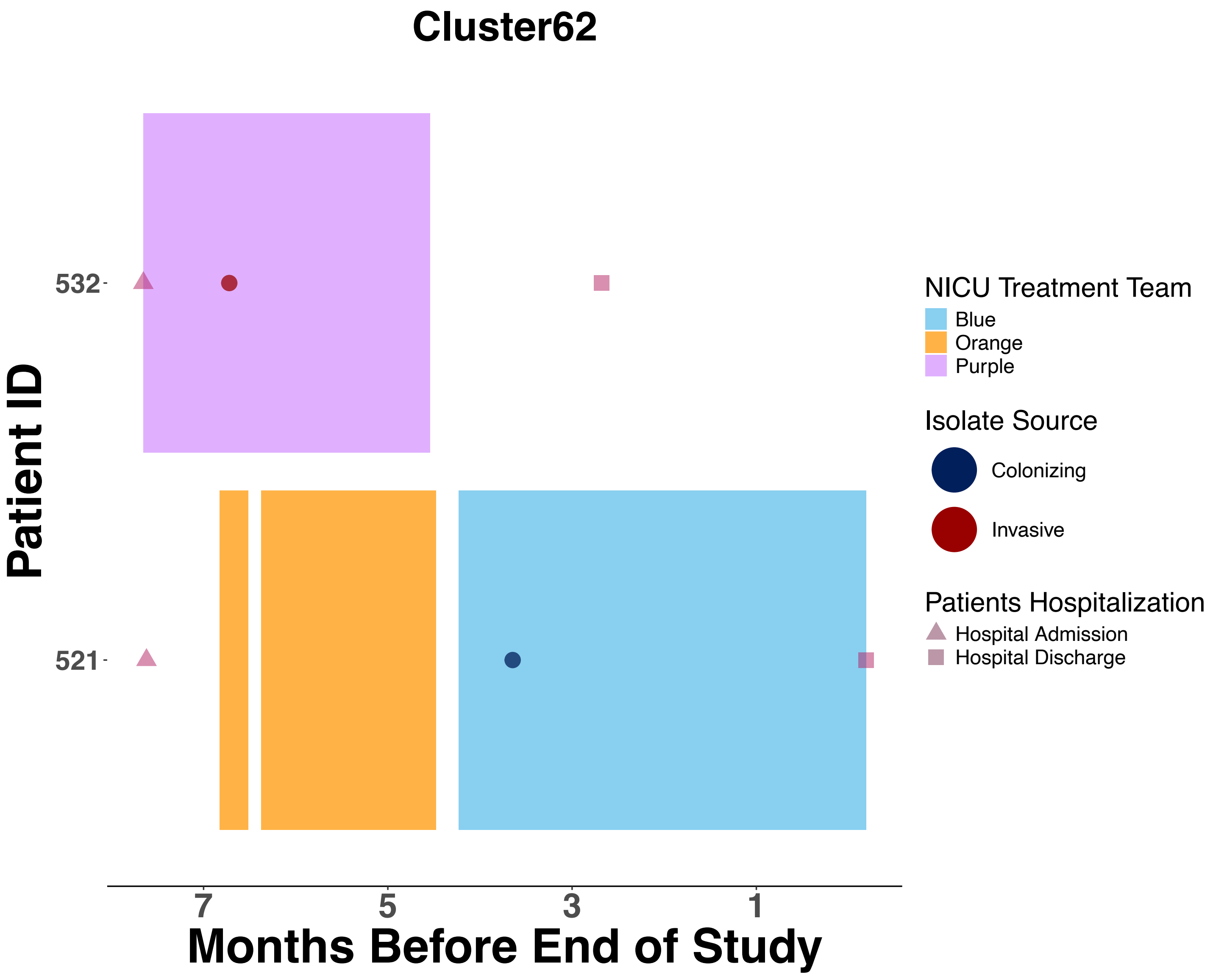

Cluster62
Patient ID
532
521
7
5
3
1
Months Before End of Study
NICU Treatment Team
Blue
Orange
Purple
Isolate Source
Colonizing
Invasive
Patients Hospitalization
Hospital Admission
Hospital Discharge

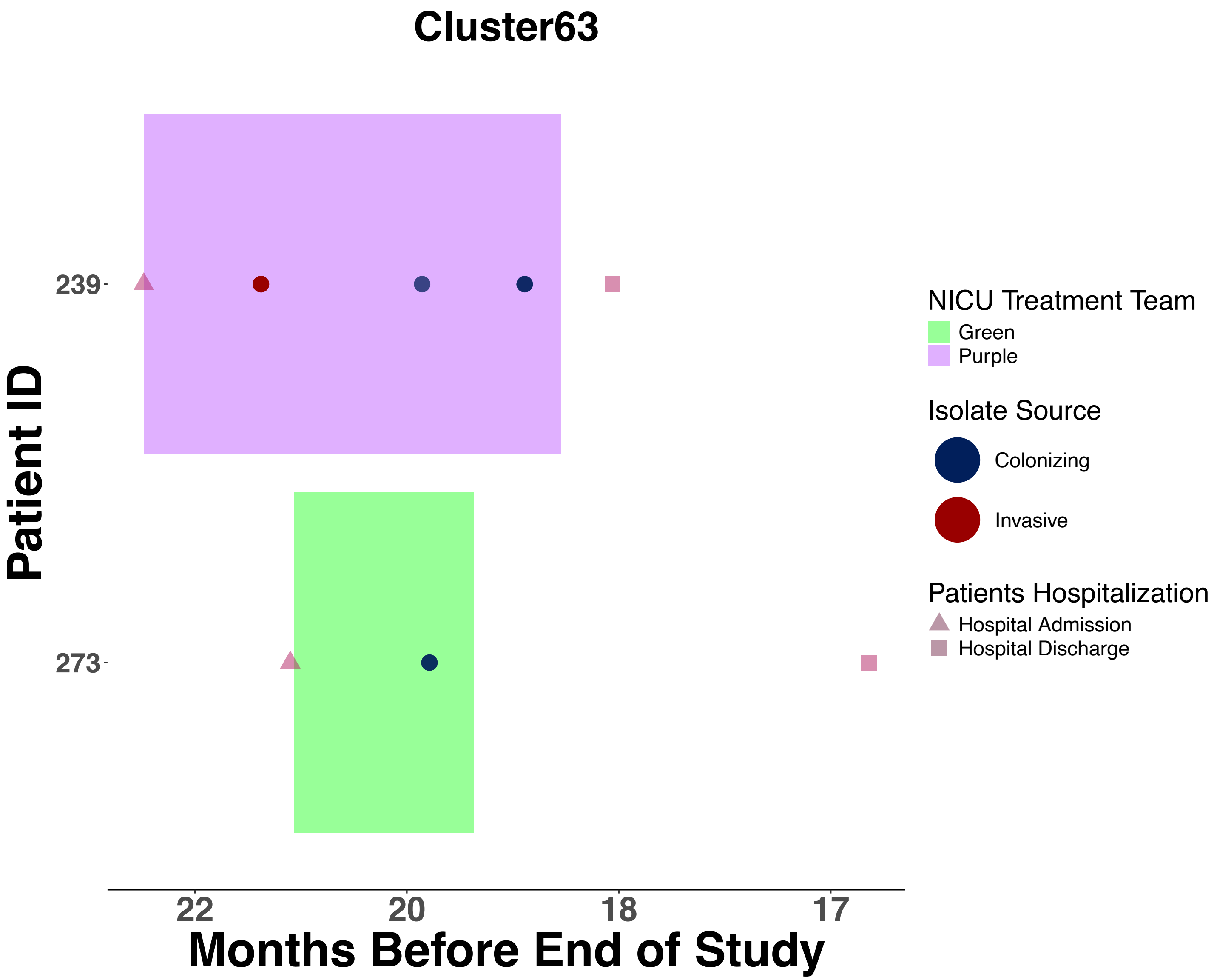

Cluster63
NICU Treatment Team
Green
Purple
Isolate Source
Colonizing
Invasive
Patients Hospitalization
Hospital Admission
Hospital Discharge
Patient ID
239
273
22
20
18
17
Months Before End of Study

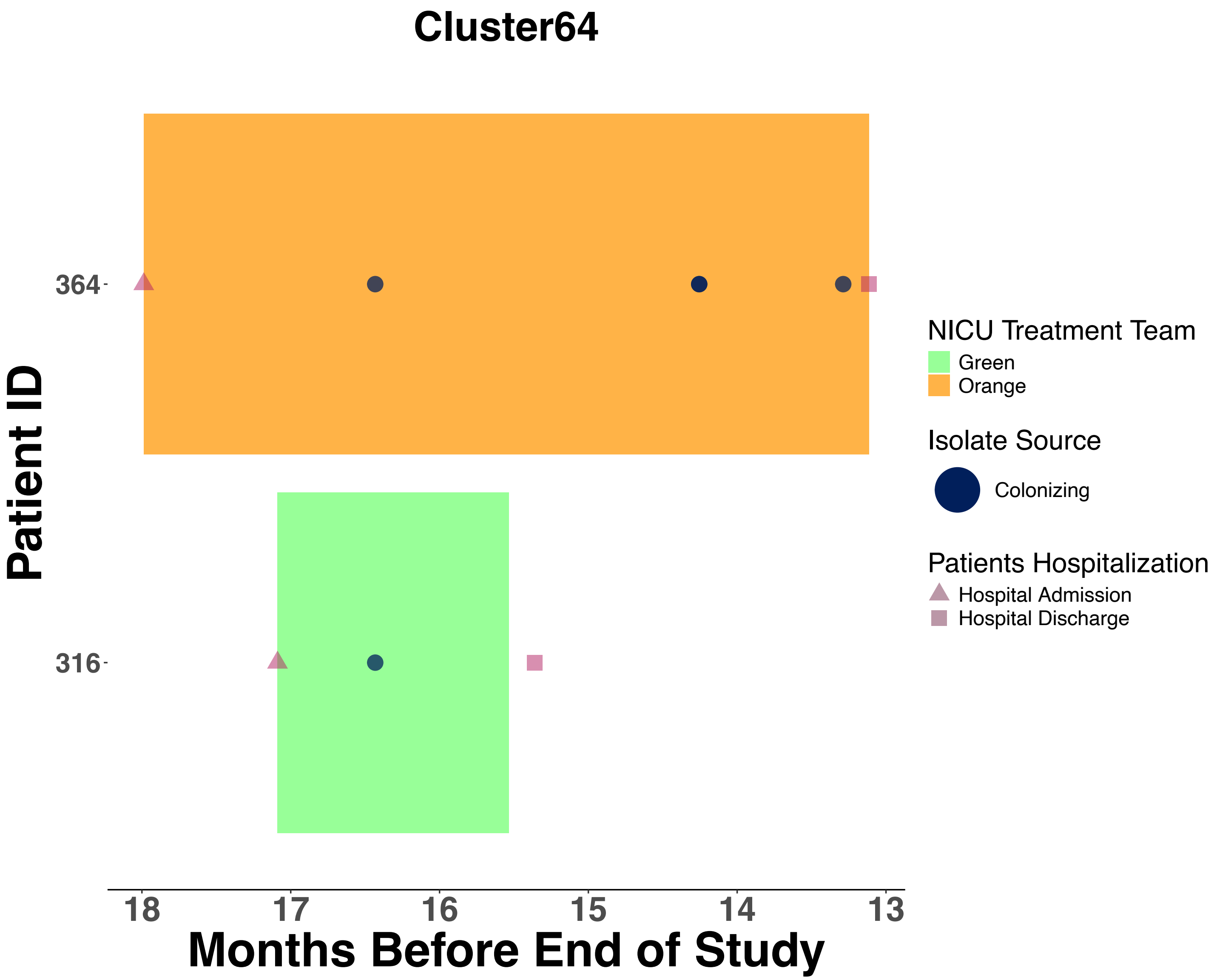
Cluster64
Patient ID
364
316
18
17
16
15
14
13
Months Before End of Study
NICU Treatment Team
Green
Orange
Isolate Source
Colonizing
Patients Hospitalization
Hospital Admission
Hospital Discharge

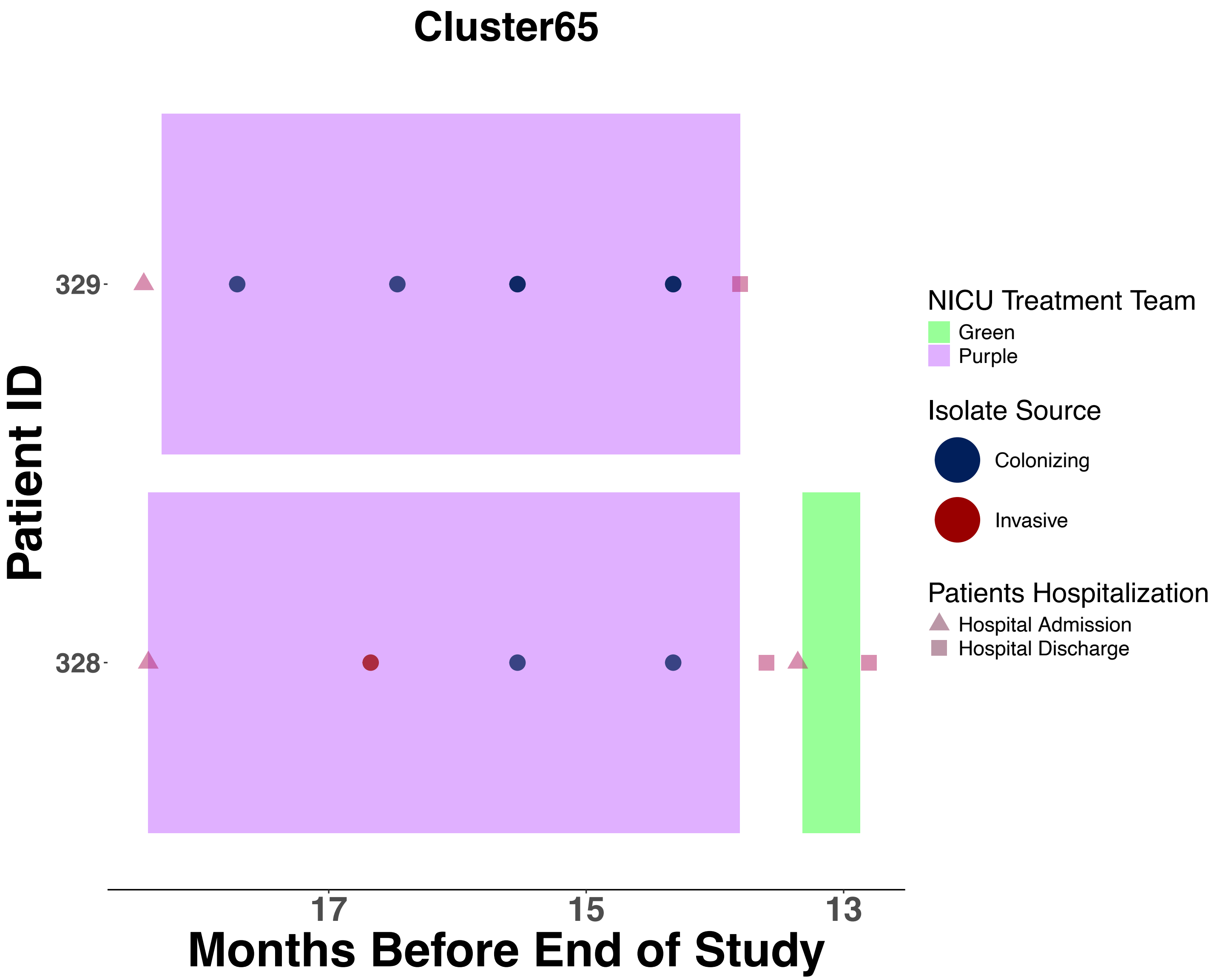
Cluster65
Patient ID
329
328
17
15
13
Months Before End of Study
NICU Treatment Team
Green
Purple
Isolate Source
Colonizing
Invasive
Patients Hospitalization
Hospital Admission
Hospital Discharge

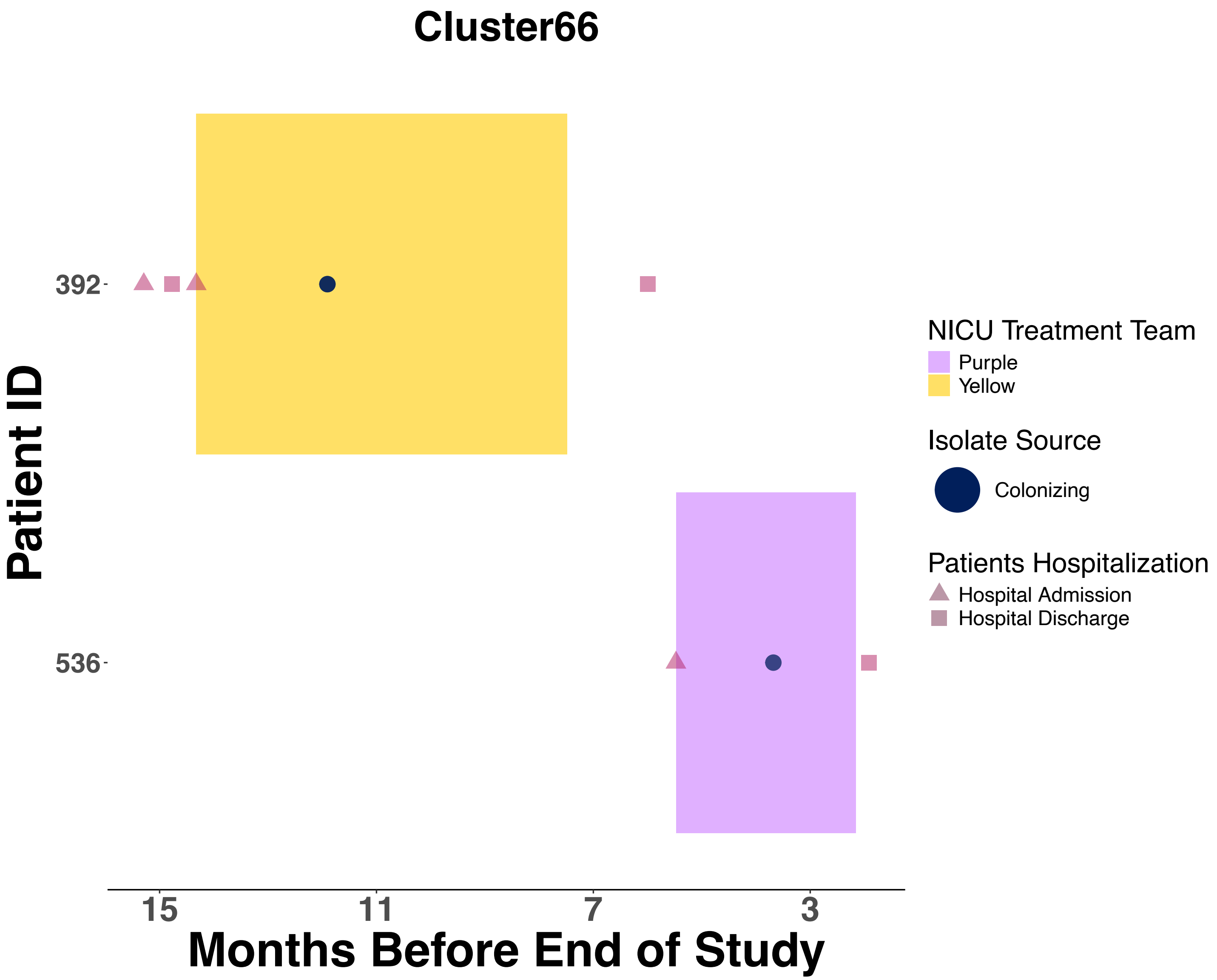

Cluster66
Patient ID
392
536
15
11
7
3
Months Before End of Study
NICU Treatment Team
Purple
Yellow
Isolate Source
Colonizing
Patients Hospitalization
Hospital Admission
Hospital Discharge

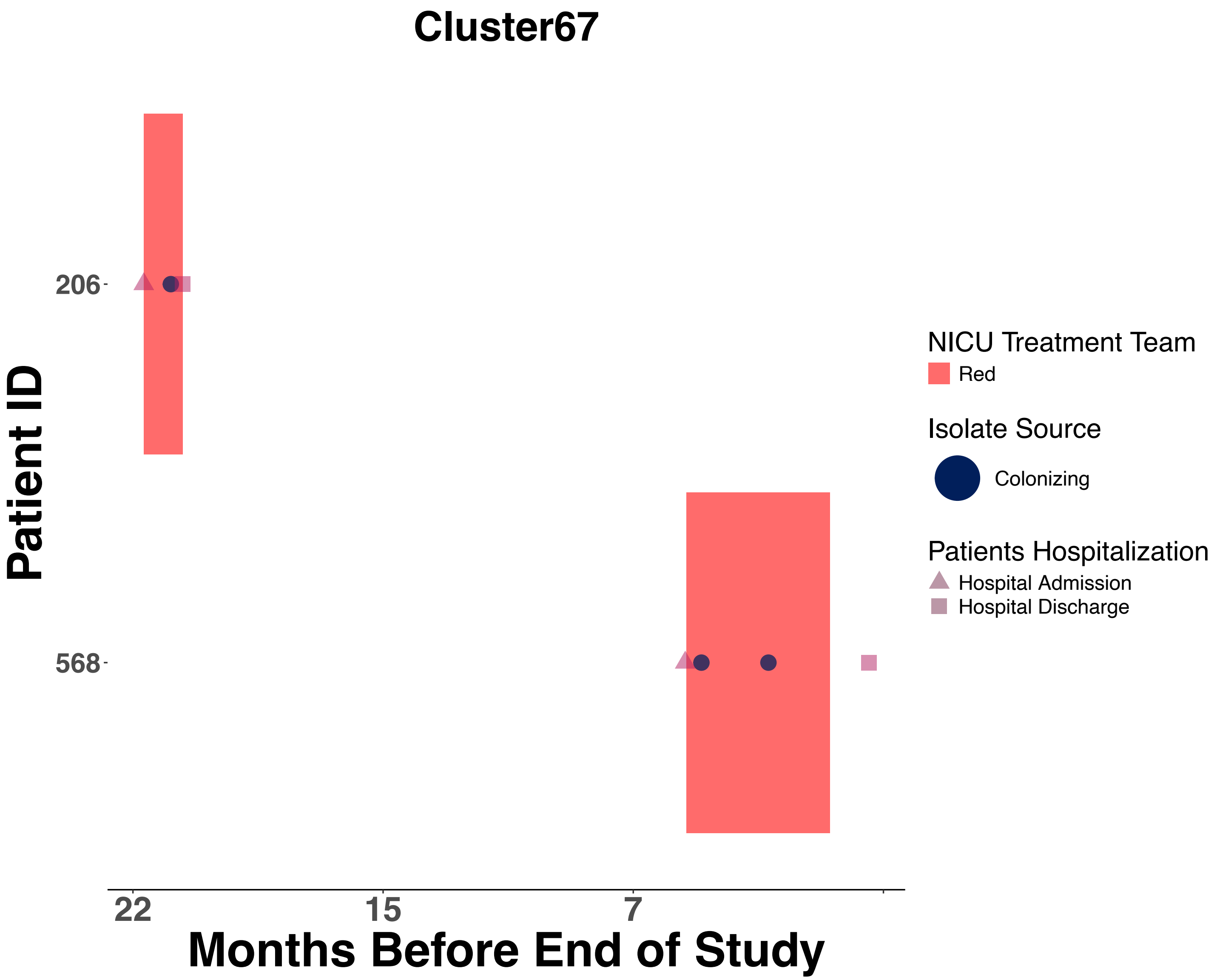
Cluster67
Patient ID
206
568
22
15
7
Months Before End of Study
NICU Treatment Team
Red
Isolate Source
Colonizing
Patients Hospitalization
Hospital Admission
Hospital Discharge

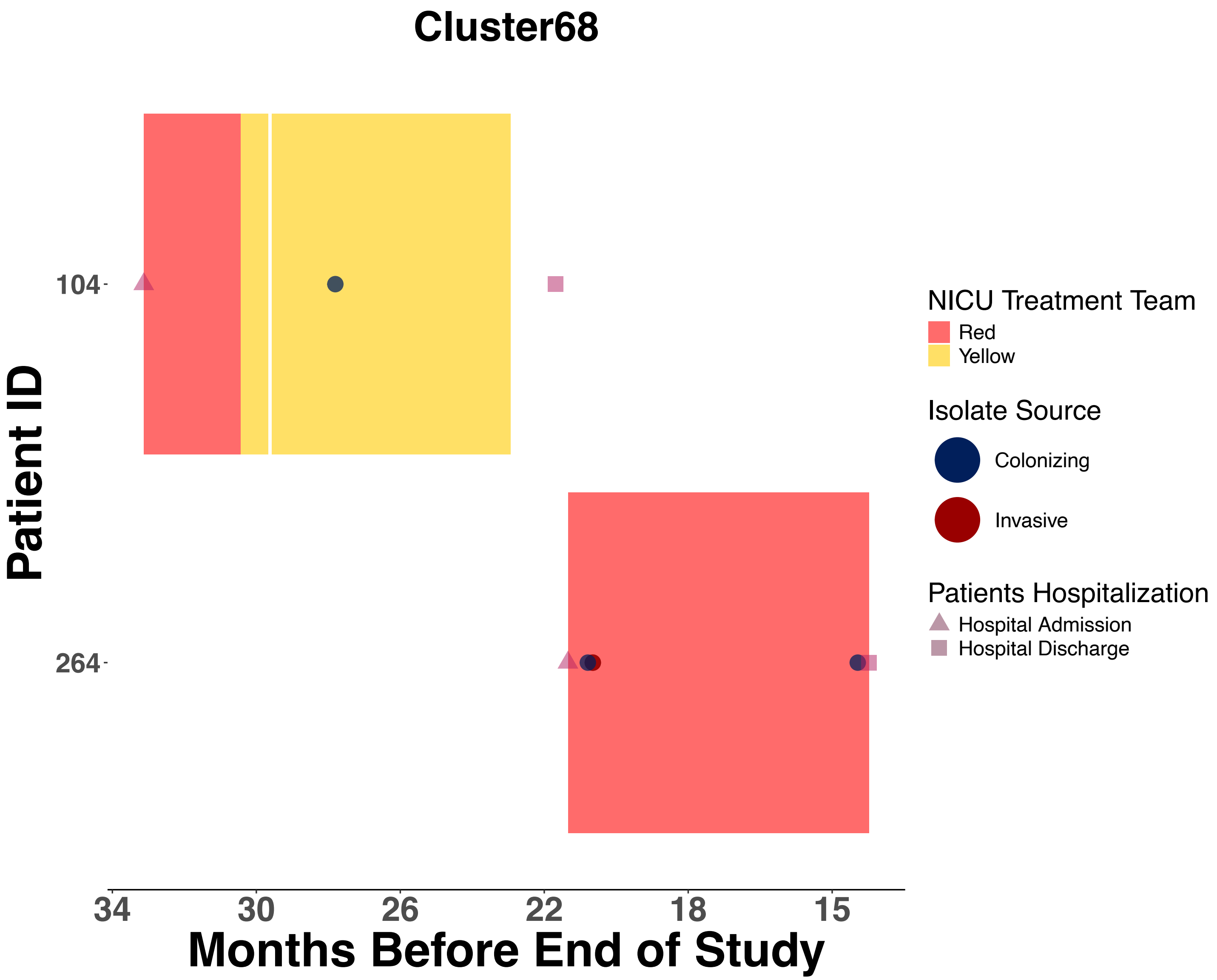
Cluster68
Patient ID
104
264
34
30
26
22
18
15
Months Before End of Study
NICU Treatment Team
Red
Yellow
Isolate Source
Colonizing
Invasive
Patients Hospitalization
Hospital Admission
Hospital Discharge

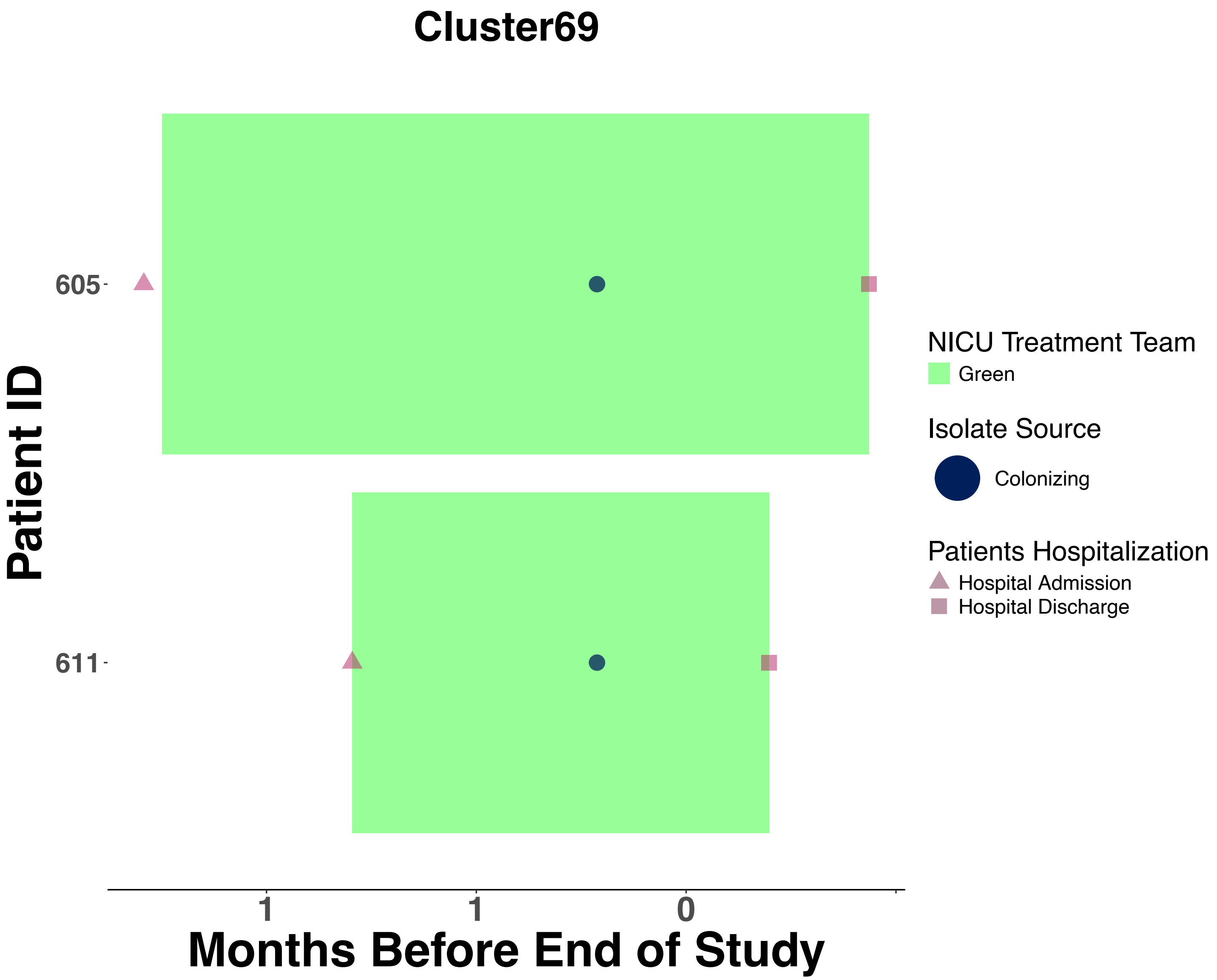
Cluster69
Patient ID
605
611
1
1
0
Months Before End of Study
NICU Treatment Team
Green
Isolate Source
Colonizing
Patients Hospitalization
Hospital Admission
Hospital Discharge

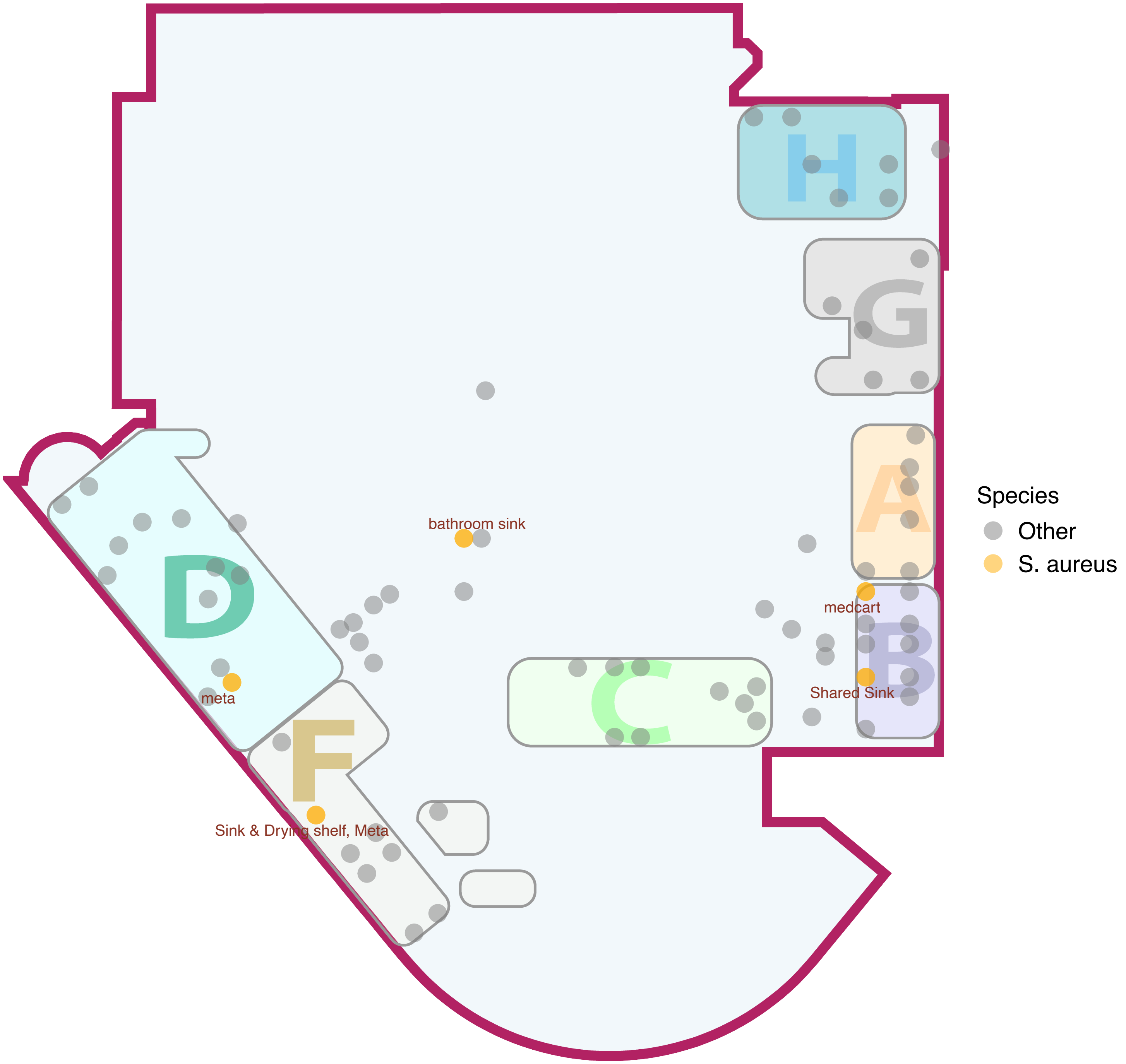

Figure S6: NICU Floorplan Indicating Environmental Sampling Locations

NICU floorplan depicting the 210 environmental sampling locations surveyed across all seven sections. Yellow dots indicate sites with positive S. aureus cultures, while gray dots represent locations where other bacterial species were detected.
